# Analyzing the multidimensionality of biological aging with the tools of deep learning across diverse image-based and physiological indicators yields robust age predictors

**DOI:** 10.1101/2021.04.25.21255767

**Authors:** Alan Le Goallec, Sasha Collin, Samuel Diai, Jean-Baptiste Prost, M’Hamed Jabri, Théo Vincent, Chirag J. Patel

## Abstract

It is hypothesized that there are inter-individual differences in biological aging; however, differences in aging among (heart images vs. electrophysiology) and across (e.g., brain vs heart) physiological dimensions have not been systematically evaluated and compared. We analyzed 676,787 samples from 502,211 UK Biobank participants aged 37-82 years with deep learning approaches to build a total of 331 chronological age predictors on different data modalities such as videos (e.g. heart magnetic resonance imaging [MRI]), images (e.g. brain, liver and pancreas MRIs), time-series (e.g. electrocardiograms [ECGs], wrist accelerometer data) and scalar data (e.g. blood biomarkers) to characterize the multiple dimensions of aging. We combined these age predictors into 11 main aging dimensions, 31 subdimensions and 84 sub-subdimensions ensemble models based on specific organ systems. Heart dimension features predict chronological age with a testing root mean squared error (RMSE) and standard error of 2.83±0.04 years and musculoskeletal dimension features predict age with a RMSE of 2.65±0.04 years. We defined “accelerated” agers as participants whose predicted age was greater than their chronological age and computed the correlation between these different definitions of accelerated aging. We found that most aging dimensions are modestly correlated (average correlation=.139±.090) but that dimensions that are biologically related tend to be more positively correlated. For example, we found that heart anatomical (from MRI) accelerated aging and heart electrical (from ECG) accelerated aging are correlated (average Pearson of .249±.005). Overall, most dimensions of aging are complex traits with both genetic and non-genetic correlates. We identified 9,697 SNPs in 3,318 genes associated with accelerated aging and found an average GWAS-based heritability for accelerated aging of 26.1±7.42% (e.g. heart aging: 35.2±1.6%). We used GWAS summary statistics to estimate genetic correlation between aging dimensions and we found that most aging dimensions are genetically not correlated (average correlation=.104±.149). However, on the other hand, specific dimensions were genetically correlated, such as heart anatomical and electrical accelerated aging (Pearson rho .508±.089 correlated [r_g]). Finally, we identified biomarkers, clinical phenotypes, diseases, family history, environmental variables and socioeconomic variables associated with accelerated aging in each aging dimension and computed the correlation between the different aging dimensions in terms of these associations. We found that environmental and socioeconomic variables are similarly associated with accelerated aging across aging dimensions (average correlations of respectively .639±.180 and .607±.309). Dimensions are weakly correlated with each other, highlighting the multidimensionality of the aging process. Our results can be interactively explored on the following website: https://www.multidimensionality-of-aging.net/

## Background

The United Kingdom population is aging because of low fertility rate and improved healthcare during the last century^1^. The same trend can be observed in the US^2^ and worldwide^3–15^. For example, the number of Americans older than 85 years is expected to triple before 2050^16^. Because a large number of diseases such as cardiovascular disease [CVD], hypertension, cancer, osteoarthritis, type 2 diabetes, osteoporosis, dementia, depression, Parkinson’s disease and Alzheimer’s disease are associated with age^16–21^, it is expected that their prevalence will increase. The burden of these diseases negatively impacts the quality of life of the elderly^22–24^, limits the gains in life expectancy^23, 25, 26^, and strongly affects healthcare costs, which are therefore projected to starkly increase in the coming decades ^1, 27, 28^.

To reduce the burden of age-related diseases, two strategies can be undertaken in parallel. The first is to develop treatments for the different diseases, and the second is to target their common root by slowing aging^29–34^. To better explain what slowing aging entails, we introduce the concept of biological age. When we casually refer to age, we refer to what will be described in this paper as “chronological age” [CA]. CA is simply the measure of how much time has passed since an individual was born and is not, in and of itself, the cause of age-related diseases. In contrast, biological age is the measure of how much damage and wear and tear the body has accumulated over time and is the true underlying cause of age-related diseases^35^. Accelerated agers are individuals whose biological age is higher than their CA. While biological age is an intuitive concept, accurately defining it and measuring it is a challenging task^35^.

Several aging biomarkers such as telomere length^36–44^ have already been identified^45–64^, and biological age predictors such as the DNA methylation clock^65–68^ have been built from them. We discuss these existing biological age predictors more in detail in the supplemental.

Aging may be a “multidimensional” process whose manifestation may be organ- or tissue-specific^69^. As the number of biological age predictors have increased, it became clear that the predictors were not all capturing the same facets of aging^35^. If they were, being an accelerated ager in terms of telomere length would be highly correlated with being an accelerated ager in terms of DNA methylation, for example. Instead, these two aging dimensions seem to be largely uncorrelated^70^. More generally, it seems that most aging dimensions evaluated so far are either uncorrelated, weakly positively correlated, or even weakly negatively correlated^71, 72^. While studying the multidimensionality of aging is a rapidly emerging field^71–82^, extensively studying the multidimensionality of aging is, however, a challenging task because most datasets only provide data from a specific organ system. Second, the analytic models required to build biological predictors of age are complex and training some of the models such as the convolutional neural networks requires expertise ^83, 84^ and use of large computational resources for extended periods of time.

In the following, we leveraged 676,787 samples from 502,211 UK Biobank^85^ [UKB] participants aged 37-82 years old to investigate the multidimensionality of aging. We built 331 age predictors that we then hierarchically ensembled into 11 aging main dimensions and 17 subdimensions, based on the organ system to which we assigned the predictive biomarkers. Then, we investigated the genetic and environmental components of these 28 aging dimensions by performing a Genome Wide Association Study [GWAS] and X-Wide Association Study [XWAS]^86^ on the 28 accelerated aging phenotypes, respectively. Finally, we evaluated how these different aging dimensions correlate. We computed the correlation between the 28 accelerated aging phenotypes before computing the genetic correlation, and the environmental correlation between these phenotypes.

## Results

### Overview of the dataset

We leveraged the UK Biobank [UKB], a dataset of 676,787 samples covering 502,211 participants, with an age range of 37 to 82 years (Supplementary Figure 1). We hierarchically classified the biological data modalities collected from these participants (Figure 1) into 11 main aging dimensions, 31 subdimensions and 84 sub-subdimensions (Table 1). The general philosophy behind this three-level hierarchy is that the main dimensions represent aging of different organ systems, such as the brain, eyes, arteries, heart and bones. The sub-dimensions represent different facets of aging for one of the main dimensions. For example, heart aging encompasses both anatomical aging, captured by MRI videos, and electrical aging, captured by ECGs. Finally, the sub-sub dimensions represent different views or data preprocessing of the same subdimension. For example, heart anatomical aging is captured by MRI videos taken under three different views (two-chamber, three-chamber and four-chamber views) and using two different preprocessing methods (raw images and contrasted images). We tried to stick to this organization as much as possible, but we had to make exceptions for some of the datasets. For example, brain cognitive is a subdimension for which a large number of different measures were collected by UKB, such as reaction time tests, memory tests and fluid intelligence tests. Therefore, we treated these different facets of cognitive performance as sub-subdimensions, even though they are not different views and preprocessing of the cognitive subdimension.

**Figure 1:**
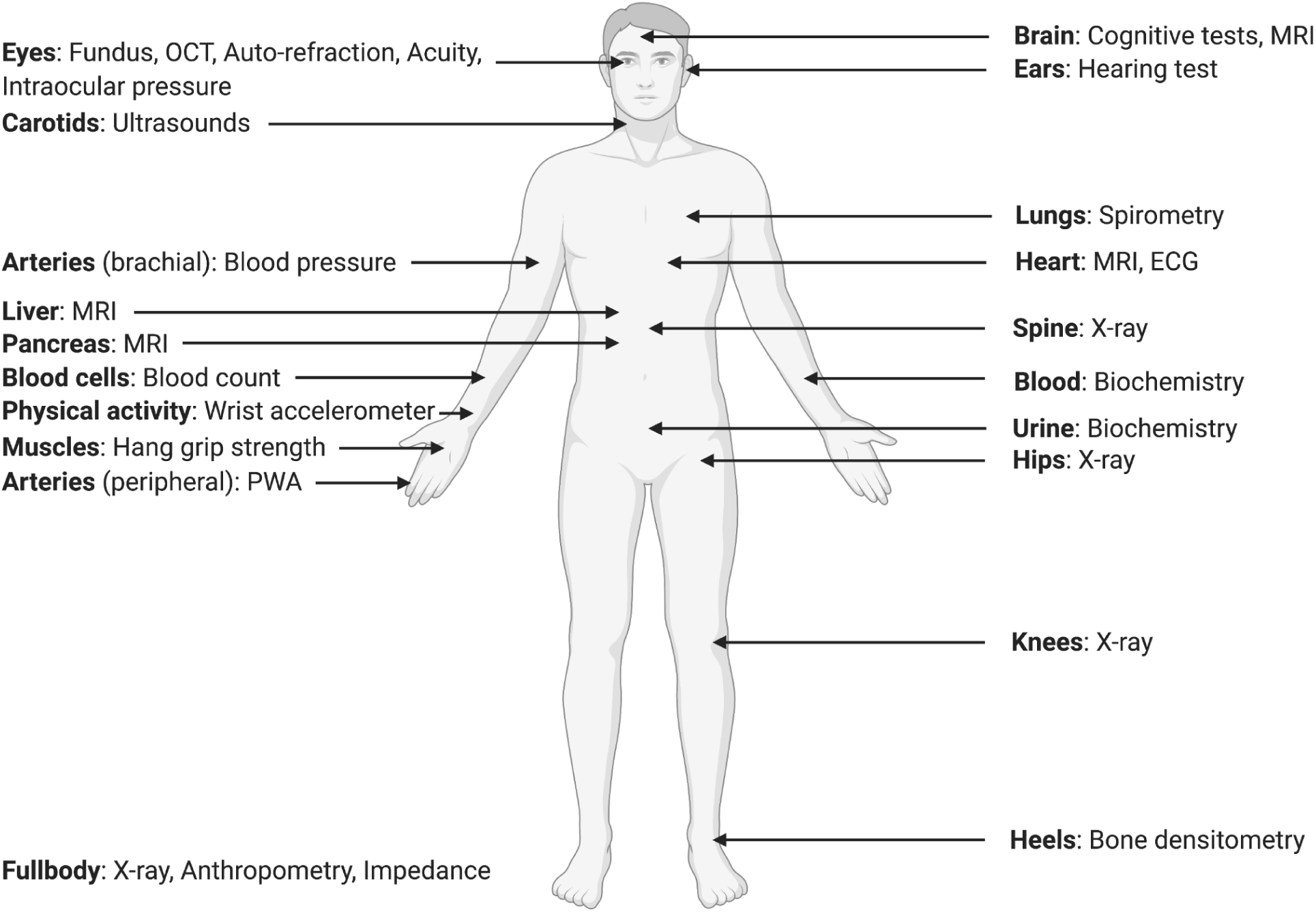
Overview of the different UK Biobank data modalities leveraged to build chronological age predictors

**Table 1:**
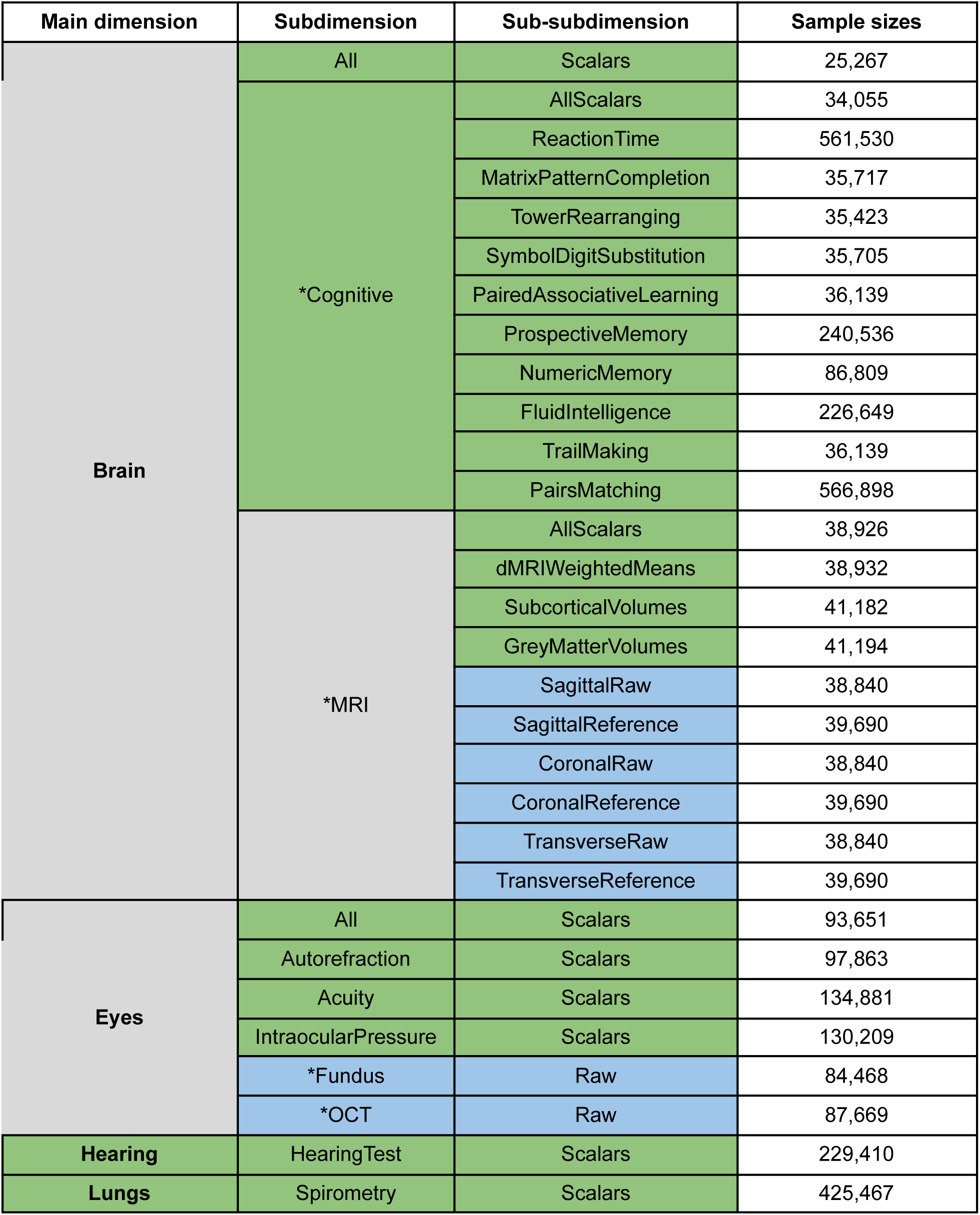
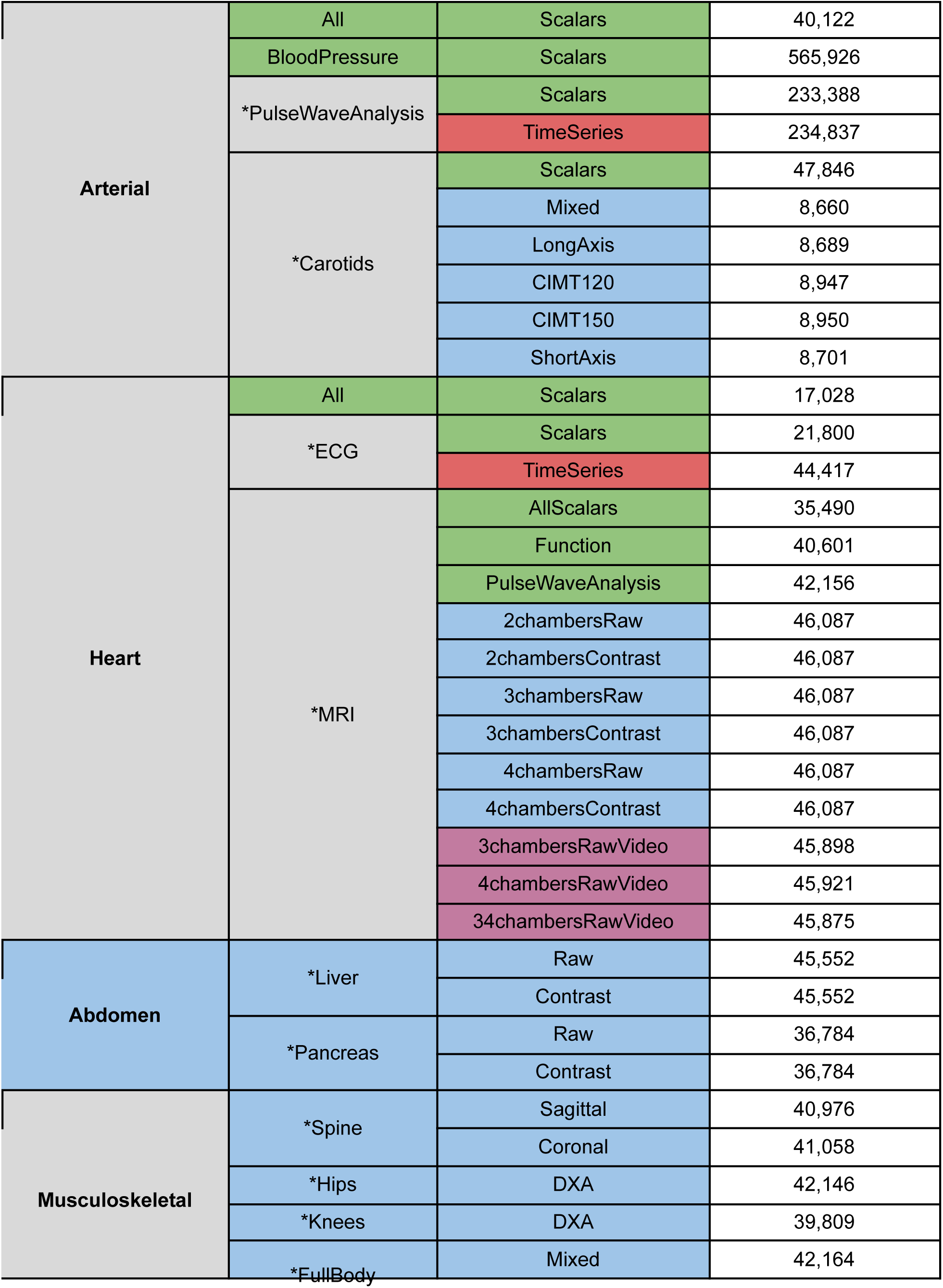
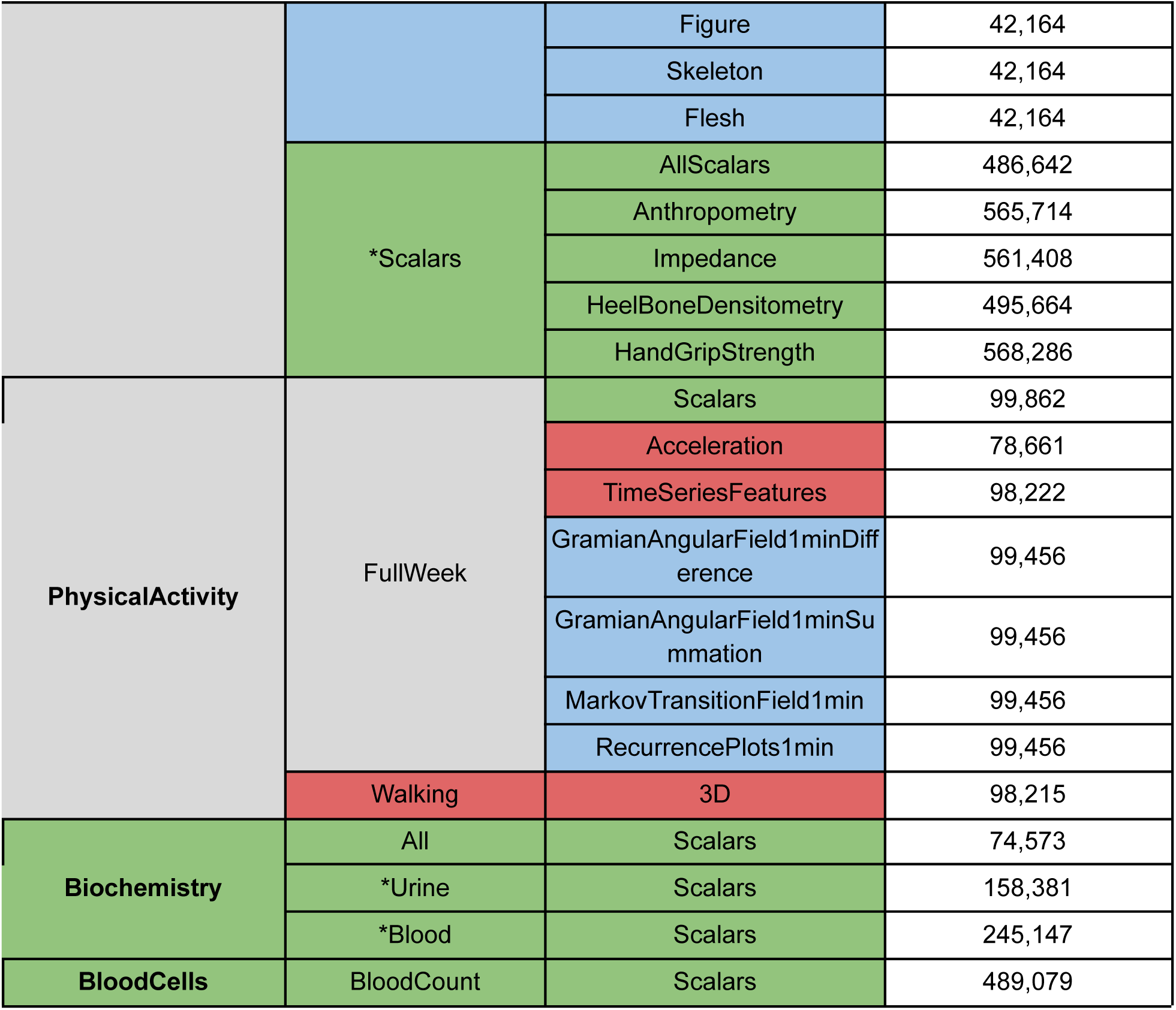
Hierarchy between the different aging dimensions, subdimensions and sub-subdimensions. Legend: Green - Scalar biomarkers; Red - Time Series; Blue - Images; Purple - Videos; Gray - Mixed types

| Main dimension | Subdimension | Sub-subdimension | Sample sizes |
| --- | --- | --- | --- |
| Brain | All | Scalars | 25,267 |
|  | *Cognitive | AllScalars | 34,055 |
|  |  | ReactionTime | 561,530 |
|  |  | MatrixPatternCompletion | 35,717 |
|  |  | TowerRearranging | 35,423 |
|  |  | SymbolDigitSubstitution | 35,705 |
|  |  | PairedAssociativeLearning | 36,139 |
|  |  | ProspectiveMemory | 240,536 |
|  |  | NumericMemory | 86,809 |
|  |  | FluidIntelligence | 226,649 |
|  |  | TrailMaking | 36,139 |
|  |  | PairsMatching | 566,898 |
|  | *MRI | AllScalars | 38,926 |
|  |  | dMRIWeightedMeans | 38,932 |
|  |  | SubcorticalVolumes | 41,182 |
|  |  | GreyMatterVolumes | 41,194 |
|  |  | SagittalRaw | 38,840 |
|  |  | SagittalReference | 39,690 |
|  |  | CoronalRaw | 38,840 |
|  |  | CoronalReference | 39,690 |
|  |  | TransverseRaw | 38,840 |
|  |  | TransverseReference | 39,690 |
| Eyes | All | Scalars | 93,651 |
|  | Autorefraction | Scalars | 97,863 |
|  | Acuity | Scalars | 134,881 |
|  | IntraocularPressure | Scalars | 130,209 |
|  | *Fundus | Raw | 84,468 |
|  | *OCT | Raw | 87,669 |
| Hearing | HearingTest | Scalars | 229,410 |
| Lungs | Spirometry | Scalars | 425,467 |
| Arterial | All | Scalars | 40,122 |
|  | BloodPressure | Scalars | 565,926 |
|  | *PulseWaveAnalysis | Scalars | 233,388 |
|  |  | TimeSeries | 234,837 |
|  | *Carotids | Scalars | 47,846 |
|  |  | Mixed | 8,660 |
|  |  | LongAxis | 8,689 |
|  |  | CIMT120 | 8,947 |
|  |  | CIMT150 | 8,950 |
|  |  | ShortAxis | 8,701 |
| Heart | All | Scalars | 17,028 |
|  | *ECG | Scalars | 21,800 |
|  |  | TimeSeries | 44,417 |
|  | *MRI | AllScalars | 35,490 |
|  |  | Function | 40,601 |
|  |  | PulseWaveAnalysis | 42,156 |
|  |  | 2chambersRaw | 46,087 |
|  |  | 2chambersContrast | 46,087 |
|  |  | 3chambersRaw | 46,087 |
|  |  | 3chambersContrast | 46,087 |
|  |  | 4chambersRaw | 46,087 |
|  |  | 4chambersContrast | 46,087 |
|  |  | 3chambersRawVideo | 45,898 |
|  |  | 4chambersRawVideo | 45,921 |
|  |  | 34chambersRawVideo | 45,875 |
| Abdomen | *Liver | Raw | 45,552 |
|  |  | Contrast | 45,552 |
|  | *Pancreas | Raw | 36,784 |
|  |  | Contrast | 36,784 |
| Musculoskeletal | *Spine | Sagittal | 40,976 |
|  |  | Coronal | 41,058 |
|  | *Hips | DXA | 42,146 |
|  | *Knees | DXA | 39,809 |
|  | *FullBody | Mixed | 42,164 |
|  |  | Figure | 42,164 |
|  |  | Skeleton | 42,164 |
|  |  | Flesh | 42,164 |
|  | *Scalars | AllScalars | 486,642 |
|  |  | Anthropometry | 565,714 |
|  |  | Impedance | 561,408 |
|  |  | HeelBoneDensitometry | 495,664 |
|  |  | HandGripStrength | 568,286 |
| **PhysicalActivity** | FullWeek | Scalars | 99,862 |
|  |  | Acceleration | 78,661 |
|  |  | TimeSeriesFeatures | 98,222 |
|  |  | GramianAngularField1minDifference | 99,456 |
|  |  | GramianAngularField1minSummation | 99,456 |
|  |  | MarkovTransitionField1min | 99,456 |
|  |  | RecurrencePlots1min | 99,456 |
|  | Walking | 3D | 98,215 |
| **Biochemistry** | All | Scalars | 74,573 |
|  | *Urine | Scalars | 158,381 |
|  | *Blood | Scalars | 245,147 |
| **BloodCells** | BloodCount | Scalars | 489,079 |

The datasets can also be classified into four different data modalities: (1) scalar features (non-dimensional variables such as laboratory results), (2) time-series (one-dimensional variables such as ECGs), (3) images (two-dimensional variables such as Liver MRI) and (4) videos (three-dimensional predictors such as heart MRI videos).

### Definition of biological aging and accelerated aging

We used machine learning algorithms to predict chronological age [CA] from each of the 84 sub-subdimensions. We analyzed scalar features with elastic nets [ENs], gradient boosted machines [GBMs] and neural networks [NNs]. We analyzed time series, images and videos with respectively one-dimensional, two-dimensional and three-dimensional convolutional neural networks [CNNs]. We hierarchically ensembled the predictions to build CA predictors for each biological dimension, subdimension, and sub-subdimension. We used a 10-folds cross-validation to generate an unbiased testing prediction for each sample.

For each aging dimension, we estimated each participant’s biological age in this dimension as the testing prediction outputted by the corresponding ensemble model. Similarly, we calculated the accelerated aging for each participant in each aging dimension by taking the difference between the participant’s chronological age and biological age (the prediction residuals computed on the testing set). For example, we estimated the heart age for a participant as the testing prediction generated by the ensemble model built on all the heart data modalities. A participant with a chronological age of 50 years and a biological heart age of 60 years has an accelerated heart age of ten years. We observed a bias in the residuals as a function of chronological age. Young participants are on average predicted older than they are, whereas old participants are on average predicted younger than they are. We corrected both the biological ages and the accelerated aging phenotypes for the bias.

In total, we estimated 331 dimensions or definitions of biological age made available as phenotypes to other researchers through the UK Biobank.

### Phenotypic, genetic and environmental correlation between the different aging dimensions

#### Phenotypic correlation between aging dimensions

We estimated the phenotypic correlation structure between the 331 aging dimensions by computing the correlation matrix between the different accelerated aging phenotypes (Figure 2). The correlation matrix between the 11 aging main dimensions and 17 selected aging subdimensions along its hierarchical structure can be found in Figure 3. The average Pearson correlation between the 331 aging dimensions is .156±.149, based on 54,615 correlations (min=-.185; max = 1.000; median=.095; interquartile range=.116). The minimum correlation is -.185, the maxThis average number does not capture the structure within and between the hierarchy of the dimensions. We identified six levels in the hierarchy of the models: the main biological dimensions (mean correlation .139±.090; 55 correlations), the biological subdimensions (mean correlation .247±.135; 22 correlations), the biological sub-subdimensions (mean correlation .387±.094; 51 correlations), the different views of the same dimension (mean correlation .541±.161; 59 correlations), the images preprocessing methods (mean correlation .658±.083; 18 correlations) and the algorithms (mean correlation .838±.120; 158 correlations). In general, the correlation between the accelerated aging dimensions increases as one goes down the hierarchy, since the models on which the predictors were built become more and more similar. This hierarchy in terms of correlation between the different models can for example clearly be observed for the brain accelerated aging dimensions (Supplementary Figure 2).

**Figure 2:**
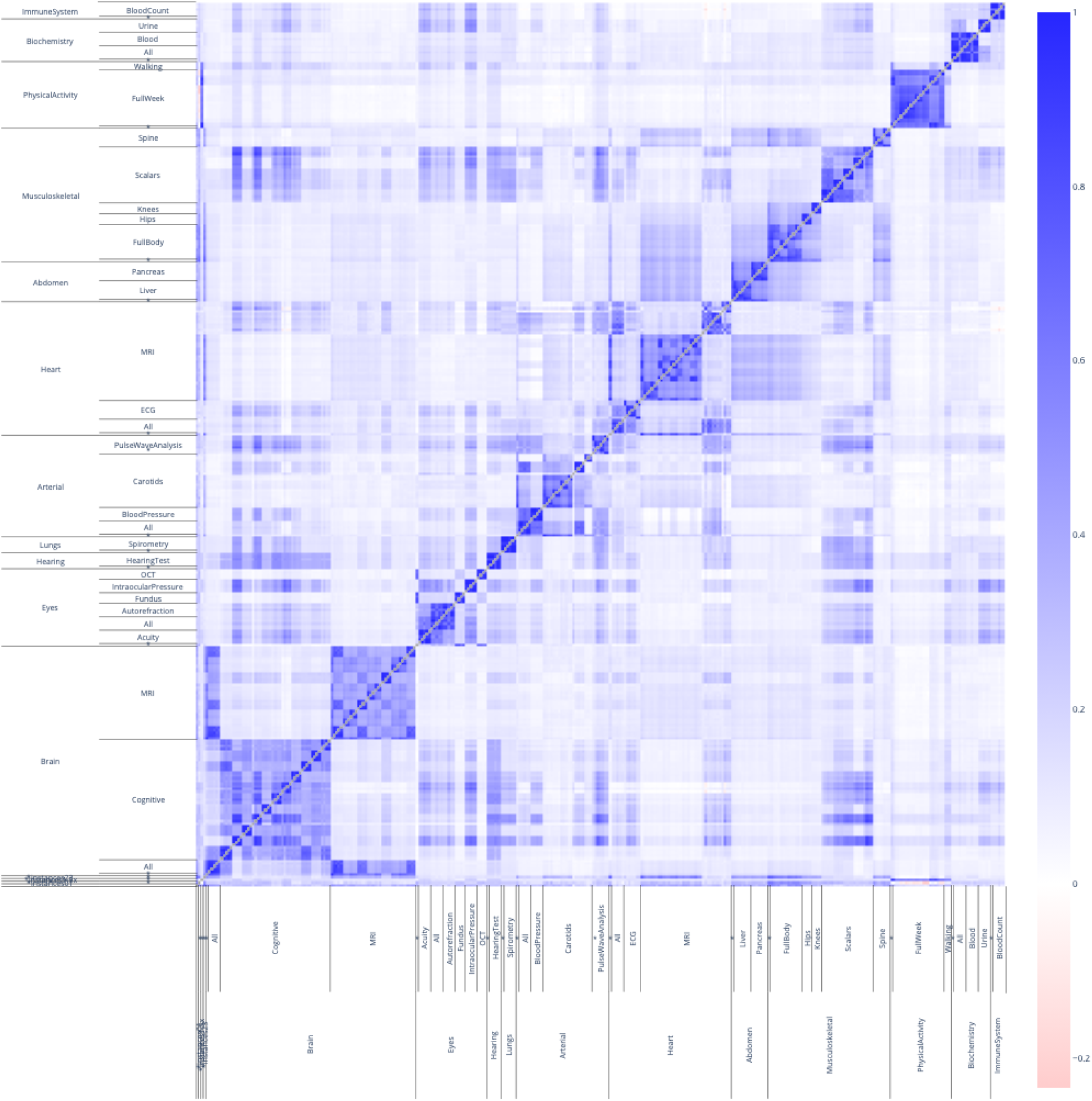
**Phenotypic correlation between accelerated aging as defined by each of the 331 models** The correlation heatmap displays the Pearson correlation between accelerated aging for each pair of aging dimensions. The x and y axes are hierarchically organised to display the aging main dimensions at the higher level, then the aging subdimensions, and finally the aging sub-subdimensions. Each aging subdimension corresponds to several rows and columns because of the different algorithms that were trained to predict chronological age from this dataset. For the sake of readability, these are not displayed.

**Figure 3:**
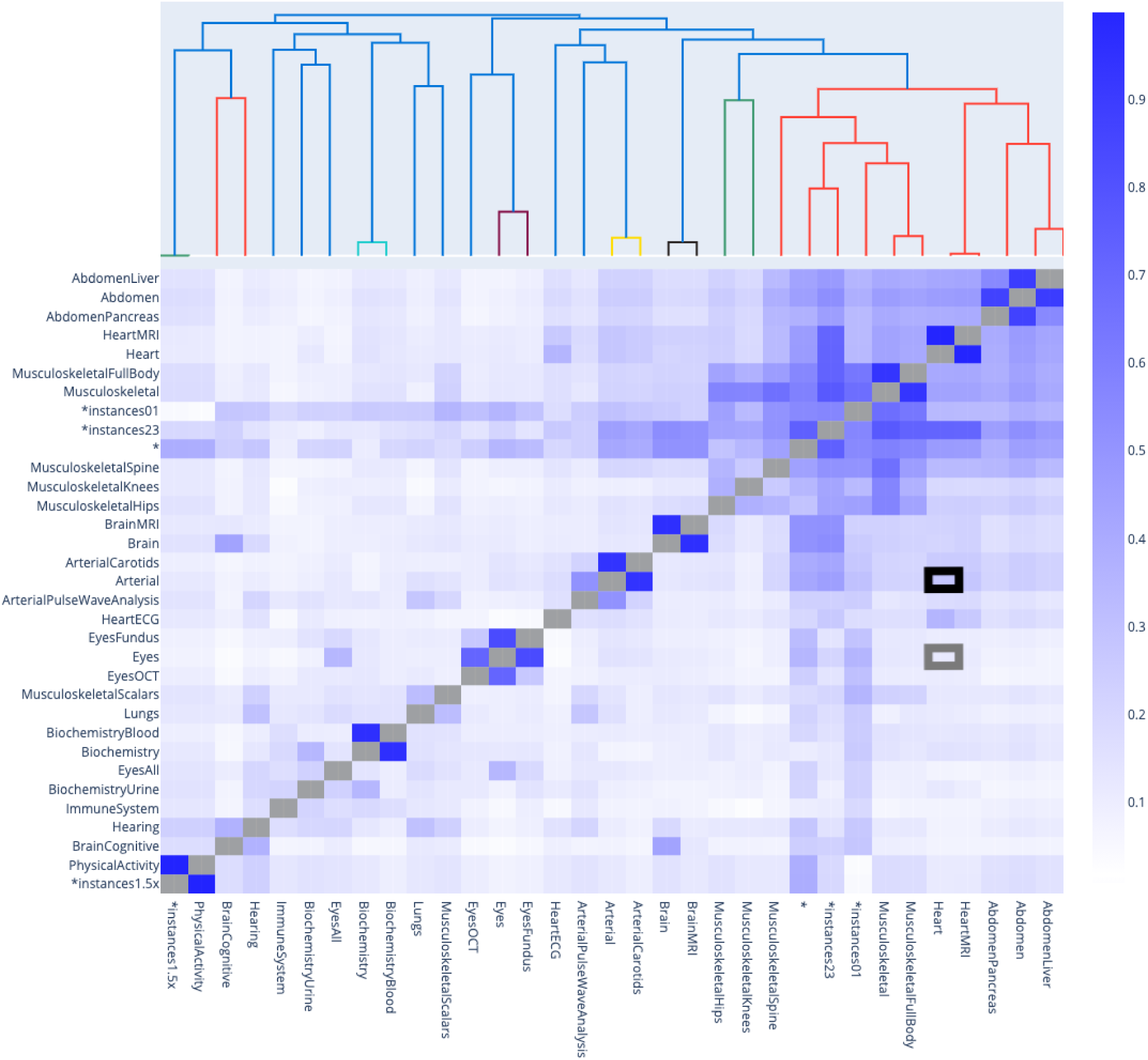
**Phenotypic correlation between accelerated aging in the main aging dimensions and 17 selected aging subdimensions** The gray box highlights the correlation between accelerated heart aging and accelerated eye aging. The black box highlights the correlation between accelerated heart aging and accelerated arterial aging.

The first level is the correlation between the 11 main aging dimensions, such as accelerated brain aging, eye aging, heart aging, arterial aging and musculoskeletal aging. We found that the 11 aging main dimensions are weakly correlated with an average correlation of .139±.090 (55 correlations). For example, eye accelerated aging and heart accelerated aging are .100±.012 correlated (see gray box in Figure 3). We found that accelerated aging dimensions built on biologically similar datasets tend to be more correlated. For example, arterial accelerated aging and heart accelerated aging are .260±.160 correlated (see black box in Figure 3).

The second level is the correlation between biologically driven subdimensions. For example, brain aging encompasses both cognitive aging and anatomically measured (MRI-based) aging. We found that the average correlation between biologically driven aging subdimensions is .247±.135 (22 correlations). Specifically, brain subdimensions (cognitive and anatomical, one correlation) are .113 correlated, eye subdimensions (fundus, OCT, intraocular pressure, autorefraction and acuity) are .211±.119 correlated (10 correlations), arterial subdimensions (carotids ultrasounds and pulse wave analysis recorded on the finger) are .203 correlated (one correlation), heart subdimensions (anatomical and electrical) are .248 correlated (one correlation), abdomen subdimensions (liver and pancreas) are .539 correlated (one correlation), musculoskeletal subdimensions (spine, hips, knees, full body and scalar predictors) are .272±.130 correlated (10 correlations), physical activity subdimensions (full week and walking) are .309 correlated (one pair) and biochemistry subdimensions (blood and urine) are .090 correlated.

The third level is the correlation between biologically related sub-subdimensions. For example, brain cognitive is a subdimension of brain aging that encompasses ten sub-subdimensions such as reaction time and fluid intelligence. We found that the average correlation between biologically driven aging sub-subdimensions is .387±.094 (51 correlations). Specifically, brain cognitive sub-subdimensions (reaction time, matrix pattern completion, tower rearranging, symbol digit substitution, paired associative learning, prospective memory, numeric memory, fluid intelligence, trail making and paris matching) are .391±0.098 correlated (45 correlations) and musculoskeletal sub-subdimensions (anthropometry, impedance, heel bone densitometry and hand grip strength) are .345±.092 correlated (6 correlations).

The fourth level is the correlation between different views of the same aging dimensions. For example, we used brain MRI images from the sagittal, the coronal and the transverse planes, and the information contained in the 3D MRI images were also summarized by UKB in three sets of scalar features: diffusion MRI weighted means, subcortical volumes and gray matter volumes. We found that the average correlation between different views of the same dimension is .541±.161 (59 correlations). Specifically, brain MRI-based views (sagittal plane, coronal plane, transverse plane, diffusion MRI weighted means, subcortical volumes and gray matter volumes) are .499±.062 correlated (18 correlations), arterial pulse wave analysis views (raw time series and scalar features extracted from these time series) are .634 correlated (one correlation), carotid ultrasound views (short axis, long axis, CIMT120, CIMT150) are .392±.167 correlated (10 correlations), heart ECG views (time series and scalar features extracted from these time series) are .439 correlated (one correlation), heart MRI views (two-chamber, three-chamber and four-chamber views) are .542±.067 correlated (six correlations), musculoskeletal spine views (sagittal and coronal) are .487 correlated (one pair), musculoskeletal full body views (flesh and figure) are .543 correlated (one correlation) and physical activity full week views (raw acceleration times series, features time series, scalar features extracted from the time series, Gramian angular field (difference and summation) generated from the time series, Markov transition field generated from the time-series and recurrence plot generated from the time series) are .693±.111 correlated (21 correlations).

The fifth level is the correlation between different preprocessings on the same data. For example, we used both raw heart MRI images, and the same images on which we applied a contrast filter. We found that the average correlation between image preprocessing is .658±.083 (18 pairs). Specifically, we found that raw and “reference” (the video is summarized into a single time frame) brain MRI images are .547±.026 correlated (six correlations), raw and contrasted images are .714±.021 correlated (10 correlations) and full body “figure” and skeleton (figure images with the silhouette around the skeleton removed) images are .714±.004 correlated (two correlations).

The sixth (and last) level is the correlation between different algorithms trained on the same dataset. We found that the average correlation between algorithms is .838±.120 (158 correlations). Specifically, elastic net and light GBM are .837±.113 correlated (41 correlations), elastic net and neural network are .859±.11 correlated (41 correlations), light GBM and neural network are .908±.075 correlated (41 correlations) and InceptionV3 and InceptionResNetV2 are .731±.111 correlated (35 correlations).

We did not identify any negative correlations between the 28 selected aging dimensions. When considering the 331 aging dimensions, we found that 2.1% (116 out of 54,615) of the pairwise correlations were negative. These low correlations tend to be driven by small sample size and/or poor prediction performance by at least one of the two models. For example, the lowest correlation (-0.234, between the ensemble model built on datasets from UKB’s first instance and a full week physical activity-based predictors) has a sample size of 97 (mean sample size for the correlations: 42,371). If we discard this cluster of negative correlations, the next lowest correlation is -.136 (correlation between accelerated aging as defined by the elastic net built on heart function scalar data (R^2^=0.9±0.1%) and accelerated aging as defined by the elastic net built on blood counts (R^2^=5.8±0.1%)).

#### Genetic correlation between aging dimensions

We computed genetic correlations between the aging dimensions and hierarchically clustered them (Figure 4). After filtering out the correlations for which the sample size was smaller than 15,000 participants, we found that the average genetic correlation between the 11 main dimensions is .104±.149 (18 correlations). The highest genetic correlation is between musculoskeletal aging and abdominal aging (.573±.042). The next three highest correlations are approximately .2 (heart and blood cells: .200±.105; hearing and physical activity: .189±.09; musculoskeletal and brain: .186±.062). The remaining correlations are lower than .15. We found four negative correlations, none of them being significantly negative. For example, the genetic correlation between accelerated eye aging and accelerated hearing aging is -.035±.044.

**Figure 4:**
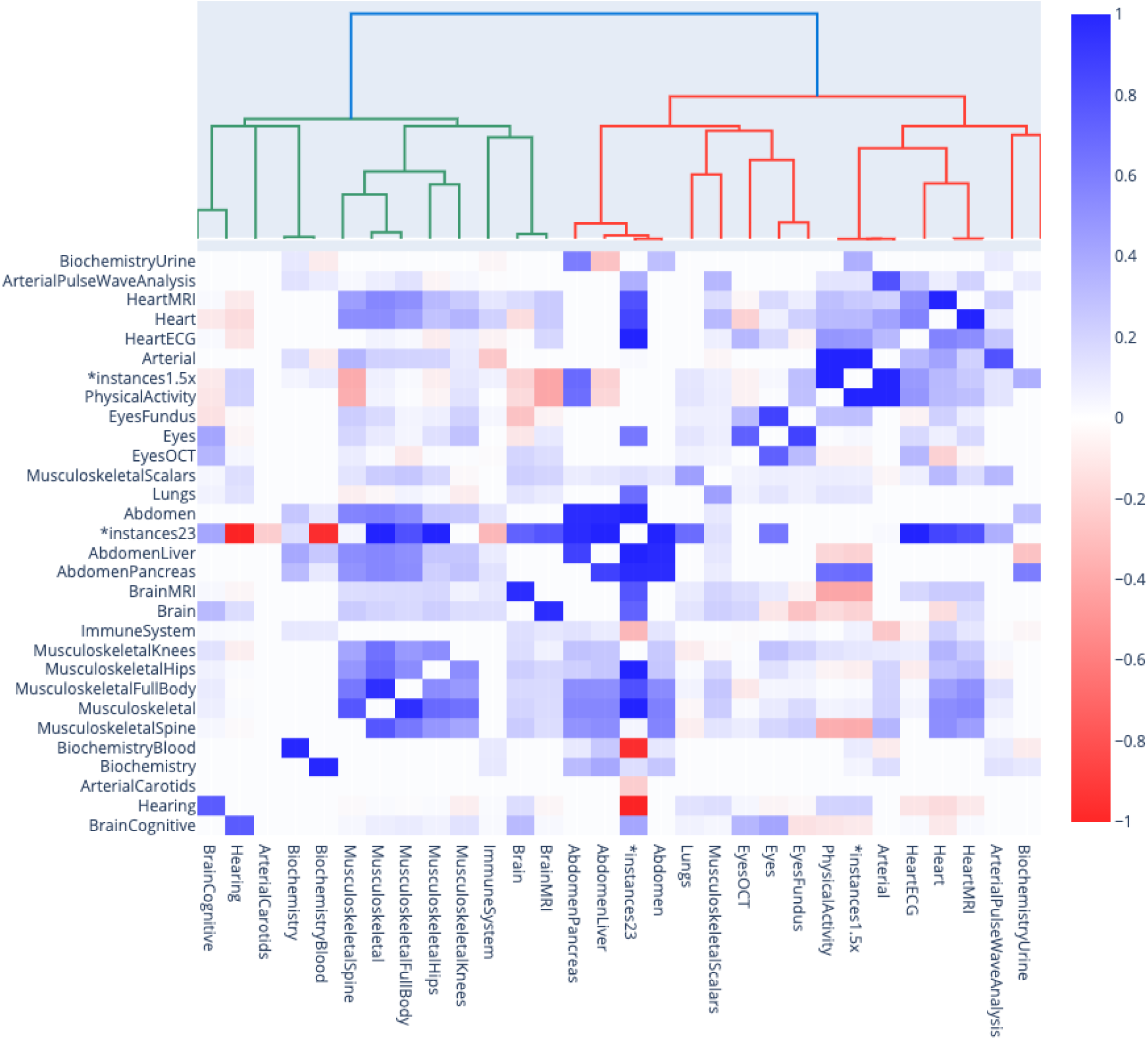
**Genetic correlation between the main aging dimensions and 17 selected aging subdimensions.** *instances23 and *instances1.5x models denote ensemble models built across several main aging dimensions. For example, *instances23 refers to the model that we built on datasets primarily issued from instances 2 and 3 of the UKB dataset. See Methods for more detail.

We then computed the genetic correlations between the different subdimensions of each main dimension, and we found an average correlation of .338±.255 (15 correlations). We found that the subdimensions of the main dimensions brain and biochemistry are uncorrelated (brain: anatomical and cognitive accelerated aging are .019±.056 correlated; biochemistry: blood and urine accelerated aging are -.080±.043 correlated). In contrast, we found that the subdimensions of abdominal aging are strongly correlated (liver and pancreas: .863±.036), along with the subdimensions of heart aging (anatomical and electrical: .508±.089) and the subdimensions of eye aging (OCT-based and fundus-based: .299±.025). Finally, we found that the image-based subdimensions of musculoskeletal aging (full body, spine, hip and knee) are .494±.064 correlated (six correlations), whereas the scalar features-based musculoskeletal subdimension (anthropometry, impedance, heel bone densitometry and hand grip strength) is only .126±.098 correlated (four correlations) with the images-based musculoskeletal subdimension.

##### Comparison between phenotypic and genetic correlations between aging dimensions

For each pair of aging dimensions, we compared the genetic correlation with the phenotypic correlation (computed using the corrected chronological age prediction residuals). After filtering out the correlations for which the sample size was lower than 15,000 samples (because of the associated high standard deviation on the genetic correlation measure), we found that the Pearson correlation between the genetic and the phenotypic correlations is .781 for the main aging dimensions and .936 for the aging subdimensions. The correlation between aging dimensions for which the difference between phenotypic correlation and genetic correlation is the highest in absolute value is the correlation between brain cognitive aging and hearing aging (difference: -.392±.070; phenotypic correlation: .351±.005; genetic correlation: .743±.070).

#### Correlation between aging dimensions in terms of association with biomarkers, clinical phenotypes, diseases, family history, environmental variables and socioeconomics

We use “X” to refer to all nongenetic variables measured in the UK Biobank (biomarkers, clinical phenotypes, diseases, family history, environmental and socioeconomic variables). We performed an X-Wide Association Study [XWAS] to identify which of the 4,372 biomarkers classified in 21 subcategories (Supplementary Table 6), 187 clinical phenotypes classified in 11 subcategories (Supplementary Table 11), 2,073 diseases classified in 26 subcategories (Supplementary Table 16), 92 family history variables (Supplementary Table 21), 265 environmental variables classified in nine categories (Supplementary Table 24), and 91 socioeconomic variables classified in five categories (Supplementary Table 29) are associated (p-value threshold of 0.05 and Bonferroni correction) with accelerated aging in the selected 28 dimensions. The XWAS results are described further below under Results - Identification of nongenetic factors associated with accelerated aging.

We compared these associations across the different aging dimensions to understand if X-variables associated with accelerated aging in one aging dimension are also associated with accelerated aging in another aging dimension. For example, in terms of environmental variables, are the diets that protect against heart aging the same as the diets that protect against brain aging?

We found that the average correlation between main aging dimensions is .359±.229 in terms of biomarkers, .643±.142 in terms of associated clinical phenotypes, .488±.198 in terms of diseases, .030±.519 in terms of family history, .639±.180 in terms of environmental variables and .607±.309 in terms of socioeconomics.

We found that the average correlations between the aging main dimensions are higher than the average correlations between the aging subdimensions (Supplementary Figure 3 and Supplementary Table 1). For a greater level of detail, the average correlation between the aging main dimensions and between the aging subdimensions for each of the six main X-categories and their associated 73 X-subcategories can respectively be found in Figure 5 and Supplementary Figure 4. We summarize the key result for each of the six main X-categories below. Finally, the website can be used to display these different correlations between each specific pair of aging dimensions. For the sake of the example, we provide the correlations between brain aging and heart aging in terms of the different X-associations in Supplementary Figure 5.

**Figure 5:**
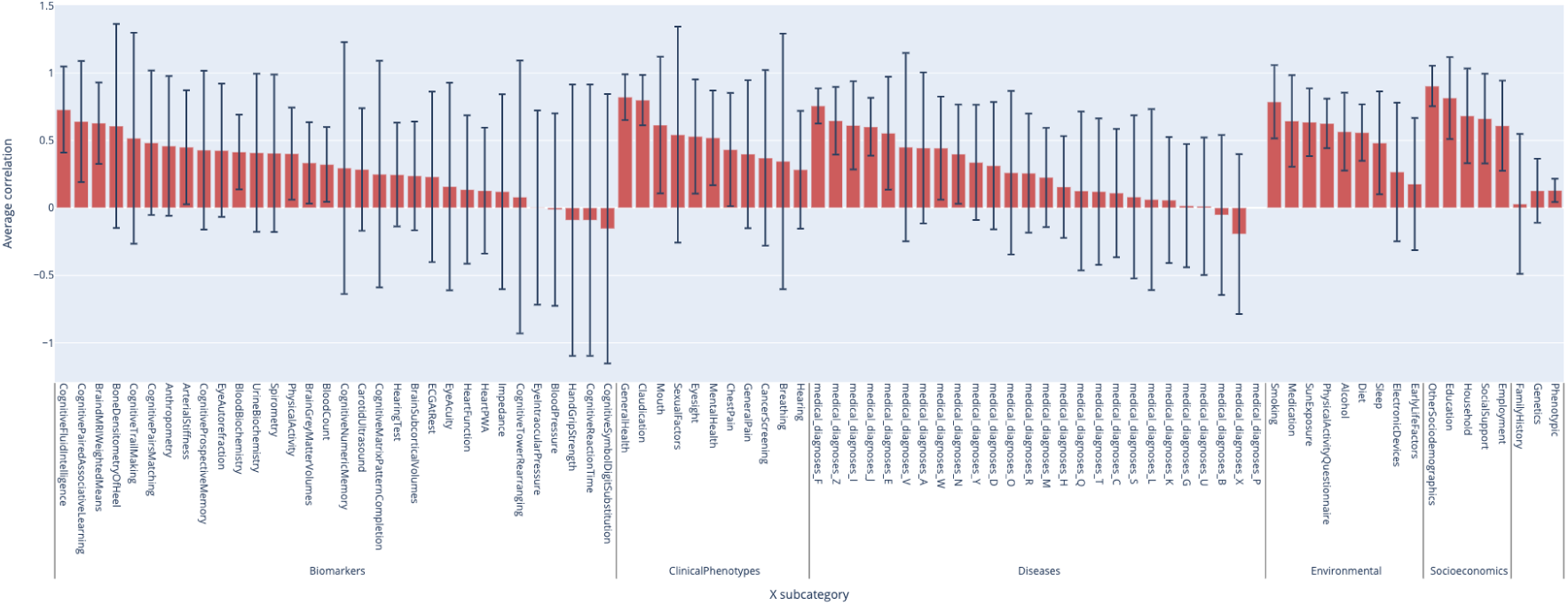
XWAS Summary - Average correlation between aging main dimensions for each X-subcategory

Finally, we predicted accelerated aging in the different aging dimensions from the non-genetic variables, as reported under Results - Predicting accelerated aging from biomarkers, clinical phenotypes, diseases, environmental variables and socioeconomic variables. We computed the correlation between the different aging dimensions in terms of feature importance (Supplementary Figure 6). The detailed results can be found on the website.

##### Biomarkers

We found that the average correlation between accelerated aging main dimensions and between accelerated aging subdimensions in terms of associated biomarkers are respectively .359±.229 and .656±.309 (Supplementary Figure 7). We found that the three biomarker categories with the highest average correlation between aging main dimensions are cognitive fluid intelligence (.729±.319), cognitive paired associative learning (.640±.449) and brain MRI weighted means (.628±.301). Conversely, we found that the three biomarker categories with the lowest average correlation between aging main dimensions are cognitive symbol digits substitution (-.154±.998), cognitive reaction time (-.091±1.005) and hand grip strength (-.091±1.005).

##### Clinical phenotypes

We found that the average correlation between accelerated aging main dimensions and between accelerated aging subdimensions in terms of associated clinical phenotypes are respectively .643±.142 and .668±.336 (Supplementary Figure 8). We found that the three clinical phenotypes categories with the highest average correlation between aging main dimensions are general health (.820±.169), claudication (.799±.186) and mouth (.614 ±.507). Conversely, we found that the three clinical phenotypes categories with the lowest average correlation between aging main dimensions are hearing (.283±.346), breathing (.345±.947) and cancer screening (.371±.650).

##### Diseases

We found that the average correlation between accelerated aging main dimensions and between accelerated aging subdimensions in terms of associated diseases are respectively .488±.198 and .585±.187 (Supplementary Figure 9). We found that the three disease categories with the highest average correlation between aging main dimensions are medical diagnoses F - mental diseases (.756±.130), medical diagnoses Z - diverse diseases (.646±.251) and medical diagnoses I - cardiovascular diseases (.612±.327). Conversely, we found that the three disease categories with the lowest average correlation between aging main dimensions are medical diagnoses X - diseases linked to exposure, poisoning, assault and others (-.194±.592), medical diagnoses B - infectious diseases (-.053±.593) and medical diagnoses U - drug resistance and others (.123±.509).

##### Family history

We found that the average correlation between accelerated aging main dimensions and between accelerated aging subdimensions in terms of associated family history are respectively .030±.519 and .728±.397.

##### Environmental variables

We found that the average correlation between accelerated aging main dimensions and between accelerated aging subdimensions in terms of associated environmental variables are respectively .639±.180 and .624±.321 (Figure 6). We found that the average correlation between aging main dimensions in terms of association with environmental variables from different subcategories is, in decreasing order, .787±.272 for smoking, .644±.339 for medication, .635±.251 for sun exposure, .626±.183 for physical activity - questionnaire, .565±.288 for alcohol, .558±.209 for diet, .482±.381 for sleep, .267±.514 for electronic devices and .179±.490 for early life factors.

**Figure 6:**
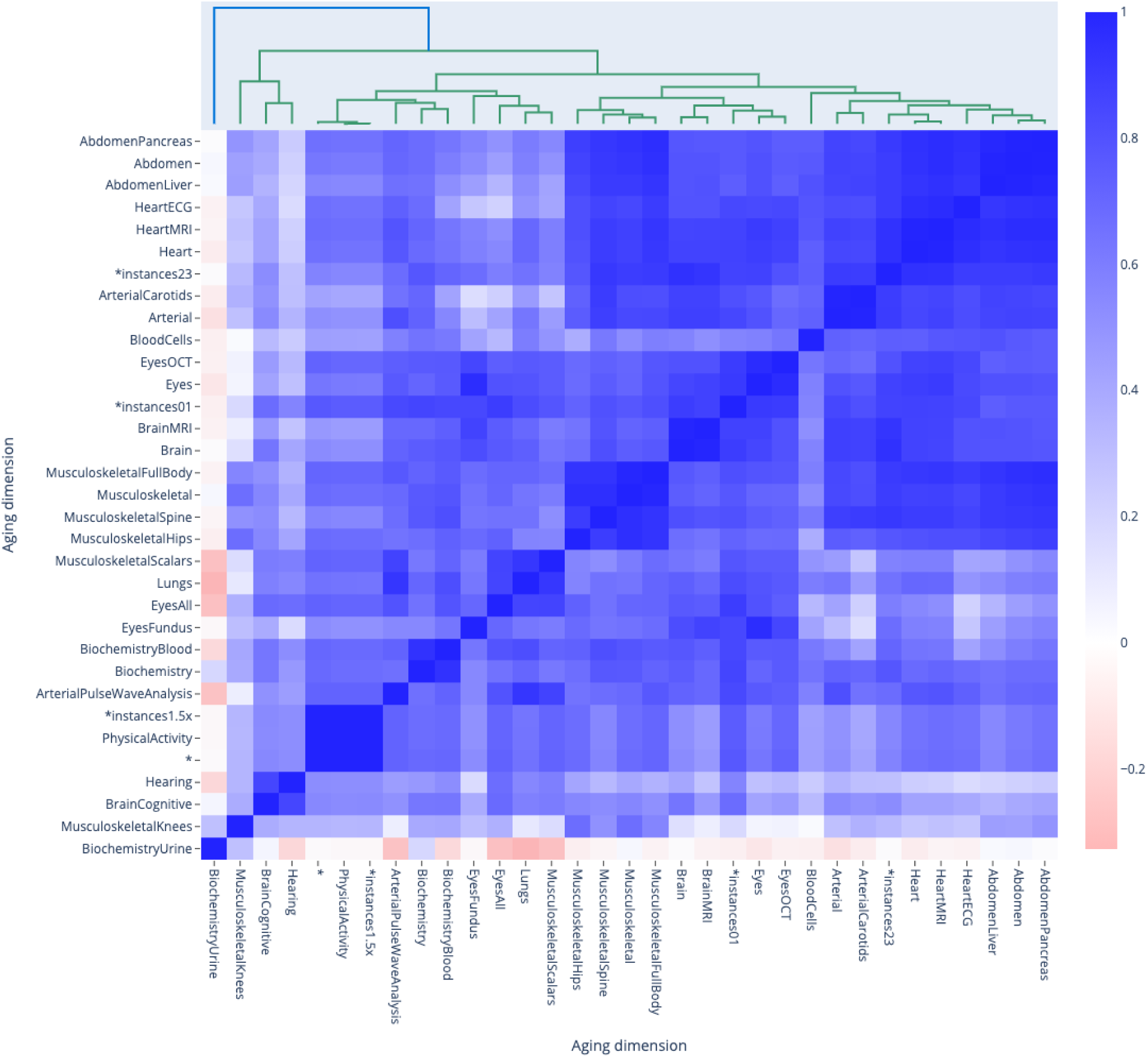
**Correlation between the main aging dimensions and 17 selected aging subdimensions in terms of associated environmental variables** *instances23 and *instances1.5x models denote ensemble models built across several main aging dimensions. For example, *instances23 refers to the model that we built on datasets primarily issued from instances 2 and 3 of the UKB dataset. See Methods for more detail.

##### Socioeconomics

We found that the average correlation between accelerated aging main dimensions and between accelerated aging subdimensions in terms of associated socioeconomic variables are respectively .607±.309 and .639±.421 (Supplementary Figure 10). We found that the average correlation between aging main dimensions in terms of association with socioeconomic variables from different subcategories is, in decreasing order, .903±.151 for “other sociodemographics”, .814±.304 for education, .683±.351 for household, .663±.333 for social support and .609±.334 for employment.

### Performance in predicting chronological age from different aging dimensions

The testing performance obtained on the main biological dimensions and selected subdimensions can be found in Figure 7, and the details of the performances obtained on the different models can be found in Supplementary Figure 11.

**Figure 7:**
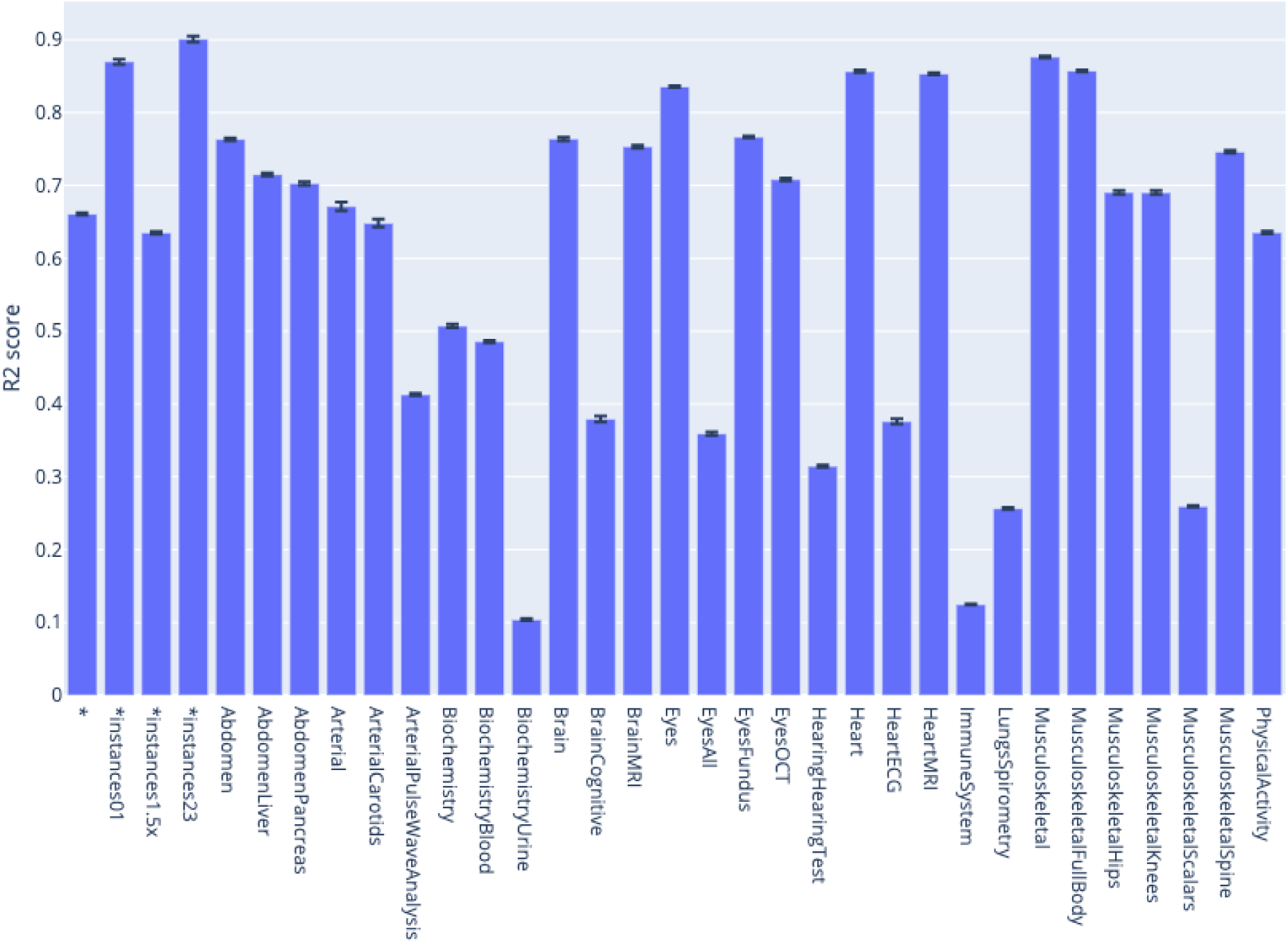
**Chronological age prediction accuracy for the main aging dimensions and selected aging subdimensions** *instances23 and *instances1.5x models denote ensemble models built across several main aging dimensions. For example, *instances23 refers to the model that we built on datasets primarily issued from instances 2 and 3 of the UKB dataset. See Methods for more detail.

We found that the ensemble model built on the data collected during instances two and three of the UKB protocol (focused on collecting medical imaging data) predicted CA with a testing R-Squared [R^2^] of 90.1±0.4% and a root mean squared error [RMSE] of 2.26±0.04 years. Five of the 11 biological dimensions accurately predicted CA with a R^2^ higher than 75%. Musculoskeletal measures (e.g. spine, hips, knees and full body X-ray images) predicted CA with a R^2^ of 87.6%±0.1%, heart data (e.g. MRI videos and ECGs) predicted CA with a R^2^ of 85.3±0.1%, eyes data (e.g. eye fundus and OCT images) predicted CA with a R^2^ of 83.6±0.1%, brain data (functional brain resting MRI and cognitive tests) predicted CA with a R^2^ of 76.4±0.3%, and abdomen images (liver and pancreas MRIs) predicted CA with a R^2^ of 76.3±0.2%. Three of the 11 biological dimensions predicted CA with moderate accuracy with R^2^ values between 50% and 75%. Arterial data (carotid ultrasounds and pulse wave analysis data) predicted age with a R^2^ of 73.3±0.2%, physical activity measurements (wrist accelerometer records) predicted age with a R^2^ of 63.5±0.2%, and biochemistry laboratory values (blood and urine biochemistry) predicted age with a R^2^ of 50.7±0.3%. Finally, three of the 11 biological dimensions poorly predicted CA, with R^2^ values lower than 50%. Hearing tests predicted CA with a R^2^ of 31.4±0.2%, lung function data(spirometry) predicted CA with a R^2^ of 25.6±0.1%, and blood cells data (blood count) predicted age with a R^2^ of 12.5±0.1%.

In total, we report the results for 331 chronological age predictors with an average R^2^ of 41.0%, a standard deviation of 25%, a maximum R^2^ of 90.1% and a minimum R^2^ of -0.1%. The detail of the performance of the different machine learning algorithms and architectures on the 84 aging sub-dimensions, as well as the performance of the ensemble models can be found on the website. We provide more detail, as well as the results of survival prediction in our supplementary.

### Identification of the features driving biological aging predictions

For scalar-based age predictors, we interpreted the models based on the absolute values of the coefficients of the elastic nets, the feature importances of the gradient boosted machines, and the effect on the prediction accuracy of a random permutation between the values of the tested feature for the neural network. The best performing algorithm was most often the GBM (36 out of 41 models), but we also relied on the signed coefficient of the elastic net to identify whether the features selected by the GBM were associated with accelerated young or old age in a linear context. The detailed importance of the features for each scalar data-based model can be found on the website under Model interpretation - Scalar data.

For the models built on time series, images, or videos we used saliency maps to identify the predictive features. For models built on images, we also used the Grad-RAM algorithm to generate a second set of attention maps. For each biological age dimension, we generated ten attention maps samples for the intersection of the three following categories: sex (male and female), age category (younger participants, older participants and participants in the middle of the age distribution), and for different aging rates (accelerated agers, normal agers and decelerated agers). An example of these attention maps can be found in Figure 8, and the rest can be found on the website under Model interpretation - Time series/Images/Videos. We provide a summary of our findings for each aging dimension in the Supplemental.

**Figure 8:**
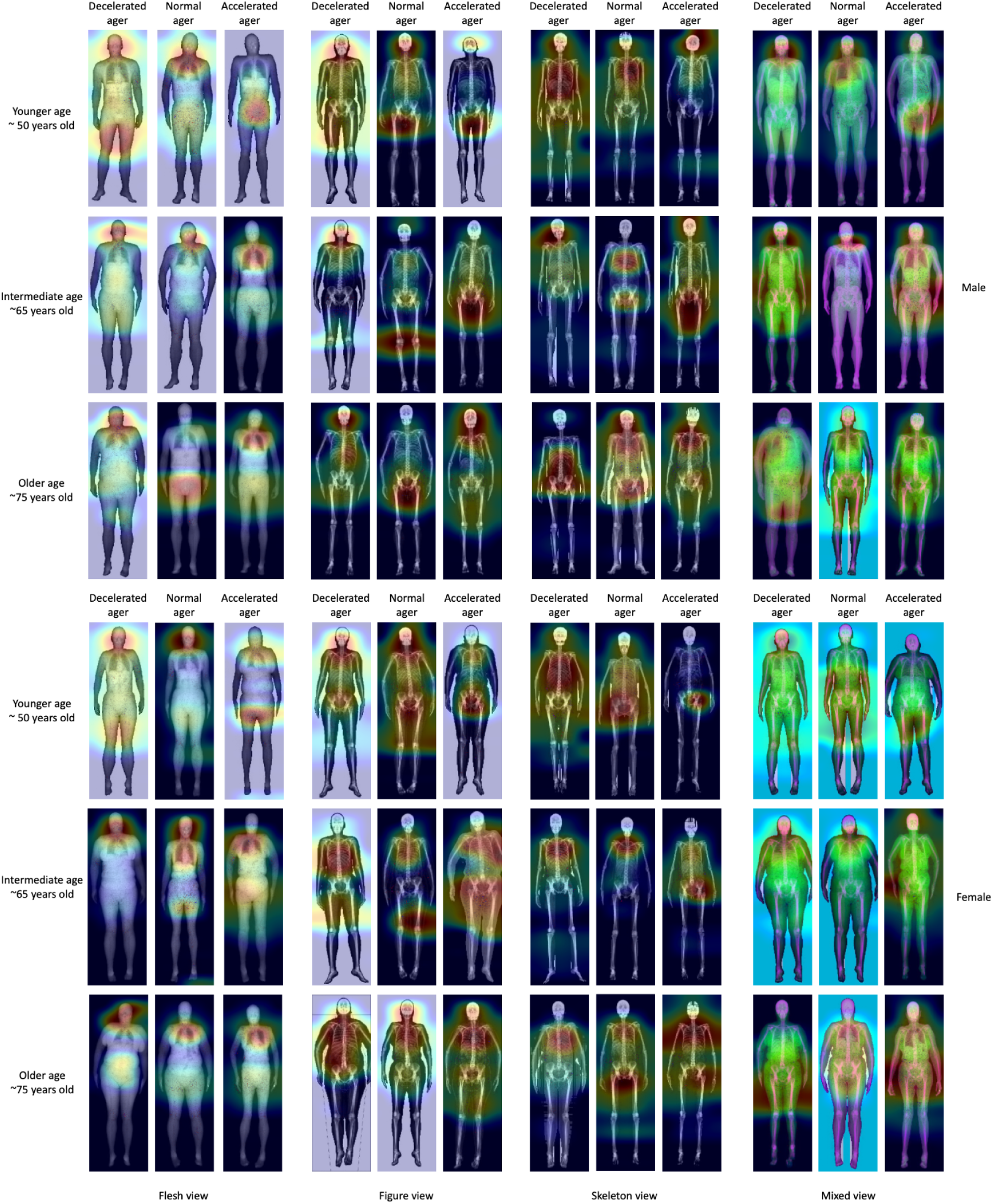
**Attention maps for full body X-ray images** Warm colors highlight regions of high importance according to the Grad-RAM map.

### Heritability and SNPs associated with the different accelerated aging dimensions

We performed 30 genome wide association studies [GWASs] on each of the 11 aging main dimensions, the selected 17 aging subdimensions and two general aging dimensions. We computed the variance of accelerated aging explained by GWAS variants (h2g) for each of the 28 accelerated aging dimensions (Figure 9A). The average GWAS-based heritability (h2g) across the 28 accelerated aging dimensions was 26.1%, and the standard deviation was 7.42%. We found that five main accelerated aging dimensions had h2g greater than 30% (Brain: 35.9±2.6%, Arterial: 32.6±7.3%, Heart: 35.6±3.6%, Musculoskeletal: 34.9±1.7%, Lungs: 30.3±0.2%). Two of the associated accelerated aging subdimensions showed higher GWAS heritability (Brain MRI: 39.1±1.7%, Heart MRI: 37.9±1.9%). In contrast, we found that two main accelerated aging dimensions had h2g less than 15% (Hearing: 14.1±3.9%, Physical activity: 12.0±0.9%). We found that the remaining four main accelerated aging dimensions had h2g between 15% and 30% (Eyes: 28.2±1.2%, Abdomen: 26.3±1.9%, Biochemistry: 25.5±1.0%, Blood cells: 18.1±0.2%).

**Figure 9:**
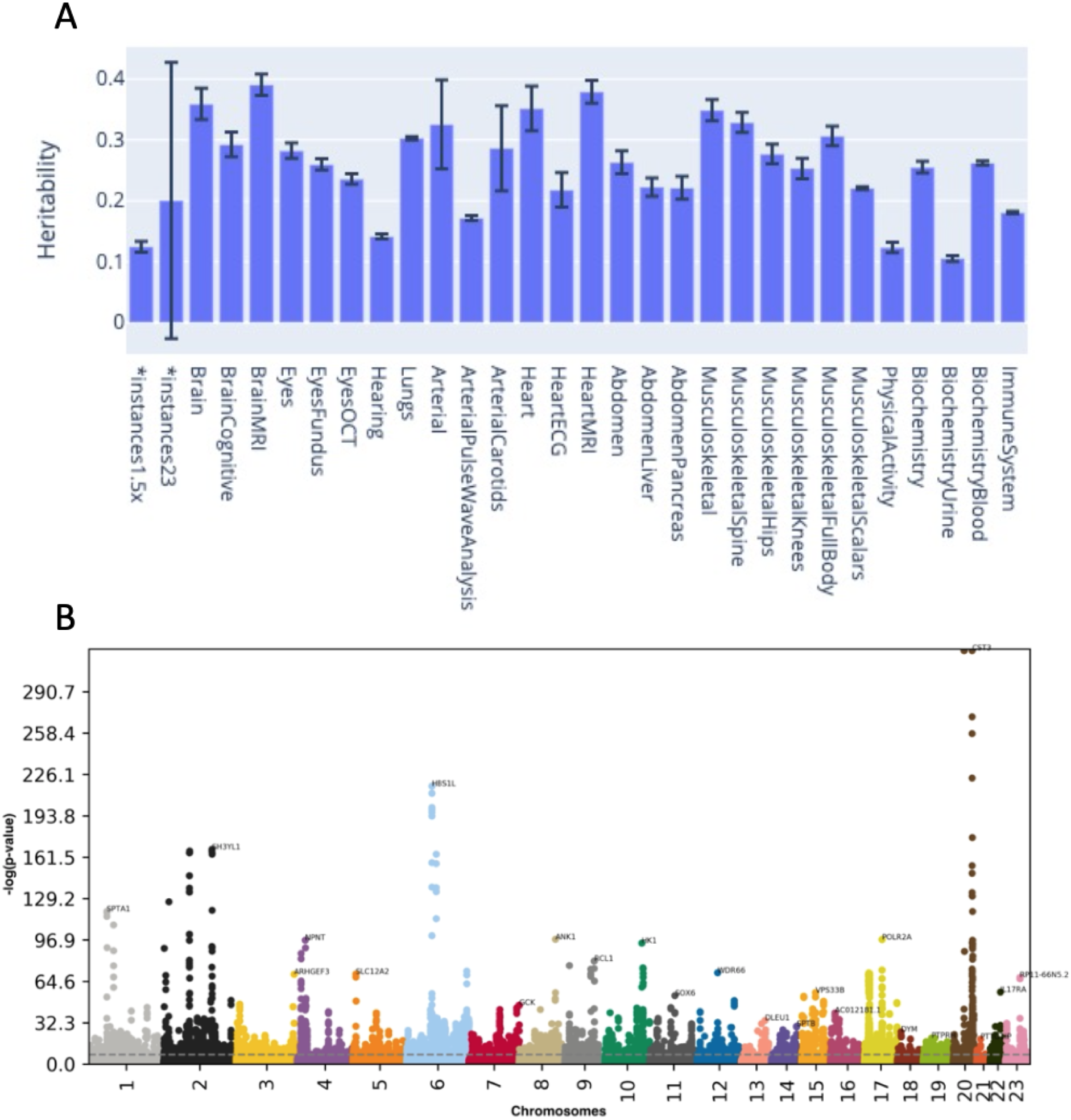
**GWAS Summary** **A - Heritability of accelerated aging for the 28 aging main and sub-dimensions on which a GWAS was performed. B - Union of the GWASs results over the 28 aging main and sub-dimensions on which it was performed.** The x axis shows the 23 human chromosomes, with 23 being the X chromosome. The y axis shows the negative log-p-value for each of the SNPs. Data points above the dotted line represent SNPs that are associated with at least one aging dimension after statistical correction.

We identified 9,697 single nucleotide polymorphisms [SNPs] in 3,318 genes significantly associated (p-value < 5e-8) with accelerated aging in at least one of the 28 dimensions (Figure 9B), with most of these significant associations being driven by datasets with large sample sizes such as lungs (measured with spirometry; sample size: 381,162; 4,286 SNPs) or blood cells (measured with blood count; sample size: 471,001; 2,636 SNPs) (Supplementary Table 5). The Manhattan plots and their associated volcano plots, along with the QQ-plots can be found on the website. We provide a summary of our findings for each of the aging dimensions in the Supplemental.

### Identification of biomarkers, clinical phenotypes, diseases, family history, environmental variables and sociodemographics associated with accelerated aging

As described under Results - Correlations between aging dimensions - Correlation between aging dimensions in terms of association with biomarkers, clinical phenotypes, diseases, family history, environmental variables and socioeconomics, we performed an XWAS to identify non-genetic correlates of accelerated aging in the main again dimensions and selected subdimensions. The full results can interactively be explored on the website. A summary of our findings for each X-category can be found in the Supplemental.

### Predicting accelerated aging from biomarkers, clinical phenotypes, diseases, environmental variables and socioeconomic variables

We predicted accelerated aging in the different aging dimensions using the different X-datasets categories (biomarkers, clinical phenotypes, diseases, environmental variables and socioeconomic variables) and their respective subcategories.

We found that accelerated aging in one aging dimension could not be efficiently predicted from biomarkers from another aging dimension (Supplementary Figure 42). The average testing R^2^ over all pairs of aging dimensions and biomarkers subcategories is 6.2±9.5%.

Similarly, we found that accelerated aging could not be efficiently predicted from clinical phenotypes (Supplementary Figure 43). The average testing R^2^ over all pairs of aging dimensions and clinical phenotype subcategories is 4.8±4.2%.

We found that diseases, environmental variables and socioeconomic variables are poor predictors of accelerated aging in all dimensions, with respective average testing R^2^ values of 4.6±4.1%, 4.9±4.1%, and 4.9±4.2% (Supplementary Figure 44, Supplementary Figure 45 and Supplementary Figure 46).

## Discussion

### Correlations

#### Phenotypic correlation between accelerated aging dimensions

We found that the main aging dimensions are weakly correlated on average (mean correlation .139±.090) but that this correlation tends to increase when the two aging dimensions being correlated are biologically similar (e.g the cognitive sub-subdimensions are .391±0.098 correlated on average). We observed notable exceptions to this general trend. Some accelerated aging dimensions are only averagely correlated despite being biologically similar, such as biochemical blood and urine aging (.090±.004), blood biochemistry and blood count (.151±.002) and brain anatomical and cognitive aging (.113±.007).

We discuss possible reasons for the low average phenotypic correlation in the Supplemental, along with the reasons for the high correlation between accelerated liver and pancreas aging, as well as the importance to correct for the bias in the model’s residuals to define biological aging.

#### Genetic correlation between accelerated aging dimensions

We found that the main aging dimensions are genetically .104±.149 correlated on average and that their associated subdimensions are genetically .338±.255 correlated on average, which shows that biologically similar aging dimensions tend to be more correlated. For example, heart anatomical and electrical aging are .508±.089 correlated, whereas brain cognitive and musculoskeletal aging are uncorrelated (.067±.079). The highest significant genetic correlation was between abdominal liver aging and abdominal pancreas aging. We hypothesize that this larger than average correlation can be explained by the similarity of liver and pancreas abdominal MRI images in terms of the surrounding anatomical features they display and that the aging phenotypes learned from them rely more on the shared surrounding tissue than on the actual organs. Brain anatomical aging and brain cognitive aging are genetically uncorrelated (.019±.056) and similarly for arterial aging and musculoskeletal aging (-.031±.158), lung aging and musculoskeletal aging (-.026±.040), and physical activity aging and musculoskeletal aging (.044±.227). Scalar data-based musculoskeletal aging (anthropometry, impedance, heel bone densitometry and hand grip strength) is also less genetically correlated with the four X-rays-based musculoskeletal agings (full body, spine, hip and knee) than we expected (.126±.098).

We compared the genetic correlations to the phenotypic correlations and found that they were highly correlated (.781 for the correlations between the aging main dimensions and .936 for the aging subdimensions). We claim that this high correlation suggests that the phenotypic correlation between the different accelerated aging dimensions can be partly explained by the genetic correlation between these phenotypes. We observed exceptions. For example, accelerated brain cognitive aging and accelerated hearing aging are significantly more genetically correlated (.743±.070) than they are phenotypically correlated (.351±.005). These genetic correlations could help identify causal links between the different aging dimensions^87^.

#### Correlation between aging dimensions in terms of association with biomarkers, clinical phenotypes, diseases, family history, environmental variables and socioeconomics

We computed the average correlation between main aging dimensions in terms of associated clinical phenotypes (.643±.142), environmental variables (.639±.180), socioeconomics (.607±.309), diseases (.488±.198), biomarkers (.359±.229) and family history (.030±.519). For each X-category, we found that the average correlation between accelerated aging subdimensions is larger than the average correlation between accelerated aging main dimensions, showing that biologically similar aging dimensions tend to be similarly associated with non-genetic factors. The results of the XWAS support both the existence of a general aging process, as well as the multidimensionality of aging, which we discuss below. We discuss the limitations of the XWAS in the Supplemental.

##### XWAS results highlighting general aging factors

We found a large number of non-genetic factors associated with accelerated aging across aging dimensions, suggesting that there are common environmental and disease factors associated (both forward and reverse causality) with all aging dimensions.

For example, we found that general health, and in particular general health rating and suffering from a long-standing illness, disability or infirmity, are two X-variables among the most strongly associated with accelerated aging across aging dimensions (average correlations of respectively .099±.036 and .073±.032)). The causality of this association likely goes both ways: accelerated agers probably suffer from lower general health, since aging is associated with the onset of numerous age-related diseases. Conversely, it is plausible that suffering from long-standing illness is taxing for the body and leads to accelerated aging across most aging dimensions.

This hypothesis is supported by our finding that poor cardiovascular health is associated with accelerated aging across aging dimensions. For example, on average and across aging dimensions, 66.7% of blood pressure biomarkers (e.g. systolic blood pressure, average correlation=.093±.083) and 40.5% of arterial stiffness biomarkers (e.g. position of the shoulder on the pulse waveform, average correlation=.073±.065) are associated with accelerated aging. Similarly, on average and across aging dimensions, 81.8% of breathing clinical phenotypes (e.g. shortness of breath walking on level ground, average correlation=.062±.029) and 59.8% of chest pain clinical phenotypes (e.g. chest pain due to walking ceases when standing still, average correlation=.041±.018) are associated with accelerated aging. Finally, on average and across aging dimensions, the disease category most associated with accelerated aging is cardiovascular disease (e.g. hypertension, average correlation=.074±.031). Others have found that hypertension is associated with MRI-observable changes in the brain^88, 89^. Sun et al. also reported that participants who suffered from hypertension and diabetes had a brain biological age 3.5 years higher on average^90^.

We found that looking younger than one’s actual chronological age is associated with decelerated heart aging, musculoskeletal aging and physical activity aging. Perceived age is an ageing biomarker that is informally but widely used in clinical practice and has been shown to predict survival and to be associated with aging biomarkers such as physical functioning, cognitive functioning and leukocyte telomere length^91–93^. A GWAS was also performed on perceived age found that it is 14% GWAS-based heritable and genetically associated with 75 traits such as adiposity^94^.

Environmental exposure variables, or variables not measured on a GWAS array, tend to be similarly correlated with accelerated aging across aging main dimensions (average correlation=.639±.180). However, it remains elusive how environmental exposure influences individual tissues. We found factors that are likely causal for chronic disease prominently associated with accelerated aging, such as smoking, alcohol consumption, and physical activity. For example, smoking is the environmental subcategory most associated with accelerated aging across aging dimensions and affects not only lungs^95^, but also blood vessels^96, 97^, heart^98–100^, brain^101^, eyes^102, 103^, hearing^104^, abdominal organs^105^ (such as liver^106^, pancreas^107, 108^ and intestines^109, 110^), musculoskeletal system^111^, blood^112^ and urine. Mamoshina et al. showed that smoking is associated with accelerated aging^112^. Similarly, alcohol consumption^113^ affects the brain^114–116^, lungs^117, 118^, blood vessels^119^, heart^119–122^, liver^123–125^, pancreas^126, 127^, bones^128, 129^, muscles^130^, blood^131–133^ and kidneys^134^. There is limited evidence that alcohol consumption affects the eyes in the long term according to the literature^135–137^, but we found significant associations between accelerated eye aging and alcohol consumption variables (e.g. average weekly beer plus cider weekly, correlation=0.031, p-value=1.1e-13, see websites for other examples). Similarly, we found significant associations between accelerated hearing aging and alcohol consumption (e.g. alcohol intake frequency, correlation=0.07, p-value=1.2e-292), despite limited evidence of this phenomenon in the literature^138, 139^. Likewise, physical activity affects the brain, eyes, lungs, blood vessels, heart^140^, abdomen (liver, pancreas), musculoskeletal system, blood and urine. Likewise, sleep quality^141^, physical activity^142^ and diet^143^ affect health across different biological dimensions.

Socioeconomic variables (social support, education, household, employment and other sociodemographics (e.g. receiving an allowance)) tend to be similarly correlated with accelerated aging across aging main dimensions (average correlation=.607±.309). This is coherent with the discrepancies in life-expectancy between socio-economic classes. In the US, the richest 1% males live on average 14.6±0.2 years longer than the poorest 1% males, and the richest 1% females live on average 10.1±0.2 years longer than the poorest 1% females^144^, which suggests that wealth is associated with decelerated aging and poverty is associated with accelerated aging. We found education to be more strongly associated with accelerated/decelerated aging than employment and household, possibly suggesting health literacy^145^ as a key factor to explain the link between socio-economic status and aging rate.

##### XWAS results highlighting the multidimensionality of aging

The average correlation between aging main dimensions for each X-category and X-subcategories is far from being one, and the distribution of X-correlations is wide (large standard deviation). This suggests that, although there are general aging factors associated with accelerated aging in main dimensions as described above, there are also aging dimension specific factors, which affect most strongly a specific aging dimension. For example, hypertension is the disease most significantly associated with accelerated aging across aging dimensions, but accelerated eye aging is “enriched” in associations with eye related diseases such as cataract, retinal disorders and glaucoma, compared to other aging dimensions. Similarly, accelerated knee aging is associated with knee diseases such as knee arthrosis and internal derangement of the knee.

Furthermore, we found that some X-variables are associated with accelerated aging in one aging dimension and associated with decelerated aging in another. For example, playing computer games is associated with accelerated eye aging (correlation=-0.38) and with decelerated brain cognitive aging (correlation=.125). Others have provocatively suggested that screen time can strain the eye (computer vision syndrome)^146^. Further, there is an emerging literature that video games can possibly slow age-related cognitive decline^147, 148^. Similarly, we found that being shorter in one’s youth is significantly associated with accelerated aging in five aging dimensions (eye, hearing, lung, PWA-based arterial and musculoskeletal (scalars biomarkers-based)), whereas being taller in one’s youth is significantly associated with accelerated aging in 11 other aging dimensions (heart anatomical (MRI-based), heart electrical (ECG-based), abdomen, liver, pancreas, musculoskeletal, spine X-rays, hip X-rays, knee X-rays, physical activity and blood cells). Finally, we found discrepancies between aging dimensions in terms of association with weight change over the last year. A stable weight during the last year was exclusively associated with decelerated aging, in six aging dimensions (eye (OCT-based), lung, arterial, musculoskeletal (scalar biomarkers-based)). Gaining weight was mostly associated with decelerated aging (four dimensions: musculoskeletal, full body X-rays, knee X-rays, and urine biochemistry), but also with accelerated aging in two dimensions (lung and arterial (PWA-based)). In contrast, weight loss was mostly associated with accelerated aging (ten dimensions: heart anatomical (MRI-based), abdomen, liver, pancreas, musculoskeletal, full body X-rays, spine X-rays, hip X-rays, knee X-rays and urine biochemistry), but also with decelerated aging in two dimensions (hearing and lung). One plausible explanation for this perhaps surprising observation can be found in the obesity paradox^149^. Alternatively, it is also possible that while being overweight is associated with poor general health, losing a significant amount of weight within a year is often caused by a serious disease. Wannamethee et al. for example reported that, in contrast to intentional weight loss, unintentional weight loss was associated with increased all-cause mortality^150^, which can often be explained by disease^151^.

Finally, we found that accelerated aging in a particular aging dimension could not be successfully predicted by biomarkers from a different dimension, highlighting the multidimensionality of aging. We observed a similar trend when predicting accelerated aging from clinical phenotypes: clinical phenotypes related to an aging dimension could more successfully predict accelerated aging in this dimension than other clinical phenotypes. For example, eyesight clinical phenotypes predicted accelerated eye aging (based on all biomarkers) with a R^2^ of 13.1%, and breathing clinical phenotypes predicted accelerated lung aging with a R^2^ of 10.3%, contrasting with the low average R^2^ (4.8±4.2%) over all aging dimensions and clinical phenotypes categories.

### Performances

#### Complementarity of the aging subdimensions

For some biological dimensions, we found that the different subdimensions contained redundant information about aging. For example, the ensemble model built on full body X-ray images predicted CA with a R^2^ of 85.7±0.1%, but building an ensemble model on full body X-ray images, spine X-ray images (R^2^=74.6±0.2%), hip X-ray images (R^2^ = 69.0±0.3%) and knee X-ray images (R^2^=69.0±0.2%) only improved the R^2^ value by 2.1% (R^2^=87.6%±0.1%). This suggests that the bone X-ray images capture similar facets of aging. Likewise, the ensemble model built on heart MRI images and videos predicted CA with a R^2^ of 85.3±0.1% but building an ensemble model on heart MRIs and on ECGs (R^2^ of 37.6±0.4%) only improved the R^2^ value by 0.3% (R^2^=85.6%±0.2%). The same phenomenon can be observed for the brain. Brain images predicted CA with a R^2^ of 75.3±0.2% but adding cognitive tests (R^2^=37.9±0.4%) to the ensemble only increased the R^2^ value by 1.1% (R^2^=76.4±0.3%). Finally, carotid ultrasound images predicted CA with a R^2^ value of 64.8±0.6%, but adding other artery-based CA predictors such as pulse wave analysis (R^2^=41.3±0.2%) and blood pressure (R^2^=22.3±0.1%) only increased the R^2^ value by 2.3% (R^2^=67.1±0.6% for the arterial model).

In contrast, eye fundus (R^2^=76.6±0.2%) and OCT images (R^2^=70.8±0.2%) along with scalar eyes measures (R^2^=35.9±0.2%) increased the R^2^ value by 7.0% when combined (R^2^=83.6±0.1% for the eye model), suggesting that these three data modalities might capture different facets of aging. Similarly, liver (R^2^=71.5±0.2%) and pancreas (R^2^=70.3±0.3%) MRI images increased the R^2^ value by 5.1% when combined (R^2^=76.3±0.1% for the abdomen model).

#### Comparison between our models and the literature in terms of prediction accuracy Novelty

We are, to our knowledge, the first to build a chronological age predictor on eye OCT images (R^2^=70.1±0.2%; RMSE=4.44±0.01 years), hearing tests (R^2^=31.4±0.2%; RMSE=7.10±0.01 years), spirometry (R^2^=25.6±0.1; RMSE=7.16±0.01 years), heart MRI images and videos (R^2^=85.6±0.2%; RMSE=2.89±0.01 years), liver MRI images, (R^2^=71.5±0.2%; RMSE=4.08±0.01 years), pancreas MRI images (R^2^=70.3±0.3%; RMSE=4.15±0.02 years), spine X-rays (R^2^=74.6±0.2%; RMSE=3.81±0.01 years), musculoskeletal measurements (R^2^=25.9±0.01%; RMSE=7.10±0.01 years) and blood count (R^2^=12.5±0.01%; RMSE=7.59±0.01 years).

We also outperformed the best chronological age predictors in the literature for full body MRIs (R^2^=85.7±0.1%; RMSE=2.85±0.01 years), hip X-rays (R^2^=69.0±0.3%; RMSE=4.20±0.02 years), adult knee X-rays, (R^2^=69.0±0.3%; RMSE=4.20±0.02 years), eye fundus images (R^2^=76.6±0.2%; RMSE=3.97±0.01 years), arterial health (R^2^=67.1%±0.6%; RMSE=4.29±0.04 years), cognitive tests (R^2^=37.9±0.4%; RMSE=5.95±0.02 years) and physical activity (R^2^=63.5±0.2%; RMSE=4.71±0.1 years).

A summary of the comparison between our models and the models reported in the literature can be found in Table 2. We describe and discuss these comparisons more in detail in the Supplemental.

**Table 2:**
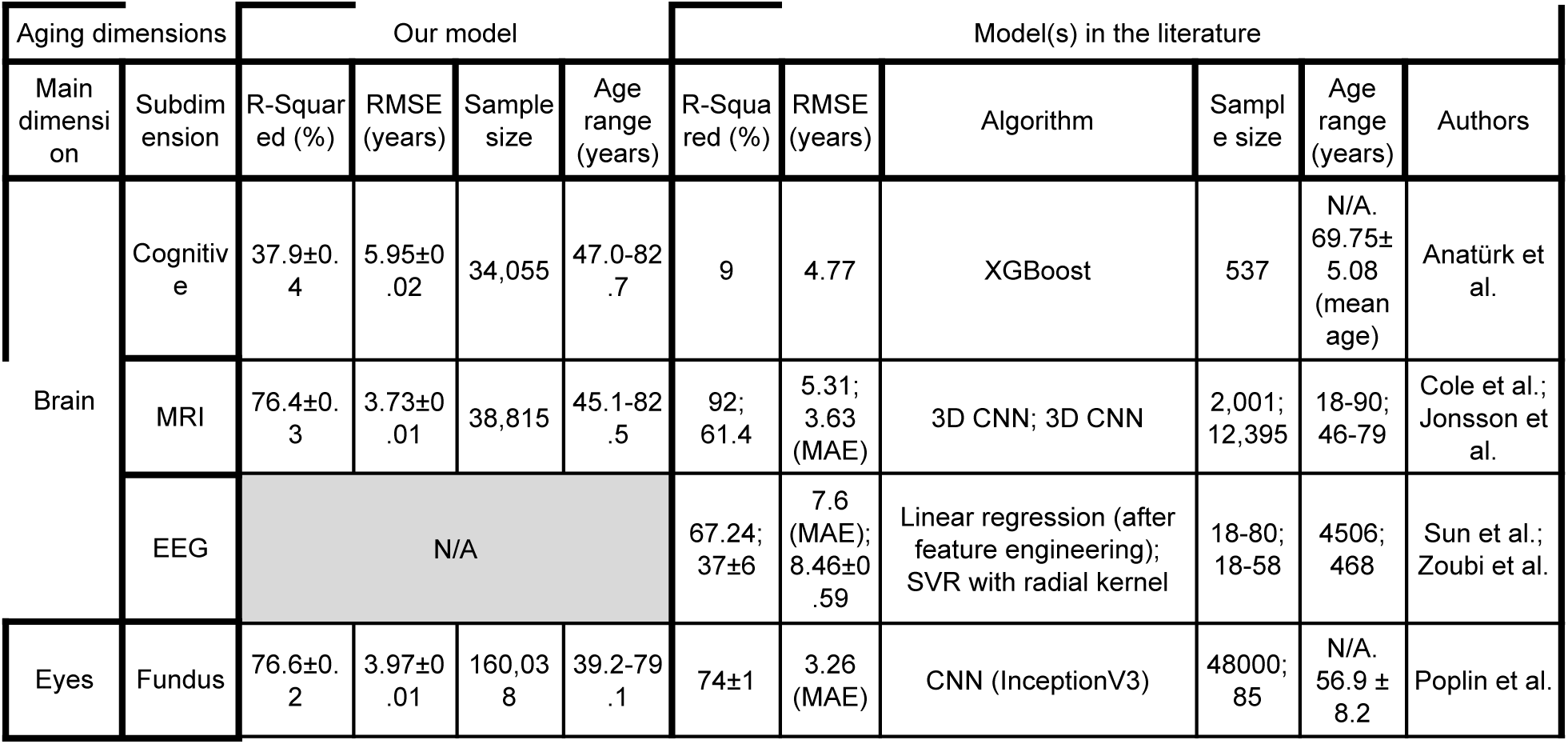
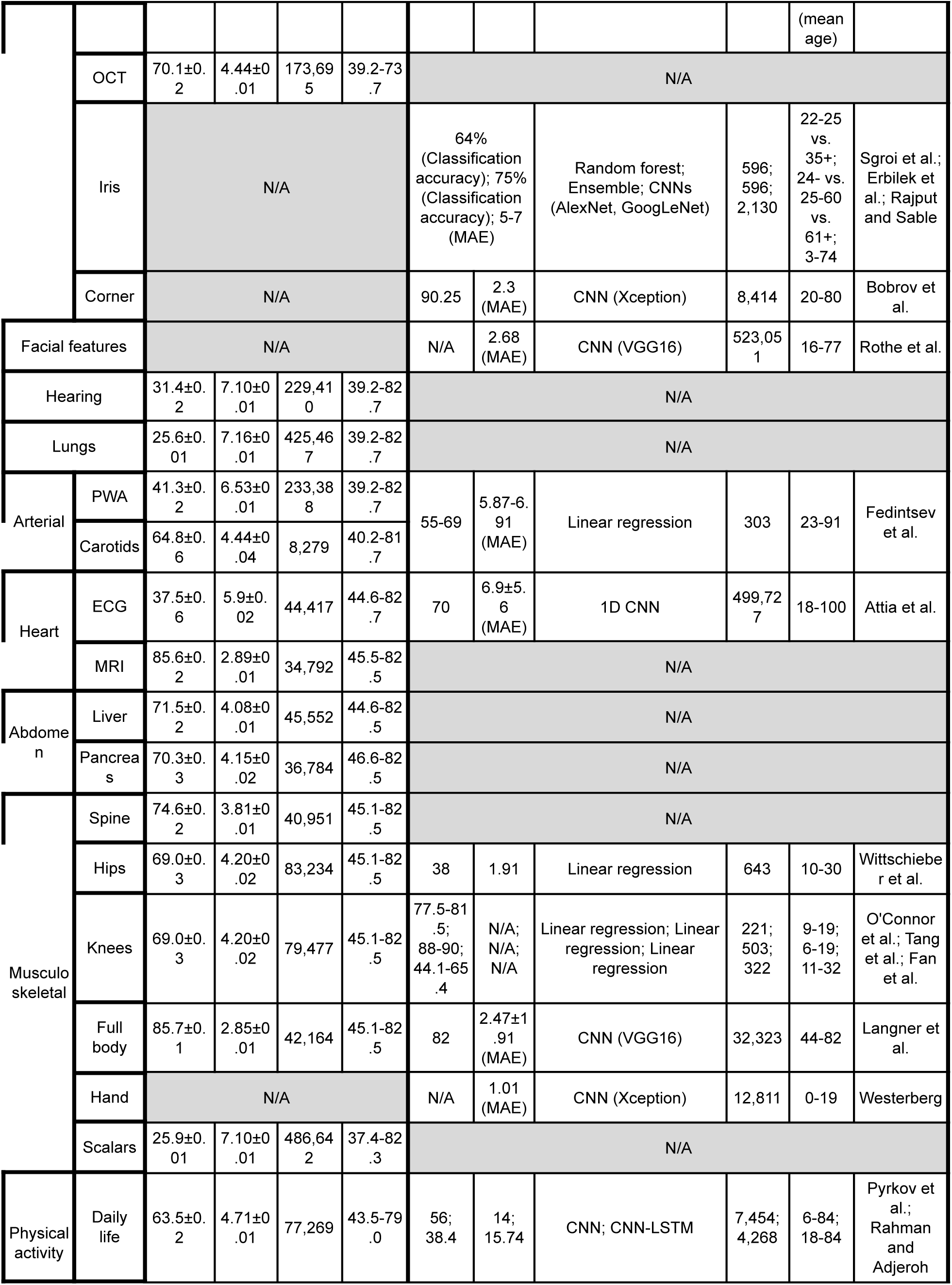
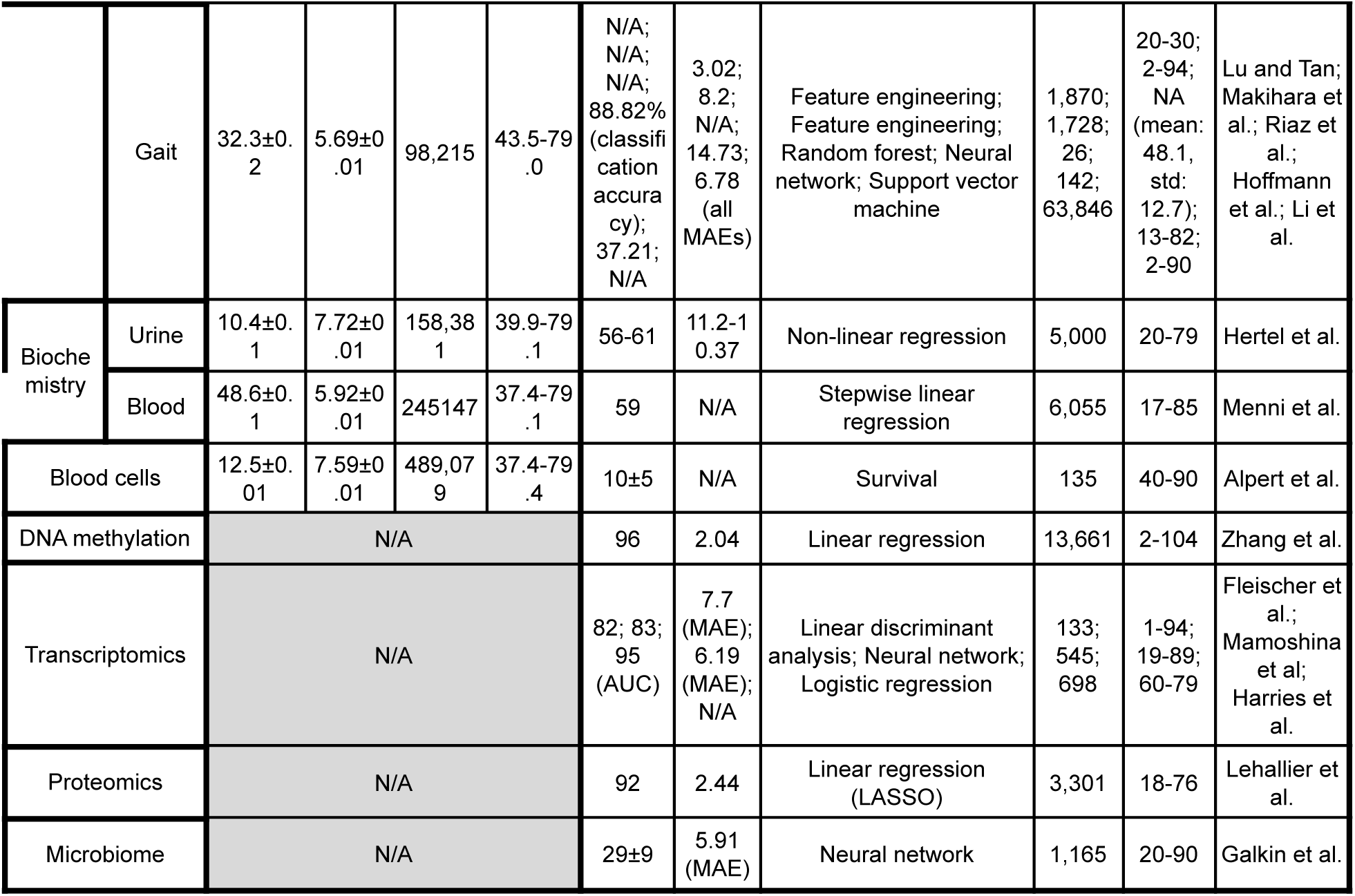
Comparison between our age predictors and the literature in terms of prediction performance.

### Identifying genetic variants associated with aging dimensions: heritability and genes linked to accelerated aging

We found that, on average, 25.6±7.7% of the variance of accelerated aging in the thirty aging dimensions on which we performed a GWAS could be explained by genetic factors. Heritability was as low as 12.3±0.9% for accelerated urine biochemistry aging and as high as 39.1±1.7% for accelerated heart anatomical aging. This difference in heritability was in great part driven by the difference in chronological age prediction performances for the different dimensions (correlation between R^2^ values obtained when predicting chronological age and h2_g values: .664), which suggests that improving the chronological age predictors would lead to an increase of the estimated genetic heritability of the associated accelerated aging dimensions. We discuss our findings in more detail in the Supplemental.

### Strengths and limitations

#### Strengths

We leveraged a large dataset to build what is, to the best of our knowledge, the most exhaustive correlation map between the different aging dimensions. In doing so, we built several new accurate chronological age predictors and outperformed the prediction accuracy of some of the pre-existing predictors. We also performed GWASs and XWASs to estimate not only the phenotypic correlation between the aging dimensions, but also the genetic and the environmental correlations. These GWASs and XWASs allowed us to identify genetic, biological and environmental factors associated with accelerated aging in the different dimensions.

#### Limitations

Despite the breadth of the biomarkers collected by UKB, we were unable to correlate our new biological age definitions to the gold standards such as DNA methylation clock^152^ and telomere length^43^ (among others). Therefore, we are unable to estimate how DNA methylation age would correlate with the other aging dimensions presented in this paper. Other datasets currently unavailable in UKB include proteomics, microbiome and saliva, but they are expected to soon be available.

Some of our predictors predicted chronological age with high accuracy (e.g musculoskeletal features: RMSE=2.65±0.04; heart: RMSE=2.83±0.04), and it has been suggested that, as chronological age prediction accuracy increases, the associated biological age predictors lose their clinical significance^153, 154^. A model that perfectly predicts chronological age only outputs chronological age, not biological age.

For the images-based age predictors, we did not perform image segmentation to isolate the organ of interest from its surrounding tissue. For example, heart MRI videos and images include the full thoracic cavity. As a consequence, and as shown by the attention maps, the predictions are therefore not exclusively driven by the heart, which reduces the interpretability and the utility of the predictor as an estimator of heart health.

Most biological age predictors do not significantly outperform chronological age as survival predictors.

In terms of associations between accelerated aging and non-genetic variables such as environmental exposures, UKB is an observational study, which means that observing these correlations does not allow us to infer causality. Each correlation could potentially be explained by direct causality (e.g not exercising leads to accelerated heart aging), reverse causality (e.g having poor heart function incites limits physical activity) or confounding factors (e.g exposure to a chemical could lead to both decreased heart function and lung function, the later leading to decreased physical activity).

We discuss our future directions in the Supplemental, as well as the potential for biological age predictors to be used to monitor the clinical trials of rejuvenating therapies and to detect new anti-aging target candidates.

## Methods

### Hardware and software

We performed the computation for this project on Harvard Medical School’s compute cluster, with access to both central processing units [CPUs] and general processing units [GPUs] (Tesla-M40, Tesla-K80, Tesla-V100) via a Simple Linux Utility for Resource Management [SLURM] scheduler. We coded the project in Python^155^ and used the following libraries: NumPy^156, 157^, Pandas^158^, Matplotlib^159^, Plotly^160^, Python Imaging Library^161^, SciPy^162–164^, Scikit-learn^165^, LightGBM^166^, XGBoost^167^, Hyperopt^168^, TensorFlow 2^169^, Keras^170^, Keras-vis^171^, iNNvestigate^172^. We used Dash^173^ to code the website on which we shared the results. We set the seed for the os library, the numpy library, the random library and the tensorflow library to zero. For the genetics analysis, we used the BOLT-LMM^174, 175^ and BOLT-REML^176^ softwares. We coded the parallel submission of the jobs in Bash^177^. Our code can be found at https://github.com/alanlegoallec/Multidimensionality_of_Aging.

### Cohort Dataset: Participants of the UK Biobank

For this project, we leveraged the UK Biobank^85^ cohort (project ID: 52887). The UKB cohort consists of data originating from a large biobank collected from 502,211 de-identified participants in the United Kingdom that were aged between 40 years and 74 years at enrollment (starting in 2006). The Harvard internal review board (IRB) deemed the research as non human subjects research(IRB: IRB16-2145).

The initial data collection at enrollment is described as “instance 0”, and some of the participants were followed up with, so more data was collected in later instances (instances 1, 2, and 3). For example, the majority of scalar biomarkers such as blood biomarkers, anthropometrics, hearing test or lung function were collected during instance 0 and sometimes repeated during instance 1. In contrast, the majority of medical images (e.g. brain MRI) were collected during instance 2 and repeated during instance 3. Data was also collected during specific programs, such as the collection of physical activity recorded by wrist accelerometers during one week, which we refer to this paper as instance 1.5x. The instance 1.50 refers to the bulk of the 103,688 participants who took part in the main study, while the instances 1.51, 1.52, 1.53 and 1.54 refer to the four smaller seasonal repeats that involved only 2,633-3,094 of the participants. Because some participants were recruited for more than one instance, the total number of samples available is 676,787, with an age range of 37 to 82 years old. The number of samples available in each instance for each dataset can be found in Table 1.

The UKB dataset contains not only biomarkers (e.g. blood biomarkers, anthropometrics, ECGs, medical images, accelerometer data) but also genetic data^178^, pathologies and environmental data (e.g. socioeconomic status, diet, physical activity). Each scalar variable’s distribution and correlation with age for different demographics can be interactively explored on the website under the tab “Biomarkers” and the subtab “Scalars”.

### Definition of the different aging dimensions

We defined our 11 aging main dimensions by hierarchically grouping the different datasets (Table 1) at three different levels: we call the first level “aging main dimensions”, the second level “aging subdimensions” and the third level “aging sub-subdimensions”. The complete hierarchy between the 11 aging main dimensions, the 31 subdimensions and the 84 sub-subdimensions is described in Table 1. For example, one of the aging main dimensions we investigated was “Heart age”. This main dimension consists of three subdimensions: “ECG”, “MRI”, and “All”. “ECG” encompasses two “sub-subdimensions”: (1) the scalar features (the length of the different intervals, such as PP, PQ and QT), and (2) the time series (the raw 12 leads ECG signal recorded for 600 timesteps). Similarly, “MRI” encompasses 12 sub-subdimensions that include three based on scalar data (e.g., Size, Pulse Wave Analysis), six of them based on MRI images (e.g., 2chambersContrast, and three of them based on MRI videos (e.g. 3chambersRawVideo). Finally, the “All” dimension has a single sub-subdimensions that encompasses all the scalar biomarkers from both the ECG and the MRI subdimensions.

### Data types and Preprocessing

The data preprocessing step is different for the different data modalities: demographic variables, scalar predictors, time series, images and videos. We define scalar predictors as predictors whose information can be encoded in a single number, such as height, as opposed to data with a higher number of dimensions such as time series (one dimension, which is time), images (two dimensions, which are the height and the width of the image) and videos (three dimensions, which are the height and width of the video, along with time). The data extracted and preprocessed from the wrist accelerometer is described separately under “Physical Activity”, as both scalar features, time series and images were generated from it. The preprocessing is described in the Supplemental.

### Machine learning algorithms

For scalar datasets, we used elastic nets, gradient boosted machines [GBMs] and fully connected neural networks. For times series, images and videos we used one-dimensional, two-dimensional, and three-dimensional convolutional neural networks, respectively. We describe these models in detail in the Supplemental.

### Training, tuning and predictions

We split the entire dataset into ten data folds. We then tune the models built on scalar data, on time series, on images and on videos using four different pipelines. For scalar data-based models, we performed a nested-cross validation. For time series-based, images-based and video-based models, we manually tuned some of the hyperparameters before performing a simple cross-validation. We describe the splitting of the data into different folds and the tuning procedures in greater detail in the Supplemental.

### Interpretability of the predictions

To interpret the models, we used the regression coefficients for the elastic nets, the feature importances for the GBMs, a permutation test for the fully connected neural networks, and attention maps (saliency and Grad-RAM) for the convolutional neural networks. We provide more detail in the Supplemental.

### Models ensembling

We built a four-levels hierarchy of ensemble models to improve prediction accuracies. At the lowest level, we combined the predictions from different algorithms on the same aging sub-subdimension. For example, we combined the predictions generated by the elastic net, the gradient boosted machine and the neural network on the brain (dimension) cognitive test (subdimension) “TowerRearranging” (sub-subdimension). At the second lowest level, we combined the predictions from different sub-subdimensions of a unique subdimension. For example, we combined the predictions from the different cognitive tests (e.g. ReactionTime, MatrixPatternCompletion and TowerRearranging) into an ensemble prediction for the “Cognitive” subdimension. At the third level, we combined the predictions from different subdimensions of a unique dimension. For example, we combined the brain subdimensions “All”, “Cognitive” and “MRI” into an ensemble prediction for the dimension “Brain”. Finally, at the fourth and highest level, we combined the predictions from the different aging main dimensions into general chronological age predictions. The ensemble models from the lower levels are hierarchically used as components of the ensemble models of the higher models. For example, the ensemble models built by combining the algorithms at the lowest level for each of the cognitive sub-subdimensions are leveraged when building the “Cognitive” ensemble model at the second lowest level. We provide more detail about the ensembling process in the Supplemental.

### Performance evaluation

We evaluated the performance of the models using two different metrics: R-Squared [R^2^] and root mean squared error [RMSE]. We computed a confidence interval on the performance metrics in two different ways. First, we computed the standard deviation between the different data folds. The test predictions on each of the ten data folds are generated by ten different models, so this measure of standard deviation captures both model variability and the variability in prediction accuracy between samples. Second, we computed the standard deviation by bootstrapping the computation of the performance metrics 1,000 times. This second measure of variation does not capture model variability but evaluates the variance in the prediction accuracy between samples.

### Biological age definition

We defined the biological age of participants as the prediction generated by the model corresponding to the aging main dimension, aging subdimension or aging sub-subdimension, after correcting for the bias in the residuals.

We indeed observed a bias in the residuals. For each model, participants on the older end of the chronological age distribution tend to be predicted younger than they are. Symmetrically, participants on the younger end of the chronological age distribution tend to be predicted older than they are. This bias does not seem to be biologically driven. Rather it seems to be statistically driven, as the same 60-year-old individual will tend to be predicted younger in a cohort with an age range of 60-80 years, and to be predicted older in a cohort with an age range of 60-80. We ran a linear regression on the residuals as a function of age for each model and used it to correct each prediction for this statistical bias.

After defining biological age as the corrected prediction, we defined accelerated aging as the corrected residuals. For example, a 60-year-old whose brain data predicted an age of 70 years old after correction for the bias in the residuals is estimated to have a biological brain age of 70 years, and an accelerated brain aging of ten years.

It is important to understand that this step of correction of the predictions and the residuals takes place after the evaluation of the performance of the models but precedes the analysis of the biological ages properties.

### Survival prediction

UKB collects mortality data from its 502,492 participants, 30,263 of which (6%) are already dead. We highlight the fact that these death events are surprisingly unevenly distributed between data modalities. For example, out of the 207,932 participants for whom pulse wave analysis data was recorded, only 6,000 were already dead when we performed this analysis, half less than the 12,500 death events we would have observed if the death events were evenly distributed between the different datasets.

We leveraged the mortality data to compute the Concordance Index [CI] associated with each of the 331 biological age definitions. The CI measures whether the predictor successfully predicts which participants will die first by computing the percentage of participant pairs for which the participant with the higher biological age dies before the participant with a lower biological age. Accordingly, CI values are usually between 0.5 (useless survival predictor) and 1.0 (perfect survival predictor, at least in terms of ranking the death events). CA is an effective survival predictor, so for each biological age dimension we computed the difference between the CI obtained using biological and the CI obtained using CA to estimate the added value contributed by the biomarkers used to define the biological age dimension. We computed the CA-based control CI separately for each biological age dimension, because CA yields different CI values depending on the age range and distribution of the cohort on which it was used as a predictor. For example, larger CI values can be obtained when the cohort includes a large number of young participants, since it is safe to predict that a young (e.g. 35 years old) participant is unlikely to die before an old (e.g. 80 years old) participant. As the number of such pairs of participants in the cohort increases, obtaining higher CI values using CA as a predictor becomes easier.

We computed the standard deviations for the CI values using the same protocol as the one described under “Methods - Performance evaluation”. We computed an associated two-tailed p-value for each CI difference between the biological age dimension and chronological age assuming a Gaussian distribution and using the associated z-value.

### Correlation between aging dimensions

To compute the correlation between the different aging main dimensions, subdimensions and sub-subdimensions, we computed the Pearson correlation between each pair of accelerated aging phenotypes as defined under Methods - Biological age definition, and we bootstrapped this calculation 1,000 times to obtain confidence intervals.

The average correlation between the 331 accelerated aging phenotypes can be misleading as some correlations are not driven by biology. For example, ensemble models tend to be correlated with the models on which they are built, and the models trained on the same datasets using different algorithms tend to be correlated. To distinguish between the different types of correlations, we hierarchically organized the models into six levels. The first level is for the main biological dimensions, the second level is for the biological subdimensions, the third level is for the biological sub-subdimensions, the fourth level is for the views or features of the same biological dimension, the fifth level is for the image preprocessing methods and the sixth level is for the different algorithms trained on the same dataset.

We describe the computation of the correlations at the different levels in the Supplemental.

### Hierarchical clustering

We hierarchically clustered the different accelerated aging dimensions using complete-linkage clustering and one minus the correlation between two dimensions as the distance, which we did by adapting the dendrogram function from the plotly library^160^.

### Genome-wide association of accelerated aging

The UKB contains genome-wide genetic data for 488,251 of the 502,492 participants^178^ under the hg19/GRCh37 build. We used the v3 imputed genetic data to increase the power of the GWAS, and we corrected all of them for the following covariates: age, sex, ethnicity, the assessment center that the participant attended when their DNA was collected, and the 20 genetic principal components precomputed by the UKB. We used the linkage disequilibrium [LD] scores from the 1,000 Human Genomes Project^179^. To avoid population stratification, we performed our GWAS on individuals with White ethnicity.

Since the DNA of an individual does not significantly change over their lifetime, for each participant and for each aging dimension, we used the average accelerated aging value over the different samples collected over time (see Models ensembling - Generating average predictions for each participant). Next, we performed genome wide association studies [GWASs] to identify single-nucleotide polymorphisms [SNPs] associated with the different accelerated aging dimensions using BOLT-LMM^174, 175^. Finally, we computed the heritability for each of our biological age phenotypes, and we calculated their genetic pairwise correlations using BOLT-REML^176^. We describe these analyzes in further detail below.

We selected the 11 aging main dimensions and 17 out of the 31 aging subdimensions for the GWAS. These 17 subdimensions were selected based on their prediction performance and their scientific interest and are noted with a “*” in Table 1. For example, we selected the two eyes subdimensions that predicted CA with high accuracy (fundus images: R^2^=76.6±0.2%; OCT images: R^2^=70.8±0.2%) and discarded the four subdimensions with low predictive accuracy (all scalar predictors: R^2^=35.9±0.2%; autorefraction: R^2^=29.0±0.2%; visual acuity: R^2^=15.5±0.2%; intraocular pressure: R^2^=6.9±0.1%). For the brain subdimensions, we selected the brain anatomical subdimension (MRI: R^2^=75.3±0.2%) and the functional brain subdimension (cognitive: R^2^=37.9±0.4%) but we did not select the model built on all the scalar predictors (all: R^2^=67.3±0.3%) because of its redundancy, since the predictors included in this model are the union between the scalar predictors included in the first two models. We refer to the 28 selected phenotypes as accelerated aging dimensions in the context of the genetic and environmental analyses.

#### Identification of SNPs associated with accelerated aging

We identified the SNPs associated with each of the 28 selected accelerated aging main dimensions and subdimensions using the BOLT-LMM^174, 175^ software. The sample size for the genotyping of the X chromosome is one thousand samples smaller than for the autosomal chromosomes. We therefore performed two GWASs for each aging dimension. (1) excluding the X chromosome, to leverage the full autosomal sample size when identifying the SNPs on the autosome. (2) including the X chromosome, to identify the SNPs on this sex chromosome. We then concatenated the results from the two GWASs to cover the entire genome, at the exception of the Y chromosome. We used a threshold for significance of 5e-8 for the p-values to correct for the false discovery rate.

We annotated the significant SNPs with their matching genes using the following four steps pipeline. (1) We annotated the SNPs based on the rs number using SNPnexus^180–184^. When the SNPs was between two genes, we annotated it with the nearest gene. (2) We used SNPnexus to annotate the SNPs that did not match during the first step, this time using their genomic coordinates. After these two first steps, 30 out of the 9,697 significant SNPs did not find a match. (3) We annotated these SNPs using LocusZoom^185^. Unlike SNPnexus, LocusZoom does not provide the gene types, so we filled this information with GeneCards^186^. After this third step, four genes were not matched. (4) We used RCSB Protein Data Bank ^187^ to annotate three of the four missing genes. One gene on the X chromosome did not find a match (position 56,640,134).

We plotted the results using a Manhattan plot and a volcano plot. We used the bioinfokit^188^ python package to generate the Manhattan plots. We generated quantile-quantile plots [Q-Q plots] to estimate the p-value inflation as well.

#### Heritability and genetic correlation

We estimated the heritability of the accelerated aging dimensions using the BOLT-REML^176^ software. We included the X chromosome in the analysis and corrected for the same covariates as we did for the GWAS.

Using the same software and parameters, we computed the pairwise genetic correlations between the 28 aging dimensions.

#### Correlation between phenotypic correlation and genetic correlation

We computed the correlation between phenotypic correlations and genetic correlations separately for aging main dimensions and subdimensions because the main dimensions are built on the subdimensions, and some of the main dimensions tend to strongly rely on a single of their subdimensions. For example, heart aging is based on MRI-based anatomical aging and ECG-based electrical aging, but because chronological age is significantly better predicted by MRI images than by ECGs, general heart aging is very similar to heart anatomical aging (correlation: .991±.000). Therefore, the correlations involving heart age are statistically linked to the correlations involving heart anatomical age. To estimate the correlation between phenotypic and genetic correlations, we used sets of aging dimensions for which these statistical artifacts were minimized: the aging main dimensions on one hand, and the aging subdimensions on the other hand.

### Non-genetic correlates of accelerated aging

We identified non-genetically measured (i.e factors not measured on a GWAS array) correlates of each aging dimension, which we classified in six categories: biomarkers, clinical phenotypes, diseases, family history, environmental, and socioeconomic variables. We refer to the union of these association analyses as an X-Wide Association Study [XWAS]. (1) We define as biomarkers the scalar variables measured on the participant, which we initially leveraged to predict age (e.g. blood pressure). We refer to this segment of the XWAS as a Biomarkers Wide Association Study [BWAS]. See Supplementary Table 6 for an exhaustive list of the BWAS variables. (2) We define clinical phenotypes as other biological factors not directly measured on the participant, but instead collected by the questionnaire, and which we did not use to predict chronological age. For example, one of the clinical phenotypes categories is eyesight, which contains variables such as “wears glasses or contact lenses”, which is different from the direct refractive error measurements performed on the patients, which are considered “biomarkers”. We refer to this segment of the XWAS as a Clinical Phenotypes Wide Association Study [CWAS]. See Supplementary Table 11 for an exhaustive list of the CWAS variables. (3) Diseases include the different medical diagnoses categories listed by UKB. We refer to this segment of the XWAS as a Diseases Wide Association Study [DWAS]. See Supplementary Table 16 for an exhaustive list of the DWAS variables. (4) Family history variables include illnesses of family members. We refer to this segment of the XWAS as a Family History Wide Association Study [FWAS]. See Supplementary Table 21 for an exhaustive list of the FWAS variables. (5) Environmental variables include alcohol, diet, electronic devices, medication, sun exposure, early life factors, medication, sun exposure, sleep, smoking, and physical activity variables collected from the questionnaire. We refer to this segment of the XWAS as an Environmental Wide Association Study [EWAS]. See Supplementary Table 24 for an exhaustive list of the EWAS variables. (6) Socioeconomic variables include education, employment, household, social support and other sociodemographics. We refer to this segment of the XWAS as a Socioeconomics Wide Association Study [SWAS]. See Supplementary Table 29 for an exhaustive list of the SWAS variables.

We provide information about the preprocessing of the XWAS in the Supplementary Methods.

#### X-Wide Association Studies

First, we tested for associations in an univariate context by computing the partial correlation between each X-variable and each of the 28 accelerated aging dimensions. To compute the partial correlation between an X-variable and an aging, we followed a three steps process. (1) We ran a linear regression on each of the two variables, using age, sex and ethnicity as predictors. (2) We computed the residuals for the two variables. (3) We computed the correlation between the two residuals and the associated p-value if their intersection had a sample size of at least ten samples. We used a threshold for significance of 0.05 and corrected the p-values for multiple testing using the Bonferroni correction. We plotted the results using a volcano plot. We refer to this pipeline as an X-Wide Association study [XWAS].

For each aging dimension, we list the biomarkers and phenotypes and environmental variables categories that are significantly associated with both accelerated aging and decelerating aging after Bonferroni correction for multiple testing. We order these X-categories by increasing p-value of their most significantly associated representative, and we provide up to three examples by category. If more than three variables were significantly associated with accelerated aging, we only list the first three and precede the list by “e.g”. If the same variable is present in several X-categories (e.g. heart rate can be found in heart function, ECG, pulse wave analysis and blood pressure), we only report it for the first category listed. For the sake of brevity, we shorten the name of the variables when possible and we do not report highly similar features such as features for paired organs (e.g. left arm and right arm impedance) or brain MRI weighted means highlighting the same region (e.g. weighted mean ICVF in tract anterior thalamic radiation and weighted mean OD in tract anterior thalamic radiation are reported as “anterior thalamic radiation”).

#### Prediction of accelerated aging

We leveraged the pipeline we built to predict chronological age as a function of scalar biomarkers (see Methods - Machine learning algorithms - Scalar data; and Methods - Training, tuning and predictions - Scalar data) to predict accelerated aging for the 28 aging dimensions as a function of the biomarkers, the pathologies and the environmental variables. We leveraged the pipeline to identify which features were driving the predictions, as well (see Methods - Training, tuning and predictions - Interpretability of the predictions - Scalar data-based predictors).

We built a model for each X-variables category (Supplementary Table 6, Supplementary Table 11, Supplementary Table 16, Supplementary Table 21, Supplementary Table 24, Supplementary Table 29). In doing so, we encountered the same difficulty as described under Methods - Data types and Preprocessing - Scalar data - Hierarchical clustering of the predictors: taking the intersection of all the variables yielded small sample sizes. To mitigate this issue and find a compromise between the number of variables included in the model and the sample size of their intersection, we leveraged the hierarchical clustering algorithm described under Methods - Data types and Preprocessing - Scalar data - Hierarchical clustering of the predictors.

#### X-Correlations between the accelerated aging dimensions

We estimated the X-correlations between each pair of aging dimensions, for each X-variables category in two different ways. (1) By computing the correlation between the partial correlations generated during the XWAS, that is in a univariate context. (2) By computing the correlation between the feature importances of the models built to predict accelerated aging, that is in a multivariate context.

##### X-Correlations based on the XWAS results

For the sake of clarity, let us walk through an example. Let us say we want to compute the lifestyle correlation between accelerated brain aging and heart aging. The XWAS generates a vector whose components are the partial correlations between the accelerated aging phenotype and each lifestyle variable, for both accelerated aging and heart aging. We compute three different Pearson correlations between these two partial correlation vectors. (1) The “All” correlation, using all the components of the two vectors. This correlation tends to be inflated by the large number of X-variables whose correlation with both accelerated aging dimensions is close to zero. (2) The “Intersection” correlation, using only the lifestyle variables that were significantly associated with both of the accelerated aging dimensions. Because the cardinality of the intersection can be small, a small number of X-variables can yield very high or very low correlations. (3) The “Union” correlation, using only the lifestyle variables that were significantly associated with at least one of the two accelerated aging dimensions. The “Union” correlation represents a compromise between the “All” and the “Intersection” correlations. The figures in this paper were generated using the “Union” correlation, but all three correlations can be explored on the website.

##### X-Correlations based on the feature importances

We then computed the correlations between the feature importances for different accelerated aging dimensions to estimate the X-correlation between the different dimensions. We used the same method as described above under “X-Correlations based on the XWAS results”, replacing the coefficient obtained for each X-variable in a univariate context (using partial correlation with accelerated aging) with the coefficient obtained in a multivariate context (as an accelerated aging predictor in a multivariate model).

## Data Availability

We used the UK Biobank. We will make the phenotypes we generated available upon publication in a peer-reviewed journal.

https://www.multidimensionality-of-aging.net/

https://github.com/alanlegoallec/Multidimensionality_of_Aging

## Abbreviations

UKB: UK Biobank
NHANES: National Health and Nutrition Examination Survey
ECG: electrocardiogram
EEG: Brain electroencephalography
MRI: magnetic resonance image
OCT: optical coherence tomography
DXA: dual-energy X-ray absorptiometry
PWA: pulse wave analysis
CA: chronological age
BA: biological age
AA: accelerated aging
CPU: central processing unit
GPU: graphics processing unit
SLURM: Simple Linux Utility for Resource Management
CV: cross-validation
OCV: outer cross-validation
ICV: inner cross-validation
NCV: nested cross-validation
EN: elastic net
GBM: gradient-boosted machine
NN: neural network
DNN: deep neural network
CNN: convolutional neural network
1D CNN: one-dimensional convolutional neural network
2D CNN: convolutional neural network
3D CNN: convolutional neural network
RNN: recurrent neural network
LSTM: long short-term memory neural network
GRU: gated recurrent units neural network
ReLU: rectified linear unit
SELU: scaled exponential linear unit
CAM: class activation mapping
RAM: regression activation mapping
Grad-CAM: gradient-weighted class activation mapping
Grad-RAM: gradient-weighted regression activation mapping
TPE: Tree-structured Parzen Estimator Approach
R^2^: R-Squared
MSE: mean squared error
RMSE: root mean squared error
MAE: mean absolute error
GWAS: genome wide association study
SNP: single-nucleotide polymorphism
LD: linkage disequilibrium
Q-Q plot: quantile-quantile plot
BWAS: biomarkers wide association study
EWAS: environmental wide association study
PEWAS: phenotypic and environmental wide association study
XWAS: X-wide association study
CVD: cardiovascular disease

## Author Contributions

**Alan Le Goallec:** (1) Designed the project. (2) Supervised the project. (3) Predicted chronological age from images. (4) Computed the attention maps for the images. (4) Generated preliminary results for the chronological age predictors built on ECGs and physical activity. (5) Ensembled the models, evaluated their performance, computed biological ages and estimated the correlation structure between the aging dimensions. (6) Performed the genome wide association studies. (5) Designed the website. (6) Wrote the manuscript.

**Sasha Collin:** (1) Preprocessed the pulse wave analysis and ECG time series. (2) Preprocessed the physical activity data to generate the scalar features, the time series and the images. (3) Predicted chronological age using the time series datasets. (4) Computed the attention maps for the time series. (5) Preprocessed the following images datasets: brain MRIs, eye fundus, eye OCT, carotid ultrasounds, pancreas, full body X-rays, spine X-rays, hip X-rays and knee X-rays.

**Samuel Diai:** (1) Predicted chronological age from scalar features. (2) Coded the algorithm to obtain balanced data folds across the different datasets. (3) Wrote the python class to build an ensemble model using a cross-validated elastic net. (4) Performed the X-wide association study. (5) Built the website.

**Jean-Baptiste Prost:** (1) Preprocessed the datasets for heart MRI videos and liver MRI images. (2) Predicted chronological age from heart MRI videos. (3) Computed the attention maps for videos. (4) Predicted chronological age from spirometry data as time series.

**M’Hamed Jabri:** (1) Generated 147 scalar features based on physical activity time series. (2) Generated preliminary results for the chronological age predictors based on physical activity. (3) Built a preliminary pipeline to generate recurrence plots from physical activity data.

**Théo Vincent:** (1) Improved and polished the website, with a particular emphasis on the XWAS section.

**Chirag J. Patel:** (1) Supervised the project, (2) edited the manuscript, (2) Provided funding.

## Acknowledgments

We would like to thank Dr. Yassine Moustarhfir for helping us interpret the heart MRI images and videos, Raffaele Potami from Harvard Medical School research computing group for helping us utilize O2’s computing resources, and Pauline Dimaano for proofreading the manuscript. We also want to acknowledge UK Biobank for providing us with access to the data they collected. The UK Biobank project number is 52887. We thank HMS RC for computing support.

## Conflicts of Interest

None.

## Funding

NIEHS R00 ES023504

NIEHS R21 ES25052.

NIAID R01 AI127250

NSF 163870

MassCATS, Massachusetts Life Science Center

Sanofi

The funders had no role in the study design or drafting of the manuscript(s).

## Notes

### Competing Interest Statement

The authors have declared no competing interest.

### Author Declarations

IRB#: Harvard University IRB16-2145

