## Supplementary Material for "Analyzing the multidimensionality of biological aging with the tools of deep learning across diverse image-based and physiological indicators yields robust age predictors"

### Supplementary Background, Results, Methods, Figures, Tables and References

#### Table of contents

**Table of contents** 1

**Background** 8

**Results** 10
Prediction performances 10
Comparison between the algorithms trained on scalar features 10
Survival prediction performances 11
Identification of the features driving biological aging 12
Brain 12
Eyes 13
Hearing 14
Spirometry 14
Arteries 15
Heart 16
Abdomen 17
Musculoskeletal 17
Physical activity 18
Biochemistry 18
Blood cells 19
Sex aging rates 19
Heritability and SNPs associated with the different accelerated aging dimensions 20
Abdomen 20
Arteries 20
Biochemistry 21
Brain 24
Eyes 26
Hearing 27
Heart 28
Blood cells 28
Lungs 29

Musculoskeletal 30
Physical activity 33
Identification of biomarkers, clinical phenotypes, diseases, family history, environmental variables and sociodemographics associated with accelerated aging 33
Biomarkers 34
Biomarkers associated with accelerated aging 34
Biomarkers associated with decelerated aging 35
Clinical phenotypes 47
Clinical phenotypes associated with accelerated aging 36
Clinical phenotypes associated with decelerated aging 38
Diseases 39
Diseases associated with accelerated aging 39
Diseases associated with decelerated aging 40
Family history 42
Family history associated with accelerated aging 42
Family history associated with decelerated aging 42
Environmental variables 43
Environmental variables associated with accelerated aging 43
Environmental variables associated with decelerated aging 45
Socioeconomics 47
Socioeconomic variables associated with accelerated aging 47
Socioeconomic variables associated with decelerated aging 49
**Discussion** 51
Correlations 51
Phenotypic correlation between accelerated aging dimensions 51
The low average phenotypic correlation between aging dimensions is partly explained by high model variance and limited prediction accuracy 51
Explaining the high phenotypic correlation between liver and pancreas accelerated aging 53
Correcting for the bias in the residuals prevents an artificial inflation of the correlation between the different aging dimensions 54
Genetic correlation between accelerated aging dimensions 55
Correlation between aging dimensions in terms of association with biomarkers, clinical phenotypes, diseases, family history, environmental variables and socioeconomics 56
XWAS results highlighting general aging factors 56
XWAS results highlighting the multidimensionality of aging 59
Limiting factors of the XWAS 61
Confounding factors 61
Bias due to the sample size 61
Correlation between aging dimensions in terms of association with biomarkers, clinical phenotypes, diseases, family history, environmental variables and socioeconomics 62
Limiting factors of the XWAS 62

Confounding factors 62
Bias due to the sample size 63
Comparison between our models and the literature in terms of prediction accuracy 63
Recommended considerations when comparing the prediction performances between different datasets 63
R2 vs. RMSE 63
Development vs. Aging 65
Comparison with the literature by aging dimension 66
Cognitive tests 66
Brain MRIs 67
Eye fundus images 68
Carotid ultrasounds and pulse wave analysis 68
Electrocardiograms 69
Full body X-rays 70
Hip X-rays 71
Knee X-rays 71
Physical activity: daily life 71
Physical activity: Gait analysis 74
Laboratory biomarkers 75
Blood biochemistry 77
Urine biochemistry 77
Blood cells 77
Chronological age-based organ age predictors 78
Data modalities not covered by UKB 78
DNA methylation 78
Telomere length 79
Transcriptomics 79
Proteomics 80
Microbiome 80
Brain electroencephalography 80
Eye iris and corner images 80
Facial features 81
Hand X-rays 81
Other skeletal structures 82
Physical activity: stress testing 82
Comparison between the models' performance in terms of algorithms and data modalities 82
Algorithms that consider linear vs. non-linear feature representations 82
Raw data vs. pre-processed features 82
Performance of ensemble models 83
Analysis of the bias in the residuals 84

Survival prediction performance 86
Interpretation of the models 88
Analysis of the features selected by the algorithms with respect to the literature 88
Brain 88
Eyes 89
Hearing 90
Lungs 90
Arteries 90
Heart 91
Abdomen 92
Musculoskeletal 93
Biochemistry 93
Blood cells 94
Sex and aging rates 94
Differences in feature importances between the different models 95
Univariate versus multivariate linear models 96
Linear versus non-linear multivariate models 98
Spurious identification of accelerated and decelerated agers due to low quality images 100
The symmetry in the attention maps for paired organs suggest that they age similarly 100
Identifying genetic variants associated with aging dimensions: heritability and genes linked to accelerated aging 101
Missing non-genetic variance 106
Biological age predictors can be used to evaluate candidates rejuvenating therapies and suggest new anti-aging targets 107
Biological age predictors can be used to quantify the effect of rejuvenating drugs or environmental effects 107
Rejuvenating drug candidates 107
Caloric restriction 108
Rapamycin 108
GH and IGF-1 108
Metformin 108
Acarbose 108
NAD+ 109
Sirtuins 109
Resveratrol 109
AFT4 109
Yamanaka factors 110
Young blood serum transfusion 110
17-α-estradiol 110
Lonafarnib 110
Telomerase reactivation 110

Thymus regeneration 111
Hyperbaric oxygen 111
Multidimensionality of rejuvenation 111
Limitations of biological age predictors when assessing rejuvenating therapies 112
Identifying new rejuvenating drug candidates 113
Future directions 113
**Methods** 114
Data types and Preprocessing 114
Demographic variables 114
Scalar data 115
Hierarchical clustering of the predictors to select subcohorts that maximize sample size and predictors 115
Time series 117
Pulse Wave Analysis 117
Electrocardiograms 118
Images 119
Brain MRIs 119
Eye fundus 120
Eyes optical coherence tomography 121
Carotid ultrasounds 122
Heart MRIs 122
Liver and Pancreas MRIs 123
Spine, Hips, Knees and Full body DXA images 123
Physical activity 124
Physical activity as time series 125
Physical activity as scalar features 127
Summary statistics features 127
Activity specific features 128
Sleeping features 128
Physical activity features 129
Weekend features 129
Metabolic Equivalent of Task (MET) features 129
Physical activity as images 130
Videos 132
Heart MRIs 132
Data augmentation 133
Images 133
Videos 133
Machine learning algorithms 134
Scalar data 134

Time series 135
Pulse wave analysis 136
Electrocardiograms 137
Physical activity 138
Architecture for raw acceleration data over the full week 138
Architecture for features extracted from the acceleration data over the full week 140
Architecture for three-dimensional high frequency walking data 140
Images 141
Convolutional Neural Networks Architectures 141
Compiler 142
Videos 143
Underperforming models 144
Training, tuning and predictions 147
Data splitting 147
Scalar data 148
Nested cross-validation 148
Bayesian hyperparameters optimization 148
Example 149
Images 151
Hyperparameters tuning upstream of the cross-validation 151
Cross-validation 154
Cross-validation example 155
Videos 156
Hyperparameters tuning upstream of the cross-validation 156
Time series 157
Hyperparameters tuning upstream of the cross-validation 158
Tuning of the seed using a cross-validation 159
Interpretability of the predictions 159
Scalar data-based predictors 159
Time series, image- and video-based predictors 160
Models ensembling 163
Ensembles hierarchy 163
Ensembling algorithm 164
Generating average predictions for each participant 165
Correlation between aging dimensions 166
Non-genetic correlates of accelerated aging 169
Imputation of the non-genetic X-variables 169
X-Wide Association Studies 170
Prediction of accelerated aging 171
X-Correlations between the accelerated aging dimensions 172

X-Correlations based on the XWAS results 172
X-Correlations based on the feature importances 173

**Supplementary Figures** 174

**Supplementary Tables** 244

**Supplementary references** 272

### Background

Several aging biomarkers such as telomere length[1–9] have already been identified[10–29]. However, although many biomarkers are significantly associated with age, they can only capture a small fraction of CA variance in the population if studied separately[15,30]. Because of this, a common way to define biological age is to train an algorithm (e.g. linear regression[31–35], tree-based method[36–38], neural network[39,40]) to predict an age-related phenotype (e.g. chronological age, survival, onset of an age-related disease) from a set of biomarkers (e.g. blood biomarkers, medical images)[41]. After the model has been trained, it is used to predict the CA of new individuals, and this prediction can be interpreted as the individual's biological age. For example, if the model has successfully learned to predict a participant's CA from their blood sample, it has learned what the typical blood sample of a 30-year-old person looks like. If presented with the blood sample of a 50-year-old whose blood sample results look similar to the average 30-year-old participant, the model will predict the 50-year-old participant to be only 30 years old, which will identify them as a decelerated ager.

A widely recognized and studied biological age predictor is the DNA methylation clock[42–45]. It predicted CA (age-related phenotype) with a linear regression (algorithm) trained on the DNA methylation state of 353 CpG islands (set of biomarkers) with a root mean square error [RMSE] of 3.6 years in 2013[46], and more accurate models were built in the following years[47]. It is associated with mortality[41,48–52], possibly associated with diseases such as cancer, CVD and dementia[41,48], is slowed down by caloric restriction[53] and is partly conserved between mammals[54–56]. Ten single nucleotide polymorphism SNPs have been associated with accelerated DNA methylation aging[57], and resetting the epigenetic clock is being considered as a rejuvenating approach[58].

As machine learning and deep learning gained in popularity, more complex algorithms such as random forests and neural networks predicted CA from transcriptomics[59–61], proteomics[62–64], metabolomics[65,66], blood biomarkers[67–70], microbiome[71–73] and other scalar biomarkers[74,75] with R-Squared values as high as 0.92[62,68]

Convolutional neural networks[76] [CNN] have also been leveraged to predict CA using medical image data, such as brain magnetic resonance imaging [MRI][77–93]. The 3D structure of the brain was processed by a 3D CNN that yielded a R-Squared of 0.92[78]. Brain age predictors appear to be complex phenotypes with a genetic basis. For example, two genetic variants are significantly associated with accelerated brain aging[79]. Brain EEG signals[94], cortical thickness[95] and structure[96], as well as other spatial and temporal features[97–99] and cognitive tests[100], have also been used to predict CA.

Predicting age from body measurements was done as early as 1972[101], but machine learning allowed new models to be built on bone X-rays[102]. For example, full body X-rays predicted age with an R-Squared of 0.83[103] and chest X-ray images predicted age with a R-Squared of 0.91[104]. Work has also been done to predict CA from pelvis X-ray images[105] and pediatric hand X-ray images[106–110].

CNNs were also built on facial images[101,111–128] and specific facial features, such as wrinkles[122], ears[124], eye corners[123] and eye fundus[129], to predict age with mean absolute errors as low as 2.3 years[123] or 3.2 years[129]. A GWAS identified SNPs associated with accelerated perceived age[130].

Progress in machine learning with algorithms, such as recurrent neural networks[131] [RNN], long short-term memory neural networks[132–136] [LSTM] and gated recurrent units neural networks[137] [GRU] also provided researchers with tools to better analyze time series data. Several models were for example built on heart ECGs[111,138,139] with R-Squared values as high as 0.70[138]. LSTM and CNN were also used to predict age from physical activity data[140,141], along with other methods[142,143].

The same methods that were used to predict age were also leveraged to predict other age-related phenotypes, such as survival and age-related diseases. For example, eye fundus images were used to predict cardiovascular risk factors[144] and mortality was predicted using chest X-ray images[145], heart MRI videos [146], heart ECGs[147,148], physical activity [141,149] microbiome data[150] as well as other biomarkers[151–169].

### Results

#### Prediction performances

##### Comparison between the algorithms trained on scalar features

Out of the 41 models trained on scalar features, we found that the GBM performed the best for 36 models (88%), the neural network performed the best for four models (10%; brain gray matter volumes, brain MRI weighted means, brain MRI all biomarkers, heart pulse wave analysis biomarkers) and the elastic net performed the best for a single model (2%; all brain biomarkers). The average prediction performances were 20.2±16.6% for the elastic net, 24.9±16.7% for the GBM and 23.4±16.2% for the neural network. A summary table comparing the performance of the three algorithms can be found in Supplementary Table 2.

#### Survival prediction performances

For each biological age definition, we tested whether it performed better than chronological age as a survival predictor. The mean Concordance Index [CI] difference between biological age and chronological age was small (mean: 0.003; standard deviation 0.020) but significant (paired t-test p-value: 0.02). The p-value might have been inflated by the correlation structure between the 331 biological age dimensions, so we computed the same statistics while limiting ourselves to the 11 main aging dimensions. The mean CI difference between biological age and chronological age was 0.015, the standard deviation was 0.020 and the paired t-test p-value was 0.03, so the difference was still statistically significant.

We found that 197 out of the 331 biological age dimensions outperformed chronological age as survival predictors and that 134 underperformed compared to CA. Out of the 197 dimensions that outperformed CA, 38 were significantly (p-value < 0.05) different from CA, and 18 were still significant after Bonferroni correction (based on the total 331 models). Out of the 134 dimensions that underperformed compared to CA, five were significantly different from CA, and none were significantly different from CA after Bonferroni correction. Between the 18 dimensions that significantly outperformed CA as a survival predictor after Bonferroni correction, the one obtained by training an elastic net on biochemistry biomarkers (both blood and urine) outperformed CA the most, with a CI improvement of 0.028±0.007 (CI: biological age=0.708; CA=0.679). Similarly, we found that nine out of the 11 main dimensions outperformed chronological age in terms of CI and two underperformed. Out of the nine dimensions that outperformed CA, two were significantly different from CA, both before and after Bonferroni correction. Neither of the two dimensions that underperformed compared to CA were significantly different from CA. Between the two main dimensions that significantly outperformed CA as a survival predictor after Bonferroni correction, lung aging outperformed CA the most with a CI improvement of 0.017±0.003 (CI: lung age=0.700; CA=0.683).

#### Identification of the features driving biological aging

##### Brain

We best predicted brain cognitive age with a gradient boosted machine that gave a $R^2$ of 37.8±1.2% ($R^2$=37.9±0.3% for the ensemble model based on all cognitive tests). The model was trained on 135 biological features, along with sex and ethnicity. We found that the nine most predictive scalar features are all related to speed of answering questions and include, in order: (1) time to answer, (2) mean time to correctly identify matches, (3) duration to complete alphanumeric path (trail #2), (4) time to complete test, (5) duration to complete numeric path (trail #1), (6) time to complete round (second round), (7) time to complete round (first round), (8) mean time to solve puzzles and (9) mean time to press snap-button. These nine features were associated with older age based on their elastic net ($R^2$=33.0) regression coefficients. The tenth most important feature selected by the GBM was the fluid intelligence score, which is negatively associated with aging based on the elastic net regression coefficient. The fluid intelligence score (UKB field 20,016) is the number of correct answers to 13 questions: numerical addition test, identify largest number, word interpolation, positional arithmetic, family relationship calculation, conditional arithmetic, synonym, chained arithmetic, concept interpolation, arithmetic sequence recognition, antonym, square sequence recognition and subset inclusion logic.

In terms of scalar features, we best predicted brain anatomical aging using a neural network ($R^2$=62.7±1.7%). The elastic net ($R^2$=62.2±1.1%) and the GBM ($R^2$=62.0±1.0%) performed similarly well. The models were trained on 396 biological features, along with sex and ethnicity.

The neural network highlighted (1) the forceps minor, (2) the forceps major, (3; 4) the right superior longitudinal fasciculus, (5; 7) the left acoustic radiation, (6) the right inferior longitudinal fasciculus, (8) the left corticospinal tract and (9; 10) the right and left inferior fronto-occipital fasciculi. The GBM highlighted (1) the volume of gray matter in the thalamus (right side), (2; 10) the posterior thalamic radiations (right and left sides), (3) the volume of gray matter in the ventral striatum (left side), (4) the forceps minor, (5) the superior thalamic radiation (right side), (6) the corticospinal tract (left side), (7) the subcallosal cortex (right side), (8) the volume of the putamen (left side) and (9) the volume of gray matter in the insular cortex (right side). Based on the elastic net regression coefficients, the five features most associated with old age are (1) the anterior thalamic radiation, (2) the acoustic radiation, (3) the inferior longitudinal fasciculus, (4) the volume of gray matter in the subcallosal cortex and (5) the posterior thalamic radiation (all on the right side). The five features most associated with young age are (1) the acoustic radiation (left side), (2) the corticospinal tract (left side), (3) the forceps minor, (4) the ventral striatum (left side) and (5) the middle cerebellar peduncle.

In terms of brain MRI images, the attention maps highlighted different features across participants (Supplementary Figure 10).

#### Eyes

In terms of scalar features, we best predicted eye aging with a GBM ($R^2$=35.8±0.6%) trained on autorefraction (35 features), acuity (eight features) and intraocular pressure (eight features) measures, along with sex and ethnicity. Autorefraction measures alone predicted CA with a $R^2$ of 29.0±0.2%, acuity measures with a $R^2$ of 15.5±0.2% and intraocular pressure with a $R^2$ of 6.9±0.1%. Specifically, the associated most important scalar features for the model including all the predictors are (1; 2) the spherical power (right and left eyes), (3; 4) the cylindrical power (right and left eyes), (5; 6) the astigmatism (right and left eyes), (7) the 3mm asymmetry index (left eye),

(8) the 6mm cylindrical power (right eye), (9) the 3mm asymmetry angle (left eye) and (10) the 6mm weak meridian angle. In the linear context of the elastic net ($R^2$=25.0±0.4%), the spherical power, the cylindrical power, the 6mm cylindrical power and the 6mm weak meridian angle are associated with older age, whereas the astigmatism angle, the 3mm asymmetry index and the 3mm asymmetry angle are associated with younger age.

In terms of eye fundus images, the Grad-RAM attention maps tended to highlight the eye regions that were highly vascularized, whereas the saliency maps also frequently highlighted the center of the eye (Supplementary Figure 11). In terms of OCT images, Grad-RAM highlighted all the retinal layers with different regions emphasized for different participants. Similarly, saliency maps highlighted different layers for different patients, as well as the fovea. The resolution of the resized images makes it difficult to precisely identify these layers, but the list seems to include the posterior hyaloid surface, the inner limiting membrane, the external limiting membrane, ellipsoid portion of the inner segments, the cones outer segment tips line and the retinal pigment epithelium (Supplementary Figure 12).

#### Hearing

We best predicted hearing age with a GBM ($R^2$=31.1±0.9%) trained on 126 hearing test features, along with sex and ethnicity. We found that the most important scalar features are the duration of the hearing test and the time to press next for the first tests, which all had a positive coefficient in the elastic net ($R^2$=25.5±0.7%).

#### Spirometry

The spirometry dataset used to predict lung aging only included three features (peak expiratory flow, forced vital capacity and forced expiratory volume in one second) which were selected in this order by the GBM ($R^2$=25.6±0.5%). All three of these features had a negative regression coefficient for the elastic net ($R^2$=25.5±0.4%).

#### Arteries

In terms of scalar features, we best predicted arterial age with a GBM ($R^2$=36.8±1.4%) trained on a combination of blood pressure measures (three features), pulse wave analysis features (eight features) and carotid ultrasound features (12 features). We found that the most important scalar features are (1) pulse wave peak to peak time, (2) pulse wave arterial stiffness index, (3) systolic blood pressure, (4) diastolic blood pressure, (5) pulse rate, (6) pulse wave reflection index, (7) mean carotid intima-medial thickness [IMT] at 120 degrees, (8) mean carotid IMT at 240 degrees, (9) position of pulse wave notch and (10) mean carotid IMT at 210 degrees. For the elastic net ($R^2$=31.3±1.3%), pulse wave peak to peak time, systolic blood pressure, mean carotid IMT at 240 degrees have a positive regression coefficient. In contrast, pulse wave arterial stiffness index, diastolic blood pressure pulse rate, pulse wave reflection index and position of the pulse wave notch have a negative regression coefficient.

In terms of carotid ultrasound images, the attention maps highlighted different features based on the carotid view. For the short axis view, Grad-RAM mostly highlighted the skin tissue whereas saliency maps tended to also highlight the artery or jugular vein. For the long axis view, the saliency maps mostly highlighted the artery, and Grad-RAM highlighted both the artery, the skin tissue and some of the surrounding tissues. The CIMT120, CIMT150 and Mixed views are similar to the long axis view. (Supplementary Figure 13)

A sample of the attention maps for pulse wave analysis data can be found in Supplementary Figure 14, and more are available on the website.

#### Heart

In terms of scalar features, we best predicted both general cardiac aging, anatomical cardiac aging and electrical cardiac aging with a GBM (general: $R^2$=31.7±2.3%; anatomical: $R^2$=26.7±1.8%, electrical: $R^2$=11.1±1.9%). We predicted general heart aging from 23 MRI features and 12 ECG features, along with sex and ethnicity. We found that the most important scalar features to predict general cardiac aging are (1) mean arterial pressure, (2) diastolic blood pressure, (3) sex of the participant, (4) body surface area, (5) R axis, (6) PWA augmentation index, (7) end systolic pressure index, (8) PQ interval length, (9) P duration and (10) central systolic blood pressure. The most important scalar features to predict anatomical cardiac aging are the mean arterial pressure, the systolic pressure, and the diastolic pressure. Finally, the most important scalar features to predict electrical cardiac aging are the rotation of the R axis, the rotation of the T axis and the length of the QTC interval. The elastic net predicted CA with a $R^2$ of 25.9±2.7%. It assigned a positive regression coefficient to diastolic blood pressure, sex, augmentation index, PQ interval and central systolic pressure, and a negative regression coefficient to mean arterial pressure, body surface area, R axis, end systolic pressure index and P duration.

In terms of heart MRI images, the anatomical features highlighted by the attention maps differed between views (Supplementary Figure 15 and Supplementary Figure 16). For the two chamber view, which is close to a sagittal view, Grad-RAM highlighted most of the thoracic cavity, with a particular emphasis on the spine. Three-chamber and four-chamber views of the heart also highlighted extra-cardiac features in the thoracic cavity, but tended to highlight cardiac features more consistently than the two-chamber views. These features included the interventricular septum, the mitral and tricuspid valves and the aorta. In terms of heart MRI videos, the saliency map highlighted the same features more clearly, and with a lesser emphasis on extra-cardiac features (Supplementary Figure 17 and Supplementary Figure 18).

A sample of the attention maps for the ECGs can be found in Supplementary Figure 19, and more can be found on the website.

#### Abdomen

For liver MRI-based models, attention maps highlighted the liver along with other abdominal structures such as the stomach, the spleen, muscle and adipose tissue (Supplementary Figure 20). Similarly, for pancreas MRI-based models, attention maps did not particularly focus on the pancreas but instead highlighted diverse abdominal regions across patients, including the liver (Supplementary Figure 21).

#### Musculoskeletal

In terms of scalar features, we best predicted accelerated musculoskeletal aging using a GBM or a neural network (25.2±0.4%). Impedance measures (five features) and heel bone densitometry (six features) were poor predictors of chronological age, with respective $R^2$ values of 4.2±0.2% and 4.9±0.2%. Prediction accuracy was driven by anthropometric measures ($R^2$=15.1±0.3%; eight features) and hand grip strength ($R^2$=13.1±0.3%; two features). Specifically, the most important features were (1) White ethnicity, (2) waist circumference, (3) weight, (4) impedance of whole body, (5) British ethnicity, (6) body mass index [BMI], (7) left hand grip strength, (8) Sex, (9) right arm impedance and (10) Indian ethnicity. The elastic net ($R^2$=20.5±0.4%) assigned a positive regression coefficient to White ethnicity, waist circumference, and British ethnicity and a negative regression coefficient to weight, impedance of whole body, BMI, left hand grip strength and impedance of right arm.

Attention maps for full body X-ray images highlighted diverse body parts across participants. Regions frequently highlighted included the neck, the upper torso, the hips and the knees (Figure 8). Attention maps for sagittal view spine X-ray images most commonly highlighted the lumbar region (Supplementary Figure 22), whereas the coronal view images highlighted diverse vertebras across individuals (Supplementary Figure 23). Attention maps for hip X-rays consistently highlighted the greater trochanter of the femur as well as the joint (Supplementary Figure 24). Attention maps for knee X-rays highlighted diverse regions across participants, such as the femur, the shin, the joint itself and occasionally the fibula (Supplementary Figure 25).

#### Physical activity

In terms of scalar features, we best predicted accelerated "physical activity" aging using a GBM ($R^2$=59.4±0.6%) trained on 2,271 features, along with sex and ethnicity. The most predictive scalar features are features extracted from the UKB preprocessing software (e.g. fft8_std, fft12_std, fft8_75%). In terms of images (recurrence plots, Markov transition plots and Gramian angular field plots), sample attention maps can be found in Supplementary Figure 26, and more can be found on the website. Similarly, sample attention maps for the models built on physical activity data as time series can be found in Supplementary Figure 27 (raw acceleration data) and Supplementary Figure 28 (high intensity 3D acceleration data during walking).

#### Biochemistry

We best predicted accelerated biochemical aging using a GBM ($R^2$=49.1±0.7%), and this prediction was largely driven by blood biomarkers (blood: $R^2$=47.6±0.5%; urine: $R^2$=10.4±0.3%). The model was trained on 28 blood features and four urine features, along with sex and ethnicity.

Specifically, the most important features were (1) testosterone, (2) sex hormone binding globulin (SHBG), (3) alanine aminotransferase, (4) glycated hemoglobin (HbA1c), (5) urine enzymatic creatinine, (6) cystatin C, (7) apolipoprotein B, (8) creatinine, (9) insulin-like growth factor 1 [IGF-1] and (10) urea. The elastic net ($R^2$=31.5±1.0%) assigned a positive regression coefficient to SHBG, HbA1c, cystatin C, apolipoprotein B and urea, and a negative regression coefficient to testosterone, alanine aminotransferase, urine enzymatic creatinine, creatinine, IGF-1.

#### Blood cells

We best predicted accelerated blood cells aging with a GBM ($R^2$=12.2±0.2%) trained on 31 features, along with sex and ethnicity. The most predictive scalar features are (1) red blood cell distribution width, (2) red blood cell count, (3) mean corpuscular volume, (4) mean sphered volume, (5) platelet crit, (6) lymphocyte percentage, (7) mean reticulocyte volume, (8) neutrophil count, (9) hemoglobin concentration and (10) mean corpuscular hemoglobin concentration. The elastic net ($R^2$=5.8±0.4%) assigned a positive regression coefficient to red blood cell distribution width, mean corpuscular volume, mean sphered cell volume, mean reticulocyte volume, neutrophil count and mean corpuscular hemoglobin concentration and a negative regression coefficient to red blood cell count, platelet crit, lymphocyte percentage and hemoglobin concentration.

#### Sex aging rates

All the elastic nets models that we trained to predict chronological age assigned a positive regression coefficient to the sex for feature, with the one exception of the model built on blood cell features.

### Heritability and SNPs associated with the different accelerated aging dimensions

#### Abdomen

We found that accelerated abdominal (liver and pancreas MRIs based) aging is 26.3±1.9% heritable, and that one SNP is significantly associated with this phenotype: PNPT1. PNPT1 is a RNA-binding protein involved in RNA metabolic processes[170].

We found that accelerated liver aging is heritable at 22.3±1.5% and that seven SNPs in six genes are significantly associated with this phenotype. The four peaks on the Manhattan plot highlight: (1) PNPT1 (Polyribonucleotide Nucleotidyltransferase 1, involved in RNA metabolic processes); (2) RP11-998D10.4 (a long intergenic non-coding RNA gene with unknown functions). Other genes in linkage disequilibrium with RP11-998D10.4 include ARHGEF40 (a protein coding gene for a guanine nucleotide exchange factor [GEF] possibly involved in G-protein-coupled receptors [GPCR] signaling[170]) and ZNF219 (a zinc finger protein that modulates chromatin's transcriptional activation[170]); (3) TSEN2 (TRNA Splicing Endonuclease Subunit 2, involved in RNA splicing to remove introns[170]); and (4) RN7SL793P (RNA, 7SL, Cytoplasmic 793, Pseudogene, a pseudogene with unknown function).

Accelerated pancreas aging is heritable at 22.1±1.9% but we did not find any SNP significantly associated with it.

#### Arteries

Accelerated arterial aging is heritable at 32.6±7.3% but we did not find any SNPs significantly associated with it.

Similarly, we found that arterial carotid aging (based on ultrasound images) is heritable at 28.6±7.0%, but we did not find any SNPs significantly associated with it.

We found that arterial pulse wave analysis aging is heritable at 17.2±0.4% and that 192 SNPs in 109 genes are significantly associated with this phenotype. The ten highest GWAS peaks are: (1) AC073115.6 (a long intergenic non-coding RNA). AC073115.6 is in linkage disequilibrium with CDK6 (Cyclin Dependent Kinase 6, involved in cell cycle regulation); (2) CNNM2 (Cyclin and CBS Domain Divalent Metal Cation Transport Mediator 2, involved in magnesium homeostasis); (3) IGFBP3 (Insulin Like Growth Factor Binding Protein 3, a growth factor regulator); (4) TEX41 (a long non-coding RNA linked to aortic valve disease 1); (5) HMGA2 (High Mobility Group AT-Hook 2, a transcription regulator); (6) ATP1B3 (ATPase Na+/K+ Transporting Subunit Beta 3, involved in maintaining Na and K ions' electrochemical gradients and in cell-surface interactions at the vascular wall); (7) MTND3P1 (Mitochondrially Encoded NADH:Ubiquinone Oxidoreductase Core Subunit 3, a pseudogene); (8) NPR3 (Natriuretic Peptide Receptor 3, involved in blood volume and pressure and linked to hypertension); (9) LINC00521 (Coiled-Coil Domain Containing 197 a protein coding gene with unknown function); and (10) CNBD2 (Cyclic Nucleotide Binding Domain Containing 2, involved in cyclic AMP binding and important to male fertility).

#### Biochemistry

We found that accelerated biochemistry aging is 25.5±1.0% heritable and that 165 SNPs in 83 genes are significantly associated with this phenotype. The ten highest peaks on the Manhattan plot are: (1) RP11-218C14.2 (a pseudogene with unknown function). RP11-218C14.2 is in linkage disequilibrium with cystatin (cysteine protein inhibitors) encoding genes (CST3: cystatin C, a marker of kidney function; CST4: cystatin S and CST1: cystatin SN, both involved in salivary secretion; CST9: cystatin 9, possibly involved in hematopoietic differentiation or inflammation;

CST9L: cystatin 9 like, likely involved in tissue reorganization during testis development)[170]; (2) UDP glucuronosyltransferase encoding genes (UGT1A1, UGT1A3, UGT1A4, UGT1A5, UGT1A6, UGT1A7, UGT1A8, UGT1A9, UGT1A10), which are involved in the glucuronidation pathway, which is itself part of the cytochrome P450 mediated drug conjugation's phase of drug metabolism[170]; (3) tumor necrosis factors encoding genes (TNFSF12: TNF Superfamily Member 12, a cytokine which can induce apoptosis or regulate angiogenesis; TNFSF12-TNFSF13: a hybrid protein that stimulates T-cell and B-cell cell lines)[170]. TNFSF12 and TNFSF12-TNFSF13 are in linkage disequilibrium with MPDU1 (involved in the synthesis of oligosaccharides)[170], SHBG (sex hormone binding globulin, a steroid binding protein transporting androgens and estrogens, involved in steroid response)[170], POLR2A (a subunit of RNA polymerase II, a protein synthesizing mRNA)[170], ZBTB4 (Zinc Finger And BTB Domain Containing 4, a zinc fingers protein that represses transcription)[170] and SLC35G6 (Solute Carrier Family 35 Member G5), (4) JMJD1C (Jumonji Domain Containing 1C, involved in the DNA-damage response pathway)[170]; (5) G6PC2 (Glucose-6-Phosphatase Catalytic Subunit 2, an enzyme catalyzing the last step in gluconeogenic and glycogenolytic pathways, which produces glucose to be released into the bloodstream)[170]. G6PC2 is in linkage disequilibrium with SPC25 (SPC25 Component Of NDC80 Kinetochore Complex, possibly involved in kinetochore-microtubule interaction and spindle checkpoint activity during cell replication)[170] and ABCB11 (ATP Binding Cassette Subfamily B Member 11, an ABC transporter that catalyzes the secretion of conjugated bile salts and is involved in statins metabolism[171]); (6) RP11-66N5.2 (a pseudogene). RP11-66N5.2 is in linkage disequilibrium with GS1-542M4.4 (a long intergenic non-coding RNA); (7) ALDH1A2 (Aldehyde Dehydrogenase 1 Family Member A2, involved in the synthesis of retinoic acid); (8) RGAG1 (Retrotransposon Gag Like 9, an ortholog of MUC4, involved in the production of mucus). RGAG1 is in linkage disequilibrium with TDGF1P3 (a pseudogene) and CHRDL1 (Chordin Like 1, involved in retina angiogenesis); (9) SHROOM3 (Shroom Family Member 3, involved in cell shape regulation) and

(10) DGKB (Diacylglycerol Kinase Beta, involved in intracellular diacylglycerol mediated signaling).

We found that accelerated blood biochemistry aging is 26.2±0.3% heritable and that 1,081 SNPs in 450 genes are significantly associated with this phenotype. The ten highest GWAS peaks are: (1) CST3 (cystatin C); (2) UDP glucuronosyltransferase encoding genes (involved in drug metabolism); (3) POLR2A (RNA polymerase II, involved in mRNA synthesis); (4) HK1 (Hexokinase 1, involved in glucose metabolism); (5) G6PC2 (Glucose-6-Phosphatase Catalytic Subunit 2, involved in glucose metabolism); (6) RP11-66N5.2 (a pseudogene); (7) ALDH1A2 (Aldehyde Dehydrogenase 1 Family Member A2, involved in the synthesis of retinoic acid); (8) GCK (Glucokinase, involved in glucose metabolism); (9) AC012181.1 (a microRNA). AC012181.1 is in linkage disequilibrium with HERPUD1 (Homocysteine Inducible ER Protein With Ubiquitin Like Domain 1, involved in ubiquitinated protein degradation) and CETP (Cholesteryl Ester Transfer Protein, involved in cholesterol metabolism); and (10) TMEM258 (Transmembrane Protein 258, involved in protein N-glycosylation).

We found that accelerated urine biochemistry aging is 10.5±0.5% heritable and that 24 SNPs in 12 genes are significantly associated with this phenotype. Eight GWAS peaks can be identified, from most significantly associated to least significantly associated: (1) CUBN (cubilin, an endocytic receptor involved in vitamin B12, lipoprotein and iron metabolism)[170]; (2) COL4A4 COL4A4 (Collagen Type IV Alpha 4 Chain, a component of collagen that is only found in basement membranes)[170]; (3) CYP1A1 and CYP1A2 (Cytochrome P450 Family 1 Subfamily A Members 1 and 2, involved in drug metabolism)[170]; (4) GIPR (Gastric Inhibitory Polypeptide Receptor, stimulates insulin release when blood glucose is high)[170]. GIPR is in linkage disequilibrium with FUT2 (Fucosyltransferase 2, a Golgi stack protein involved in A and B antigen synthesis[170]); (5) AF146191.4 (5) AF146191.4 (long intergenic non-coding RNA with unknown function); (6) AC098592.8 (pseudogene with unknown function); (7) TRIM46 (Tripartite Motif Containing 46, involved in microtubules fasciculation in the axon)[170]. TRIM46 is in linkage disequilibrium with RP11-201K10.3 (a protein coding gene with unknown function), MUC1 (Mucin 1, Cell Surface Associated, involved in mucous secretion), THBS3 (Thrombospondin 3, a glycoprotein involved in cell-to-cell and cell-to-matrix interactions)[170] and MTX1 (Metaxin 1, involved in mitochondrial protein import); and (8) FGF5 (Fibroblast Growth Factor 5, involved in cell mitogenesis and survival).

#### Brain

We found that accelerated brain aging is 35.9±2.6% heritable and that 219 SNPs in 25 genes are significantly associated with this phenotype. The GWAS highlighted a single peak with the gene CRHR1 (Corticotropin Releasing Hormone Receptor 1, a GPCR protein involved in the hypothalamic-pituitary-adrenal pathway). CRHR1 is in linkage disequilibrium with other genes such as CRHR1-IT1, (CRHR1 intronic transcript 1), RP11-105N13.4 and RP11-669E14.4 (a processed transcript with unknown functions and an antisense gene linked to Alzheimer's disease, both in the Pre-MRNA Processing Factor 31 [PRPF31] gene. PRPF31 is a component of the spliceosome complex and is linked to Retinitis Pigmentosa) and LRRC37A2 (Leucine Rich Repeat Containing 37 Member A2, linked to progressive myoclonic epilepsy-6 and Koolen-De Vries syndrome).

We found that accelerated anatomical (MRI-based) brain aging is 39.1±1.7% heritable and that 235 SNPs in 35 genes are significantly associated with this phenotype. The GWAS highlighted six peaks: (1) KANSL1 (KAT8 Regulatory NSL Complex Subunit 1, involved in chromatin organization through histone acetylation and linked to Koolen-De Vries Syndrome). KANSL1 is in linkage disequilibrium with other genes such as MAPT (Microtubule Associated Protein Tau, linked to diverse neurodegenerative disorders such as frontotemporal dementia, progressive supranuclear palsy, Alzheimer's disease, Pick's disease and cortico-basal degeneration), MAPT-AS1 (MAPT Antisense RNA 1, a long non-coding RNA linked to diseases such as late onset Parkinson Disease and Koolen-De Vries Syndrome) and CRHR1 (Corticotropin Releasing Hormone Receptor 1, a GPCR protein involved in the hypothalamic-pituitary-adrenal pathway); (2) RUNX2 (RUNX Family Transcription Factor 2, involved in osteoblastic differentiation and skeletal morphogenesis); (3) RP11-213G21.2 (a long intergenic non-coding RNA). RP11-213G21.2 is in linkage disequilibrium with RPS23P3 (Ribosomal Protein S23 Pseudogene 3, involved in RNA synthesis); (4) MED8 (Mediator Complex Subunit 8, involved in RNA synthesis); (5) COLEC10 (Collectin Subfamily Member 10, a C-lectin cytosolic protein); and (6) PLIN4 (Perilipin 4, involved in lipid storage).

We found that accelerated cognitive aging is 29.3±2.0% heritable and that 27 SNPs in 20 genes are significantly associated with this phenotype. The GWAS highlighted three peaks: (1) PVRL2 (Poliovirus Receptor-Related 2 (Herpesvirus Entry Mediator B), involved in Herpes cell to cell transmission). PVRL2 is in linkage disequilibrium with CTB-129P6.4 (an antisense gene), TOMM40 (Translocase Of Outer Mitochondrial Membrane 40, involved in the transport of protein precursors into mitochondria) and APOC1 (Apolipoprotein C1, involved in high density lipoprotein (HDL) and very low density lipoprotein (VLDL) metabolism); (2) UBQLN1P1 (Ubiquilin 1 Pseudogene 1, a pseudogene). UBQLN1P1 is in linkage disequilibrium with several genes such as GNL1 (G Protein Nucleolar 1, part of the major histocompatibility complex), ATAT1 (Alpha Tubulin Acetyltransferase 1, involved in microtubule dynamics), C6orf136 (Chromosome 6 Open Reading Frame 136, linked to Central Nervous System Germ Cell Tumor and Spinocerebellar Ataxia), PPP1R18 (Protein Phosphatase 1 Regulatory Subunit 18, linked to deafness), MDC1 (Mediator Of DNA Damage Checkpoint 1, involved in DNA damage response); and (3) UBDP1 (Ubiquitin D Pseudogene 1, a pseudogene). UBDP1 is in linkage disequilibrium with OR5V1

(Olfactory Receptor Family 5 Subfamily V Member 1, involved in olfactory transduction) and XXbac-BPG13B8.10 (a long intergenic non-coding RNA).

#### Eyes

We found that accelerated eye aging is 28.2±1.2% heritable and that 55 SNPs in 27 genes are significantly associated with this phenotype. The GWAS highlighted nine peaks: (1) H3YL1 (SH3 And SYLF Domain Containing 1, linked to Meier-Gorlin syndrome); (2) HERC2 (HECT And RLD Domain Containing E3 Ubiquitin Protein Ligase 2, associated with eye pigmentation); (3) HTRA1 (HtrA Serine Peptidase 1, involved in cell growth regulation); (4) COL4A4 (Collagen Type IV Alpha 4 Chain, a component of collagen that is only found in basement membranes[170]); (5) TTK (TTK Protein Kinase, involved in mitotic checkpoint and linked to retinoblastoma); (6) SLC16A1 (Solute Carrier Family 16 Member 1, a monocarboxylate transporter); (7) LINC01072 (Long Intergenic Non-Protein Coding RNA 1072, a long intergenic non-coding RNA); (8) RDH5 (Retinol Dehydrogenase 5, involved in visual pigment synthesis and linked to Fundus Albipunctatus and Fundus Dystrophy); and (9) ABHD2 (Abhydrolase Domain Containing 2, Acylglycerol Lipase, linked to Retinitis Pigmentosa).

We found that accelerated eye fundus aging is 26.0±0.9% heritable and that 109 SNPs in 44 genes are significantly associated with this phenotype. The ten highest peaks highlighted by the GWAS are (1) SH3YL1 (linked to Meier-Gorlin syndrome); (2) HERC2 (associated with eye pigmentation); (3) LINC01072 (a long intergenic non-coding RNA); (4) IRF4 (Interferon Regulatory Factor 4, involved in immune response to viruses); (5) ST3GAL6-AS1 (ST3GAL6 Antisense RNA 1, a long intergenic non-coding RNA); (6) PPL (Periplakin, present in keratinocytes); (7) PDE6G (Retinal Rod Rhodopsin-Sensitive CGMP 3',5'-Cyclic, involved in the phototransduction signaling cascade of rod photoreceptors); (8) LGR6 (Leucine Rich Repeat Containing G Protein-Coupled Receptor 6, involved in GPCR signaling); (9) MACF1 (Microtubule Actin Crosslinking Factor 1, involved in actin-microtubules interactions); and (10) TCF4 (Transcription Factor 4, involved in corneal dystrophy).

We found that accelerated eye OCT aging is 23.6±0.9% heritable and that 149 SNPs in 80 genes are significantly associated with this phenotype. The ten highest peaks highlighted by the GWAS are (1) ABHD2 (Abhydrolase Domain Containing 2, Acylglycerol Lipase, linked to Retinitis Pigmentosa.); (2) ARMS2 (Age-Related Maculopathy Susceptibility 2, a component of the eye's choroidal extracellular matrix, linked to age-related macular degeneration and retinal drusen); (3) RDH5 (Retinol Dehydrogenase 5, involved in visual pigment synthesis and linked to Fundus Albipunctatus and Fundus Dystrophy); (4) RP1L1 (RP1 Like 1, a retinal-specific protein involved in photosensitivity and rod photoreceptors' morphogenesis, and linked to Occult Macular Dystrophy and Retinitis Pigmentosa); (5) COL4A4 (Collagen Type IV Alpha 4 Chain, a component of collagen that is only found in basement membranes[170]); (6) TTK (TTK Protein Kinase, involved in mitotic checkpoint and linked to retinoblastoma); (7) CFH (Complement Factor H, involved in innate immune response to microbial infections); (8) ZNF593 (Zinc Finger Protein 593, involved in transcriptional activity regulation); (9) OCA2 (OCA2 Melanosomal Transmembrane Protein, linked to iris pigmentation); and (10) APOC1 (Apolipoprotein C1, involved in high density lipoprotein (HDL) and very low density lipoprotein (VLDL) metabolism).

#### Hearing

We found that accelerated hearing aging is 14.1±0.4% heritable and that 662 SNPs in 243 genes are significantly associated with this phenotype. The six peaks highlighted by the GWAS are (1) OR2B4P (Olfactory Receptor Family 2 Subfamily B Member 4 Pseudogene, a GPCR involved in smell perception); (2) DDR1 (Discoidin Domain Receptor Tyrosine Kinase 1, a cell surface receptor); (3) APOC1 (Apolipoprotein C1, involved in high density lipoprotein (HDL) and very low density lipoprotein (VLDL) metabolism); (4) TSNARE1 (T-SNARE Domain Containing 1, involved in vesicle fusion and linked to schizophrenia); (5) AKAP6 (A-Kinase Anchoring Protein 6); and (6) OPCML (Opioid Binding Protein/Cell Adhesion Molecule Like, possibly involved in opioid receptor function).

#### Heart

We found that accelerated heart aging is 35.2±3.6% heritable and that three SNPs in three genes are significantly associated with this phenotype. The GWAS highlighted two peaks: (1) RP11-3N13.2 (a long intergenic non-coding RNA). RP11-3N13.2 is in linkage disequilibrium with MIR2052HG (a long intergenic non-coding RNA associated with breast cancer); and (2) TTN (Titin, protein present in striated muscle and linked to dilated cardiomyopathy and myofibrillar myopathy).

We found that accelerated heart anatomical (MRI-based) aging is 37.9±1.9% heritable and that 24 SNPs in eight genes are significantly associated with this phenotype. The GWAS highlighted four peaks: (1) TTN (Titin, protein present in striated muscle and linked to dilated cardiomyopathy and myofibrillar myopathy); (2) RP11-3N13.2 (a long intergenic non-coding RNA); (3) RP11-224P11.1 (a pseudogene); and (4) MASP1 (Mannan Binding Lectin Serine Peptidase 1, involved in lectin pathway of complement activation).

We found that accelerated heart electrical (ECG-based) aging is 21.8±2.9% heritable but we did not find any SNPs significantly associated with this phenotype.

#### Blood cells

We found that accelerated blood cell aging is 18.1±0.2% heritable and that 2,636 SNPs in 1,056 genes are significantly associated with this phenotype. The ten highest peaks highlighted by the

GWAS are (1) TUBB1 (Tubulin Beta 1 Class VI, involved in microtubule formation, expressed in thrombocytes and megakaryocytes and linked to macrothrombocytopenia); (2) HBS1L (HBS1 Like Translational GTPase, involved in erythrocyte volume, hemoglobin content, erythrocyte count, platelet count and monocyte count, and linked to sickle cell disease and beta-thalassemia); (3) AC020595.1 (a long intergenic non-coding RNA); (4) SPTA1 (Spectrin Alpha, Erythrocytic 1, a component of red blood cell plasma membrane and linked to erythrocyte diseases such as elliptocytosis 2, pyropoikilocytosis, and type 3 spherocytosis); (5) ANK1 (Ankyrin 1, Erythrocytic, a protein attaching membrane proteins to cytoskeletal elements, linked to spherocytosis - a red blood cell disease); (6) RCL1 (RNA Terminal Phosphate Cyclase Like 1, involved in RNA processing and linked to breast lipoma); (7) RP11-202G18.1 (a long intergenic non-coding RNA); (8) WDR66 (Cilia And Flagella Associated Protein 251, involved in mean platelet volume and spermatozoa motility, and linked to spermatogenic failure and non-syndromic male infertility due to sperm motility disorder); (9) SLC12A2 (Solute Carrier Family 12 Member 2, involved in sodium and chloride transport); and (10) ARHGEF3 (Rho Guanine Nucleotide Exchange Factor 3, a guanine nucleotide exchange factor for RhoA and RhoB GTPases).

#### Lungs

We found that accelerated lung aging is 30.3±0.2% heritable and that 4,286 SNPs in 1,460 genes are significantly associated with this phenotype. The ten highest peaks highlighted by the GWAS are (1) NPNT (Nephronectin, involved in kidney development); (2) KANSL1 (KAT8 Regulatory NSL Complex Subunit 1, involved in chromatin organization through histone acetylation and linked to Koolen-De Vries Syndrome); (3) ADAMTSL3 (A Disintegrin-Like And Metalloprotease Domain With Thrombospondin Type I Motifs-Like 3, involved in extracellular matrix remodelling[172]); (4) EFEMP1 (EGF Containing Fibulin Extracellular Matrix Protein 1, an extracellular matrix glycoprotein linked to eye disease familial Drusen and Doyne Honeycomb Retinal Dystrophy); (5) HMGA2 (High Mobility Group AT-Hook 2, involved in transcription regulation); (6) CDC123 (Cell Division Cycle 123, involved in cell cycle regulation and linked to Wolfram syndrome); (7) RP13-539F13.2 (a pseudogene). RP13-539F13.2 is in linkage disequilibrium with ANAPC10 (Anaphase Promoting Complex Subunit 10, involved in cell cycle regulation); (8) HTR4 (5-Hydroxytryptamine Receptor 4, involved in neurotransmitter regulation and linked to irritable bowel syndrome and constipation); (9) CYCSP55 (Cytochrome C, Somatic-Like 1 (Pseudogene), a pseudogene); and (10) KDM2A (Lysine Demethylase 2A, involved in ubiquitination).

#### Musculoskeletal

We found that accelerated musculoskeletal aging is 34.9±1.8% heritable and that 11 SNPs in nine genes are significantly associated with this phenotype. The GWAS highlighted six peaks: (1) FGFR3P1 (Fibroblast Growth Factor Receptor 3 Pseudogene 1, a pseudogene involved in vascular endothelial growth factor signaling); (2) TBX15 (T-Box Transcription Factor 15, transcription factor involved in development and linked to Cousin syndrome and acromegaloid facial appearance syndrome); (3) CCDC91 (Coiled-Coil Domain Containing 91, involved in the trans-Golgi network associated with ossification of the posterior longitudinal ligament of the spine and diffuse idiopathic skeletal hyperostosis); (4) AAGAB (Alpha And Gamma Adaptin Binding Protein, involved in vesicle trafficking and linked to palmoplantar keratoderma); (5) FAM3C (FAM3 Metabolism Regulating Signaling Molecule C, linked to deafness and pancreatic cancer); and (6) SCEL (Sciellin, expressed in keratinocytes and linked to extrahepatic bile duct adenocarcinoma).

We found that accelerated full body musculoskeletal aging is 30.7±1.6% heritable and that eight SNPs in eight genes are significantly associated with this phenotype. The ten highest peaks highlighted by the GWAS are (1) TBX15 (T-Box Transcription Factor 15, transcription factor involved in development and linked to Cousin syndrome and acromegaloid facial appearance syndrome); (2) FGFR3P1 (Fibroblast Growth Factor Receptor 3 Pseudogene 1, a pseudogene involved in vascular endothelial growth factor signaling); (3) SCEL (Sciellin, expressed in keratinocytes and linked to extrahepatic bile duct adenocarcinoma); and (4) FGFR1 (Fibroblast Growth Factor Receptor 1, a fibroblast growth factor receptor linked to osteoglophonic dysplasia and Hartsfield syndrome).

We found that accelerated spine aging is 32.9±1.7% heritable and that 22 SNPs in 13 genes are significantly associated with this phenotype. The GWAS highlighted nine peaks: (1) TNFSF11 (Tumor Necrosis Factor Ligand Superfamily Member 11, a ligand for osteoprotegerin involved in osteoclast differentiation and activation, and linked to osteopetrosis); (2) HTRA1 (HtrA Serine Peptidase 1, involved in cell growth regulation); (3) FAM3C (FAM3 Metabolism Regulating Signaling Molecule C, linked to deafness and pancreatic cancer); (4) SUPT3H (SPT3 Homolog, SAGA And STAGA Complex Component, involved in chromatin organization and linked to dysostosis and hepatic adenoma); (5) RP11-438P9.2 (a pseudogene); (6) SOX5 (SRY-Box Transcription Factor 5, involved in chondrocytes differentiation and cartilage formation, linked to Lamb-Shaffer syndrome); (7) ESR1 (Estrogen Receptor 1, an estrogen receptor involved in sexual functions and linked to osteoporosis, breast cancer and endometrial cancer); (8) COLGALT2 (Collagen Beta(1-O)Galactosyltransferase 2, involved in degradation of the extracellular matrix and linked to porencephaly); and (9) AAGAB (Alpha And Gamma Adaptin Binding Protein, involved in vesicle trafficking and linked to palmoplantar keratoderma).

We found that accelerated hip aging is 27.7±1.6% heritable and that 20 SNPs in seven genes are significantly associated with this phenotype. The GWAS highlighted four peaks: (1) SUPT3H (SPT3 Homolog, SAGA And STAGA Complex Component, involved in chromatin organization and linked to dysostosis and hepatic adenoma); (2) CCDC91 (Coiled-Coil Domain Containing 91, involved in the trans-Golgi network associated with ossification of the posterior longitudinal ligament of the spine and diffuse idiopathic skeletal hyperostosis); (3) TNFSF11 (Tumor Necrosis

Factor Ligand Superfamily Member 11, a ligand for osteoprotegerin involved in osteoclast differentiation and activation, and linked to osteopetrosis); and (4) RP11-396020.1 (a long intergenic non-coding RNA).

We found that accelerated knee aging is 25.3±1.7% heritable and that 16 SNPs in six genes are significantly associated with this phenotype. The GWAS highlighted four peaks: (1) WNT16 (Wnt Family Member 16, involved in signaling, cell fate and and oncogenesis); (2) CCDC91 (Coiled-Coil Domain Containing 91, involved in the trans-Golgi network associated with ossification of the posterior longitudinal ligament of the spine and diffuse idiopathic skeletal hyperostosis); and (3) HLA-DRB9 (Major Histocompatibility Complex, Class II, DR Beta 9 Pseudogene, a pseudogene).

We found that accelerated musculoskeletal scalar features-based (anthropometry, impedance, heel bone densitometry and hand grip strength) aging is 22.1±0.2% heritable and that 2,631 SNPs in 954 genes are significantly associated with this phenotype. The ten highest peaks highlighted by the GWAS are (1) GDF5 (Growth Differentiation Factor 5, involved in the development of cartilage, joints, brown fat, teeth, and neurons, linked to osteoarthritis, acromesomelic dysplasia, brachydactyly, proximal symphalangism, chondrodysplasia and multiple synostoses syndrome); (2) CRHR1 (Corticotropin Releasing Hormone Receptor 1, a GPCR protein involved in the hypothalamic-pituitary-adrenal pathway); (3) HOTTIP (HOXA Distal Transcript Antisense RNA, involved in cell proliferation and linked to tumors); (4) SOCS2 (Suppressor Of Cytokine Signaling 2, involved in cytokine signaling and linked to polycythemia); (5) PKD2L1 (Polycystin 2 Like 1, Transient Receptor Potential Cation Channel, involved in cell-cell and cell-matrix interactions and linked to polycystic kidney disease); (6) GPR126 (Adhesion G Protein-Coupled Receptor G6, a G protein-coupled receptor involved in body height, linked to distal arthrogryposis); (7) AC013480.2 (an antisense gene). AC013480.2 is in linkage disequilibrium with PKDCC (Protein Kinase Domain Containing, Cytoplasmic, involved in development and linked to rhizomelic limb shortening with dysmorphic features); (8) POLD3 (DNA Polymerase Delta 3, Accessory Subunit, involved in DNA replication and repair and linked to Ruijs-Aalfs syndrome); (9) ZBTB38 (Zinc Finger And BTB Domain Containing 38, a transcriptional activator involved in height); and (10) FUBP3 (Far Upstream Element Binding Protein 3, involved in gene expression).

##### Physical activity

We found that accelerated physical activity aging is 12.3±0.9% heritable and that nine SNPs in seven genes are significantly associated with this phenotype. The GWAS highlighted two peaks: (1) RSL24D1P1 (a pseudogene). RSL24D1P1 is in linkage disequilibrium with HIST1H4L (H4 Clustered Histone 13, an histone, involved in chromatin organization); and (2) OR5V1 (Olfactory Receptor Family 5 Subfamily V Member 1, involved in olfactory transduction).

#### Identification of biomarkers, clinical phenotypes, diseases, family history, environmental variables and sociodemographics associated with accelerated aging

Below, we report for each X-category the three subcategories most associated with general accelerated aging, that is whose variables are most often positively associated with accelerated aging across aging dimensions. The reader should keep in mind that not all X-variables have the same sample size, and that our selection is therefore biased towards X-subcategories with large sample sizes. For each of these three subcategories, we list the three variables most often positively and statistically significantly associated with accelerated aging across aging dimensions, along with the average correlation between the variable and accelerated aging (between aging dimensions for which the association was both positive and statistically significant). We then list the top three subcategories and their associated top three variables for a specific aging dimension.

We provide the above summary for both associations with accelerated aging, and with decelerated aging.

#### Biomarkers

##### Biomarkers associated with accelerated aging

Supplementary Figure 30 displays which biomarker categories are most associated with accelerated aging for each aging dimension. For each biomarkers subcategory, we list the three biomarkers most associated with accelerated aging across all aging dimensions in Supplementary Table 7. For example, the three biomarker categories most associated with general accelerated aging are blood pressure, body impedance and arterial stiffness. Specifically, on average 66.7% of blood pressure biomarkers are associated with accelerated aging across aging dimensions, with the three most frequent associations being systolic blood pressure (average correlation=.093±.083), diastolic blood pressure (average correlation=.067±.022) and pulse rate (average correlation=.057±.022). 41.2% of body impedance biomarkers are associated with accelerated aging across aging dimensions on average, with the three most frequent associations being right arm impedance (average correlation=.051±.031), left leg impedance (average correlation=.048±.037), and whole-body impedance (average correlation=.052±.038)). 40.5% of arterial stiffness biomarkers are associated with accelerated aging across aging dimensions on average, with the three most frequent associations being position of the pulse wave peak (average correlation=.068±.066), position of the shoulder on the pulse waveform (average correlation=.073±.065), and absence of notch position in the pulse waveform (average correlation=.067±.071).

We list the three biomarkers most associated with accelerated aging in each specific aging dimension in Supplementary Table 8. For example, accelerated blood biochemistry aging is most associated with urine biochemistry, blood biochemistry and blood count biomarkers. Specifically, 100.0% of urine biochemistry biomarkers are associated with accelerated blood biochemistry aging, with the three largest associations being creatinine in urine (correlation=.083), sodium in urine (correlation=.076), and potassium in urine (correlation=.070)). 71.4% of blood biochemistry biomarkers are associated with accelerated blood biochemistry aging, with the three largest associations being cystatin (correlation=.364), urea (correlation=.265), and glycated hemoglobin (correlation=.231). 61.3% of blood count biomarkers are associated with accelerated blood biochemistry aging, with the three largest associations being eosinophil count (correlation=.076), white blood cell count (correlation=.075), and red blood cell distribution width (correlation=.069).

#### Biomarkers associated with decelerated aging

Supplementary Figure 31 displays which biomarker categories are most associated with decelerated aging for each aging dimension. For each biomarker subcategory, we list the three biomarkers most associated with decelerated aging across all aging dimensions in Supplementary Table 9. For example, the three biomarker categories most associated with general decelerated aging are hand grip strength, spirometry and cognitive symbol digit substitution - a cognitive test. Specifically, on average 97.0% of hand grip strength biomarkers are associated with accelerated aging across aging dimensions, with the associations being left hand grip strength (average correlation=.088±.093), and right hand grip strength (average correlation=.088±.094). 79.8% of spirometry biomarkers are associated with accelerating aging across aging dimensions on average, with the three largest associations being peak expiratory flow (average correlation=.080±.098), forced expiratory volume in 1-second (average correlation=.113±.159), and forced vital capacity (average correlation=.124±.167). 68.2% of symbol digit substitution biomarkers are associated with accelerating aging across aging dimensions on average, with the associations being number of symbol digit matches made correctly (average correlation=.100±.132), and number of digits matches attempted (average correlation=.103±.144).

We list the three biomarkers most associated with decelerated aging in each specific aging dimension in Supplementary Table 8. For example, decelerated knee aging is most associated with hand grip strength, heel bone densitometry and anthropometry. Specifically, 100.0% of hand grip strength biomarkers are associated with accelerated knee aging, with the associations being right hand grip strength (correlation=.049) and left hand grip strength (correlation=.037). 100.0% of heel bone densitometry biomarkers are associated with accelerated knee aging, with the three largest associations being heel broadband ultrasound attenuation (correlation=.106), heel bone mineral density (correlation=.104), heel quantitative ultrasound index (correlation=.103). 75.0% of anthropometry biomarkers are associated with accelerated knee aging, with the three largest associations being body mass index (correlation=.177), weight (correlation=.140) and hip circumference (correlation=.126).

#### Clinical phenotypes

##### Clinical phenotypes associated with accelerated aging

Supplementary Figure 32 displays which clinical phenotypes categories are most associated with accelerated aging for each aging dimension. For each clinical phenotype subcategory, we list the three clinical phenotypes most associated with accelerated aging across all aging dimensions in Supplementary Table 12. For example, the three clinical phenotypes categories most associated with general accelerated aging are breathing, chest pain and general health. Specifically, on average 81.8% of breathing clinical phenotypes are associated with accelerated aging across aging dimensions, with the most frequent associations being shortness of breath walking on level ground (average correlation=.062±.029), and wheeze or whistling in the chest in last year (average correlation=.047±.033). 59.8% of chest pain clinical phenotypes are associated with accelerated aging across aging dimensions on average, with the three most frequent associations being chest pain or discomfort walking normally (average correlation=.036±.013), chest pain due to walking ceases when standing still (average correlation=.041±.018), and chest pain or discomfort (average correlation=.035±.016). 47.3% of general health clinical phenotypes are associated with accelerated aging across aging dimensions on average, with the three most frequent associations being long-standing illness, disability or infirmity (average correlation=.073±.032), overall health rating (average correlation=.099±.036), and falls in the last year (average correlation=.039±.015).

We list the three clinical phenotypes most associated with accelerated aging in each specific aging dimension in Supplementary Table 13. For example, accelerated brain cognitive aging is most associated with breathing, chest pain and mental health. Specifically, 100.0% of breathing clinical phenotypes are associated with accelerated cognitive aging, with the associations being shortness of breath walking on level ground (correlation=.056), and wheeze or whistling in the chest in last year (correlation=.036). 50% of chest pain clinical phenotypes are associated with accelerated cognitive aging, with the associations being chest pain or discomfort walking normally (correlation=.031), and chest pain or discomfort (correlation=.027). 41.3% of mental health clinical phenotypes are associated with accelerated cognitive aging, with the three largest associations being nervous feelings (correlation=.607), worrier/anxious feelings (correlation=.065), and frequency of tiredness/lethargy in last two weeks (correlation=.056).

#### Clinical phenotypes associated with decelerated aging

Supplementary Figure 33 displays which clinical phenotypes categories are most associated with decelerated aging for each aging dimension. For each clinical phenotype subcategory, we list the three clinical phenotypes most associated with decelerated aging across all aging dimensions in Supplementary Table 14. For example, the three clinical phenotypes categories most associated with general decelerated aging are cancer screening, sexual factors and general health. Specifically, on average 18.2% of cancer screening clinical phenotypes are associated with decelerated aging across aging dimensions, with the most frequent associations being most recent bowel cancer screening (average correlation=.030±.008), ever had prostate specific antigen test (average correlation=.028±.009), and ever had bowel cancer screening (average correlation=.014±.003). 15.9% of sexual factors clinical phenotypes are associated with decelerated aging across aging dimensions on average, with the most frequent associations being age first had sexual intercourse (average correlation=.034), ever had same sex intercourse (average correlation=.023), and lifetime number of sexual partners (average correlation=.011). 11.0% of general health clinical phenotypes are associated with decelerated aging across aging dimensions on average, with the most frequent associations being no weight change compared to one year ago (average correlation=.034±.014), gained weight compared to one year ago (average correlation=.042±.018) and lost weight compared to one year ago (average correlation=.016±.001).

We list the three clinical phenotypes most associated with decelerated aging in each specific aging dimension in Supplementary Table 15. For example, decelerated brain cognitive aging is most associated with mouth, general pain and mental health. Specifically, 12.5% of mouth clinical phenotypes are associated with decelerated cognitive aging, with the association being no mouth/teeth dental problems (correlation=.056). 5.6% of general pain clinical phenotypes are associated with decelerated cognitive aging, with the association being no particular pain experienced in the last month (correlation=.032). 4.3% of mental health clinical phenotypes are associated with accelerated cognitive aging, with the associations being no manic/hyper symptoms (correlation=.066), and no problems in terms of severity of manic/irritable episodes (correlation=.058).

#### Diseases

##### Diseases associated with accelerated aging

Supplementary Figure 34 displays which disease categories are most associated with accelerated aging for each aging dimension. For each disease subcategory, we list the three diseases most associated with accelerated aging across all aging dimensions in Supplementary Table 17. For example, the three clinical phenotypes categories most associated with general accelerated aging are medical diagnoses I (cardiac diseases), medical diagnoses Z (various diseases) and medical diagnoses R (various diseases). Specifically, on average 18.9% of medical diagnoses I diseases are associated with accelerated aging across aging dimensions, with the three most frequent associations being hypertension (average correlation=.074±.031), chronic ischemic heart disease (average correlation=.040±.017), and atrial fibrillation and flutter (average correlation=.040±.013). 14.2% of medical diagnoses Z diseases are associated with accelerated aging across aging dimensions on average, with the three most frequent associations being personal history of certain other diseases (average correlation=.045±.016), problems related to lifestyle (average correlation=.035±.012), and personal history of medical treatment (average correlation=.043±.017). 14.2% of medical diagnoses R diseases are associated with accelerated aging across aging dimensions on average, with the three most frequent associations being pain in throat and chest (average correlation=.033±.012), abnormalities of breathing (average correlation=.025±.009), and abdominal and pelvic pain (average correlation=.024±.009).

We list the three diseases most associated with accelerated aging in each specific aging dimension in Supplementary Table 18. For example, accelerated eye fundus aging is most associated with medical diagnoses H (eye diseases), medical diagnoses Z (various diseases), and medical diagnoses E (metabolic diseases). Specifically, 15.7% of medical diagnoses H diseases are associated with accelerated eye fundus aging, with the three largest associations being other cataract (correlation=.051), retinal disorders in diseases classified elsewhere (correlation=.050) and retinal detachment and breaks (.032). 11.9% of medical diagnoses Z diseases are associated with accelerated eye fundus aging, with the three largest associations being personal history of certain other diseases (correlation=.029), problems related to lifestyle (correlation=.027), and dependence on enabling machines and devices, not classified elsewhere (correlation=.026). 10.1% of medical diagnoses E diseases are associated with accelerated eye fundus aging, with the three largest associations being non-insulin dependent diabetes mellitus (correlation=.070), insulin dependent diabetes mellitus (correlation=.058), and disorders of lipoprotein metabolism and other lipidemias (correlation=.029).

##### Diseases associated with decelerated aging

Supplementary Figure 35 displays which disease categories are most associated with decelerated aging for each aging dimension. For each disease subcategory, we list the three diseases most associated with decelerated aging across all aging dimensions in Supplementary Table 19. For example, the three clinical phenotypes categories most associated with general decelerated aging are medical diagnoses O (obstetrics diseases), medical diagnoses Z (various diseases) and medical diagnoses K (gastrointestinal tract diseases). Specifically, on average 2.8% of medical diagnoses O diseases are associated with decelerated aging across aging dimensions, with the three most frequent associations being perineal laceration during delivery (average correlation=.027±.010), single spontaneous delivery (average correlation=.021±.005), and long labor (average correlation=.018±.005). 0.9% of medical diagnoses Z diseases are associated with decelerated aging across aging dimensions on average, with the three most frequent associations being outcome of delivery (average correlation=.038±.011), supervision of normal pregnancy (average correlation=.014±.004), and postpartum care and examination (average correlation=.009±.001). 0.3% of medical diagnoses K diseases are associated with decelerated aging across aging dimensions on average, with the three most frequent associations being inguinal hernia (average correlation=.009±.003), diverticular disease of intestine (average correlation=.012), and cholelithiasis (average correlation=.015).

We list the three diseases most associated with decelerated aging in each specific aging dimension in Supplementary Table 20. For example, decelerated blood biochemistry aging is most associated with medical diagnoses O (obstetrics diseases), medical diagnoses Z (various diseases), and medical diagnoses N (urinary and reproductive systems diseases). Specifically, 36.1% of medical diagnoses O diseases are associated with decelerated blood biochemistry aging, with the three largest associations being perineal laceration during delivery (correlation=.047), labor and delivery complicated by fetal stress (correlation=.034) and single spontaneous delivery (.032). 6.0% of medical diagnoses Z diseases are associated with accelerated blood biochemistry aging, with the three largest associations being outcome of delivery (correlation=.063), antenatal screening (correlation=.017), and supervision of normal pregnancy (correlation=.017). 3.7% of medical diagnoses N diseases are associated with accelerated blood biochemistry aging, with the three largest associations being prostate hyperplasia (correlation=.015), female infertility (correlation=.013), and erosion and ectropion of cervix uteri (correlation=.009).

### Family history

#### Family history associated with accelerated aging

On average 1.3% of family history variables are associated with accelerated aging across aging dimensions, with the three most frequent associations being siblings' illnesses: prefer not to answer (average correlation=.012±.002), sibling's severe depression (average correlation=.010±.000), and mother's diabetes (average correlation=.037±.008).

We list the three family history variables most associated with accelerated aging in each specific aging dimension in Supplementary Table 22. For example, 6.5% of family history variables are associated with accelerated lung aging, with the three largest associations being father's bronchitis/emphysema (correlation=.121), mother's bronchitis/emphysema (correlation=.092) and adopted mother's diabetes (correlation=.054).

#### Family history associated with decelerated aging

On average 1.4% of family history variables are associated with decelerated aging across aging dimensions, with the three most frequent associations being father's prostate cancer (average correlation=.020±.009), adopted sibling's stroke (average correlation=.114±.025), and father's lung cancer (average correlation=.101±.057).

We list the three family history variables most associated with decelerated aging in each specific aging dimension in Supplementary Table 23. For example, 2.2% of family history variables are associated with decelerated lung aging, with the associations being mother's Alzheimer's disease/dementia (correlation=.022), and father's prostate cancer (correlation=.012).

### Environmental variables

#### Environmental variables associated with accelerated aging

Supplementary Figure 36 displays which environmental categories are most associated with accelerated aging for each aging dimension. For each environmental subcategory, we list the three environmental categories most associated with accelerated aging across all aging dimensions in Supplementary Table 25. Because we find environmental exposures to be of particular interest, we present all subcategories, by decreasing association. 36.4% of smoking variables are associated with accelerated aging across aging dimensions on average, with the three most frequent associations being currently smoking tobacco on most or all days (average correlation=.056±.023), pack years adult smoking as proportion of life span exposed to smoking (average correlation=.084±.031), and difficulty not smoking for one day (average correlation=.058±.023). 24.7% of sleep variables are associated with accelerated aging across aging dimensions on average, with the three most frequent associations being napping during the day (average correlation=.047±.025), daytime dozing/sleeping - narcolepsy (average correlation=.039±.016), and sleeplessness/insomnia (average correlation=.038±.016). 19.7% of sun exposure variables are associated with accelerated aging across aging dimensions on average, with the three most frequent associations being facial aging: about your age (average correlation=.032±.025), time spent outdoors in summer (average correlation=.046±.014), and facial aging: older than you are (average correlation=.025±.007). 15.9% of alcohol intake variables are associated with accelerated aging across aging dimensions on average, with the three most frequent associations being average weekly beer plus cider intake (average correlation=.043±.021), reason for reducing amount of alcohol drunk: illness of ill health (average correlation=.045±.017), and average weekly spirits intake (average correlation=.035±.011). 12.2% of early life factors variables are associated with accelerated aging across aging dimensions on average, with the three most frequent associations being taller comparative height size at age ten (average correlation=.044±.012), plumper comparative body size at age ten (average correlation=.028±.013), and does not know whether was breastfed as a baby (average correlation=.024±.008). 11.6% of physical activity (questionnaire) variables are associated with accelerated aging across aging dimensions on average, with the three most frequent associations being no physical activity in the last four weeks (average correlation=.039±.017), time spent watching television (average correlation=.061±.033), and types of transport used (excluding work): public transport (average correlation=.052±.014). 11.3% of diet variables are associated with accelerated aging across aging dimensions on average, with the three most frequent associations being major dietary changes in the last five years because of illness (average correlation=.047±.021), bread type: white (average correlation=.041±.020), and processed meat intake (average correlation=.037±.011). 8.4% of medication variables are associated with accelerated aging across aging dimensions on average, with the three most frequent associations being taking a cholesterol lowering medication (average correlation=.060±.036), taking aspirin (average correlation=.046±.021), and taking a blood pressure medication (average correlation=.045±.019). 7.9% of electronic devices variables are associated with accelerated aging across aging dimensions on average, with the three most frequent associations being playing computer games (average correlation=.029±.010), usual side of head for mobile phone use: left (.018±.006), and weekly usage of mobile phone in last three months (.011±.001).

We list the three environmental categories most associated with accelerated aging in each specific aging dimension in Supplementary Table 26. For example, accelerated eye aging is most associated with sleep, smoking and physical activity (questionnaire). Specifically, 57.1% of sleep variables are associated with accelerated eye aging, with the three largest associations being napping during the day (correlation=.027), being a morning/evening person - chronotype (correlation=.026) and sleeplessness/insomnia (correlation=.026). 37.5% of smoking variables are associated with accelerated eye aging, with the three largest associations being pack years adult smoking as proportion of life span exposed to smoking (correlation=.092), pack years of smoking (correlation=.090), and past tobacco smoking: smoked on most or all days (correlation=.066). 11.4% of physical activity variables are associated with accelerated eye aging, with the three largest associations being time spent watching television (correlation=.063), no physical activity during the last four weeks (correlation=.051), and types of transport used (excluding work): public transport (correlation=.042).

#### Environmental variables associated with decelerated aging

Supplementary Figure 37 displays which environmental categories are most associated with decelerated aging for each aging dimension. For each environmental subcategory, we list the three environmental variables most associated with decelerated aging across all aging dimensions in Supplementary Table 27. Because we find environmental exposures to be of particular interest, we present all subcategories, by decreasing association. 34.6% of physical activity (questionnaire) variables are associated with decelerated aging across aging dimensions on average, with the three most frequent associations being usual walking pace (average correlation=.070±.037), drive faster than motorway speed limit (average correlation=.053±.029), and duration of other exercises (average correlation=.053±.024). 24.9% of smoking variables are associated with decelerated aging across aging dimensions on average, with the three most frequent associations being time from waking to first cigarette (average correlation=.058±.023), age started smoking in current smokers (average correlation=.057±.023), and not currently smoking tobacco (average correlation=.054±.021). 13.0% of sleep variables are associated with decelerated aging across aging dimensions on average, with the three most frequent associations being snoring (average correlation=.030±.010), getting up in the morning (average correlation=.031±.012), and sleep duration (.020±.005). 11.8% of electronic devices variables are associated with decelerated aging across aging dimensions on average, with the three most frequent associations being hands-free device/speakerphone use with mobile phone in the last three months (average correlation=.032±.020), length of mobile phone use (average correlation=.044±.026), and length of mobile phone use compared to two years previously (average correlation=.035±.018). 10.5% of medication variables are associated with decelerated aging across aging dimensions on average, with the three most frequent associations being not taking any medication for cholesterol, blood pressure or diabetes (average correlation=.094±.042), not taking any medication for cholesterol, blood pressure, diabetes or taking exogenous hormones (average correlation=.057±.027), and not taking medication for pain relief, constipation or heartburn (average correlation=.041±.017). 9.3% of diet variables are associated with decelerated aging across aging dimensions on average, with the three most frequent associations being no major dietary change in the last five years (average correlation=.034±.012), cereal intake (average correlation=.047±.020), and eating both eggs, dairy, wheat and sugar (average correlation=.036±.014). 9.1% of alcohol variables are associated with decelerated aging across aging dimensions on average, with the three most frequent associations being alcohol usually taken with meals (average correlation=.041±.018), alcohol drinker status (average correlation=.035±.011), and average weekly red wine intake (average correlation=.039±.014). 8.9% of sun exposure variables are associated with decelerated aging across aging dimensions on average, with the three most frequent associations being facial aging: younger than you are (average correlation=.045±.023), skin colour (average correlation=.026±.013), and natural hair color before graying: dark brown (average correlation=.018±.006). 7.6% of early life factors variables are associated with decelerated aging across aging dimensions on average, with the three most frequent associations being comparative height size at age ten: shorter (average correlation=.039±.012), being breastfed as a baby (average correlation=.031±.010), and about average comparative body size at age ten (average correlation=.024±.008).

We list the three environmental categories most associated with decelerated aging in each specific aging dimension in Supplementary Table 28. For example, decelerated cognitive aging is most associated with electronic devices, physical activity (questionnaire) and sleep. Specifically, 40.0% of electronic devices variables are associated with decelerated cognitive aging, with the three largest associations being playing computer games (correlation=.125), hands-free device/speakerphone use with mobile phone in the last three months (correlation=.039) and usual side of head for mobile phone use: equality left and right (correlation=.025). 34.3% of physical activity variables are associated with decelerated cognitive aging, with the three largest associations being driving faster than motorway speed limit (correlation=.125), usual walking pace (correlation=.070), and time spent using the computer (correlation=.054). 14.3% of physical activity variables are associated with accelerated eye aging, with the association being getting up in the morning (correlation=.047).

#### Socioeconomics

##### Socioeconomic variables associated with accelerated aging

Supplementary Figure 38 displays which socioeconomic variable categories are most associated with accelerated aging for each aging dimension. For each socioeconomic variable subcategory, we list the three socioeconomic variables most associated with accelerated aging across all aging dimensions in Supplementary Table 30. Because there are only five socioeconomic variables categories, we present all of them, by decreasing association. 16.5% of “other sociodemographics” variables are associated with accelerated aging across aging dimensions on average, with the three most frequent associations being private healthcare (average correlation=.035±.011), receiving attendance allowance (average correlation=.026±.008), and not knowing whether one receives attendance/disability/mobility allowance (average correlation=.016±.005). 13.9% of social support variables are associated with accelerated aging across aging dimensions on average, with the three most frequent associations being not having leisure/social activities (average correlation=.029±008), leisure/social activities: pub or social club (average correlation=.034±.014) and leisure/social activities: religious group (average correlation=.065±.023). 11.0% of education variables are associated with accelerated aging across aging dimensions on average, with the three most frequent associations being qualifications: none of the above (average correlation=.056±.031), qualifications: CSEs or equivalent (average correlation=.039±.012), and qualifications: prefer not to answer (average correlation=.021±.008). 8.5% of household variables are associated with accelerated aging across aging dimensions on average, with the three most frequent associations being renting ones accommodation from local authority, local council or housing association (average correlation=.048±.020), living in a flat, maisonette or apartment (average correlation=.035±.010), and gas or solid-fuel cooking/heating: a gas fire that you use regularly in winter time (average correlation=.039±.014). 5.0% of employment variables are associated with accelerated aging across aging dimensions on average, with the three most frequent associations being job involves night shift work (average correlation=.026±.007), job involves heavy manual or physical work (average correlation=.025±.014), and current employment status: prefer not to answer (average correlation=.014±.004).

We list the three socioeconomic variables categories most associated with accelerated aging in each specific aging dimension in Supplementary Table 31. For example, accelerated cognitive aging is most associated with “other sociodemographics”, employment and education. Specifically, 28.6% of “other sociodemographics” variables are associated with accelerated cognitive aging, with the associations being private healthcare (correlation=.027), and receiving an attendance allowance (correlation=.025). 13.0% of employment variables are associated with accelerated cognitive aging, with the three largest associations being job involves heavy manual or physical work (correlation=.039), job involves night shift work (correlation=.036), and job involves mainly walking or standing (correlation=.030). 12.5% of education variables are associated with accelerated cognitive aging, with the association being qualifications: none of the above (correlation=.071).

#### Socioeconomic variables associated with decelerated aging

Supplementary Figure 39 displays which socioeconomic variable categories are most associated with decelerated aging for each aging dimension. For each socioeconomic variable subcategory, we list the three socioeconomic variables most associated with decelerated aging across all aging dimensions in Supplementary Table 32. Because there are only five socioeconomic variables categories, we present all of them, by decreasing association. 16.4% of employment variables are associated with decelerated aging across aging dimensions on average, with the three most frequent associations being current employment status: in paid employment or self-employed (average correlation=.061±.057), length of working week for main job (average correlation=.065±.068), and frequency of travelling from home to job workplace (average correlation=.057±.068). 16.3% of education variables are associated with decelerated aging across aging dimensions on average, with the three most frequent associations being qualifications: college or university degree (average correlation=.059±.021), qualifications: A levels/AS levels or equivalent (average correlation=.044±.013), and qualifications: O levels/GCSEs or equivalent (average correlation=.034±.015). 13.9% of social support variables are associated with decelerated aging across aging dimensions on average, with the three most frequent associations being leisure/social activities: sports club or gym (average correlation=.051±.024), able to confide (average correlation=.026±.011), and leisure/social activities: pub or social club (average correlation=.048±.003). 11.7% of "other sociodemographics" variables are associated with decelerated aging across aging dimensions on average, with the association being not receiving any allowance for either attendance, disability or mobility (average correlation=.056±.025). 10.0% of household variables are associated with decelerated aging across aging dimensions on average, with the three most frequent associations being the average total household income before tax (average correlation=.079±.037), the number of vehicles in the household (average correlation=.052±.019), and the number of persons in the household (average correlation=.039±.014).

We list the three socioeconomic variables categories most associated with decelerated aging in each specific aging dimension in Supplementary Table 33. For example, decelerated cognitive aging is most associated with education, “other sociodemographics” and social support. Specifically, 37.5% of education variables are associated with accelerated cognitive aging, with the three largest associations being qualifications: college or university degree (correlation=.078), qualifications: A level/AS levels or equivalent (correlations=.061), and qualifications: O levels/GCSEs or equivalent (correlation=.051). 14.3% of “other sociodemographics” variables are associated with accelerated cognitive aging, with the association being not receiving any allowance for either attendance, disability or mobility (correlation=.039). 11.1% of social support variables are associated with accelerated cognitive aging, with the association being leisure/social activities: sports club or gym (correlation=.031).

### Discussion

#### Correlations

##### Phenotypic correlation between accelerated aging dimensions

###### The low average phenotypic correlation between aging dimensions is partly explained by high model variance and limited prediction accuracy

We expected that the accelerated aging correlation between biologically similar aging dimensions would be higher. For example, the hips and the knees are joints that tend to be used jointly during physical activity and we therefore expected that their accelerated aging correlation would be higher than .8, but we found that they only had a correlation of .360±.005. Similarly, both serum assays and urine assays are biochemical assays which include biomarkers of metabolism and kidney function[173], but we found that these two accelerated aging dimensions had a correlation of .090±.004. The relatively low correlations between related accelerated aging subdimensions (.247±.135) should however be interpreted while taking into account (1) the weak performance of some of the predictors, (2) the high variance of chronological age predictors, and (3) the small and sometimes negative correlations already reported in the literature between some of the accelerated aging dimensions[174–176]. We discuss these three factors in more detail below.

First, some of the unexpectedly weak correlations between a priori similar aging dimensions could be explained by the small prediction performances of the models. The low correlation between biochemical blood and urine aging (.090±.040) can, for example, partly be explained by the weak performance of the model predicting chronological age as a function of the urine biomarkers ($R^2$=10.4±1.0%), which itself can be explained by the limited number of urine biomarkers available in UKB (four of them: creatinine, microalbumin, potassium and sodium). Re-estimating our models in a cohort covering a different set of blood and urine biomarkers would likely affect the correlation between these two aging dimensions.

Second, our chronological age predictors are highly variable. For example, we found that two models built on different anatomical views of the heart (three-chamber view: $R^2$=82.1%; four-chamber view: $R^2$=81.0%) have only a correlation of .672±.003. More generally, we found that the models trained on different views or using different features from the same biological dimension were only .541±.161 correlated. Similarly, the correlation between models built on the same images, using different preprocessing methods (e.g. raw vs. contrasted images) was only .658±.083. Further, different algorithms trained on the same dataset also tend to yield different predictors (mean correlation .838±.120). For example, the correlation between accelerated heart aging defined by two different CNN architectures (InceptionResNetV2: $R^2$=70.4%; InceptionV3 $R^2$=70.0%), both trained on the same heart MRI images (“four-chamber” view, raw preprocessing) is only .723±.002.

Finally, the correlations we observed are significantly larger than the correlations reported by Li et al.[174]. The correlations between the nine biological age residuals reported in Li et al. are between 0.35 and -0.32, with 75% of the 36 pairwise correlations being smaller than 10% in absolute value and more than half being negative. Four of the nine biological age definitions reported by Li et al. are based on DNA methylation (Horvath, Hannum, PhenoAge and GrimAge). The average correlation between these four dimensions is only 11%, and one of these six pairwise correlations (0.35, -0.02, <0.01, 0.11, 0.13, 0.09) is negative.

In this context, our finding that accelerated hip aging and accelerated knee aging had a correlation of .36±0.5 suggests that the aging process of both joints is strongly linked. We draw similar conclusions for other accelerated aging dimensions pairs whose correlation is higher than 25%.

##### Explaining the high phenotypic correlation between liver and pancreas accelerated aging

The phenotypic correlation between accelerated liver aging and accelerated pancreas aging is particularly high (.539±.004). We hypothesize that this correlation may be driven by the shared anatomical features that surround these organs (and therefore shared in the MRI images). The similarity between the anatomical features highlighted by the attention maps in the liver and pancreas images, as well as the high correlation between these two accelerated aging dimensions led us to classify them as two subdimensions of “Abdomen aging”. Based on the MRI attention maps, we do not believe that the pancreas and liver aging dimensions are strongly organ specific (Supplementary Figure 29 A and B and website: Models interpretability - Images). Instead, we hypothesize that the CNN is capturing the general aging of the abdominal cavity, its different organs and surrounding tissues.

Similarly, we hypothesize that images-based age predictors also capture aging of the tissue surrounding the organ. For example, different image-based accelerated aging dimensions (carotid ultrasounds, heart MRIs, liver MRIs, pancreas MRIs, full body X-rays, spine X-rays, hip x-rays, knee x-rays) tend to be more correlated on average. For example, accelerated aging of the connective tissue surrounding the liver, the pancreas, the heart and the musculoskeletal system is likely correlated, which increases the correlation between these predictors (see Feature importances - Images for these aging dimensions on the website). This explanation is coherent with the fact that accelerated eye aging is only weakly correlated with the other images-based accelerated aging dimensions (5-11%).

##### Correcting for the bias in the residuals prevents an artificial inflation of the correlation between the different aging dimensions

We observed a bias in the residuals when predicting chronological age. Younger participants tend to be predicted older than they are while older participants tend to be predicted younger than they are. Correcting for this bias before evaluating the performance of the models would artificially increase the performance. Indeed, estimating the bias for a specific sample to correct for it requires knowing the chronological age for this sample, which is precisely what the model is trying to predict. On the other hand, when estimating biological age as a phenotype to assess its genetic and environmental correlates, not correcting for this statistical bias in the predictions could lead to spurious results. For example, all accelerated aging dimensions would be artificially positively correlated because they would share a similar bias. This is because the default behavior for most algorithms when they cannot extract relevant information from the predictors is to predict the mean of the target variable over the training set, to minimize the mean squared error. As the algorithm learns to leverage the predictors, it progressively moves away from this initial prediction. For predictors that only captured a small percentage of the age variance (e.g. urine biochemistry: $R^2$=10.4±0.1%; blood cells: $R^2$=12.5±0.1%), we feared that the predictions were only marginally different from these initial baseline predictions and therefore almost identical between the two models, which would have spuriously increased the correlation between accelerated urine biochemistry aging and accelerated blood cell aging, independently of the biological correlation between the two aging processes. By correcting for the bias in the residuals, we remove from all prediction sets the statistical artefact that artificially increases the correlation between the different dimensions. After correction, we therefore report a correlation of only 19.8±0.2% between accelerated urine biochemistry aging and accelerated blood cells aging.

#### Genetic correlation between accelerated aging dimensions

We found that the main aging dimensions are genetically .104±.149 correlated and that their associated subdimensions are genetically .338±.255 correlated on average, which shows that biologically similar aging dimensions tend to be more correlated. For example, heart anatomical and electrical aging are .508±.089 correlated, whereas brain cognitive and musculoskeletal aging are uncorrelated (.067±.079). The highest significant genetic correlation was between abdominal liver aging and abdominal pancreas aging. This large correlation can be explained by the fact that liver and pancreas abdominal MRI images are highly similar, and that the aging phenotypes learned from them rely more on the shared surrounding tissue than on the actual organs. Perhaps surprisingly, brain anatomical aging and brain cognitive aging are genetically uncorrelated (.019±.056) and similarly for arterial aging and musculoskeletal aging (-.031±.158), lung aging and musculoskeletal aging (-.026±.040), and physical activity aging and musculoskeletal aging (.044±.227). Scalar data-based musculoskeletal aging (anthropometry, impedance, heel bone densitometry and hand grip strength) is also less genetically correlated with the four X-rays-based musculoskeletal agings (full body, spine, hip and knee) than we expected (.126±.098).

We compared the genetic correlations to the phenotypic correlations and found that they were highly correlated (.781 for the correlations between the aging main dimensions and .936 for the aging subdimensions). This high correlation suggests that the phenotypic correlation between the different accelerated aging dimensions is driven by the genetic correlation between these phenotypes. We observed exceptions. For example, accelerated brain cognitive aging and accelerated hearing aging are significantly more genetically correlated (.743±.070) than they are phenotypically correlated (.351±.005). These genetic correlations could help identify causal links between the different aging dimensions[177].

#### Correlation between aging dimensions in terms of association with biomarkers, clinical phenotypes, diseases, family history, environmental variables and socioeconomics

We found that the average correlation between main aging dimensions is .643±.142 in terms of associated clinical phenotypes, .639±.180 in terms of environmental variables, .607±.309 in terms of socioeconomics, .488±.198 in terms of diseases, .359±.229 in terms of biomarkers, and .030±.519 in terms of family history. For each X-category, the average correlation between accelerated aging subdimensions is larger than the average correlation between accelerated aging main dimensions, showing that biologically similar aging dimensions tend to be similarly associated with non-genetic factors.

##### XWAS results highlighting general aging factors

We found a large number of non-genetic factors associated with accelerated aging across aging dimensions, suggesting that there is a general aging process that affects all aging dimensions.
For example, we found that general health, and in particular general health rating and suffering from a long-standing illness, disability or infirmity, are two X-variables among the most strongly associated with accelerated aging across aging dimensions (average correlations of respectively .099±.036 and .073±.032)). The causality of this association likely goes both ways: accelerated agers probably suffer from lower general health, since aging is associated with the onset of numerous age-related diseases. Conversely, it is plausible that suffering from long-standing illness is taxing for the body and leads to accelerated aging across most aging dimensions.

This hypothesis is supported by our finding that poor cardiovascular health is associated with accelerated aging across aging dimensions. For example, on average and across aging dimensions, 66.7% of blood pressure biomarkers (e.g. systolic blood pressure, average correlation=.093±.083) and 40.5% of arterial stiffness biomarkers (e.g. position of the shoulder on the pulse waveform, average correlation=.073±.065) are associated with accelerated aging. Similarly, on average and across aging dimensions, 81.8% of breathing clinical phenotypes (e.g. shortness of breath walking on level ground, average correlation=.062±.029) and 59.8% of chest pain clinical phenotypes (e.g. chest pain due to walking ceases when standing still, average correlation=.041±.018) are associated with accelerated aging. Finally, on average and across aging dimensions, the disease category most associated with accelerated aging is cardiovascular disease (e.g. hypertension, average correlation=.074±.031). This association between poor cardiovascular health and accelerated aging in other dimensions could be causal. For example, hypertension leads to MRI-observable changes in the brain[178,179], and Sun et al. also reported that participants who suffered from hypertension and diabetes had a brain biological age 3.5 years higher on average[180].

We found that looking younger than one's actual chronological age is associated with decelerated heart aging, musculoskeletal aging and physical activity aging. Perceived age is an ageing biomarker that is informally but widely used in clinical practice and has been shown to predict survival and to be associated with aging biomarkers such as physical functioning, cognitive functioning and leukocyte telomere length[181–183]. A GWAS was also performed on perceived age found that it is 14% GWAS-based heritable and genetically associated with 75 traits such as adiposity[130].

Environmental variables tend to be similarly correlated with accelerated aging across aging main dimensions (average correlation=.639±.180). It is likely that these associations are causal and that they positively or negatively affect overall health, which reflects on most aging dimensions. For example, smoking is the environmental subcategory most associated with accelerated aging across aging dimensions and affects not only lungs[184], but also blood vessels[185,186], heart[187–189], brain[190], eyes[191,192], hearing[193], abdominal organs[194] (such as liver[195], pancreas[196,197] and intestines[198,199]), musculoskeletal system[200], blood[201] and urine. Mamoshina et al. showed that smoking is associated with accelerated aging[201]. Similarly, alcohol consumption[202] affects the brain[203–205], lungs[206,207], blood vessels[208], heart[208–211], liver[212–214], pancreas[215,216], bones[217,218], muscles[219], blood[220–222] and kidneys[223]. There is limited evidence that alcohol consumption affects the eyes in the long term according to the literature[224–226], but we found significant associations between accelerated eye aging and alcohol consumption variables (e.g. average weekly beer plus cider weekly, correlation=0.031, p-value=1.1e-13, see websites for other examples). Similarly, we found significant associations between accelerated hearing aging and alcohol consumption (e.g. alcohol intake frequency, correlation=0.07, p-value=1.2e-292), despite limited evidence of this phenomenon in the literature[227,228]. Likewise, physical activity affects the brain, eyes, lungs, blood vessels, heart[229], abdomen (liver, pancreas), musculoskeletal system, blood and urine. Likewise, sleep quality[230], physical activity[231] and diet[232] affect health across different biological dimensions.

Socioeconomic variables (social support, education, household, employment and other sociodemographics) tend to be similarly correlated with accelerated aging across aging main dimensions (average correlation=.607±.309). This is coherent with the discrepancies in life-expectancy between socio-economic classes. Specifically in the US, the richest 1% males live on average 14.6±0.2 years longer than the poorest 1% males, and the richest 1% females live on average 10.1±0.2 years longer than the poorest 1% females[233], which suggests that wealth is associated with decelerated aging and poverty is associated with accelerated aging. We found education to be more strongly associated with accelerated/decelerated aging than employment and household, possibly suggesting health literacy[234] as a key factor to explain the link between socio-economic status and aging rate.

#### XWAS results highlighting the multidimensionality of aging

The average correlation between aging main dimensions for each X-category and X-subcategories is far from being one, and the distribution of X-correlations is wide (large standard deviation). This suggests that, although there are general aging factors associated with accelerated aging in main dimensions as described above, there are also aging dimension specific factors, which affect most strongly a specific aging dimension. For example, hypertension is the disease most significantly associated with accelerated aging across aging dimensions, but accelerated eye aging is "enriched" in associations with eye related diseases such as cataract, retinal disorders and glaucoma, compared to other aging dimensions. Similarly, accelerated knee aging is associated with knee diseases such as knee arthrosis and internal derangement of the knee.

Furthermore, we found that some X-variables are associated with accelerated aging in one aging dimension and associated with decelerated aging in another. For example, playing computer games is associated with accelerated eye aging (correlation=-0.38) and with decelerated brain cognitive aging (correlation=.125). This finding is coherent with the fact that screen time can strain the eye (computer vision syndrome)[235], and that video games can possibly slow age-related cognitive decline[236,237]. Similarly, we found that being shorter in one's youth is significantly associated with accelerated aging in five aging dimensions (eye, hearing, lung, PWA-based arterial and musculoskeletal (scalars biomarkers-based)), whereas being taller in one's youth is significantly associated with accelerated aging in 11 other aging dimensions (heart anatomical (MRI-based), heart electrical (ECG-based), abdomen, liver, pancreas, musculoskeletal, spine X-rays, hip X-rays, knee X-rays, physical activity and blood cells). Finally, we found discrepancies between aging dimensions in terms of association with weight change over the last year. A stable weight during the last year was exclusively associated with decelerated aging, in six aging dimensions (eye (OCT-based), lung, arterial, musculoskeletal (scalar biomarkers-based)). Gaining weight was mostly associated with decelerated aging (four dimensions: musculoskeletal, full body X-rays, knee X-rays, and urine biochemistry), but also with accelerated aging in two dimensions (lung and arterial (PWA-based)). In contrast, weight loss was mostly associated with accelerated aging (ten dimensions: heart anatomical (MRI-based), abdomen, liver, pancreas, musculoskeletal, full body X-rays, spine X-rays, hip X-rays, knee X-rays and urine biochemistry), but also with decelerated aging in two dimensions (hearing and lung). One plausible explanation for this perhaps surprising observation can be found in the obesity paradox[238]. Alternatively, it is also possible that while being overweight is associated with poor general health, losing a significant amount of weight within a year is often caused by a serious disease. Wannamethee et al. for example reported that, in contrast to intentional weight loss, unintentional weight loss was associated with increased all-cause mortality[239], which can often be explained by disease[240].

Finally, we found that accelerated aging in a particular aging dimension could not be successfully predicted by biomarkers from a different dimension, highlighting the multidimensionality of aging. We observed a similar trend when predicting accelerated aging from clinical phenotypes: clinical phenotypes related to an aging dimension could more successfully predict accelerated aging in this dimension than other clinical phenotypes. For example, eyesight clinical phenotypes predicted accelerated eye aging (based on all biomarkers) with a $R^2$ of 13.1%, and breathing clinical phenotypes predicted accelerated lung aging with a $R^2$ of 10.3%, contrasting with the low average $R^2$ (4.8±4.2%) over all aging dimensions and clinical phenotypes categories.

#### Limiting factors of the XWAS

##### Confounding factors

Socioeconomic status is an important confounding factor for some of the associations. For example, we found that working longer hours was associated with decelerated heart aging (correlation: .057). This might sound counterintuitive, since long working days are associated with stroke[241], poor sleep quality and mental health[242], as well as depression, weight gain, smoking and drinking[243], all of which tend to be associated with accelerated aging in our XWAS. This paradox can likely be explained by the fact that unemployed participants are considered to work zero hours and are likely accelerated agers as an indirect consequence of their socioeconomic status, as explained above. These observations suggest that performing a new XWAS while correcting for additional factors such as socioeconomic status would likely affect the findings and their interpretation.

Similarly, taking a cholesterol-lowering medication is positively associated with accelerated heart aging, despite the fact that drugs, such as statins, are taken to improve cardiovascular health[244] and could therefore be seen as a heart rejuvenating therapy. This finding can be explained by the fact that participants taking statins likely already suffer from high cholesterol and poor cardiovascular health, making them heart accelerated agers. Their heart age might decrease after taking cholesterol lowering medications, but they remain accelerated agers compared to the participants who do not need to take cholesterol lowering drugs.

##### Bias due to the sample size

Similarly to the GWAS, the difference between the number of exposures associated with each aging dimension is largely driven by the difference in sample sizes. For example, accelerated heart aging and accelerated heart anatomical aging are .991±.000 correlated, but because of the smaller sample size when taking the intersection between the heart MRIs and the ECGs, the PEWAS identified nine exposures associated with accelerated heart aging versus 32 for accelerated heart anatomical aging.

#### Correlation between aging dimensions in terms of association with biomarkers, clinical phenotypes, diseases, family history, environmental variables and socioeconomics

##### Limiting factors of the XWAS

###### Confounding factors

Socioeconomic status is an important confounding factor for some of the associations. For example, we found that working longer hours was associated with decelerated heart aging (correlation: .057). This might sound counterintuitive, since long working days are associated with stroke[241], poor sleep quality and mental health[242], as well as depression, weight gain, smoking and drinking[243], all of which tend to be associated with accelerated aging in our XWAS. This paradox can likely be explained by the fact that unemployed participants are considered to work zero hours and are likely accelerated agers as an indirect consequence of their socioeconomic status, as explained above. These observations suggest that performing a new XWAS while correcting for additional factors such as socioeconomic status would likely affect the findings and their interpretation.

Similarly, taking a cholesterol-lowering medication is positively associated with accelerated heart aging, despite the fact that drugs, such as statins, are taken to improve cardiovascular health[244] and could therefore be seen as a heart rejuvenating therapy. This finding can be explained by the fact that participants taking statins likely already suffer from high cholesterol and poor cardiovascular health, making them heart accelerated agers. Their heart age might decrease after taking cholesterol lowering medications, but they remain accelerated agers compared to the participants who do not need to take cholesterol lowering drugs.

###### Bias due to the sample size

Similarly to the GWAS, the difference between the number of exposures associated with each aging dimension is largely driven by the difference in sample sizes. For example, accelerated heart aging and accelerated heart anatomical aging are .991±.000 correlated, but because of the smaller sample size when taking the intersection between the heart MRIs and the ECGs, the PEWAS identified nine exposures associated with accelerated heart aging versus 32 for accelerated heart anatomical aging.

#### Comparison between our models and the literature in terms of prediction accuracy

##### Recommended considerations when comparing the prediction performances between different datasets

###### $R^2$ vs. RMSE

When comparing different chronological age predictors it is crucial to take into account the age range of the datasets on which their performance was evaluated, as wide age ranges will yield higher R-Squared [$R^2$] values, while shorter age ranges will yield lower root mean squared error [RMSE] values.

First, we provide intuition as to why obtaining high $R^2$ values is more easily achieved on datasets with wide age ranges. As observed in the formula $R^2 = 1 - \frac{sum\ of\ squares\ of\ residuals}{total\ sum\ of\ squares}$, high $R^2$

values are reached when the prediction of the model is significantly more accurate than predicting the mean of the target distribution. Let us assume that a predictor can predict age with an error of ±5 years. Such a predictor can efficiently distinguish participants who are twenty years old from patients who are eighty years old. If the age range is 20-80 years, the model will therefore be able to explain a large percentage of the variance and yield a high $R^2$ value, as the average residual absolute value will be approximately five years, whereas predicting the mean of the chronological age distribution (30 years) for every sample will give an average residual value of approximately fifteen years assuming a uniform distribution. In contrast, if the age range is 40-50 years, predicting the mean of the distribution (55 years) for every sample will give an average residual absolute value of approximately 2.5 years. Using the same predictor with an error of ± 5 years could give a negative $R^2$ value. The reasoning above does not take into account the squaring of the errors or the fact that a model trained on a dataset with an age range of ten years is unlikely to yield a predictor with a mean absolute error of five years, but it provides intuition as to why a model built on the same set of biomarkers will reach higher $R^2$ values when its performance is evaluated on datasets with a larger age range.

Second, we provide intuition as to why lower RMSE values are more easily obtained on datasets with narrow age ranges. If a model is only provided with useless predictors, it will learn to minimize the RMSE by predicting the mean of the distribution. For example, a linear regression will set the coefficient for all the predictors to values close to zero and set the bias term to equal the mean of the chronological distribution. If the age range of this distribution is 20-80 years, a model without informative predictors at its disposal will predict a chronological age of 30 years for every sample which will give a mean absolute error of 15 years assuming a uniform chronological age distribution. As the model is provided with more informative features as predictors, it will leverage them to progressively affine its prediction and decrease its mean absolute error [MAE]. If the age range of the chronological age distribution is 40-50 years, the mean absolute error of a model built on the same set of non-informative features will be 2.5 years. The calculation to compute the RMSE rather than the MAE is more complicated because of the non-linearity, but the reasoning above provides intuition as to why lower RMSE values are more easily obtained on datasets with narrow age ranges.

This concept is illustrated in a publication that leveraged 356 laboratory biomarkers of the National Health and Nutrition Examination Survey[245–252] to predict chronological age. NHANES covers ages from one to 85 years old, and the model built on the whole cohort gave a $R^2$ value of 92% and a mean absolute error [MAE] of 4.76 years. Sagers et. al also built submodels on different age categories. The models built on the age range 45-64 and on the age range 65-85 gave lower MAE values (respectively 2.20 and 4.30 years) which could be mistakenly interpreted as the sign of better performing models. However, the models also gave lower $R^2$ values (respectively 77% and 25%). The MAE values were decreased by the narrower age range, while the initial larger $R^2$ value of 92% of the model built on the full dataset was a consequence of the larger width of the age range covered.

The age range covered by the UKB cohort is 37 to 82 years, which is typically shorter than the age range covered by other datasets. For example, NHANES[245–252] covers ages from one to 85 years, and the ChestX-ray8 datasets[253] covers ages from one to 90 years.

##### Development vs. Aging

Predicting age may be "easier" depending on the age inclusion criteria. For example, height is a predictor of chronological age during youth and adolescence[254], but its correlation with chronological age is reduced during adulthood. Each model has its own particularity based on the predictors included, but models trained on cohorts with an age range of 0-19 years (children and teenagers) provide more accurate estimates of chronological age versus models built on adults.

For example, in a previous publication we predicted the chronological age of infants younger than three years old with a $R^2$ of 62.5%[72]. Pediatric hand bones can also be used to predict chronological age with a mean absolute error of less than five months; however, models such as BoneXpert can only be used for boys younger than 17 years old and for girls younger than 15 years old[106,107]. Sagers et al.[68], in their laboratory-based age predictor found the highest $R^2$ value for participants younger than 18 years old ($R^2$=0.94 for age group [1,18), $R^2$=0.44 for [18,45), $R^2$=0.77 for [45, 65), $R^2$=0.25 for 65+). When comparing several chronological age predictors, it is therefore important to take into account not only the width of the age range covered but also the age inclusion criterion of the cohort. We note that the UKB cohort's age range is 37-82 years which makes predicting chronological age on this dataset a more challenging task than a cohort that samples individuals from a wider age range.

#### Comparison with the literature by aging dimension

A summary of the comparison between the performance of our chronological age predictors and the predictors found in the literature can be found in Table 2.

##### Cognitive tests

Anatürk et al. used XGBoost[38] on cognitive tests collected on 537 participants with a mean age of 69±5 years to predict chronological age with a $R^2$ value of 9% and a RMSE of 4.77 years[100]. Using UKB's cognitive test data we predicted chronological age with a $R^2$ value of 37.9±0.4% and a RMSE of 5.95±0.02 years. Our $R^2$ value is four times higher than the one reported by Anatürk et al., which is likely driven by UKB's sample size, which is approximately six times larger than the one reported by Anatürk et al. (UKB: 31,720; Anatürk et al.: 537). The lower RMSE reported by Anatürk et al. can be explained by the narrower age range covered by their dataset, with a standard deviation of five years.

##### Brain MRIs

A large number of models have been built to predict chronological age using brain MRI images[77–93]. We selected two of them, based on three-dimensional CNN architectures, to compare our prediction performance.

The first was built by Cole et al. on 2,001 samples aged between 18 and 90 years old and yielded a $R^2$ value of 92% and a RMSE of 5.31 years[78]. In contrast, our ensemble model for a population between ages 37-82 years gave a $R^2$ value of 76.4±0.3% and a RMSE of 3.73±0.01 years. Our lower $R^2$ value suggests that Cole et al.'s model outperformed our model, while our lower RMSE value suggests the opposite. This apparent contradiction can be explained by the significantly larger age range covered by Cole et al.'s dataset (18-90 years) (see Discussion - Performances - Comparison between our models and the literature in terms of prediction accuracy - Recommended considerations when comparing the prediction performances between different datasets - $R^2$ vs. RMSE).

A fairer comparison can be made between our model and the model built by Jonsson et al. as they used transfer learning to test its accuracy on 12,395 UKB samples aged 46-79 years. Their three-dimensional (3D) CNN architecture gave a MAE of 3.63 years and a $R^2$ value of 61.4% after being re-trained on the UKB dataset[79]. This $R^2$ value is significantly lower than the one we obtained with our ensemble model ($R^2$=76.4±0.3%). This is perhaps surprising because, unlike Jonsson et al.'s model which used a 3D scan, our model is based on three two-dimensional sliced images of the brain in the three anatomical planes. Even without ensembling, we found that our 2D models built on the three brain anatomical planes outperformed the model reported by Jonsson et al. (Sagittal plane: $R^2$=61.9±0.3%, Coronal plane: $R^2$=62.6±0.3%, Transverse plane: $R^2$=64.3±0.3%). One plausible explanation is that relying on 2D images allowed us to leverage deeper and more complex architectures. The architectures we built on heart videos for example had to be kept significantly shallower and simpler (see Methods - Machine learning algorithms - Videos).

##### Eye fundus images

Chronological age was predicted on UKB using eye fundus images with a $R^2$ value of 74±1% by Poplin et al.[129]. We found that our ensemble model outperformed this prediction accuracy ($R^2$=76.6±0.2%). One potential explanation for this difference is that Poplin et al.'s model was trained on only 48,000 UKB patients, whereas our model was trained on 90,000 participants. Poplin et al.'s model was however also trained on 236,000 EyePACS[255] images, so it is unclear if the sample size benefited their model or ours and/or from the different CNN architectures. For example, our InceptionV3 model slightly outperformed their model ($R^2$=75.2±0.1%), whereas our InceptionResNetV2 model significantly underperformed ($R^2$=69.1±0.2%).

More recently Nagasato et al. used transfer learning to train a VGG16 architecture on a far smaller sample size (n=85) using ultra-wide-field pseudo-color eye fundus images collected from participants aged 57.5±20.9 years to predict chronological age. However, they did not report their R-Squared value, only the slope of the regression coefficient between their predictor and chronological age (standard regression coefficient=0.833)[256].

##### Carotid ultrasounds and pulse wave analysis

Fedintsev et al. predicted chronological age from a linear combination of scalar features extracted from carotid artery duplex scan and pulse wave velocimetry and collected over 303 participants aged 23-91 years. They obtained a $R^2$ value of 55% for males and 69% for females, and a RMSE of 6.91 years for males and 5.87 years for females[74]. We approximate the performance of this model on the general population using a weighted average between males (104 participants) and females (199 participants) for comparison purposes: $R^2$=64.2%; RMSE=6.23 years We predicted chronological age using carotid ultrasound images ($R^2$=64.8±0.6; RMSE=4.44±0.04 years), pulse wave analysis data ($R^2$=41.3±0.2%; RMSE=6.53±0.01 years) and blood pressure ($R^2$=22.3±0.1%; RMSE=7.34±0.01 years). The ensemble model built on these three predictors gave a $R^2$ value of 67.1%±0.6% and a RMSE of 4.29±0.04 years. We obtained a lower RMSE value, which is partly explained by the narrower age range covered by UKB. We also obtained a larger $R^2$ value despite the narrower age range covered by UKB. This better performance of our model can be explained by the larger sample size and the different data modalities over which we built our ensemble model. However, Fedintsev et al.'s model performs very similarly to ours despite being built on 26 times fewer samples (303 vs. 7,797) and only using a small number of scalar features. This suggests that the features reported by Fedintsev et al. (medium intima thickness, percent of the stenosis, arterial stiffness) are reliable aging biomarkers that efficiently summarize the biological features observed in the carotid ultrasounds and the pulse wave velocity.

##### Electrocardiograms

Attia et al. predicted chronological age from 499,727 12-lead ECGs with a $R^2$ of 70% and an average error of 6.9±5.6 years[138]. The difference between the $R^2$ value of our ensemble model (37.5±0.6%) and their $R^2$ of 70% can be partly explained by the significantly larger age range that their cohort covers (18-100 years). However, despite the wider age range, their model obtained a similar average error to ours (RMSE=5.9±0.02 years). Their model outperformed our model, which can probably be largely explained by the difference in sample sizes (499,727 samples for the Mayo Clinic digital data vs. 44,417 samples for UKB). One possible way to improve the prediction accuracy of ECG based chronological age prediction on UKB would be to use transfer learning[257] to leverage the model developed by Attia et al. and to retrain it on the UKB cohort.

ECGs were also leveraged by Wiggins et. al to build a bayesian age classifier with an area under the curve [AUC] of 84.75%[139].

##### Full body X-rays

Langner et al. predicted chronological age with a testing $R^2$ of 82% and MAE of 2.47±1.91 years after training the VGG16 CNN architecture[258] on 32,323 UKB's full body Dual-energy X-ray absorptiometry [DXA] images[103]. We found that our preprocessing and the architectures that we used did not outperform their model individually. However, we found that our ensemble model yielded a statistically significant higher final testing $R^2$ of 85.7±0.1% (RMSE=2.85±0.01 years). We also found that adding to this ensemble predictions generated on spine DXA images ($R^2$=74.6±0.2%), hip DXA images ($R^2$=69.0±0.3%), Knee DXA images ($R^2$=69.0±0.3%) and scalar musculoskeletal features such as anthropometry, heel bone densitometry and hand grip strength ($R^2$=25.9±0.1%) yielded a chronological age predictor with a $R^2$ of 87.6±0.1%.

Karargyris et al. used chest X-rays and the DenseNet121 CNN architecture to predict chronological age with a $R^2$ of 90%[104]. After testing several architectures on the ChestX-ray8[253] dataset using transfer learning we found that the best architecture was indeed DenseNet121, which yielded the same $R^2$ of 90%. We also found that our ensemble model predicted chronological age with a $R^2$ of 91%. These $R^2$ values are slightly higher than the one we obtained on the UKB datasets (85.7±0.1% for the full body DXA images), but as previously explained this difference in prediction accuracy is largely driven by the difference in age ranges. Figure 1 of the publication shows that the age range for the ChestX-ray8 dataset is 1-90 years, which includes the childhood and the teenagerhood during which significant anatomical changes happen, compared to 37-82 years for UKB.

##### Hip X-rays

Wittschieber et al.[259] used a linear regression on features extracted from 643 pelvic X-rays collected from participants aged 10-30 years and obtained a $R^2$ value of 38%. Similar work was performed on the pelvis[105], the iliac crest[260,261] and the ilium[262]. Our model ($R^2$=69.0±0.3%; RMSE=4.20±0.02) significantly outperformed the one developed by Wittschieber et al. despite our age range not covering teenagerhood years, which can be explained by the 65 times larger sample size we leveraged.

##### Knee X-rays

Two chronological age predictors were built on knee X-ray images collected from young participants[263,264]. O'Connor et al. used a linear regression on features extracted from 221 knee X-rays collected from participants aged 9-19 years and obtained a $R^2$ value of 77.5-81.5%[263]. Tang et al. used a linear regression on features extracted from 503 knee X-rays collected from participants aged 6-19 years and obtained a $R^2$ value of 88-90%[264]. Fan et al. used a linear regression on features extracted from 322 knee X-rays and MRIs collected from participants aged 11-30 years and obtained $R^2$ values ranging from 44.1% to 65.4%[265] and other researchers performed similar studies[266–269]. We trained CNNs on 79,477 knee X-ray images collected from participants aged 45-82 and obtained a $R^2$ value of 69.0±0.3% and a RMSE value of 4.20±0.02 years. The significantly higher accuracy of the two first models described above can be explained by the age range over which they were trained and tested, as the human body changes significantly faster during development. We are, to our knowledge, the first to build a chronological age from knee X-rays for the older adult population.

##### Physical activity: daily life

Other investigators have built a survival model on UKB's wrist accelerometer data[149] predicting all-cause mortality, but we are, to our knowledge, the first to predict chronological age using this dimension/data modality. We compare our performance to two chronological age predicting models built on NHANES's wrist accelerometer data. These two publications are the ones from which we adapted the CNN architectures to build our own pipeline (see Methods - Machine learning algorithms - Time series - Physical activity)[140,141].

First, Pyrkov et al. predicted chronological age on 7,454 samples with an age range of 6-84 years and obtained a $R^2$ value of approximately 56% (Pearson correlation: 75%) and a RMSE of 14.0 years using a CNN[141].

Second, Rahman and Adjeroh predicted chronological age on 4,268 samples with an age range of 18-84 years with a RMSE of 15.74 years[140]. Rahman and Adjeroh also reported $R^2$ values as high as 0.93. We want to be very careful in raising this point, but we suggest there might have been an error in the code when computing the $R^2$ values for some of the models, and we provide four pieces of evidence to support this suggestion. (1) The order of magnitude for the $R^2$ values does not match the other metrics (Table 10 in Rahman et al's publication). For example, the training RMSE, correlation and $R^2$ values for the 1D CNN model are respectively 18.81 years, 52% and 88%. For a linear regression, the training $R^2$ value is the square of the training Pearson correlation, which gives us an order of magnitude of 27% rather than the 88% reported. (2) The testing MAE (15.49 years), RMSE (18.81 years) and correlation (45%) values reported show that the prediction accuracy on the testing set is lower than on the training set, as expected. However, the testing $R^2$ value reported is 93%, higher than the training $R^2$ value (88%). Similar anomalies can be observed for the other architectures (DNN, ConvLSTM, CNN+LSTM). (3) Sagers et al. reported a similar testing $R^2$ value of 92% on NHANES using laboratory biomarkers[68]. They, however, reported a corresponding MAE value of 4.76 years for a comparable age range that was more than three times lower than the testing MAE reported by Rahman and Adjeroh for their 1D CNN (MAE: 15.49 years, $R^2$=93%). (4) Rahman and Adjeroh themselves noticed the discrepancy in their results: “While 1D CNN has the best $R^2$ (0.93) followed by the DNN network ($R^2$=0.89), for MAE and correlation ConvLSTM* and CNN+LSTM model performed better.”. Based on the four aforementioned arguments, we will approximate Rahman et al’s testing $R^2$ value by the square of their best model’s testing correlation ($0.62^2$=38.4%) to compare the performance of their models with ours.

Our ensemble model predicted chronological age using wrist accelerometer data with a RMSE of 4.71±0.1 years and a $R^2$ value of 63.5±0.2%. Our RMSE is therefore approximately three times smaller than the RMSEs reported by Pyrkov et al. (RMSE=14.0) and Rahman and Adjeroh (RMSE=15.74), but our better performance is in part driven by the narrower age range covered by UKB (UKB: 43-79 years; Pyrkov et al.: 6-84 years; Rahman and Adjeroh: 18-84 years). Our model also outperformed the two NHANES based models in terms of $R^2$ values (our model: 63.5±0.2%; Pyrkov et al.: 56%; Rahman and Adjeroh: 38.4%) despite being built on a narrower age range.

We hypothesize that four factors can explain our better performance. (1) The sample size we leveraged from the UKB dataset after preprocessing and filtering the samples is twelve to twenty times larger the sample sizes that Pyrkov et al. and Rahman and Adjeroh could extract from the NHANES dataset (UKB: 90,000-100,000, NHANES: 4,268-7,454). (2) Unlike UKB, NHANES accelerometer data is a frequency measure that does not capture the absolute value of the acceleration. UKB data is also available at two different sampling frequencies (0.2 Hz and 100 Hz), which we both leveraged. (3) We found that our best predictive non-ensemble model was the GBM we trained on 2,271 scalar features ($R^2$=59.4±0.2%, RMSE=5.11±0.1 years). In contrast, our best non-ensemble model built on time series gave a $R^2$ value of 41.8±0.3% and a RMSE of 6.11±0.1 years. The models built by Pyrkov et al. and Rahman and Adjeroh rely on time series data compared to our scalar based model. (4) Building ensemble models increased our $R^2$ value from $R^2$=59.4±0.2% to 63.5±0.2%. Both Pyrkov et al. and Rahman and Adjeroh built several models but since, to our knowledge, they did not create an ensemble, we report the performances of their non-ensembled models. (5) We built both the CNN architecture designed by Pyrkov et al. and the ConvLSTM* architecture designed by Rahman and Adjeroh, which is their best performing architecture. We found that the architecture designed by Pyrkov et al. outperformed the architecture designed by Rahman and Adjeroh on the UKB dataset, which explains in part why we report higher prediction performances than Rahman and Adjeroh. Our better RMSE values than Pyrkov et al. (6.11±0.1 years vs. 14.0 years) despite using the same CNN architecture is likely driven by the narrower UKB's age, as well as reasons (1) and (2) mentioned above (sample size and difference in time series types).

##### Physical activity: Gait analysis

Gait changes as one gets older[270] and several research teams predicted chronological age from the gait[142,271–278]. Lu and Tan analyzed the gait of 1,870 silhouette gaits collected from participants aged 20-30 years to predict chronological age with a MAE of 3.02 years[274]. Makihara et al. analyzed 1,728 gait videos collected from participants aged 2-94 years and predicted chronological age with a MAE of 8.2 years[276]. Riaz et al. attached integrated measurement units to the lower back of 26 participants (mean age: 48.1 years old; standard deviation: 12.7 years) and recorded their gait. They then trained a random forest on the features extracted from these records and classified the participants into three age categories (age < 40 years, 40 years < age < 50 years and age > 50 years) with 88.82% accuracy[277]. Hoffmann et al. recorded the gait of 142 participants aged 13-82 years as they walked over a sensor floor. They then built a neural network and predicted chronological age with a MAE of 9.55 years and a $R^2$ value of 37.21%[142]. Li et al. built a support vector machine on features extracted from 63,846 gait energy image[279] recordings of participants aged 2-90 years. They predicted chronological age with a MAE of 6.78 years[278].

We analyzed the gait of the 77,269 UKB participants using wrist accelerometer recordings and predicted chronological age with a $R^2$ of 32.3±0.2% and a RMSE of 5.69±0.01 years. Only Hoffmann et al. reported a $R^2$ value (37.21%)[142], larger than the one we obtained (32.3±0.2%), which can be explained by the wider age range covered by their dataset. The RMSE we obtained is lower than the MAE values reported by the aforementioned publications, which can be explained by the narrower age range covered by UKB (37-82 years). Comparing the performance of our model with the ones reported in the literature is therefore difficult, but we hypothesize that the gait is more efficiently analyzed using gait videos than wrist accelerometer recordings and that Li et al.'s predictor is therefore likely more accurate than the one we built[278].

##### Laboratory biomarkers

We predicted chronological age using blood count biomarkers ($R^2$=12.5±0.01%, RMSE=7.59±0.01 years) and biochemistry biomarkers ($R^2$=50.7±0.3%, RMSE=5.81±0.02 years), which include blood biochemistry biomarkers ($R^2$=48.6±0.1%, RMSE=5.92±0.01 years) and urine biochemistry biomarkers ($R^2$=10.4±0.1%, RMSE=7.72±0.01 years). UKB's laboratory biomarkers (blood count, blood biochemistry and urine biochemistry) have previously been analyzed to predict survival[151,280], but we are, to our knowledge, the first to leverage these to predict chronological age. We built separate models on biochemistry biomarkers (blood and urine biochemistry) and blood cells biomarkers (blood count), which complicates our task of comparing the performances of our models to the literature, as most publications did not make this distinction. Below, we compare the results of our biochemistry-based chronological age predictors with models found in the literature that leveraged diverse laboratory biomarkers. The comparison between our blood biochemistry-based chronological age predictor, our urine biochemistry-based chronological age predictor and our blood cells-based chronological age predictors are treated independently and can be found subsequently.

We briefly summarize the results of five studies which predicted chronological age using laboratory biomarkers. (1-3) Three papers leveraged the NHANES[245–252] dataset to predict chronological age as a function of biochemical and blood count biomarkers[68,69,281]. (1) Sagers et al. used a random forest to predict chronological age from 356 biomarkers collected from 67,563 participants aged 1-85 years. They obtained a $R^2$ value of 92% and a MAE of 4.76 years[68]. (2) Kendiukhov built a neural network on 926 predictors measured in 86,311 samples aged 10-85 years and predicted chronological age with a $R^2$ of 78% and a MAE of 6.5 years[281]. (3) Wood et al. used XGBoost[38] on 39 biomarkers collected on 46,739 participants aged 19-85 years. They predicted chronological age with a Pearson correlation of 75% for males and 0.77% in females[69], which is approximately equivalent to a $R^2$ of 58%. (4) Putin et al. trained an ensemble of neural networks on 46 blood biomarkers from 62,419 blood biochemistry and cell count tests collected from patients aged 1-100 years and predicted chronological age with a $R^2$ of 80% and a MAE of 6.07 years[282]. (5) Wang et al. combined anthropometric measurements or vital signs with body fluid biomarkers. They leveraged 30 vital signs and 1,891 lab tests from 377,686 electronic medical records from patients of ages 18-85 to predict chronological age with a MAE of 7.58 years[75].

The diversity of the biomarkers covered by these models means that they should be compared to our models with caution; however, they still provide orders of magnitude for the prediction accuracy that can be reached when leveraging laboratory values to predict chronological age. Between the age predictors that we built, the model that is most similar to the ones found in the literature is the one that we trained on blood and urine biochemistry biomarkers. It predicted chronological age with a $R^2$ value of 50.7±0.3% and a RMSE of 5.81±0.02 years. We hypothesize that the underperformance of our model can be explained by (1) the greater the number of biomarkers included in the models found in the literature (n=46–1,921 compared to ours, 32), (2) the types of biomarkers leveraged by the different publications and (3) the differences in age ranges covered.

##### Blood biochemistry

We predicted chronological age with a $R^2$ value of 48.6±0.1% and a RMSE of 5.92±0.01 years using blood biochemistry biomarkers. Menni et al. predicted chronological age with a $R^2$ of 59% using 22 blood metabolites collected on 6,055 participants aged 17-85 years[283]. We hypothesize that the smaller $R^2$ value of our blood biochemistry model (48.6±0.1%) can be explained by the larger age range of the cohort leveraged by Menni et al.

##### Urine biochemistry

We predicted chronological age with a $R^2$ value of 10.4±0.1% and a RMSE of 7.72±0.01 using four urine biochemistry biomarkers. Hertel et al. used 59 urine nuclear magnetic resonance metabolomic biomarkers collected on 5,000 participants aged 20-79 years to predict chronological age with a $R^2$ values of 56% for men and 61% for women (respective RMSEs: 11.2 and 10.37 years)[66]. We hypothesize that our significantly smaller $R^2$ value is driven by both the narrower age range covered by UKB and the smaller number of variables included in our model (four vs. 59). Our RMSE is smaller (7.72±0.01 years) than the RMSEs reported by Hertel et al., which could spuriously lead the reader to believe that our model performed better, but this is another consequence of UKB's narrower age coverage.

##### Blood cells

We used blood count biomarkers to predict chronological age and obtained a $R^2$ value of 12.5±0.01% and a RMSE of 7.59±0.01 years. To our knowledge, we are the first to predict chronological age from blood cell biomarkers. Alpert et al. built a predictor of survival based on the age trajectories of immune biomarkers (IMM-AGE) collected from 135 participants but did not directly predict chronological age[169]. A scatter plot showing the correlation between IMM-AGE and chronological age in the Framingham Heart Study [FHS] dataset (age range 40-90 years)[284] can be found in Extended Data Fig.10b of their paper, but no correlation coefficient is reported. Both the correlation of our predictor and IMM-AGE with chronological age are modest (estimated $R^2$=10±5% based on the figure). Because both models seem to be comparatively accurate despite IMM-AGE benefiting from the wider age range covered by the FHS dataset and considering that our model was specifically trained to predict chronological age, we hypothesize that our model slightly outperforms IMM-AGE as a chronological age predictor.

###### Chronological age-based organ age predictors

Other organ age indicators have been designed. For example, the concept of lung age[285] was developed to encourage patients to stop smoking, and data from the participants of the Framingham Heart Study[286] was used to develop the concept of heart age[284]. We do not compare the performance of our models with these indicators because they are not chronological age predictors but calibrated on disease or behavior risk. Further, they define organ age as the participant's age minus or plus a given number of years based on a list of criteria. For example, lung age is chronological age adjusted for spirometry measures[285], and heart age is chronological age modified for HDL levels, total cholesterol, blood pressure, smoking habits and diabetic status[284].

##### Data modalities not covered by UKB

We did a brief literature review on age predictors capturing aging dimensions not currently assayed in UKB participants to highlight the unexplored zones of the multidimensionality of aging map and as a possible source of inspiration for future data modalities collection.

###### DNA methylation

The most widely known and accurate chronological age predictor is the DNA methylation clock[46]. Since its discovery by Horvath in 2013 using blood-derived DNA, much work has been done to improve its precision. Zhang et al. for example used linear regressions to build chronological age predictors on 13,661 participants aged 2-104 that gave RMSE values as low as 2.04 years and $R^2$ values as high as 96% (estimated from their reported Pearson correlations)[47]. They also showed that near perfect chronological age prediction was theoretically possible, but that the association between the biological age derived from the prediction and mortality decreases as the prediction accuracy approaches one.

##### Telomere length

Telomere length shortens with life and has therefore been used as an aging biomarker[3,7,8]. Telomere length has for example been used to suggest that shift work leads to accelerated aging[2]. It has been shown that telomere length is also associated with age-related diseases[1,4–6] and survival[9]. Telomere shortening is correlated but distinct across different tissues, suggesting a multidimensionality of telomere aging[287].

##### Transcriptomics

Transcriptomic measures (e.g., genome-wide gene expression) have been used to predict chronological age[59–61]. Fleischer et al. trained an ensemble of linear discriminant analysis classifiers [LDA] on 133 dermal fibroblast RNA-seq profiles collected from participants aged 1-94 years to predict chronological age with a $R^2$ value of 81% and an MAE of 7.7 years[59]. Mamoshina et al. trained a neural network on 545 muscle transcriptomic samples collected from participants aged 19-89 years and predicted chronological age with a $R^2$ value of 83% and a MAE of 6.19 years[60]. Harries et al. showed that they could classify young participants (aged < 65 years) from old participants (aged > 74 years) with an AUC of 95% using only six RNA probes from 698 samples[61].

##### Proteomics

Proteomics measurements have also been used to predict chronological age[62–64]. For example, Lehallier et al. recently trained a LASSO[34] on 2,978 plasma proteins collected from 3,301 participants aged 18-76 years. The LASSO selected 491 proteins and predicted chronological age with a $R^2$ value of 92% (estimated from the Pearson correlation) and a RMSE value of 2.44 years[63].

##### Microbiome

Chronological age predictors were built on the microbiome[71–73] from an array of body locations, including the gut and oral cavities. For example, Galkin et al. trained a neural network on 1,165 gut microbiome samples collected from participants aged 20-90 years to predict chronological age with a $R^2$ value of 29±9% and an MAE of 5.91 years[71].

##### Brain electroencephalography

Brain electroencephalography [EEG] signals were used to predict chronological age[94,180]. Sun et al. engineered features from EEGs collected from 4,506 participants aged 18-80 years and used a linear regression to predict chronological age with a $R^2$ value of 67.24% and a MAE of 7.6 years[180]. Zoubi et al. engineered features from EEGs collected from 468 participants aged 18-58 years and used a support vector regression [SVR] with radial kernel to predict chronological age with a $R^2$ value of 37±6% and a RMSE of 8.46±0.59[94].

##### Eye iris and corner images

Sgroi et al., Erbilek et al. and Rajput and Sable[288–290] predicted chronological age from iris images. Sgroi et al. trained a random forest on 630 scalar texture features extracted from 596 sample iris images to classify participants aged 22-25 years versus participants older than 35 years with an accuracy of 64%. Erbilek et al. built an ensemble of five simple geometric features extracted from the same iris images dataset. Using what they refer to as the sensitivity negotiation method, inspired by game theory, they obtained a classification accuracy of 75%. Rajput and Sable trained CNNs on 2,130 iris images collected from 213 participants aged 3-74 years and predicted chronological age with an MAE of 5-7 years[290].

Bobrov et al. trained CNNs on 8,414 eye corner images collected from participants aged 20-80 and predicted chronological age with a $R^2$ value of 90.25% (estimated from the Pearson correlation) and a MAE of 2.3 years[123].

##### Facial features

In 1972, Damon predicted chronological age with a MAE of 5.88 years by building a linear regression on 51 facial (ear breath and length, nose breadth, grayness of hair, baldness) and body (sitting height, abdominal depth, bideltoid breadth, hand grip) anthropometric measurements collected from 600 men aged 20-80 years[101]. Similarly, and more recently, Dayanithi et al. predicted chronological age from ear morphometry[124].

With the recent advances of machine learning and computer vision, more complex models have been trained to predict chronological age from wrinkle features[122], eye corners[123] and full facial pictures[111–121,125–128,273]. For example, Rothe et al. trained a VGG16[258] architecture on 523,051 face images and predicted chronological age with a MAE of 2.68 years on the MORPH[291] benchmark dataset (sample size: 55,134, age range: 16-77 years)[113].

##### Hand X-rays

Several researchers leveraged hand X-ray images to predict chronological age in infants, children, and teenagers during the last six years[102,106–110,292,293]. Westerberg for example trained the Xception[294] architecture on 12,811 images from patients aged 0-19 years and predicted chronological age with a MAE of 1.007 years[108].

##### Other skeletal structures

Other skeletal structures have been leveraged to predict age such as the clavicles epiphyses[295–297], the fourth cervical vertebra[298], the mandibular ramus length[299] or the molars[300–302].

##### Physical activity: stress testing

Chronological age was also predicted as a function of exercise stress testing performance, but we were unable to access the publication[143].

#### Comparison between the models' performance in terms of algorithms and data modalities

##### Algorithms that consider linear vs. non-linear feature representations

We found that the non-linear algorithms (GBM and neural network) outperformed the elastic net - a linear model - for 40 models out of the 41 models (98%) trained on scalar features. The GBM outperformed the elastic net by 4.7% on average in terms of $R^2$ (Elastic net=20.2%, GBM=24.9%) which suggests that part of the age variance is captured by non-linear effects and complex interactions between the features.

##### Raw data vs. pre-processed features

We found that the models built on the "extracted" or pre-processed scalar features did not perform as well as the deep learning models built on the corresponding raw time series or images. This suggests that some of the information was not captured by the pre-processed features and building models on the raw signals is preferred for these data modalities. For example, our GBM predicted chronological age from the PWA scalar features (e.g. positions of the pulve wave peak and notch) with a $R^2$ of 22.9±0.2%, whereas a CNN built on the raw time series gave a $R^2$ of 41.0±0.2%, explaining almost twice the variance of chronological age compared to the scalar and pre-processed features-based model. Clinical uses of the PWA scalar features could benefit from integrating CNN models to extract more information from the signal[303]. Similarly, the CNN built on raw ECGs signal (GBM: $R^2$=11.1±0.4%; CNN: $R^2$=37.6±0.4%) significantly outperformed the GBM built on ECG scalar features (e.g. pulse rate and length of the different segments), explaining more than three times more of the variance in chronological age than the pre-processed features-based model.

For some aging dimensions, features extracted from the raw time series or images by the CNN demonstrated complementarity with the scalar features input by the elastic nets, GBMs, and dense neural networks. For example, the ensemble model built on anatomical brain anatomical features (dMRI weighted means, subcortical volumes and gray matter volumes) gave a $R^2$ value of 64.7±0.3%, whereas the ensemble model built on both the brain images and the anatomical scalar features gave a $R^2$ value of 75.3±0.2%.

Finally, we found during our preliminary analysis that some aging dimensions did not benefit from building CNNs on the raw data. For example, we built a one-dimensional CNN on the raw spirometry data, which presents as a time series. This model did not outperform the GBM built on the scalar features (peak expiratory flow, forced vital capacity and forced expiratory volume in one second, $R^2$=23.6%). We hypothesize that this can be explained by the shape of the spirometry curves, which are highly similar for all the individuals and can be efficiently summarized by the three scalar features.

##### Performance of ensemble models

We found that ensembling several architectures built on the same data modality increased the performance of the predictions. For example, when predicting chronological age as a function of knee X-rays, the InceptionResNetV2 and the InceptionV3 architecture obtained $R^2$ values of

64.9±0.3% and 65.7±0.3% respectively. When ensembling these two models, we predicted chronological age with a $R^2$ value of 69.0±0.03, increasing the best $R^2$ value by 3.3%.

Similarly, we observed that building ensemble models on different data modalities significantly increased the performance of the prediction. For example, the best model built on a single musculoskeletal modality was the ensemble model built on the full body "figure" (showing the skeleton as well as the silhouette of the participant) X-ray images, which predicted chronological age with a $R^2$ value of 81.9±0.2%. By building an ensemble model on all the musculoskeletal modalities-based predictors (other full body X-ray images, hip, knee and spine X-ray images, anthropometry, impedance, heel bone densitometry and hand grip strength measures) we predicted age with a $R^2$ value of 87.6±0.01%, a 6.7% improvement on the previous best $R^2$ value.

For some aging dimensions, ensembling different data modalities only yielded marginal gains of $R^2$ value. For example, the predictor built on heart MRI videos and images predicted chronological age with a $R^2$ value of 85.3±0.1%. Adding the ECG-based predictors ($R^2$=37.5±0.5%) only improved the prediction performance by 0.3% ($R^2$=85.6±0.2%).

#### Analysis of the bias in the residuals

We observed a bias in the residuals for all models. Participants on the younger end of the cohort's chronological age distribution tend to be predicted older than they are, whereas participants on the older end of the distribution tend to be predicted younger than they are. This can be observed in other publications, as well[68,138,282]. It could be tempting to interpret this bias as a sign that young people tend to be accelerated agers and that old people tend to be decelerated agers. We however concluded that this observation is solely a statistical artifact after making the following four observations. (1) The bias is relative. If a model is trained on participants aged 20 to 50, the participants aged 50 years will tend to be predicted younger than they are. If we train a second model on participants aged 50 to 80, the same participants aged 50 years will tend to be predicted older than they are. No age group is intrinsically accelerated aging or decelerated aging, the bias for an age group depends on the position of the age group relative to the cohort on which the model was trained. This can for example be observed in the models trained on different age groups by Sagers et al.[68]. (3) A similar pattern is observed for all aging dimensions. (3) The bias gets smaller as the model's accuracy increases. (4) We predicted other quantitative variables such as serum cholesterol and blood pressure, and we found a similar bias pattern. Aycheh et al. also interpreted the bias as a statistical artefact and hypothesized that it was the result of a sample size imbalance between the different age groups[95]. We refuted this hypothesis by training models on datasets with different chronological age distributions (e.g. uniform, Gaussian), as the bias was largely unaffected by these preprocessing steps. We therefore formulate a hypothesis that is coherent with the five aforementioned observations. The default behavior of a model to minimize the loss function (mean squared error) if it is not provided with informative predictors with respect to the target is to predict the mean of the target variable for every sample. Therefore, every participant that is younger than the mean cohort age will be predicted older than they are, and inversely for the older participants. The bias will also be proportional to the difference between the age of the participant and the mean age, which is what we observe in our models. As the model starts extracting relevant knowledge from the predictors, it must balance the potential improvement for the new prediction with the safety of its initial default guess, which behaves like a Bayesian prior, "pulling" every sample's prediction towards the mean target value on the dataset. Therefore, the bias remains approximately proportional to the difference between the participant's chronological age and the cohort's mean age, while progressively decreasing in amplitude as the $R^2$ increases.

Some other researchers who built biological age predictors by training models to predict chronological age reported this bias[95,103,304], but, to the best of our knowledge, most did not, although it can be observed in the scatter plot when one is present in the publication[59,63,68,78,80,92,94,96,138,140,142,144,282]. In some publications the bias does not seem to be present[60,64,71,98,141], one explanation being that, as the $R^2$ value of the model increases, it becomes harder to discern it. The fact that most researchers do not mention this bias does not necessarily mean that they did not notice it, but considering that it represents an important hazard for all the association studies with mortality and diseases down the road when not corrected, we recommend that researchers report it when they observe it in the future.

One key point we would like to highlight here is that it is crucial to correct for the bias in the residuals when defining accelerated aging before performing the association with survival, diseases, biomarkers or other phenotypes. If that is not done, older participants' biological age will tend to be lower than their chronological age, making them decelerated agers. In contrast, younger participants will tend to be accelerated agers. Because older participants are at a higher risk for death than younger participants, not accounting for the bias will likely lead to spurious and counterintuitive findings, such as decelerated aging being associated with increased mortality.

#### Survival prediction performance

We found that only 18 biological ages out of 331 (5.4%) significantly outperformed CA as survival predictors after Bonferroni correction, and none significantly underperformed. The average difference in CI between biological age and chronological age was small (0.020) but significant (p-value=0.03). The biological age that significantly outperformed CA the most (elastic net trained on biochemistry biomarkers) improved the CI by only 0.028±0.007. Most biological ages therefore represent a negligible improvement over CA as survival predictors, which we hypothesize can possibly be explained in the following ways. (1) Aging and survivability are different phenotypes. Aging affects all organs, whereas death is usually linked to the failure of one or several vital organs such as the brain, the heart, the lungs, the kidneys or the liver. This could for example explain that accelerated eye aging and accelerated hearing aging are not better predictors of survival than chronological age. We however find this hypothesis unconvincing for two reasons. First, accelerated hearing aging is correlated with accelerated aging in other dimensions linked to vital organs such as brain cognitive aging (.351±.005) and lung accelerated aging (.283±.002). Similarly, accelerated eye aging is for example correlated with accelerated heart aging (.100±.012). Second, accelerated aging in a non-vital organ can indirectly lead to increased mortality. For example, eye aging is associated with a greater likelihood to fall, and falls are the leading cause of accidental death among older adults[305,306]. Therefore, we expected that accelerated aging in all dimensions would be significantly associated with increased mortality. (2) Building on (1), aging and survival do not happen on the same time scale. It is for example possible that aging can be better understood as the slow, early onset and progressive wear and tear on the body, whereas death is associated with the sudden onset of an event that will jeopardize the function of one or several vital organ(s) most commonly occurring during old age. The two processes are not disconnected. For example, atherosclerosis leads to the progressive stiffening of arteries that happens through life and is associated with hypertension. This process begins during childhood and can therefore be leveraged to predict chronological age and build an arterial age predictor, but the life-threatening conditions associated with atherosclerosis will not manifest themselves until middle age and will represent a significantly greater threat during the elder years[307]. Consequently, being a ten-years accelerated arterial ager at the age of 82 (biological age of 92) might be associated with a significant increase in mortality risk, whereas being a ten-years accelerated ager at the age of 37 years (biological age of 47) might only be associated with a comparatively modest increase in mortality risk. It is therefore plausible that the lack of association between aging and survival we observe is driven by the relatively young age of the participants and that recomputing the CI on an older cohort would increase the added value of biological age predictors as survival predictors. (3) Some of the models we trained predicted chronological age with RMSE values as low as 2.26±0.04 years. For such highly accurate models, the difference between biological age and chronological age will be small for most participants, which could explain the limited added value of biological age predictors compared to chronological age when predicting survival. The decreased association between accelerated aging and mortality as CA predictors become more accurate has been reported for DNA methylation-based[47] and physical activity-based chronological age predictors[141]. However, we find this explanation unconvincing because the 331 chronological age predictors cover RMSE values between 2.26 years and 8.72 years, with a mean of 5.92 years and a standard deviation of 1.53 years. Similarly, the 11 age predictors based on the main aging dimensions cover RMSE values between 2.65 years and 7.60 years with a mean of 4.80 years and a standard deviation of 1.82 years.

Other researchers also found a limited association between accelerated aging and survival: a systematic review and meta-analysis on DNA methylation-based age predictors and mortality risk concluded that the association was inconclusive[48]. The DNA-methylation based clock corrects for the bias in the residuals (see Analysis of the bias in the residuals), but, to the best of our knowledge, other association studies did not, which likely strongly hindered the robust discovery of associations between accelerated aging and other phenotypes.

### Interpretation of the models

#### Analysis of the features selected by the algorithms with respect to the literature

##### Brain

We found that attention maps highlighted diverse regions of the brain, which is consistent with the systemic brain changes that happen with aging (atrophy, white matter hyperintensities, impaired white matter integrity, microbleeds, small vessel changes)[308] and the fact that the scalar predictors' feature importance was distributed between the different features, with most features being considered important by at least one of the algorithms. This overall suggests that no specific part of the brain is more significantly affected by aging than the others.

Our finding that brain cognitive aging is associated with reduced processing speed and fluid intelligence is coherent with the literature[309,310]. Age-related changes in processing speed are linked to changes in the cerebellar and the prefrontal cortex[311] and we therefore expected that accelerated brain cognitive aging and accelerated brain anatomical aging would be more strongly correlated (correlation: .113±.007).

##### Eyes

The fact that eye age prediction is driven by spherical power and that the elastic net's regression coefficient for this feature is positive is coherent with the fact that, with age, presbyopia becomes more prevalent[312]. Presbyopia –also called age-related farsightedness– is a consequence of the loss of the ability for the eye to accommodate to focus on nearby objects and it corresponds to positive spherical power. Similarly, cylindrical power is a measure of astigmatism and increases with age[313–318], which explains its selection by the GBM and the positive regression coefficient in the elastic net.

Our attention maps highlighted the eye fundus vascular features, which is consistent with Poplin et al.'s attention maps [144]. Ege et al. reported that with age, eye fundus images tend to become more yellow because of optimal imperfections in the refractive media[319], which was possibly leveraged by our neural networks and could explain why some non-vascularized regions were also highlighted. Finally, our attention maps highlighted most retinal layers in the OCT images.

The diversity of the features highlighted by the different models is coherent with the fact that age-related changes occur in all ocular tissues[320].

#### Hearing

Presbycusis, or age-related hearing loss, can be diagnosed with hearing tests[321]. During hearing tests, different frequencies are played to the patients, who must press stop after they start hearing the sound. The volume of the sound being played increases over time, so the longer it takes the patient to press the button, the stronger their hearing loss. Considering this, it is intuitive that the GBM we trained on hearing tests highlighted the total duration of the test and the time it took the participant to press next as the most predictive features of chronological age.

#### Lungs

The strength of respiratory muscles such as the diaphragm decreases with age, which reduces the peak expiratory flow (PEF), the forced vital capacity (FVC) and the forced expiratory volume in one second (FEV1). The support tissue that keeps the airways open during expiration also loses its integrity with age, which reduces the FEV1. The alveoli lose their elasticity, which makes expiration more difficult[322]. These factors explain why the elastic net assigned a negative regression coefficient to PEF, FVC and FEV1 when predicting chronological age from spirometry measures.

#### Arteries

With age, atherosclerosis develops, so intima media thickness increases [323] and arterial vessels stiffen, which leads to an increase in systolic blood pressure and decrease in diastolic blood pressure[324]. Kostis et al. reported no change of resting heart rate with age[325], but in the UKB cohort resting heart rate is negatively correlated with age (-.048±.018). These observations explain the feature importances reported by the GBM as well as the sign of the elastic net regression coefficients for the models built on arterial scalar features (positive for systolic blood pressure and carotid intima media thickness, negative for diastolic blood pressure and heart rate). The attention maps showed that the CNNs trained on short axis carotid ultrasound images (a cross-sectional view of the carotid) do not leverage the arterial features such as the carotid artery or the jugular vein, but instead rely on the superficial skin tissue. In contrast, the attention maps for the models trained on long axis carotid ultrasound images (a longitudinal view of the carotid) do highlight the carotid arteries. The difference in prediction performances (short axis: $R^2$=34.7±0.8%; long axis: $R^2$=47.1±0.8%) suggests that approximately 75% of "arterial age" prediction is actually driven by the surrounding tissue.

##### Heart

As discussed above, blood pressure is strongly affected by aging, which explains its importance as a predictor for heart age. The ECG R axis and PQ interval are respectively negatively (-.165±0.02) and positively (.161±.026) correlated with age which explains their selection by the models.

The attention maps for the models trained on heart MRI images and videos highlighted the interventricular septum, which can be explained by the fact that its thickness increases with age[326] and that a sigmoid-shaped interventricular septum is part of normal aging[327]. The attention maps also highlighted the mitral and tricuspid valves, which undergo significant changes with age[328], such as calcification. Calcification is rare for the tricuspid valve[329], but common for the mitral one[330]. The attention maps highlighted the aorta as well, whose composition, stiffness, thickness and diameter change with age[331,332]. Finally, the attention maps showed that the models trained on heart MRI images did not exclusively rely exclusively on heart features to predict chronological age, but also on the surrounding tissue and anatomical features. This particularly affected the two-chamber view images, for which the attention maps highlighted the whole thoracic cavity with a particular emphasis on the spine, which explains the .047 higher phenotypic correlation between sagittal raw spine X-ray images-based accelerated aging and two-chamber view raw heart MRI images-based accelerated aging (correlation between spine-sagittal and heart-two-chamber view: .322±.005; three-chamber view: .275±.006; four-chamber view: .274±.007).

#### Abdomen

The attention maps of the models built on liver MRI images highlighted various abdominal regions including the liver, the stomach, the spleen, as well as muscle, bones and adipose tissue. The attention maps of the models built on pancreas MRI images highlighted similar features aside from the pancreas, including the liver. The similarities between liver-based and pancreas-based attention maps suggest that our models do not properly capture liver aging and pancreas aging, but instead capture general abdominal aging. The abdomen undergoes significant macroscopic changes as we age[333], which were likely leveraged by the convolutional neural networks. In terms of liver aging, it is known that liver function decreases with age[334] and that the liver ages at the cellular level, which is for example associated with low-grade inflammation[335]. There is less evidence that the liver undergoes clear macroscopic changes that could be captured by MRI images[336], but it has been reported that with age the color of the liver gets darker, blood flow decreases, liver volume decreases[337,338], and that the prevalence of liver diseases, such as nonalcoholic fatty liver disease, alcoholic liver disease, cirrhosis and fibrosis, increase with age[339], which might have been leveraged by our models to predict chronological age[340]. In terms of pancreas aging, age-related change visible on MRI images include pancreatic atrophy, fatty degeneration, and lobulation[341]. Finally, aging is also associated with abdominal changes in adipose tissue[342,343], muscles[344–346] and bones[347].

#### Musculoskeletal

With age, the body composition changes, as muscle mass decreases whereas fat mass increases[348–350], which explains why the models selected hand grip strength, waist circumference, weight, whole-body impedance and BMI as important CA predictors.

The attention maps of the models trained on knee X-ray images highlighted the joint, possibly relying on osteoarthritis[351,352]. Similarly, the attentions maps of the models trained on hip X-ray images highlighted both the joint and the proximal femur, which can possibly be explained by osteoporosis[353,354] and changes in the shapes of the femoral neck[355] and medullary cavity[356] with age. The attention maps of the models trained on spine X-ray images highlighted different vertebrae, which can be explained by osteoporosis and degenerative changes[357]. The lumbar spine was particularly highlighted and is known to suffer from disc degeneration, among other changes[347]. Finally, the attention maps of the models trained on full body X-ray images highlighted the neck, the upper torso, the hips and the knees. The highlighting of the neck can be possibly explained by the fact that the neck tilt and the cervical angle tend to increase with age[358] and the highlighting of the upper torso can be explained by increased osteoporosis and thoracic kyphosis with age, as well as changes in the ribs shapes and angles[359,360].

#### Biochemistry

Testosterone and sex hormone binding globulin levels decrease with age in men[361–363]. Alanine aminotransferase is a marker of liver injury that decreases with age[364]. Glycated hemoglobin is a marker of glycemic control and diabetes that increases with age[365]. Urine creatinine is a biomarker of kidney function which decreases with age as creatinine clearance declines [366]. Cystatin C is a biomarker of kidney function whose levels increase with age[367]. Apolipoprotein B is a protein involved in the transport of cholesterol and a biomarker for atherosclerosis whose levels increase with age[368]. Serum creatinine is a waste product of muscles whose levels increase with age[369].

Insulin-like growth factor 1 is an anabolic hormone involved in growth whose serum levels decrease with age[370]. Serum urea is a market of kidney function whose levels increase with age as reduced kidney function results in decreased urea excretion[371]. These observations explain the importance of the features selected by the models.

##### Blood cells

According to the literature red blood cell distribution width[372] and mean corpuscular volume[373], increase with age whereas red blood cell count[374], platelet crit[375], lymphocyte percentage[376] and hemoglobin levels[377] decrease with age. Mean sphered volume, mean reticulocyte volume and neutrophil count are positively correlated with age in UKB's cohort (respective correlations: .107, .099 and .022). These observations explain the importance of the features selected by the models.

#### Sex and aging rates

All but one (blood cells) elastic net models, the regression coefficient for sex when predicting CA is positive. A positive coefficient for the sex variable means that, all other biomarkers included in the model being kept equal, a male will be predicted to be older than its female counterpart. It is important to note that, perhaps counterintuitively, this suggests that males are decelerated agers, not accelerated agers. For example, if the regression coefficient for sex in a model build on arterial health is +5 years, it means that the arterial health of a 45-year-old male is similar to the arterial health of a younger 40-year-old female. Since females tend to live several years longer than males[378], we expected to find that males are accelerated agers compared to females. We hypothesize that the counterintuitive sign of the sex regression coefficients can be explained by the positive correlation between sex and age in the UKB cohort: the average male is 59.0±8.6 years old and the average female is 58.3±8.3 years old. It is therefore possible that the regularized linear regression (elastic net) relies on the sex coefficient to correct for this discrepancy while keeping the regularization cost to a minimum.

#### Differences in feature importances between the different models

For the models built on scalar features, we computed the correlation between the feature importances estimated in four different ways: (1) in a univariate, linear context, using the Pearson correlation between chronological age and each scalar predictor, (2) in a multivariate, linear context, using the regression coefficients of the elastic net model, (3) and (4) in a multivariate, non-linear context using the feature importance from GBMs (3) and neural networks (4). The correlations can be found in Supplementary Table 3 (Pearson) and Supplementary Table 4 (Spearman).

The average correlation between these four measures of feature importance was .48±.27 (Pearson) and .50±.19 (Spearman). This low average correlation should however be interpreted while keeping in mind that intra-model variance is not null, that is that the feature importances for the same algorithm trained on two similar datasets would be lower than 1.0. For example, GBMs rely on bootstrap aggregating (also known as bagging), which is a stochastic procedure. Therefore, the algorithm could generate two different forests despite being trained on the same dataset. The feature importances generated by a GBM also tend to be close to zero for many predictors, which means that the ranking of these predictors shows some variability and will decrease the Spearman correlation between the feature importances of two GBMs trained on identical datasets. Similarly, the elastic nets and the neural networks are trained using gradient descent which is a stochastic process and can result in the algorithm converging to two different local optima despite being trained on the same dataset. Additionally, elastic nets use ridge and LASSO to shrink the coefficients of variables that are either poor predictors or correlated with another good predictor. LASSO in particular tends to select one variable out of each group of highly correlated variables and to set the regression coefficient for the other variables to zero. On slightly different datasets, this procedure can result in the same algorithm selecting different variables as representatives of their clusters, which introduces important variations in the feature importances outputted by the model.

##### Univariate versus multivariate linear models

We found that, in a linear context, the average correlation between the feature importance in a univariate versus multivariate context is approximately .52±.25 (Pearson) and .471±.213 (Spearman), which can be explained by the difference of interpretation between the two measures of feature importance, as explained below.

In contrast to a raw Pearson correlation (univariate model), a linear regression coefficient (multivariate model) corrects for the effect of the other variables included in the model. For example, a typical exercise in machine learning is to predict the price of houses using features such as the number of rooms and the total surface area[379]. In a univariate context, the correlation between the number of rooms and the price of the house is positive, because the total surface area is a confounding factor: houses with a larger number of rooms tend to have a larger total surface area on average, which itself means that the house will tend be more expensive, on average. However in the multivariate context of a model that includes both the number of rooms and the total surface area, the interpretation of the regression coefficient for the number of rooms is not “How much does the price of a house increase on average if the house has more rooms?” anymore but, instead, “How much does the price of the house increase if more walls are built to split the total surface areas into a larger number of rooms, without increasing the total surface area?”. In the latter case, it is possible that some people will prefer a house with few large rooms rather than several small rooms, and the regression coefficient will therefore be negative or at least smaller in magnitude: the total surface area is “explaining away” the initial importance of the number of rooms. The inverse effect can be observed, too, as adding variables to a linear regression to correct for confounding factors can increase the importance of a feature that was initially or poorly correlated with the target variable, which is known as a Simpson's paradox[380].

Another difference between a correlation coefficient and a regression coefficient is that the regression coefficient takes into account the average effect of a change in the predictor value on the target variable but not the variation between samples. For example, if increasing the surface of the house by a square foot increases the price of $1,000 on average, the regression coefficient will have the same value in the two following cities: (1) a city in which the price of the house is solely determined by the surface area (Pearson correlation of 1.0 between the surface area and the price), and (2) a city in which the price of the house is determined by the surface area, after what a random integer N between 0 and 100 is generated and the price is increased by $1,000,000xN. In the second city, the average effect of the surface area on the price is still the same, so the regression coefficient is unchanged. In contrast, the Pearson correlation between the surface area and the price is now close to zero because the predictor only explains a negligible part of the variance of the target. Indeed, the Pearson correlation between two variables indeed measures how well one of the variables can be predicted from the other. For example, if the price of the houses is based solely on the surface area, with a base price of $1,000,000 plus one cent for each square foot of surface area, the correlation between the predictor and the target will be maximal (Pearson correlation of 1.0) since the price can be perfectly predicted from the surface area, even though the absolute effect of the surface area on the price is negligible. The small magnitude of the effect will be captured by the regression coefficient, which will take a small value. This second difference between the Pearson correlation and linear regression coefficient can however be accounted for by centering and scaling the target and the predictors, which we did. Therefore, the differences between univariate Pearson correlation-based versus multivariate linear regression-based feature importances should solely be interpreted by remembering that linear regression coefficients account for the confounding of the other variables included as predictors in the model.

We provide two examples, selected from the elastic net that we trained to predict chronological age from 53 blood biochemistry and demographic (sex and ethnicity) variables, and for which the correlation between feature importances based on Pearson correlations versus linear regression (elastic net) coefficients was .595 (Pearson correlation) and .491 (Spearman correlation). (1) Serum glucose levels is the 8th most correlated feature with chronological age (Pearson correlation: .109±.005), but it was only ranked as the 30th most important predictor by the elastic net. This can possibly be explained by the observation that the elastic net ranked glycated hemoglobin [HbA1c] as the 10th most important predictor. HbA1c is correlated with both chronological age (Pearson correlation: .200±.008) and blood glucose levels (Pearson correlation: .623±.005), and it is the 5th most correlated feature with chronological age, so it possibly explained away the importance of glucose as a chronological age predictor. Additionally, the elastic net is a regularized linear regression whose ridge and LASSO effects shrink the regression coefficient of the features it does not select, so the partial redundancy between blood glucose and HbA1c further explains why the former was deemed of little importance by the elastic net. (2) Cholesterol was ranked as the 8th most important feature by the elastic net, but it is only the 43rd most correlated predictor with chronological age (Pearson correlation: 0.016). This is an example of Simpson's paradox[380].

#### Linear versus non-linear multivariate models

We found that, in a multivariate context, the average correlation between the feature importance in a linear (elastic net) versus non-linear (GBM) context is approximately .6 (Pearson: .62±.21; Spearman: .55±.14), which can be explained by two limitations of a linear model compared to a non-linear model. (1) A linear model cannot capture non-linear relationships between a predictor and the target variable. For example, testosterone levels for males increase with chronological age during adolescence and decrease with chronological age in old age. If one tries to predict testosterone levels from chronological age, a linear model such as a correlation or a linear regression will be poorly suited as it can only estimate the average effect of getting older over the entire age range with a single regression coefficient. To capture this non-linearity and improve the prediction accuracy, one must either introduce quadratic terms in the linear regression or adopt more complex models such as a GBM. GBMs rely on trees, and they can therefore split the age range into several age leaves, which can then be used to model the non-linearity. (2) A linear model also cannot learn complex interactions between the variables such as “exclusive or” [XOR] types. That is, if we try predicting chronological age from two variables A and B in the case that older participants are participants who have either A and not B, or B and not A, a linear model such as a logistic regression will not be able to properly classify the participants without introducing interaction terms, whereas a tree-based algorithm will easily learn to leverage this pattern with a simple tree of depth two.

Inversely, if the relation between the predictor and the target is strictly linear, then a linear model such as a linear regression might be better suited to discover the relationship. A linear regression could for example easily learn to predict the perimeter of a circle from a radius of any value after being trained on a small number of datapoints. In contrast, a tree would have to be trained on a significantly larger number of data points to cover the space, it would not be able to predict the target as accurately on the testing set even after creating a leaf for every data point in the training set, and it would not be able to extrapolate what it learned to other radius ranges.

We provide two examples, selected from the elastic net and the light GBM that we trained to predict chronological age from 53 blood biochemistry and demographic (sex and ethnicity) variables, and for which the correlation between feature importances based on linear regression (elastic net) coefficients versus GBM feature importances was .601 (Pearson correlation) and .792 (Spearman correlation). (1) SHGB was ranked as the first most important predictor by the GBM. In contrast it was only ranked as the 26th most important predictor by the elastic net. This can possibly be explained by the fact that SHBG levels are positively correlated with chronological age for males (Pearson correlation: .269) and negatively correlated with chronological age for females (Pearson correlations: -.094), which could not be leveraged by the elastic net but might have exploited by the GBM. (2) In contrast, sex was ranked as the second most important predictor by the elastic net but was only ranked as the 30th most important predictor by the GBM.

#### Spurious identification of accelerated and decelerated agers due to low quality images

We found that low quality images were overrepresented in the most accelerated and decelerated patients. This can for example be observed in the most decelerated ager among older age male participants for transverse brain MRI images (Supplementary Figure 10) and in the most decelerated ager among older age male participants for contrasted liver MRI images (Supplementary Figure 20). This is most likely a statistical artefact: low quality images are rare, and the model cannot accurately estimate the chronological age of the patient, which makes it more likely that the prediction will either be significantly higher (spurious accelerated ager) or lower (spurious decelerated ager) than the target.

#### The symmetry in the attention maps for paired organs suggest that they age similarly

The attention maps highlighted very different features across participants. For example, brain MRI attention maps highlighted different brain regions (Supplementary Figure 10), and similar differences can be observed for attention maps based on eye fundus (Supplementary Figure 11), eye OCT (Supplementary Figure 12), carotid ultrasound (Supplementary Figure 13), heart MRI (Supplementary Figure 15 and Supplementary Figure 16), liver MRI (Supplementary Figure 20), pancreas MRI (Supplementary Figure 21), full body X-rays (Figure 8), spine X-rays (Supplementary Figure 22 and Supplementary Figure 23), hip X-rays (Supplementary Figure 24) and knee X-rays (Supplementary Figure 25). These variations could suggest that attention maps are a highly noisy tool to identify which features are driving the CNN predictions. However, and in contrast with this important variation across participants, the attention maps for paired organs (eyes, hips and knees) tend to be symmetrical, which implies that (1) attention maps are not as noisy as initially suggested above and (2) the left and the right-side organs tend to age similarly.

#### Identifying genetic variants associated with aging dimensions: heritability and genes linked to accelerated aging

The GWAS we performed on the accelerated biochemistry aging and accelerated blood biochemistry aging phenotypes identified the genes encoding SHBG and cystatin C as significantly associated with these aging phenotypes. This finding is intuitive considering that these two biomarkers were respectively ranked as the second and the sixth most important predictors by the model we trained to predict chronological age as a function of biochemical biomarkers. SHBG and cystatin C levels increase with age (respective correlations of .06±.01 and .30±.02) so accelerated biochemistry agers are likely to have higher SHBG and cystatin C levels than an average person of the same chronological age. Genes for which some alleles are associated with a higher/lower serum SHBG and cystatin C levels are therefore likely to be associated with accelerated/decelerated biochemistry aging by the GWAS. Similarly, the model ranked glycosylated hemoglobin [HbA1c] as the fourth most important predictor and glucose as the 14th most important predictors for CA, which possibly explains why the GWAS identified MPDU1, HK1 and G6PC2 as significantly associated with accelerated aging, since mutations in MPDU1 lead to glycosylation disorders[170] and both HK1 and G6PC2 are enzymes involved in the glucose pathway[170]. The GWAS also highlighted UDP glucuronosyltransferase genes, which are linked to liver function and more specifically to drug metabolism[170]. Deficiency in UDP glucuronosyltransferase can lead to increased drug hepatotoxicity, which increases alanine aminotransferase [ALT] levels[381]. ALT is a biomarker for liver damage and was ranked as the third most important predictor for chronological age by our model, which offers a potential explanation as to why the GWAS highlighted UDP glucuronosyltransferase genes as significantly associated with accelerated biochemical aging. Finally, the GWAS highlighted CETP (involved in cholesterol metabolism) and ABCB11 (involved in statins metabolism, which regulate cholesterol level), which can be explained by the fact that the blood biochemistry model included cholesterol, LDL, HDL and triglycerides as predictors (respectively ranked as 15th, sixth, 20th and 15th most important predictors).

Similarly to what we observed for accelerated biochemistry aging, the genes highlighted by the GWAS were frequently linked to the accelerated aging dimension on which it was performed. For blood cell aging, the GWAS highlighted TUBB1 (expressed in thrombocytes and megakaryocytes and linked to macrothrombocytopenia), HBS1L (involved in erythrocyte volume, hemoglobin content, erythrocyte count, platelet count and monocyte count, and linked to sickle cell disease and beta-thalassemia), SPTA1 (a component of red blood cell plasma membrane and linked to erythrocyte diseases such as elliptocytosis 2, pyropoikilocytosis, and type 3 spherocytosis), ANK1 (linked to spherocytosis), WDR66 (involved in mean platelet volume). For arterial (pulse wave analysis-based) aging, the GWAS highlighted ATP1B3 (involved in cell-surface interactions at the vascular wall) and NPR3 (involved in blood volume and pressure and linked to hypertension). For accelerated anatomical brain aging the GWAS highlighted CRHR1 (hypothalamic-pituitary-adrenal pathway), RP11-669E14.4 (linked to Alzheimer's disease), LRRC37A2 (linked to progressive myoclonic epilepsy-6 and Koolen-De Vries syndrome), KANSL1 (linked to Koolen-De Vries Syndrome and already associated with Alzheimer's disease risk by another GWAS[382]), MAPT (linked to diverse neurodegenerative disorders such as frontotemporal dementia, progressive supranuclear palsy, Alzheimer's disease, Pick's disease and cortico-basal degeneration), MAPT-AS1 (linked to Parkinson Disease and Koolen-De Vries Syndrome) and RUNX2 (involved in skeletal morphogenesis). For accelerated anatomical brain aging the GWAS highlighted C6orf136 (linked to Central Nervous System Germ Cell Tumor and Spinocerebellar Ataxia). For accelerated eye aging, the GWAS highlighted HERC2 (associated with eye pigmentation), TTK (linked to retinoblastoma), RDH5 (involved in visual pigment synthesis and linked to fundus albipunctatus and fundus dystrophy) and ABHD2 (linked to retinitis pigmentosa). For accelerated eye fundus aging, the GWAS highlighted HERC2 (associated with eye pigmentation), PPL (present in keratinocytes), PDE6G (involved in the phototransduction signaling cascade of rod photoreceptors) and TCF4 (involved in corneal dystrophy). For accelerated eye OCT aging, the GWAS highlighted ABHD2 (linked to Retinitis Pigmentosa), ARMS2 (a component of the eye's choroidal extracellular matrix, linked to age-related macular degeneration and retinal drusen), RDH5 (involved in visual pigment synthesis and linked to Fundus Albipunctatus and Fundus Dystrophy), RP1L1 (a retinal-specific protein involved in photosensitivity and rod photoreceptors' morphogenesis, and linked to Occult Macular Dystrophy and Retinitis Pigmentosa), TTK (linked to retinoblastoma) and OCA2 (linked to iris pigmentation). For accelerated anatomical heart aging, the GWAS highlighted TTN (linked to dilated cardiomyopathy and myofibrillar myopathy). For accelerated musculoskeletal aging, the GWAS highlighted TBX15 (linked to Cousin syndrome and acromegaloid facial appearance syndrome), CCDC91 (associated with ossification of the posterior longitudinal ligament of the spine and diffuse idiopathic skeletal hyperostosis) and AAGAB (linked to palmoplantar keratoderma). For accelerated full body musculoskeletal aging, the GWAS highlighted TBX15 (linked to Cousin syndrome and acromegaloid facial appearance syndrome) and FGFR1 (linked to osteoglophonic dysplasia and Hartsfield syndrome). For accelerated spine aging, the GWAS highlighted TNFSF11 (linked to osteopetrosis), SUPT3H (linked to dysostosis), SOX5 (involved in cartilage formation and linked to Lamb-Shaffer syndrome) and ESR1 (linked to osteoporosis). For accelerated hip aging, the GWAS highlighted SUPT3H (linked to dysostosis), CCDC91 (associated with ossification of the posterior longitudinal ligament of the spine and diffuse idiopathic skeletal hyperostosis) and TNFSF11 (linked to osteopetrosis). For accelerated knee aging, the GWAS highlighted CCDC91 (associated with ossification of the posterior longitudinal ligament of the spine and diffuse idiopathic skeletal hyperostosis). For accelerated musculoskeletal scalar features-based (anthropometry, impedance, heel bone densitometry and hand grip strength) aging, the GWAS highlighted GDF5 (linked to osteoarthritis, acromesomelic dysplasia, brachydactyly, proximal symphalangism, chondrodysplasia and multiple synostoses syndrome), GPR126 (involved in body height and linked to distal arthrogryposis), PKDCC (involved in development and linked to rhizomelic limb shortening with dysmorphic features), POLD3 (linked to Ruijs-Aalfs syndrome) and ZBTB38 (involved in height).

The GWASs also highlighted genes with no direct connection to the aging dimension on which the GWAS was performed. These findings can possibly be explained in four different ways. (1) The gene highlighted has a complex and unknown link to the aging dimension. For example, a large number of genes involved transcription regulation, nucleotide exchange factors and GPCRs with unknown end effects. As a specific example, half the genes (three out of six) associated with accelerated abdominal (liver view) aging were related to transcription: PNPT1 (involved in RNA metabolic processes), ZNF219 (a zinc finger protein that modulates chromatin’s transcriptional activation) and TSEN2 (involved in RNA splicing to remove introns). (2) The gene highlighted by the GWAS is in linkage disequilibrium with another gene, the other gene being linked to the aging dimension on which the GWAS was performed. For example, the most significant gene associated with accelerated biochemistry aging is RP11-218C14.2. RP11-218C14.2 is a pseudogene with unknown function but is in linkage disequilibrium with cystatin encoding genes (CST3, CST4, CST1, CST9, CST9L). (3) Accelerated aging in the dimension on which the GWAS was performed (let us call it dimension A) is correlated with accelerated aging in another dimension (let us call it dimension B). Therefore, the GWAS might detect genes associated with accelerated aging in dimension B, despite being performed on accelerated aging in dimension A. For example, the gene most significantly associated with accelerated hearing aging is OR2B4P, involved in smell perception. It is possible that accelerated smell aging (which we could not measure using the UKB cohort) and accelerated hearing aging are correlated, which would explain this finding. (4) The aging dimension might not solely rely on biomarkers related to its intended definition. This is particularly true for aging dimensions for which we trained the models on images. For example, heart MRI images capture the tissue surrounding the heart, so some of the genes detected by the GWAS might not be linked to heart aging but to adipose tissue, bones, muscles, skin and other tissue aging.

Finally, the GWAS highlighted pseudogenes, long intergenic non-coding RNAs and protein coding genes with unknown functions which are likely either in linkage disequilibrium with the causal gene (for example), or whose function still remains to be discovered. For example, accelerated blood cells aging is associated with AC020595.1 and RP11-202G18.1.

In conclusion, the genes whose biological connection to their associated aging dimension is clear provide insight about the way genetics can influence the aging rates of the different organ systems. In contrast, the genes that were highlighted by the GWAS but whose connection to their associated aging dimension is unclear provide insights about general factors of aging, the multidimensionality of aging, and are potentially promising target candidates for rejuvenating therapies.

#### Missing non-genetic variance

We found that we could not successfully predict accelerated aging from biomarkers, clinical phenotypes, diseases, family history, environmental variables or socioeconomic variables ($R^2$ values lower than 5% on average), so the majority of the environmental (nongenetic) variance identified by the GWAS remains unexplained. This can possibly be explained by four different factors. (1) We could only leverage a limited number of X-variables. Unlike the genetic information, which can be fully sequenced and stored, non-genetic variables are diverse and vary through life which makes it impossible to fully collect it. (2) All X-variables were not collected by UKB from the same participants and the sample size of their intersection was close to zero. This problem was strengthened after taking the intersection with the aging dimension on which the XWAS was performed. We therefore had to build a separate model for each set of X-variables. Leveraging the union of all the environmental variables on a large cohort of participants would have likely allowed us to explain a larger part of the environmental variance. (3) We estimate the percentage of variance explained by environment exposures using the testing $R^2$ obtained from the predictions outputted by a model that we trained to predict accelerated aging, which is different from the variance component analysis performed by the BOLT-REML software to directly estimate the variance explained by the genetic factors. Elastic nets, GBMs and neural networks allow us to identify which environmental variables play a key role in predicting accelerated aging. They can be used to output an accelerated aging prediction for new samples, but they likely underestimate the amount of variance that can be explained by environmental variables, compared to the restricted maximum likelihood [REML] method.

### Biological age predictors can be used to evaluate candidates rejuvenating therapies and suggest new anti-aging targets

#### Biological age predictors can be used to quantify the effect of rejuvenating drugs or environmental effects

Biological age predictors can be used to monitor the effect of rejuvenating therapies. For example, Elysium is using a DNA-methylation clock to monitor the effect of NAD+ as a rejuvenating drug in clinical trials[383]. Similarly, Fahy et al. used several DNA-methylation clocks to show that they could rejuvenate the immune system[384], Horvath et al. developed DNA-methylation clocks in mice to measure whether young blood plasma transfusion could reverse aging[55], and Lu et al. showed that DNA methylation age decreased when Yamanaka factors were used to reprogram mouse ganglion cells to restore vision[385].

In contrast, biological age predictors can also be used to measure the effect of environmental variables on the aging rates of different organ systems. For example, Horvath et al. used a DNA methylation clock to show that obesity accelerates liver aging[386].

##### Rejuvenating drug candidates

A large number of rejuvenating therapies are currently under development[387], leveraging diverse approaches such as systemic factors, metabolic metallic manipulation, senescent cell ablation, and cellular reprogramming[388]. Preclinical research is identifying new candidates as well[389]. Developing accurate age predictors for the different aging dimensions such as ours will be key to assessing the properties of each of these treatments. We briefly summarize some of them below.

##### Caloric restriction

Caloric restriction robustly increases lifespan and healthy lifespan in many species. The effect is likely mediated by the mechanistic target of rapamycin [mTOR] pathway, Insulin-like growth factor-1 [IGF-1] signaling and sirtuins[390]. It is also possible that diet and the microbiome affects cellular senescence by modulating the epigenome[391].

##### Rapamycin

Rapamycin, which targets the TOR signaling pathway, has been shown to slow general aging[392] and hearing aging[393] in mice and is a popular rejuvenating candidate[394,395]. Specifically, a prospective clinical trial showed that topical rapamycin slows skin aging in humans[396].

##### GH and IGF-1

Growth hormone [GH] and Insulin-like growth factor-1 [IGF-1] levels decrease with age. GH and IGF-1 mediate the anti-aging effects of caloric restriction and are potential rejuvenating drugs for the musculoskeletal, metabolic and cognitive aging dimensions, especially in males[397].

##### Metformin

Metformin is an antidiabetic drug that affects the IGF-1 signaling pathway and is a promising rejuvenating drug candidate[398–402]. Its use is associated with increased life expectancy[403,404], a decreased incidence of cancer[405], cardiovascular diseases, inflammation and is possibly slowing cognitive aging (it improves cerebral angiogenesis and neurogenesis in mice[406]) and musculoskeletal aging[407]. It also attenuates the hallmarks of aging at the cellular level[400].

##### Acarbose

Acarbose, like Metformin, is an antidiabetic drug that improves lifespan and health in mice by improving carbohydrate metabolism[408,409] and mimicking caloric restriction[410]. Acarbose has been tested in combination with Rapamycin to slow aging[411].

#### NAD+

Nicotinamide adenine dinucleotide [NAD+] is an enzyme mediating redox reactions that plays a key role in metabolism regulation. NAD+ levels decrease with age and cause defects in mitochondrial and nuclear functions, leading to age-related diseases. It is therefore hypothesized that NAD+ supplementation could slow aging and protect against age-related pathologies, such as neurodegenerative diseases[412]. The effect of NAD+ is possibly linked to caloric restriction through its activation of sirtuins[413], and its levels could, for example, be increased by taking a nicotinamide riboside supplementation[414].

#### Sirtuins

Sirtuins work in tandem with NAD+[412,415,416] and are also a potential target for rejuvenating therapies[417] that could mimic the rejuvenating effect of caloric restriction[417,418] and delay age-related telomere shortening[418]. Sirtuins could for example rejuvenate metabolic and cardiovascular age[419]. There is controversial evidence that sirtuins are linked to caloric restriction[420–423].

#### Resveratrol

Resveratrol activates the sirtuin gene Sirt1, which possibly mimics the benefits of caloric restriction[424,425]. Thaung Zaw et al. performed a pilot study in postmenopausal women and claim that Resveratrol could reduce cognitive aging by up to ten years[426].

#### AFT4

ATF4 has been shown to be altered in long-lived mice and is found in higher levels in mice treated with rejuvenating drugs candidates such as acarbose, rapamycin, caloric restriction or methionine restriction[427].

##### Yamanaka factors

Ocampo et al. showed that forced expression of the Yamanaka factors (Oct4, Sox2, Klf4, and c-Myc)[428] in mice lead to cellular reprogramming and pluripotency through epigenetic marks remodeling, which erases cellular markers of aging, ameliorates signs of aging and increases life expectancy[429]. Similarly, Lu et al. showed that Yamanaka factors (Oct4, Sox2, Klf4) can be used to restore youthful methylation patterns, which reverses vision loss in old mice[385].

##### Young blood serum transfusion

Old mice who received a blood plasma from young mice through heterochronic parabiosis[430] showed rejuvenation in bones[431], skeletal cells[432], liver[432], kidneys[433], blood vessels[434], brain and cognitive function[434–437].

##### 17-α-estradiol

17-α-estradiol protects against metabolic decline and inflammation and slows aging in male mice[438,439]. It does not affect other hallmarks of aging and its effect is distinct from caloric restriction[440].

##### Lonafarnib

Lonafarnib decreased mortality for patients with Hutchinson-Gilford progeria syndrome, a rare premature aging disease, making a potential rejuvenating drug candidate[441].

##### Telomerase reactivation

As cells undergo mitosis, their telomeres shorten until they reach the Hayflick limit[442]. As we age, our telomeres therefore shorten. Telomerase protects the telomeres by resynthesizing them during each mitosis. Reactivating it reverses tissue generation across multiple organs (testes, spleen and intestines)[443], which makes telomerase reactivation an attractive rejuvenating candidate strategy[444,445].

##### Thymus regeneration

Fahy et al. showed that a protocol designed to regenerate the thymus could decrease DNA methylation age by 1.5 years on average and decrease GrimAge -- a DNA methylation clock designed to predict survival[51] -- by two years. This rejuvenation is associated with improved immunological function and improved risk indices for several age-related diseases[384].

##### Hyperbaric oxygen

A randomized controlled trial showed that hyperbaric oxygen can slow cognitive aging by improving cerebral blood flow, leading to improved attention, information processing speed and executive functions[446].

#### Multidimensionality of rejuvenation

As we show in this paper, most aging dimensions are weakly correlated, and it is therefore likely that different rejuvenating drugs will affect the different aging dimensions differently.

Specifically, some of the candidate rejuvenating therapies aim to expand life expectancy or revert general aging, while others focus on a specific aging dimension.

Treatments targeting general aging include Yamanaka factors, caloric restriction and the drugs involved in its related pathway, such as Rapamycin, IGF-1, Metformin, Acarbose, NAD+, sirtuins, Resveratrol and AFT4[390]. For example, rapamycin appears to slow both heart, liver, adrenal glands, endometrium and tendon aging in mice[392]. Young blood serum transfusion also affects several aging dimensions simultaneously (bones, skeletal cells, liver, kidneys, blood vessels, brain and cognitive function), telomerase reactivation reverses tissue degeneration across multiple organs (testes, spleen and intestines) in mice and Lonafarnib decreases general mortality in progeria patients. Although these drugs may affect several aging dimensions, not all are affected to the same extent. For example, Shavlakadze et al. showed that mTORC1 inhibition in rats counter-regulates age-related gene activation such as c-Myc. Such a mechanism could potentially affect all aging dimensions, but Shavlakadze et al. found that the drug is more effective at rejuvenating the kidneys than the liver or skeletal muscles[447]. Similarly, dietary restriction was shown to improve lifespan but not cognitive aging in Drosophila melanogaster[448]. A multidimensional biological age predictor could be used to shed light on which aging dimensions are indeed affected by these treatments and quantify this benefit in years.

In contrast to the drugs targeting general aging, Thymus regeneration primarily rejuvenates the immune system, 17-α-estradiol's effects seem to be limited to metabolic age and inflammation levels, and hyperbaric oxygen was only shown to improve cognitive aging so far. A multidimensional biological age predictor could be used to identify other aging dimensions affected by the aforementioned treatments. For example, considering that hyperbaric oxygen improves cognitive aging by improving the cerebral blood flow in specific regions, it would be interesting to determine whether it also improves anatomical brain aging.

##### Limitations of biological age predictors when assessing rejuvenating therapies

A limitation of biological age predictors built by training a model to predict chronological age is that they are not guaranteed to be based on causal factors and that they are therefore vulnerable to "cheating". For example, a sigmoid-shaped interventricular septum wall is a common finding in older patients, but it is of little clinical significance[327]. A drug or surgical intervention aiming at remodeling the interventricular septum wall could therefore artificially decrease the biological age predicted by our heart MRI-based models and make spurious claims about improved heart health, despite being unlikely to lead to better heart-related health outcomes. This limitation of our biological age predictors should therefore be kept in mind, especially if they are used in clinical trials.

##### Identifying new rejuvenating drug candidates

Biological age predictors can provide insights about the aging mechanisms and suggest targets for new rejuvenating therapies. For example, the DNA methylation clock suggests that it might be possible to slow or revert aging by demethylating the CpG islands[58,449], which Lu et al. recently confirmed by using Yamanaka factors to restore vision in mice[385]. Similarly, the features highlighted as important by the models we trained to build the age predictors (see regression coefficients and feature importances for the models built on scalar predictors; see attention maps for models built on time series, images or videos) suggest potential targets to slow or reverse aging. Additionally, the genes identified by the GWASs are potential targets to reverse aging, as well. These could be targeted in the future using gene therapy as suggested by the recent rescue of Hutchinson-Gilford progeria in mice[450]. Finally, the environmental variables identified by the PEWAS can suggest potential rejuvenating lifestyles and treatments. With all these potential targets, one must keep in mind that correlation is not causation.

#### Future directions

As UKB will release new datasets, we plan to build new age predictors that will enrich the aging correlation map. We also plan to compute the genetic correlation between accelerated aging in the different dimensions and age-related disease traits, such as diabetes, heart disease and cancer and to follow up on the genes highlighted by the GWAS to identify potential new rejuvenating targets. We plan to leverage other datasets to validate our findings and compare our aging dimensions to other predictors such as the DNA methylation clock, as well.

Finally, we hope to adapt our pipeline to predict survival and estimate both the multidimensionality of mortality (Is being at a higher risk for cancer associated with being at a higher risk for other causes of deaths such as cardiovascular disease, after correcting for chronological age?) and the correlation between aging and survivability. Are accelerated agers also accelerated “diers”? That is, if we build a chronological age predictor and a survival predictor on the same dataset, will accelerated agers, according to the age predictor, also have a higher mortality risk factor according to the survival model? And are the features driving these two predictions the same? Our preliminary results however show that this task will prove challenging, as outperforming chronological age as a survival predictor is difficult. Specifically, we leveraged the 233,388 pulse wave analysis samples to predict survival and found that we could only outperform chronological by a non-significant 1.9% percent in terms of Concordance-Index (71.4±0.9% vs. 69.5±0.9%).

Alternatively, our pipeline could also be leveraged to predict diseases.

### Methods

#### Data types and Preprocessing

##### Demographic variables

First, we removed out the UKB samples for which age or sex was missing. For sex, we used the genetic sex when available, and the self-reported sex when genetic sex was not available. We computed age as the difference between the date when the patient attended the assessment center and the year and month of birth of the patient to estimate the patient’s age with greater precision. For physical activity, the age of the patient when they attended the assessment center was different from the age of the patient when they wore the wrist accelerometer, so we computed their age based on the date they started wearing the device. For ECG, we found 26 samples for which the date at which the participant attended the assessment center was not recorded in the main data frame, so we imputed it using data from the ECG files. We one-hot encoded ethnicity.

#### Scalar data

We define scalar data as a variable that is encoded as a single number, such as height or blood albumin level, as opposed to data with a higher number of dimensions, such as time series, images and videos. The complete list of scalar biomarkers can be found in Supplementary Table 6. We did not preprocess the scalar data, aside from the normalization that is described under cross-validation further below.

##### Hierarchical clustering of the predictors to select subcohorts that maximize sample size and predictors

As we will describe under Methods - Models ensembling, we built ensemble models to leverage the full breadth of the UKB dataset when predicting chronological age. These models rely on a linear combination of predictions outputted by different submodels (Table 1), which prevents them from learning more complex interactions between the features that were initially used to build the submodels. For example, the ensemble model for the aging main dimension “brain” and subdimension “cognitive” is built as a linear combination of the predictions outputted by the submodels built on sub-subdimension datasets such as “reaction time”, “matrix pattern completion” and “tower rearranging” but does not have a direct access to the scalar features that were initially used to train these sub-submodels (e.g. “number of puzzles attempted” and “number of puzzles correct” for the sub-subdimension “tower rearranging”). To address this shortcoming, we aimed to build models on as many scalar features as possible. Because the different scalar features were not all collected on the same participants, the sample size of the intersection was zero. To find a compromise between the breadth (the number of features included) and the depth (the sample size of their intersection) of the model, we designed a greedy hierarchical clustering algorithm.

Starting with every variable as a leaf, we first merge the two features for which the ratio between the sample size of intersection over the sample size of the union is the highest. We then recursively merge the different branches until we obtain the intersection of all the datasets as the root node. After building the tree, we assign a score to each node that is the product between the number of features included in the node and the sample size of their intersection. For each node, if the ratio between its score and the score of its parent node (closer to the root) is smaller than or equal to one, then the node is not selected. We select and mark with an asterisk "*" the nodes for which the ratio between the node's score and its parent node's score is strictly greater than one. We then rank the selected nodes by their score, and we train ten models corresponding to the top ten selected nodes.

The intuition behind this algorithm is the following. (1) The number of models we can reasonably train under time and computing resources constraints is significantly smaller than the number of distinct combinations of predictive features. We therefore need to select promising feature combinations. (2) The most promising models will be those for which both the sample size and the number of predictive features included is large. (3) The feature combinations selected should not be redundant. If the top ten selected models have highly similar predictors and participants coverage, they will likely yield similar performance. A model built on a smaller number of features with a smaller sample size still has the potential to outperform a model with a higher score, if the predictive features are better. To maximize the chance of discovering such combinations, the ten models trained should be used to explore non-redundant feature combinations. The exclusion of nodes whose score is lower than their parent's score, even if the child node's score might be high in absolute value, serves this purpose.

For the sake of clarity, let us walk through an example. Let us assume that we have three features A (sample size 100), B (sample size 120) and C (sample size 150), and we are deciding whether to first merge A and B, A and C, or B and C. Let us assume that the sample size of the intersection between A and B is 80, the sample size of the intersection between A and C is 100, the sample size of the intersection between B and C is 90, and the sample size of the intersection between A, B and C is 10. The score of the intersection between A and B is therefore 80/140=0.57; the score of the intersection between A and C is 100/150=0.67; and the score of the intersection between B and C is 90/180=0.5. We therefore merge the two leaves associated with the highest score: A and C. We now have two nodes: AC with a sample size of 100 and C with a sample size of 150. After merging the two nodes we obtain the root node, ABC, with a sample size of 20. We can now compute the score for each node as the product between the sample size and the number of features. A: 100*1=100, B: 120*1=120, C: 150*1=150, AC: 100*2=200, ABC=20*3=60. For each node, we compare its score to its parent score. The selected nodes are AC (200>60), B (120>60) and ABC (it does not have parents, so 60>0), while the rejected nodes are A (100 < 200) and C (150 < 200). We then rank the three selected nodes by their score: #1 AC (200), #2 B (120), #3 ABC (60). If our computing resources allow us to train two models, we will therefore train a first model using the features A and C as predictors and a second model using only the feature B as predictor.

During our preliminary analysis, we trained several of the highly ranked clusters. For the sake of simplicity, efficiency, and because we observed limited gains in $R^2$ values, we only trained the model with the highest score for our final pipeline.

#### Time series

##### Pulse Wave Analysis

The UKB contains 234,837 samples with pulse wave analysis measurements, a measure of arterial stiffness (field 4205). Each sample is a time series of 100 measurements, collected over one second (Supplementary Figure 45). We scaled each sample independently between 0 and 1 and used the normalizing constant as a "side predictor" for the predictive algorithms. We name side predictors scalar predictors that can be fed to a neural network along with other data modalities (time series, images, videos) to improve the predictions. Here, the normalized pulse wave analysis measurements encode the information about the shape of the time series function (Volume=f(time)), while the side predictor encodes the information about the actual height of this function (the maximum value it takes). We did not filter out any samples.

#### Electrocardiograms

The UKB contains 44,463 10 second-long 12-lead resting electrocardiograms [ECGs] (field 20205), stored as .xml files. We extracted the raw signal from the .xml files using the Beautiful soup python package[451] (Supplementary Figure 46) along with the participants' heart rate. We then used the heart rate to compute the length of every participant's heart cycle. We used this information to decompose each ECG into four components called the "level" (the mean value), the "trend" (the linear bias over time), the "seasonality" (the periodical component of the signal) and the "noise" (the allegedly random remaining variation), using the seasonal_decompose function from the statsmodel python library[452] (Supplementary Figure 47). To generate our signal, we subtracted the trend component from the raw signal. We filtered out the samples for which the heart rate was lower than 30 beats per minute or higher than 125 beats per minute, as we found by observing the ECGs that these outlying values were associated with corrupted signal (Supplementary Figure 48). After completing the preprocessing, we were left with 44,417 samples.

During our preliminary analysis, we preprocessed the data in two other ways. These preprocessing methods were outperformed by the preprocessing described above, so we did not use them for our final pipeline, but we briefly describe them here as other researchers might find them of interest. First, we use the median ECG signal for each participant. Unlike the ten-second-long signal described above, this signal is a single heartbeat for each participant, stored as 600 time steps. It can be directly extracted from the .xml files provided by the UKB. Second, we split each ten-second-long ECG signal into several single beat signals. To do this, we extracted the heart rate from the .xml for each sample, we used it to compute the length of a cardiac cycle for the participant, and we split the sample accordingly. This generated approximately ten times more samples for each participant.

#### Images

All the preprocessed images were stored as .jpg. We rescaled the values of the pixels to be between 0 and 1 instead of between 0 and 255 to mitigate the explosion of gradients in the neural networks (see Methods-Algorithms). When images were in grayscale, we saved them as RGB by duplicating the values three times. As explained in detail under Methods-Algorithms-Images, we used transfer learning to analyze the images. In the context of transfer learning, the size of the architecture of the model is affected by the dimension of the input. Large images contain more information but require larger deep learning architectures that take longer to train. To resolve prohibitory long training times, we resized the images so that the total number of pixels for each channel would be below 100,000. For example, we resized all square images larger than 316*316 pixels to this dimension. We also observed that models trained on small images (e.g. brain images) performed poorly. We found that enlarging small images yielded better results. The size of the different images before and after resizing can be found in Supplementary Table 34. The detail of the preprocessing for each of the specific image datasets can be found below.

##### Brain MRIs

The UKB contains 42,098 resting functional 3D brain MRIs videos, covering 40,597 participants (field 20227). The dimension of the files are 88*88*64 pixels, with 490 time points collected over six minutes. A “reference scan” is also available. The reference scan is a single 3D image (88*88*64 pixels) that has higher between-tissue contrast to noise and is used to correct head motion and alignment during the procedure. We selected the reference scan as well as the median time frame of the raw data (time step 245) and generated one image for each anatomical plane (sagittal, coronal, transverse). For each plane, we selected the median slice, as well as the slices that were 5 pixels before and after the median slice and saved the resulting three slices as a three-dimensional RGB image .jpeg image. We padded the images with zeros to center the brain pictures, with a resulting shape of 88*88*3 for all images. A sample of preprocessed brain MRI images can be found in Supplementary Figure 49.

##### Eye fundus

The UKB contains eye fundus RGB images, with dimensions 1536*2048*3 pixels. Data from both the left eye (field 21015, 87,562 samples for 84,760 participants were collected) and the right eye (field 21016, 88,259 samples for 85,239 participants) was collected. We took the vertical symmetry of left eye images. We cropped the images to remove the black border surrounding the actual eye fundus images, which yielded centered square images with dimensions 1388*1388*3. Some of the images were low quality on visual inspection. For example, they had very low or very high luminosity. To reduce the prevalence of such images, we developed the following two-step heuristic. First, we computed the mode of the distribution for each of the three RGB layers. If the mode is strictly larger than 250 for both the red, the green and the blue channels, and has a frequency at least as high as 100,000 for the three channels, the image is removed. Second, we compute the median of the distribution for each of the three RGB channels. If the difference between the red median and the maximum between the green and blue medians is strictly smaller than ten, the image is removed. A sample of preprocessing OCT images can be found in Supplementary Figure 50.

#### Eyes optical coherence tomography

The UKB contains OCT 3D images from both the left eye (field 21017, 87,595 samples for 84,173 participants) and the right eye (field 21018, 88,282 images for 85,262 participants) (Supplementary Figure 51). Each sample contains 128 images of dimensions 650*512*1 grayscale pixels. We selected the image where the fovea pit is the deepest as the input for our models, as follows. First, for each sample, we discarded images before the 30th image and after the 105th image, because the fovea was never the deepest in the first or last images. Then we applied the fastNlMeansDenoising function from the openCV python library[453] to each of the remaining 75 images with the following hyperparameter values: h=30, templateWindowSize=7, searchWindowSize=7. We then applied OpenCV's Canny edge algorithm[454] with 30 and 150 as the two thresholds for the hysteresis procedure to detect the surface of the retina. Instead of relying on a single threshold to detect edges, the hysteresis method consists in setting both an upper and lower threshold. Pixels whose gradient value is above the upper threshold are classified as belonging to an edge, and pixels whose gradient value is below the lower threshold are classified as non-edge. The remaining pixels, whose gradient value is between the two thresholds, are classified as an edge if they are connected to at least one edge pixel. For each column of pixels, we identified the surface of the retina as the first pixel encountered set to 1 by the canny edge algorithm, starting from the top of the image. We applied two smoothing methods (triangle moving average and Savitzky–Golay filter) to detect the surface (Supplementary Figure 52).

We then computed the curvature of the detected surface (Supplementary Figure 53), and we used the maximum curvature value for each of the 75 images. Next, we identified which of these 75 images had the maximal curvature, corresponding to the pit of the fovea. The list of maximal curvature for each image is a noisy function, so we applied the Savitzky–Golay filter to identify the image with the maximal curvature (Supplementary Figure 54). Finally, we cropped the selected image to center the fovea pit vertically to obtain the final images with dimension 500*512*3 pixels (Supplementary Figure 55). A sample of preprocessed OCT images can be found in Supplementary Figure 56.

##### Carotid ultrasounds

The UKB contains carotid artery ultrasounds from both the left (field 20222, 8,992 images for 8,965 participants) and right (field 20223, 8,995 samples for 8,968 participants) sides. For each artery, four different measures are taken. One along the short axis, one along the long axis, one at an angle of 120 degrees and one at an angle of 150 degrees. The images are screenshots of the software used to record the ultrasound images (Supplementary Figure 57), so we extracted the actual images with a resulting final dimension of 506*437 pixels. We superimposed the images taken along the long axis at different angles (long axis, 120 degree and 150 degrees) to obtain RGB images that we named "Mixed". A sample of preprocessed carotid images can be found in Supplementary Figure 58.

##### Heart MRIs

The preprocessing of heart MRI videos is described under "Methods - Data types and Preprocessing - Videos - Heart MRIs". For images, we selected the first time frame of the videos before the cropping step and converted them into RGB pictures by duplicating the grayscale layer. After preprocessing, we obtained 46,987 images of dimensions 200*200*3 out of the initial grayscale videos of dimensions 200*200*50. A sample of preprocessed heart images can be found in Supplementary Figure 59.

#### Liver and Pancreas MRIs

UKB contains Liver MRI images (field 20204, 45,685 samples for 43,267 participants) of dimensions 288*384, stored as DICOM files. We removed the 83 images for which the image quality indicator had any flag on (field 22414). We applied an adaptive histogram equalizer filter to the images to enhance the contrast. We kept both images, which we named “Raw” and “Contrast”. We cropped off the legend on the right side of the images which yielded images of dimensions 288*350, that we stored as .jpg images.

The UKB also contains pancreas images (field 20259, 37,619 samples for 35,285 participants). We followed the same pipeline used for the preprocessing of the liver images to preprocess the pancreas images and obtained 36,784 images.

A sample of preprocessed abdominal (liver and pancreas) images can be found in Supplementary Figure 60.

#### Spine, Hips, Knees and Full body DXA images

The UKB contains skeletal Dual-energy X-ray absorptiometry (DXA) images from the spine, hips, knees and full body (field 20158, 42,287 images for 41,212 participants), saved as DICOM files.

For the spine, images were collected for both the sagittal plane and the coronal plane. Not all images had the same dimensions, so we resized all of them to the median size (1513*684 pixels for sagittal, 724*720 pixels for coronal) (Supplementary Figure 62).

For the hip and knee images, we took the left symmetry for the left side images. The images had different dimensions between the participants, so we resized all the images to respectively

626*680 (hips) and 851*700 (knees). A sample for hip and knee images can respectively be found in Supplementary Figure 63 and Supplementary Figure 64.

Two full body images are made available by UKB. One displays the skeleton of the participant, surrounded by the shape of their body. We refer to this image as “Figure”. The other barely displays the bone structure but makes the other tissues more visible. We refer to this image as “Flesh”. From these two images, we generated two more images for each participant. (1) An image for which we removed the shape of the body surrounding the skeleton, which we named “Skeleton”. To isolate the skeleton from the rest of the image, we used the following algorithm. For each pixel at the border of the image, we computed the position of the first bone pixel encountered in every direction. All the pixels between the border pixel and the bone pixel were set to black. We defined the bone pixels to be pixels with a value larger than or equal to 70 (white color) and smaller than 250 (to exclude non-bone bright artefact pixels). (2) An image for which we superposed the three first images (Figure, Flesh and Skeleton) to generate a three-layer RGB image that we named “Mixed”. We resized these four images to 811*272 pixels, the median image size, for all participants (Supplementary Figure 61).

#### Physical activity

103,688 UKB participants received wrist accelerometers that they wore for a week to measure their physical activity levels[455]. Some of them repeated the experience during up to four “seasonal repeats” to evaluate the variability of their physical activity pattern between seasons, so the total amount of accelerometer records amounts to 115,464. The raw data is available and consists of three leads (x, y and z axes) and a sampling frequency of 100 Hz with a dynamic range of ±8g (field 90001). A preprocessed version of the data is also available and consists of one lead that reports the average acceleration every five seconds, after filtering for noise and correcting for gravity (field 90004). For the preprocessed version, only one sample per participant is available, for a total of 103,687 samples.

#### Physical activity as time series

First, we preprocessed the data to ensure we were analyzing activity that took place during the full week. To mitigate the problems associated with excessively long time series, we started with the summarized data (one lead, 0.2 Hz, field 90004) and computed the mean acceleration every minute. We discarded the samples for which less than a full week's worth of data was available (10,080 time steps = 7 days * 24 hours * 60 minutes), and only kept the first 10080 when more data than necessary was available. This step filtered out 25,026 samples out of the initial 103,687 samples, leaving 78,661 samples. Finally, we applied a smoothing function (exponential moving average with parameter 35) to reduce the noise. We refer to this preprocessed data as "FullWeek_Acceleration" (Supplementary Figure 65).

Then, we generated a second time series ("FullWeek_TimeSeriesFeatures") over the full week by processing the raw high frequency 3D data (field 90001). We used the "biobank Accelerometer Analysis" software[456,457] to compute the probability for each sample of performing several activities (biking, sitting/standing, vehicle, mixed, sleeping and walking) every five seconds. The software generates 113 features as an intermediate step, that it uses in a later step to classify the different activities. We computed the average for these time series every 5 seconds to leverage these features as additional channels for the FullWeek_TimeSeriesFeatures model. The final dimensions of each sample was therefore 113 channels of 7 days * 24 hours * 12 5-minutes long periods = 5,373 time steps.

We then filtered low quality samples. First, we removed 6,618 samples for which data was collected for less than a week. The summary file outputted by the software that we used to preprocess the data also contains quality indicators. We used four columns (quality-calibratedOnOwnData, quality-goodCalibration, quality-goodWearTime and quality-daylightSavingsCrossover) to filter out 8,984 low quality samples. After preprocessing the dataset, we were left with 99,862 samples.

We generated a third time series from the raw high frequency 3D data to specifically compare the walking pattern between the different participants, which we refer to as "Walking_3D". We identified for each sample the continuous periods during which the activity was predicted by the software to be walking with a probability of 1. For each sample, we then randomly selected up to five of these sequences and extracted the corresponding 3D raw acceleration data (Supplementary Figure 66).

Finally, we generated a similar dataset to predict age as a function of physical activity during sleeping. The predictor was weak so it is not included in the main tables, but we describe our methods for researchers who would be interested in reusing this pipeline for different purposes. First we detected continuous periods of 180 and 360 minutes of sleep. Because the probability is the sleeping activity is never equal to 1 over these long periods of time, we first generated binary time series that specified every five seconds whether the activity was sleeping with a probability of 1, or not. Then we smoothed this curve by using a sliding window with a period of 30 minutes and set a threshold of 0.3 to determine the beginning and the end of the sleeping period. This allowed us to include brief periods of physical activity during the night as being part of the sleeping period. We then extracted the 3D high intensity acceleration data corresponding to the sleeping periods of 180 and 360 minutes. We extracted as many of these samples as possible for each participant. Neither the 180 minutes nor the 360 minutes hyperparameters yielded good predictors of chronological age (R-Squared=0.016 for both models).

#### Physical activity as scalar features

We filtered out the low-quality samples based on the quality-calibratedOnOwnData, quality-goodCalibration, quality-goodWearTime and quality-daylightSavingsCrossover indicators, as mentioned in “Physical activity as time series” above. Then we generated a total of 2,271 scalar features, from three different pipelines (Supplementary Table 35).

First, we leveraged 1,333 summary features directly outputted by the two biobank Accelerometer Analysis preprocessing softwares[456,457]. The first software[457] generated 813 features from which we filtered out 66 (Supplementary Table 36) non-biological features (e.g. features related to the calibration of the accelerometers) and kept the remaining 747 features. The second software[456] generated 896 features, from which we filtered out 67 non-biological features. We then took the union of the features generated by the two models to obtain the 1,333 features.

Second, we used the 113 time series generated by the Accelerometer Analysis software during one of its intermediate steps. For each of these time series, we computed the minimum value, the maximum value, the average value, the standard deviation, as well as the 25th, 50th and 75th percentiles, for a total of 791 features.

Finally, we built 147 custom features from six different types. (1) Summary statistics features, (2) Behavioral features, (3) Sleeping features, (4) Activity phase features, (5) Weekend features, (6) Metabolic Equivalent of Task (MET) features. The design of these features is described in detail below.

##### Summary statistics features

First, we computed the average acceleration every minute for each participant, and we computed the mean, median, interquartile range, skewness, and the fifth, tenth, 25th, 75th and 90th percentile on these time series. We then computed the average acceleration over every day of the week, and we computed the following summary statistics: the mean, standard deviation, minimum, maximum, median, and the 25th and 75th percentiles. Finally, we computed the same summary statistics on the different periods of the day: (1) night (midnight to 6am), (2) morning (6am to 9am), (3) day (9am to 7pm) and (4) evening (7pm to midnight).

###### Activity specific features

We computed the number of minutes spent on each activity (e.g. walking, sleeping). We then computed the distribution of time across the different activity types for each day of the week along with the standard deviation for these values. Finally, we computed the same features described under “Summary statistics features” for every physical activity. For each activity, we defined the time series on which to perform this analysis as the concatenation of periods of at least five minutes over which the activity classification remained constant.

###### Sleeping features

We defined night sleep as time steps classified as sleep between 10pm and 6am, and we computed the average night sleep duration. We defined deep sleep as an uninterrupted series of timesteps classified as sleeping, and we computed the average duration of deep sleep phases for each participant. Two participants could have the same average night sleep duration of eight hours, but the first participant could have a single deep sleep phase of eight hours while the second participant could briefly wake up or move in their sleep three times throughout the night, which would read as an average deep sleep duration of only two hours. The average deep sleep duration can therefore be used to assess the quality of sleep that each participant is getting. Then, we defined a sleep session as a sleeping activity longer than twenty minutes and we computed the daily average number of sleep sessions for each participant. This feature for example captures if a participant is taking naps. Finally, we selected the longest sleep session for each participant, which represents their “night sleep”, even if some participants might be working night shifts and therefore have this “night sleep” during the day. We computed the average duration of this night sleep and the average time of the day at which it started for each participant.

##### Physical activity features

We computed the daily average time spent walking, as well as the daily average duration of the longest continuous walk. We also computed the average time of the day associated with the first and the last peak of physical activity.

##### Weekend features

We defined the weekend as the time between Friday 10pm and Sunday 11:59pm and we computed the time spent on the different activities during this period. We also computed the time at which the participants went to bed on the night of Saturday evening to Sunday morning to capture Saturday night habits.

##### Metabolic Equivalent of Task (MET) features

The metabolic equivalent of a task is the rate at which a participant is expending energy when performing this task compared to when they are resting (e.g. sitting or being inactive). For example, a MET of two means that the participant is spending twice as much energy performing their current activity than they are when they are resting. Activities with a MET smaller than three (e.g. walking slowly or working at a desk) are considered to be light intensity activities. Activities with a MET between three and six qualify as moderate intensity activities (e.g. walking at a faster pace or practicing relaxing sportive activities). Activities with a MET larger than six are considered vigorous intensity activities (e.g. jogging, biking, dancing, weightlifting, competitive sportive activity).[458–462]

MET measures can be used to estimate the total energy expended over the day or the week, using the unit of MET hour. For example, if a participant performs a task with a MET of three for 40 minutes, that is equivalent to performing a task with a MET of one for two hours. In both cases, the total energy expended by the participant while practicing the activity is two MET hours.

The UKB software estimates the MET for every time step when it preprocesses the data. We leveraged this column to compute the following features. First, we computed the total MET hours over the full week. Then, we computed the daily (seven values) and the hourly (24 values) MET averages and associated standard deviations. Finally, we computed the mean and the standard deviation of the MET values for low, medium and vigorous activities over the week. We defined low, medium and vigorous intensity using the aforementioned threshold values.

##### Physical activity as images

Starting from the unidimensional, preprocessed acceleration average with a frequency of 0.2 Hz (field 90004), we computed the average acceleration for every minute. From there, we generated four "images" per sample for each participant. We describe how these "images" are created below.

For the first, we generated a custom recurrence plot[463]. We used a sliding window of length ten minutes and stride of one minute, so the value of the pixel in row i and column j is the Euclidean distance between the vector of the 10 one-minute-average accelerations starting at the minute i and the vector of the 10 one-minute-average accelerations starting at the minute j and stored them as floats. We generated two recurrence plots. One using only the time steps for which the probability of the participant sleeping was 1, and one for the remaining time steps. Because recurrence plots are symmetrical, we merged these two recurrence plots into a single image, in which the upper right triangle corresponds to the sleeping timesteps, and the lower left triangle corresponds to the remaining time steps (Supplementary Figure 67).

Second, we generated a Markov transition fields image (n_bins=8), a Gramian angular difference fields image and a Gramian angular summation field image[464] respectively using the MarkovTransitionField and the GramianAngularField functions of the pyts python package[465]. Finally, we resized all the images to be of size 316*316. A sample of these three images can be found in Supplementary Figure 67.

We generated other images that we ended up not using for the analysis. We will describe them here as knowing that they were poor age predictors might save time for researchers considering similar approaches. First, we generated the recurrence plots using the average acceleration over periods of 32 minutes instead of one minute, which generated images of dimension 316*316 directly, saving the resizing step. We did not observe significant differences between the two methods when we explored this possibility, so we discarded it. We also applied the binary filter traditionally used for recurrence plots, by setting to one the value for the pairs of points that were in the 95th percentile for proximity. We found that the performances were similar, with a possible advantage for using float values. Finally, we also applied these four methods (recurrence plot, Markov transition field and Gramian angular summation and Gramian angular difference) to selected segments of 3D data for the walking and the sleeping activities, possibly capturing the recurrence patterns in the gait, for example. During our preliminary analysis and early exploration of the hyperparameters, we found that none of these 3D data-based plots yielded R-Squared values higher than 0.05.

#### Videos

##### Heart MRIs

The UKB contains long-axis heart MRI videos (field 20,208, 46,869 samples for 44,413 participants), stored as 50 time 200*200 DICOM images. The 50 images do not correspond to a specific sampling frequency. Pictures are instead collected over several cardiac cycles (where a cycle is one heartbeat). They are selected and rearranged until the full cardiac cycle of the participant can be reconstituted. As a consequence, two participants with two different heart rates will still have their full cardiac cycle summarized by 50 images. We filtered out samples for which the number of time frames was different than 50.

Three different views of the heart were recorded by UKB: (1) two-chamber view (displaying the right atrium and ventricle), (2) three-chamber views (displaying the two atria and the left ventricle) and (3) four-chamber view (displaying the two atria and ventricles). We filtered out 844 samples for which one or more of the three views was not available. We applied a histogram equalizer filter to enhance the contrast of the images (similarly to what we did for the liver and pancreas images). We stored both the raw and the contrasted images, which we respectively refer to as "Raw" and "Contrast".

For the images, we saved the first frame of each of the remaining samples or the remaining samples without cropping the images, as mentioned under "Preprocessing - Images - Heart". For the videos, we cropped the images to obtain 150*150 images centered on the heart, with the aim to reduce the size of the model needed to train on the videos. For this same reason, we generated two 25 time frames-long samples from each 50 time frames video: one with the even numbered time frames, and one with the odd numbered time frames. This last step allowed us to halve the size of each image, therefore doubling the batch size that would fit into the model. Because our preliminary results on images showed a lesser predictive potential for the two-chamber view, we only trained 3D CNN models on three-chamber view and four chamber view videos.

#### Data augmentation

##### Images

To prevent overfitting and increase our sample size during the training we used data augmentation[466] on the images. Each image was randomly shifted vertically and horizontally, as well as rotated and zoomed. We chose the hyperparameters for these transformations' distributions to represent the variations we observed between the images in the initial dataset. For example, for the eyes OCT images, we observed a stronger variation between images in the vertical direction than in the horizontal direction, so the random vertical shift was sampled from the [-20%; +20%] uniform distribution, whereas the random horizontal shift was sampled from the [-10%; +10%] uniform distribution. A summary of the hyperparameter values for the transformations' distributions can be found in Supplementary Table 37.

The data augmentation process is dynamically performed during the training. Augmented images are not generated in advance. Instead, each image is randomly augmented before being fed to the neural network for each epoch during the training.

##### Videos

For the heart MRI videos, we introduced a random rotation of plus or minus three degrees in the spatial plane, sampled from the uniform distribution [-3,3]. Similarly to images, the augmentation process for videos is performed dynamically during the training. Each video is randomly augmented before being fed to the neural network.

### Machine learning algorithms

#### Scalar data

We used three different algorithms to predict age from scalar data (non-dimensional variables, such as laboratory values). Elastic Nets [EN] (a regularized linear regression that represents a compromise between ridge regularization and LASSO regularization), Gradient Boosted Machines [GBM] (LightGBM implementation[467]), and Neural Networks [NN]. The choice of these three algorithms represents a compromise between interpretability and performance. Linear regressions and their regularized forms (LASSO[34], ridge[33], elastic net[35]) are highly interpretable using the regression coefficients but are poorly suited to leverage non-linear relationships or interactions between the features and therefore tend to underperform compared to the other algorithms. In contrast, neural networks[39,40] are complex models, which are designed to capture non-linear relationships and interactions between the variables. However, tools to interpret them are limited[468] so they are closer to a “black box”. Tree-based methods such as random forests[36], gradient boosted machines[37] or XGBoost[38] represent a compromise between linear regressions and neural networks in terms of interpretability. They tend to perform similarly to neural networks when limited data is available, and the feature importances can still be used to identify which predictors played an important role in generating the predictions. However, unlike linear regression, feature importances are always non-negative values, so one cannot interpret whether a predictor is associated with older or younger age. We also performed preliminary analyses with other tree-based algorithms, such as random forests[36], vanilla gradient boosted machines[37] and XGBoost[38]. We found that they performed similarly to LightGBM, so we only used this last algorithm as a representative for tree-based algorithms in our final calculations.

#### Time series

We analyzed three different time series datasets for this paper: pulse wave analysis [PWA] data (a measure of arterial stiffness), ECGs and wrist accelerometer recordings. We used one architecture for PWA, one architecture for ECGs and three architectures for accelerometer recordings. The architectures are significantly different but share the following structure when they are not directly adapted from a publication. Our original architectures are composed of two blocks, (1) the convolutional block and (2) the dense block. The convolutional block consists of one-dimensional convolutional layers that extracts information from the temporal dimension of the signal. These convolutional layers use batch normalization, zero padding, kernel and bias regularization, ReLU as their activation function and are followed by max pooling to progressively halve the temporal dimension. A global max pooling is applied after the last convolutional layer to convert the temporal features into scalar features. We then concatenate the output of the convolutional block with scalar features computed by a side dense neural network built on scaling factors (the amplitudes of the time series signals before we normalized them). The concatenated scalar features are then inputted into the dense block. The dense block consists of five fully connected layers. The number of nodes in the first layer is the largest power of two that is smaller than the input size, and the number of nodes in the second layer is half this value. For example, if the input of the first dense layer is of size 1,040, the first and second dense layer will have 1,024 and 512 nodes, respectively. We then concatenate the output of the third layer with the features generated by a side neural network trained on demographic variables (sex and ethnicity) and feed the concatenation to the third layer. The number of nodes in the third layer is the largest power of two that is smaller than the input size, and the number of nodes in the fourth layer is half this value. We capped the number of nodes for these first four fully connected layers at 1,024. They all use ReLU activation, batch normalization, kernel and bias regularization, and dropout. We then added a fifth and final dense layer with a unique node with a linear activation function to predict chronological age. We plotted a figure for each architecture using the keras python library[469].

We used Adam[470] as the compiler for all the time series models.

##### Pulse wave analysis

To analyse the pulse wave analysis data we built an eight layers deep convolutional neural network [CNN] whose architecture can be found in Supplementary Figure 68 and Supplementary Figure 69. The architecture consists of a convolutional block and a dense block. The inputs for the convolutional block are time series with a single channel and 100 time steps. The convolutional block is composed of three convolutional layers with kernels of size three and strides of one. We used zero-padding so that the convolution would not reduce the spatial dimension of the input. The number of filters for these three convolutional layers are respectively 32, 64 and 128. We used one dimensional max pooling with a kernel size of 2 after each of the first two layers to reduce the dimension of the input, and we used a global max one-dimensional layer after the last convolutional layer to convert each of the 128 filters to a scalar feature. We then concatenated these 128 scalar features with the scaling factor of the imputed sample, duplicated 12 times (the scaling factor is a measure of the amplitude of the raw signal before it was normalized between zero and one, as described under Methods - Data types and Preprocessing - Time series - Pulse Wave Analysis). We inputted these 140 features into the dense block, which consists of two dense layers with respectively 128 and 64 nodes. We concatenated the 64 features outputted by second dense layer with sex and ethnicity. We inputted these 89 features into two dense layers with respectively 64 and 32 nodes. We used batch normalization, kernel regularization, bias regularization, ReLU activation for the seven first layers of the architecture. We used dropout to regularize the four dense layers, as well. Finally, we used a dense layer with a single node and linear activation as the final eighth layer to predict chronological age. We refer to the five dense layers as the dense block.

We built simple LSTM and GRU architectures, as well. These architectures underperformed during our preliminary analysis ($R^2$=14%) and we therefore did not train these models for our final pipeline.

#### Electrocardiograms

We used the architecture from the article "Prediction of mortality from 12-lead electrocardiogram voltage data using a deep neural network"[147]. The original architecture can be found under "Extended Data Fig. 2" in the original paper.

The architecture of our model can be found in Supplementary Figure 70 and Supplementary Figure 71. Our architecture is composed of two blocks: (1) the convolutional block and (2) the dense block. The convolutional block comprises five seven-layer deep one-dimensional convolutional neural networks branches, in parallel. Each of these CNN branches takes three ECG leads of 5,040 time steps (ten seconds, we padded 20 time steps before and after the raw signal to mirror the original model[147]) as its input, with some of the leads being used as input for more than one CNN. We used zero-padding for the convolutional layers. Each CNN's branch's output is processed by a max pooling to convert the 512 one-dimensional filters into 512 scalar features, which are then concatenated with 64 features built on sex and ethnicity. These 2,624 features are then inputted into the dense block, which is a six-layer-deep fully connected neural network. The number of nodes in the first four layers is halved every layer (256, 128, 64, 32). The fifth layer has eight nodes, and the sixth layer has a single node with a linear activation function to predict age. We applied batch normalization, kernel and bias regularization, and we used ReLU as activation function for every layer in the architecture, except for the final layer.

In other words, we made the following changes to adapt the original architecture to the task of predicting chronological age instead of one year survival. (1) We replaced chronological age by ethnicity as a side predictor. (2) We replaced the final sigmoid activation layer by a linear activation layer. (3) We used the same architecture for the five CNN branches, unlike the original architecture which is using different CNN architectures for the different CNN branches. (4) We used zero-padding on the convolutional layers which conserved their input's dimension. We relied on max pooling layers with a size of two and a stride of two to iteratively halve the input's size. (5) We added batch normalization and ReLU as activation functions to the dense layers, except for the final dense layer (see (2)).

During our preliminary analysis, we explored simpler architectures using one-dimensional convolutional layers, LSTM layers or GRU layers. We found that our architectures did not perform as well (custom one-dimensional CNNs: $R^2$=30%; LSTMs and GRUs: $R^2$=20%) as the architecture described above so we did not train these models for our final pipeline. In particular, we found that LSTM and GRU models significantly underperformed, with $R^2$ values lower than 10%.

##### Physical activity

We developed a different architecture for the three time series extracted from the wrist accelerometers data (see Methods - Data types and Preprocessing - Physical activity - Physical activity as time series).

###### Architecture for raw acceleration data over the full week

For the model based on the acceleration data across the full week (FullWeek acceleration), we used the architecture from the paper "Extracting biological age from biomedical data via deep learning: too much of a good thing?"[141]. The architecture of the model is described in Supplementary Figure 72 and Supplementary Figure 73 and is constituted of two blocks: (1) a convolutional block and (2) a dense block. These two blocks form a seven layers deep neural network, preceded by a batch normalization. The inputs for the convolutional block are time series with a single channel and 10,080 time steps. The convolutional block consists of four one-dimensional convolutional layers with respectively 64, 32, 32 and 32 filters, a kernel size of respectively 128, 32, eight and eight, and a stride of one. Every convolutional layer is zero-padded and followed by a max pooling with a size of respectively four, four, three and three, and with a stride of two. The output of the convolutional block is inputted into the dense block which consists of three dense layers. The first two dense layers have respectively 256 and 128 nodes and are using dropout. The first six layers of the architecture use kernel and bias regularization, and use ReLU as their activation function. The seventh layer is a dense layer with size one and with a linear activation function to predict chronological age.

During our preliminary analysis, we tested a second architecture, from the article “Deep Learning using Convolutional LSTM estimates Biological Age from Physical Activity”[140]. This architecture uses both convolutional layers and LSTM layers and takes as input the data formatted as a 3-dimensional data frame of dimensions 7x24x60. The first dimension is for the days of the week, the second for the hours of the day and the third for the minutes of the hour. The model’s architecture can be found Figure 2 of the aforementioned paper. This architecture explained 33% of the variance in chronological age but was outperformed by the first model described above and took significantly longer to train. As a result, we did not select this second architecture for our final pipeline.

##### Architecture for features extracted from the acceleration data over the full week

To analyze the 113 time series of features extracted from the raw acceleration data, we built a 14 layers deep architecture which can be found in Supplementary Figure 74 and Supplementary Figure 75. The architecture consists of a convolutional block and a dense block. The inputs for the convolutional block are time series with 113 channels and 2,016 time steps. The convolutional block comprises nine one-dimensional convolutional layers with the number of filters for each layer doubling with every layer, starting from 64 and with a maximum number of filters of 4,096 (64, 128, 256, 512, 1,024, 2,048, 4,096, 4,096, 4,096). Each convolutional layer is zero-padded and followed by a one-dimensional max pooling layer of size two and stride 2. The output of the convolutional block is converted to 4,096 scalar features using a global max pooling over each of the 4,096 filters. A single layer neural network (409 nodes) is parallelly taking as input the 113 scaling factors for the 113 normalized time series that were inputted in the convolutional block. The 4,096 convolutional based features are concatenated with the 409 scaling factor-based features, and the resulting 4,505 scalar features are inputted in the dense block. The dense block is composed of five dense layers with respectively 1,024, 1,024, 1,024, 512 and one node(s). All dense layers, except for the last one, use dropout. The last layer uses linear activation to predict chronological age. A single layer (25 nodes) neural network is trained on sex and ethnicity, and its output is concatenated with the output of the second dense layer before being inputted in the third dense layer. All the layers of the architecture aside from the final layer use batch normalization, kernel and bias regularization, and ReLU for their activation function.

##### Architecture for three-dimensional high frequency walking data

We built a 13 layers deep convolutional architecture to analyze the walking data. This architecture can be found in Supplementary Figure 76 and Supplementary Figure 77. The architecture is highly similar to the one described above. It comprises a convolutional block and a dense block.

The inputs for the convolutional block are time series with three channels and 5,700 time steps (5.7 seconds of 3-dimensional acceleration). The convolutional block consists of eight one-dimensional convolutional layers with the number of filters for each layer doubling with every layer, starting from 32 and with a maximum number of filters of 2,048 (32, 64, 128, 256, 512, 1,024, 2,048, 2,048). All convolutional kernels are zero-padded, of size three and with a stride of one. Each convolutional layer uses batch normalization, ReLU as activation function, and is followed by a one-dimensional max pooling layer of size two and stride 2. The output of the convolutional block is converted to 2,048 scalar features using a global max pooling over each of the 4,096 filters. A single layer neural network (204 nodes) is parallelly taking as input the 3 scaling factors for the x-axis, y-axis and z-axis normalized time series that were inputted in the convolutional block (the three dimensions of the accelerometer). The 4,096 convolutional based features are concatenated with the 204 scaling factor-based features, and the resulting 2,252 scalar features are inputted in the dense block. The dense block is composed of five dense layers with respectively 1,024, 1,024, 1,024, 512 and 1 nodes. The first four dense layers use dropout. The last layer uses linear activation to predict chronological age. A single layer (25 nodes) neural network is trained on sex and ethnicity, and its output is concatenated with the output of the second dense layer before being inputted in the third dense layer. All layers, except for the last one, use batch regularization, kernel and bias regularization, and use ReLU as their activation function.

#### Images

##### Convolutional Neural Networks Architectures

We used transfer learning[471–473] to leverage two different convolutional neural networks[76] [CNN] architectures pre-trained on the ImageNet dataset[474–476] and made available through the python Keras library[469]: InceptionV3[477] and InceptionResNetV2[478]. We considered other architectures such as VGG16[258], VGG19[258] and EfficientNetB7[479], but found that they performed poorly and inconsistently on our datasets during our preliminary analysis and we therefore did not train them in the final pipeline. For each architecture, we removed the top layers initially used to predict the 1,000 different ImageNet images categories. We refer to this truncated model as the “base CNN architecture”.

We added to the base CNN architecture what we refer to as a “side neural network”. A side neural network is a single fully connected layer of 16 nodes, taking the sex and the ethnicity variables of the participant as input. The output of this small side neural network was concatenated to the output of the base CNN architecture described above. This architecture allowed the model to consider the features extracted by the base CNN architecture in the context of the sex and ethnicity variables. For example, the presence of the same anatomical heart feature can be interpreted by the algorithm differently for a male and for a female. We added several sequential fully connected dense layers after the concatenation of the outputs of the CNN architecture and the side neural architecture. The number and size of these layers were set as hyperparameters. We used ReLU[480] as the activation function for the dense layers we added, and we regularized them with a combination of weight decay[481,482] and dropout[483], both of which were also set as hyperparameters. Finally, we added a dense layer with a single node and linear activation to predict age.

##### Compiler

The compiler uses gradient descent[484,485] to train the model. We treated the gradient descent optimizer, the initial learning rate and the batch size as hyperparameters. We used mean squared error [MSE] as the loss function, root mean squared error [RMSE] as the metric and we clipped the norm of the gradient so that it could not be higher than 1.0[486].

We defined an epoch to be 32,768 images. This corresponded to the length of one epoch being the same across datasets, independently of the difference in sample sizes. If the training loss did not decrease for seven consecutive epochs, the learning rate was divided by two. This is theoretically redundant with the features of optimizers such as Adam, but we found that enforcing this manual decrease of the learning rate was sometimes beneficial. During training, after each image has been seen once by the model, the order of the images is shuffled. For images that were available for both sides of the body, such as eye, carotids, hip or knee, both images would be shown to the model for each sample, and the prediction would be computed as the average prediction between the two images. If the image for one side was missing, the image from the other side was used twice. At the end of each epoch, if the validation performance improved, the model's weights were saved.

We defined convergence as the absence of improvement on the validation loss for 15 consecutive epochs. This strategy is called early stopping[487] and is a form of regularization. We requested the GPUs on the supercomputer for ten hours. If a model did not converge within this time and improved its performance at least once during the ten hours period, another GPU was later requested to reiterate the training, starting from the model's last best weights.

#### Videos

Our architecture is largely inspired by the C3D[488] architecture. We treated time as a third spatial dimension and built a three-dimensional convolutional neural network [3D CNN] to analyze it. The architecture of the model can be found in Supplementary Table 39 and is composed of a convolutional block and a dense block. The convolutional block consists of four three-dimensional convolutional layers. The first layer has 16 cubic kernels of dimensions 3x3x3, the second has 64 cubic kernels of dimensions 3x3x3, the third has 512 cubic kernels of dimensions 5x5x5, and the fourth layer has 1,024 kernels of dimensions 5x5x7, the third dimension being time. All convolutional layers have a stride of 1x1x1 and are zero padded to conserve the dimension of their input. We progressively reduce the spatial dimensions of the input sample by using a max pooling operation with dimension 2x2x1 and stride 2x2x1 after each convolutional layer. The fifth layer is a max-pooling layer in the spatial dimensions. The features are then flattened and batch normalized[489] before being fed to the dense block. The dense block consists of two dense layers. The penultimate layer is a fully connected layer with 1,024 nodes with dropout. The final layer is a fully connected layer with a single node with a linear activation to predict chronological age. All other layers use SELU[490] as their activation function.

We used the same compiler for the videos as for the images (see Methods - Machine learning algorithms - Images - Compiler), with the exception that an epoch is defined as seeing each sample once, and that the learning rate is decreased by 20% if the training performance does not improve for five consecutive epochs.

We also built a CNN-LSTM architecture on the heart MRI videos. The CNN extracts spatial features from each frame of the video. These spatial features are then fed as a time series as input to the LSTM that leverages the temporal dimension. We selected the 2D CNN architecture that performed best on heart MRI images and we used it to extract 64 features from each image. This step gave 64 50 time frames-long time series for each sample video. We then built an LSTM on this dataset to predict chronological age. We found that the CNN-LSTM architecture underperformed compared to the 3D CNN architecture ($R^2$=69.8±0.9%), so we did not include it in our final pipeline.

#### Underperforming models

As mentioned above, we explored alternative architectures and data preprocessing methods for some of our models, during our preliminary analysis. We did not select some of the underperforming models for our final pipeline, and the associated results are not found on the website. Here, we briefly summarize why these models were not selected.

We predicted chronological age using impedance-based and dual-energy X-ray absorptiometry [DXA]-based fat percentage estimations. The neural network predicted chronological age with a $R^2$ higher than 99%. After investigation, we discovered that the fat percentage estimations take chronological age into account. The neural network was able to extract this information to give almost perfect predictions. Interestingly, the elastic net and the gradient boosted machine were unable to efficiently recover the chronological age variable and gave $R^2$ values lower than 70%.

As reported in Figure 7, we predicted chronological age from spirometry scalar features with a $R^2$ value of 25.6±0.1%. We also built a one-dimensional neural network to leverage the full spirometry data as time series. We found no significant difference in prediction accuracy of the two models, so we did not include the model based on the time series in the final pipeline.

As described more in detail in the methods, we tested different architectures to predict chronological age as a function of the ECGs data as time series. We only included the best performing one in our final pipeline.

As described in the method and reported on the website, we predicted chronological age using three-dimensional wrist accelerometer data with a $R^2$ value of 32.3±0.01%. We hypothesized that the regular gait pattern could be leveraged by recurrence plots, Markov transition fields plots and Gramian angular fields plots. We found that training convolutional neural networks on these images gave $R^2$ values lower than five percents so we did not include them in the final pipeline.

In the same way that we predicted chronological age using high frequency wrist accelerometer data collected as the individuals were walking, we built one dimensional and two-dimensional CNNs on data collected during the participants' sleep. We found that these models gave $R^2$ values lower than three percent, so we did not include them in the results. We hypothesize that this can be partly explained by the fact that, unlike walking which is an activity with a clearly identifiable and comparable pattern, sleeping is irregular. A participant can stay immobile for several hours and only move for a few seconds.

We tested three two dimensional CNN architectures for which the results cannot be found on the website to predict chronological age as a function of images: VGG16, VGG19 and EfficientNetB7. During our preliminary analysis, we found that these architectures significantly underperformed. For example, we found that the VGG architectures trained on heart or liver MRIs predicted age with a validation $R^2$ value between 30% and -4%, whereas InceptionResNetV2 and InceptionV3 both gave $R^2$ values around 70%. We found that this poor performance was often driven by $R^2$ values close to zero on a subset of the ten cross validation folds. For example, a VGG19 model trained on heart MRI videos that predicted chronological age with a validation $R^2$ value lower than 10% over the ten folds gave a $R^2$ value of 60% on one of the ten validation folds. This suggests that VGG19 could have given prediction performances similar to the other models, but that successfully tuning this architecture would have required more time. It is possible that the architectures we excluded have similar optimal performances but are more sensitive to weights' initialization, gradient vanishing, gradient exploding, or nodes dying.

Aside from the three-dimensional CNN [3DCNN] architecture described under the methods, we predicted chronological age from heart MRI videos using an CNN-LSTM architecture. We found that its performance did not match the 3DCNN's, so we did not include it in our final pipeline.

### Training, tuning and predictions

#### Data splitting

We split the 676,787 samples into ten data folds, while keeping all samples from the same participant in the same fold. To ensure this, we split the 502,211 participants' ids (referred to by UKB as "eid") into ten different buckets of the same size. To generate 10 folds for each sub-dataset (e.g. ECGs), we took the intersection of the samples in each of the 10 folds with the samples for which the sub-dataset data was available. This method had however one important loophole, which is that we could not guarantee that the folds for the sub-datasets would be balanced. For example, resting ECG data was only recorded for 42,360 out of the 502,211 participants. Since the 502,211 participants are split into ten folds, a fold contains approximately 50,221 participants. Although unlikely, we could therefore not guarantee that all or most of the ECG samples would be attributed to the first data fold, leading to highly unbalanced folds for the ECGs analysis. Unbalanced folds can lead to problems during the cross-validation (see further below), as models trained on a smaller number of samples will tend to generalize worse. One solution would have been to use a different split for each dataset, but this would have generated problems when building the ensemble models fold by fold (see Methods - Models ensembling). To mitigate this issue of unbalanced data folds, we developed the following heuristic. We randomly split the 502,211 participants into ten folds, 1,000 times. For each of these 1,000 splits, we computed for each sub-dataset the variance of the percentages of samples in each fold. We then scored each of the 1,000 splits using the maximum of the variance among the different sub-datasets. For example, if the ECG samples were not evenly split for the ith split out of the 1,000 splits (e.g. fold 1: 55% of the samples, every other fold: 5% of the samples), the variance of the sample proportions would be high, which would yield a poor score for the ith split. Finally, we selected the split with the lowest score as the final split for the main dataset, and for all the sub-datasets. This selected split had a score of 5.8e-4, which means that the most unbalanced sub-dataset had a variance in its sample size proportion between its ten folds of 5.8e-4.

#### Scalar data

##### Nested cross-validation

Cross-validation is a method to tune the regularization of models and prevent overfitting[491]. For the models inputting scalar data (see Table 1 in green for examples), we tuned the hyperparameters and generated a testing prediction for each sample using a nested 10x9-folds cross-validation. We refer to the two nested cross-validations as the “outer” and the “inner” cross-validations. The outer-cross validation is used to generate an unbiased testing prediction for each sample, as opposed to a simple split of the data into a “training+validation” set on one hand, and a testing set on the other hand, which would only generate a testing prediction for one tenth of the dataset. The inner cross-validation is used to tune the hyperparameters more precisely, leveraging the full inner cross-validation dataset as a validation set, as opposed to a simple data split of the “training+validation” dataset into a training and a validation sets, which would only use one data fold as the validation set to estimate the performance associated with a specific combination of hyperparameters. The nested cross-validation is illustrated in Supplementary Table 40.

##### Bayesian hyperparameters optimization

To tune the hyperparameters, we used the Tree-structured Parzen Estimator Approach[492] [TPE] of the hyperopt python package[493]. TPE is a sequential Bayesian hyperparameters optimization method that iteratively suggests the next most promising hyperparameters combination as a function of the hyperparameters combinations that have already been tested, by building a probabilistic representation of the objective function. We set the number of iterations to 30. For each model, 30 different hyperparameter combinations are iteratively tested before selecting the best performing one. The hyperparameters names and their ranges defining the hyperparameters space can be found in Supplementary Table 38. It might be of interest to other researchers that we initially tuned the hyperparameters using a random search[494] with the same number of iterations, and we did not observe a significant improvement in the model's performance after implementing the Bayesian hyperparameters optimization.

##### Example

For the sake of clarity, let us walk through a concrete example, which is illustrated in Supplementary Table 40. Suppose we want to generate unbiased predictions for every sample in a dataset using an elastic net. First, let us generate the testing prediction for the data fold F9, which is performed by the first fold of the outer cross-validation (outer cross-validation fold 0). We select the data fold F9 out of the ten data folds as the testing fold, and we select the remaining nine data folds as "training+validation" folds for the inner cross-validation. We scale and center the target (age) and the predictors using the mean and standard deviation values of the variables on the "training+validation" dataset. We then enter the first inner-cross validation.

For the first inner cross-validation fold, we select the data fold F8 as the validation set, and the remaining eight "training+validation" data folds as the training set. We re-scale and center age and the predictors in the training and the validation sets using the mean and standard deviation values of the training set. We train the model on the eight training data folds with the first hyperparameters combination sampled by the TPE algorithm (one value for alpha and one value for l1_ratio) and generate validation predictions on the validation fold (data fold F8), which we unscale. This completes the first of the nine inner cross-validation folds (Inner CV fold 0). We then permute the nine inner data folds. We scale the age and the predictors using the mean and standard deviation computed on the new training set. Then we train the model with the same first combination of hyperparameters on eight data folds, leaving aside the data fold F9 (still being used as the testing set for the outer cross-validation) and the data fold F7 (now being used as the validation set for the inner cross-validation). We then use the new trained model to generate validation predictions on the data fold F7, which we unscale. This completes the second of the nine inner-cross validation folds (Inner CV fold 1). We then reiterate these inner permutation and training processes seven more times, until every data fold in the nine “training+validation” data folds is used as the validation set once. At this point, we concatenate the validation predictions from these nine validation folds to obtain the overall validation predictions associated with the first hyperparameters combination, and compute the associated performance metric (e.g. RMSE). This completes the inner-cross validation for the first hyperparameters combination.

We then perform the same 9-folds inner cross-validation, this time with the second hyperparameters combination suggested by the TPE algorithm. We iterate this process 28 more times, until 30 different hyperparameters combinations have iteratively been tested. Next, we select the hyperparameter combination that yielded the best validation performance (e.g. minimum RMSE), and we retrain a model on the whole nine “training+validation” data folds (all data folds except for data fold #1), using this best performing hyperparameters combination. This completes the first inner cross-validation.

We then use the model to generate unbiased predictions on the unseen testing set (data fold F9) and record these predictions. By anticipation for the ensembling algorithm (see Methods - Models ensembling) we also need to compute validation predictions on the data fold F8. We do this by training a model on all the data folds aside from the validation fold (data fold F8) and the testing fold (data fold F9), with the selected hyperparameters combination. We then use this trained model to compute predictions on the validation fold (data fold F8) and record these predictions, after unscaling them. This completes the first of the ten outer cross-validation folds (outer cross-validation 0).

We then complete the second outer cross-validation fold (outer cross-validation 1), this time using the data fold F8 as the testing dataset, to obtain unbiased testing predictions on this data fold, as well as validation predictions on the data fold F7. We reiterate the process eight more times to obtain the testing and validation predictions on the remaining data folds. We then concatenate the testing predictions from the ten data folds to obtain our final testing predictions for the model. Similarly, we concatenate the validation predictions from the ten data folds to obtain our final testing predictions for the model, which will later be used during ensemble models building and model selection (see Methods - Models ensembling).

The final validation and testing predictions for each data fold are therefore not necessarily associated with the same hyperparameters combination. It is also important to notice that we performed a single outer cross-validation, but that we performed a separate inner-cross validation for each outer cross-validation fold (hence the word "nested"), for a total of ten inner cross-validations per outer cross-validation fold.

#### Images

##### Hyperparameters tuning upstream of the cross-validation

The hyperparameters we tuned were the number of added fully connected dense layers, the number of nodes in these layers, their activation function, the optimizer, the initial learning rate, the weight decay, the dropout rate, the data augmentation amplitude and the batch size. Repeatedly tuning the values of the hyperparameters for different deep neural networks architectures and on the different cross-validation folds would have been prohibitively time and resource consuming. Instead, we sequentially explored how each hyperparameter was affecting the training and validation performances for a single architecture (InceptionV3) on a single cross validation fold (fold #0, see Methods - Training, tuning and predictions - Images - Cross-validation for the detailed description of the cross-validation) on a subset of the images datasets (raw sagittal brain images, eye fundus images, long axis carotid ultrasound images, four-chamber view heart MRI images, raw liver MRI images and coronal spine images). We then extrapolated the hyperparameter values to the other architectures, datasets and cross-validation folds. The hyperparameters combinations tested during the tuning can be found in Supplementary Table 41.

First, we maximized the batch size for each architecture. The maximum number of images per batch depends on the memory of the GPU and the size of the architecture, which itself depends on the dimensions of the image. The batch size for the different architectures can be found in Supplementary Table 43.

Then, we tested the learning rates, including 1e-6, 1e-5, 1e-4, 1e-3, 1e-2 and 1e-1. We observed that learning rates larger than 1e-4 prevented the model from converging for some runs. Second, we did not observe significant differences between the results obtained with learning rates smaller than 1e-4. We therefore set the initial learning rate to be 1e-4 for all models to shorten the time to convergence while ensuring that the learning rate was small enough to allow convergence and the finding of a local minima for the loss function.

Then we tested three different optimizers to perform the gradient descent: Adam[470], Adadelta[495] and RMSprop[496]. We did not observe any significant differences between the optimizers, so we set the optimizer to be Adam.

We then added different numbers of fully connected layers between the base CNN and side CNN's concatenated outputs and the final activation layer. We set the number of nodes to be

1,024 in the first added layer and then decreased the number of nodes by a factor of two for each successive layer. For example, if we added three fully connected layers, the number of nodes was 1024, 512 and 256. We added zero, one and five layers. We did not observe significant differences in the performance of the different architectures, so we set the number of fully connected layers to one.

We then tested powers of two from 16 to 2,048 as the number of nodes in this single layer. We did not observe significant differences between these architectures, so we set the number of nodes to be 1,024 to keep the number close to the initial number of nodes in the imported CNN architectures, as these were initially used to perform classification between 1,000 categories.

We tested two different activation functions for the activation functions of the fully connected layers we added in the side neural network and before the final linear layer. We did not observe any significant differences between the rectified linear units [ReLU][497] and the scaled exponential linear units [SELU][490] as activation functions, so we used the more common ReLU.

We then tested different levels of data augmentation. We introduced a hyperparameter that we called "data augmentation factor". The data augmentation factor modulates the amount of variation introduced by the data augmentation, while preserving the ratio between the different transformations. For example, a data augmentation factor of one is equivalent to the default data augmentation (see Preprocessing - Data augmentation - Images), but a data augmentation factor of two will double the ranges of the possible values sampled and the expected values for the vertical shift, the horizontal shift, the rotation and the zoom on the original images. We tested the following values for the data augmentation factor: 0, 0.1, 0.5, 1, 1.5 and 2. We found that different values for the data augmentation factor hyperparameter yielded similar results, as long as the data augmentation factor was not zero. We therefore set the data augmentation factor to be one when training the final models.

We then tuned the dropout rate for the fully connected layers we added. We tested the following values: 0, 0.1, 0.25, 0.3, 0.5, 0.75, 0.9 and 0.95. We observed that a dropout rate of 0.95 led to underfitting and that smaller values reduced overfitting on the training set but without improving the validation performance. As a consequence, we used a dropout rate of 0.5.

Finally, we tuned the weight decay. We tested the following values: 0, 0.1, 0.2, 0.3, 0.4, 0.5, 1, 5, 10 and 100. For the larger datasets, we found that weight decay values as low as 0.4 could lead to underfitting. We found that lower weight decay values reduced overfitting on the training set without significantly improving the validation performance. We set the weight decay to 0.1. Altogether, we found that hyperparameter tuning had little effect on the validation performance as long as extreme hyperparameters values were not selected.

##### Cross-validation

Training deep convolutional neural networks on images and videos is too time and resource consuming to perform a nested cross-validation. Therefore, we tuned the hyperparameters during the preliminary analysis, as described above. After hyperparameters tuning, we performed a simple outer cross-validation to obtain a testing prediction for each sample of the datasets, but we replaced the inner cross-validation with a simple split between the training fold and the validation fold (Supplementary Table 42). Although the hyperparameters were already tuned, a validation set was still required for two reasons: (1) to perform early stopping[487], a form of regularization. (2) to generate a set of validation predictions that are necessary for efficient ensemble building (see Methods - Models ensembling) and model selection. During the cross-validation, we scaled and centered the target variable (chronological age) as well as the side predictors (sex and ethnicity) around zero with a standard deviation of one, using the training summary statistics. Scaling the target and the input helps prevent the issues of exploding and vanishing gradients[498,499].

##### Cross-validation example

For the sake of clarity, let us walk through an example. Let us say that we want to generate unbiased predictions for every sample in a dataset using a CNN. First, we select the data fold #0 as the validation set, the data fold #1 as the testing set, and the remaining data folds (#2-9) as the training set. Then we scale and center the target (age), and the side predictors (sex and ethnicity) using the training mean and standard deviation: for each of the variables, we subtract the training mean to the variable on both the training, the validation and the testing set, and we divide it by the training standard deviation. We then train the model on the training set until convergence and select the architecture's parameters (also known as "weights") associated with the epoch that yielded the lowest validation RMSE. We then use the optimal weights to generate validation predictions for the data fold #0 and testing predictions on the data fold #1. Finally, we unscale the validation and testing predictions by multiplying them by the initial age training standard deviation before adding the initial age training mean to them. This completes the first cross-validation fold.

We then reiterate the process, this time using the data fold #1 as the validation set, the data fold #2 as the testing set, and the remaining data folds (#0 and #3-9) as the training set. We use the optimized weights to generate the validation predictions on the data fold #2, and the testing predictions on the data fold #3. We unscale the validation and testing predictions. This completes the second cross-validation fold. We reiterate the process eight more times to complete the cross-validation. We then concatenate the validation predictions from the ten data folds to obtain the final validation predictions, and the testing predictions from the ten data folds to obtain the final testing predictions.

#### Videos

We used a similar pipeline for the videos as we did for the images. First, we tuned the architecture and the hyperparameters upstream of the cross-validation, during the preliminary analysis. After setting the hyperparameters, we used the same cross-validation pipeline as for the images-based models to perform early stopping and to generate a testing prediction for every sample.

##### Hyperparameters tuning upstream of the cross-validation

Because training three-dimensional convolutional architectures to convergence requires computational times as long as one week, tuning the hyperparameters using a grid search or a random search would have been prohibitively time consuming. Instead, we sequentially tuned the hyperparameters using the data fold #0 as the validation set and the data folds #2-9 as the training set. We trained the models on the three-chamber view of the heart videos and extrapolated the hyperparameters obtained to other cross-validation folds and views of the heart.

First, we tuned the architecture of the convolutional block. We set as hyperparameter the number of filters in the last (fourth) convolutional layer. We tested 512, 758 and 1,024 filters, and we automatically set the number of filters in the penultimate (third) layer to be half that number. We found 1,024 to be the optimal number of filters. We also tested doubling the third convolutional layer. The two convolutions are stacked without using a max pooling operation, similarly to the original C3D[488] architecture. We set the number of filters for these two convolutional layers to be 128, which is twice the number of filters in the previous convolutional layer, and we set the number of filters in the fourth layer as an hyperparameter: 256, 512 and 1,024. We found that the double convolutional layer did not perform the single convolutional layer, so we conserved the latter architecture.

Then, we tuned the architecture of the dense block. We first set the number of dense layers to two, with the last layer being the final prediction layer with a single node. We set the number of nodes in the first dense layer as a hyperparameter and tested the following values: 512, 758 and 1,024. We found 1,024 to be the optimal number of nodes. We then tested adding a third fully connected layer between the first and the final dense layer with either 512, 768 or 1,024 nodes. We found that 512 was the optimal number of nodes, but that the three dense layers architecture underperformed compared to the two dense layers, so we conserved the latter.

Finally, we tuned the regularization hyperparameters. We tested dropout after the first convolutional layer with the following dropout rates: 0.2, 0.3 and 0.5. We found that a dropout rate of 0.2 was optimal and improved the generalization of the model. We then tested weight decay on this same layer with the following L2 regularizations: 1e-4, 1e-3, 1e-2. We found that the optimal value was 1e-3 but that using weight decay led to the model underperforming, so our final model does not use weight decay. Finally, we tuned the data augmentation parameter. We randomly rotated the images in the spatial plane to prevent overfitting, sampling the rotation angle from the uniform distribution [-angle, +angle]. We tested three different angle values: three degrees, five degrees and seven degrees. We found the optimal value to be three degrees.

#### Time series

We tuned the time series models in two steps. (1) We tuned the hyperparameters using a single cross-validation fold. (2) We tuned the seed and performed early stopping (patience=40) using a simple cross-validation, to generate a testing sample size for every sample. These two steps are described in detail below.

#### Hyperparameters tuning upstream of the cross-validation

The hyperparameters we tuned were the number of convolutional layers in the architecture, the number of nodes in the dropout rate and the strength of the kernel and the bias regularizations. The tuned the hyperparameters using a single cross-validation fold. Specifically, we used the data fold #0 as validation set and the data folds #2-9 as training set. We used the same pipeline as for the images and the videos, with two differences. (1) Because the training of models built on time series was significantly faster than the training of models built on images or videos, used a grid search to tune the hyperparameters rather than tune them sequentially. (2) We scaled the input differently. For the models built on the ECGs and on the raw acceleration data across the full week, we did not scale the data because we adapted architectures from publications in which the input has not been scaled. The models built on PWA, features extracted from acceleration data and three-dimensional walking data, we normalized every sample separately by dividing each lead by the absolute value of its maximal value for the sample. We then used this maximum value as a scalar predictor which we refer to in this paper as “scaling factor”.

We tested architectures with one to ten convolutional layers for the convolutional block. The default number of filters for each convolutional layer doubled with every layer, starting from 16 and capped at 1,024. For example, the default number of filters for a nine layers deep convolutional block would be 16, 32, 64, 128, 256, 512, 1,024, 1,024 and 1,024. To allow the architecture to increase its breadth without increasing its depth, we introduced another hyperparameter which we called the “filters factor”, with a value of either one, two or four. The number of filters in each convolutional layer was the default number of filters multiplied by the filters factor value. For example, the number of filters for a three layers deep convolutional block with a filter factor of four would be 64, 128 and 256, instead of 16, 32 and 64. For the dropout rate, we defined the hyperparameter space as eleven values uniformly spread between 0 and

50%, included. For the kernel and the bias regularizers, we defined the hyperparameter space as the absence of regularization (0) and every negative power of 10 between three (10e-3) and six (10e-6), included. We used the hyperparameters values selected on this first cross-validation fold for the nine remaining cross-validation folds. The hyperparameters values selected can be found in Supplementary Table 45.

##### Tuning of the seed using a cross-validation

We observed that the hyperparameters values selected on the first cross-validation fold did not always perform as well for the other cross-validation folds. Similarly, we observed that similar hyperparameter values combinations could lead to significantly different performances on the first cross-validation fold. We hypothesized that these differences could be partly driven by different random initializations of the weights of the neural network, which led to convergence to different local minima. To mitigate this effect, we tuned the Keras seed independently for each of the ten cross-validation folds. We tested every integer between zero and nine, included, and selected the seed that yielded the best model in terms of performance on the validation set.

The cross-validation also served to generate a testing prediction on every sample and to perform early stopping, as described under the pipeline for images.

### Interpretability of the predictions

#### Scalar data-based predictors

For elastic nets, we interpreted the models using the values of the regression coefficients. Large absolute values for these coefficients means they played an important role when generating the predictions. For gradient boosted machines we used the feature importances, which are based on the number of times a tree selected each of the variables. Variables with high feature importances were selected more often and are therefore likely to play a key role in predicting chronological age. For neural networks, we estimated the importance of each feature by permuting it randomly between samples before computing the performance of the model. The score of each feature is the difference between the R-Squared value before and after the random permutations. Features whose random permutation leads to a large decrease in the model's performance are estimated to be important predictors of chronological age.

We estimated the concordance between the three different algorithms by computing the Pearson and the Spearman correlations between their feature importances.

#### Time series, image- and video-based predictors

To interpret the CNNs built on time series, images and videos, we used saliency maps[500]. For time series and images, we coded the saliency maps using the keract python library. For videos, we generated a saliency map for each time frame using the iNNvestigate python library [501]. For each input sample, a saliency map uses the gradient of the final prediction with respect to each individual input pixel to estimate whether changing the value of this pixel would affect the prediction. Pixels for which the gradient is close to zero are not important, whereas pixels with a large gradient are estimated to be important. For videos, we computed both a saliency map for each time frame, which we stored as a .gif file, and an average saliency map over all time frames.

For images, we built a second attention map using a custom version of the Gradient-weighted Class Activation Mapping [Grad-CAM] algorithm[502] adapted to regression rather than multi-class classification: Gradient-weighted Regression Activation Mapping [Grad-RAM]. The intuition behind Grad-CAM maps is that they are similar to saliency maps[502], but instead of computing the gradient with respect to the input image, they compute it with respect to the activation of the last convolutional layer. As convolutional layers maintain the spatial organization of the input image,

Grad-CAM can still identify which region of the image is driving the predictions. Because Grad-CAM does not have to backpropagate the gradient all the way back to the input image, it is considered a less noisy alternative to the saliency maps. In the same way that saliency maps need to combine the attention maps generated in the different input channels (e.g. RGB) into a single activation map, Grad-CAM must combine the attention maps generated on the different filters of the last convolutional layer. For example, the last convolutional layer for InceptionResNetV2 has 1,792 filters. Grad-CAM combines these 1,792 attention maps into a single attention map using a linear combination. In the initial Class Activation Mapping [CAM] algorithm[503], generating CAM activation maps required to retrain the model after modifying the architecture and replacing all the fully connected layers after the final convolutional layer with a global max pooling operation, which converted each filter into a scalar feature. The intuition behind this substitution was that each filter could be interpreted as detecting a specific feature, and global max pooling yielded a scalar that could be interpreted as the presence (high value) or absence (low value) of the feature anywhere on the image. The scalar values were then linearly combined and activated using the softmax function to yield the probabilities of belonging to different classes. To obtain the activation map for a specific class, the filters of the last convolution layer were linearly combined using the weights connecting the scalar features obtained after the max pooling operation to the final prediction score for that class. CAM was later improved to become Grad-CAM[502]. Grad-CAM saves the need for modifying the architecture of the model and retraining it by approximating the linear regression weight for each final convolutional filter by the mean activation gradient over the pixels of the filter. The intuition behind this approximation is that a filter's pixel is important if changing its value affects the final prediction, so a high average gradient over the pixels of the filter justifies that this filter should be given a higher weight when merging all the filters into a single attention map. To adapt Grad-CAM to our regression task we (1) computed the derivatives of the chronological age prediction rather than a class' prediction and (2) removed the ReLU activation applied to the weighted sum of the last convolutional filters, which we replaced by an absolute value. The rationale is that for (Grad-)CAM maps, we only want to highlight the regions of the picture which are associated with a high probability for the class. In contrast, for (Grad-)RAM we care as much about the regions of the input image that can strongly increase the chronological age prediction as about the regions that can strongly decrease it. Because the filters in the last convolutional layer are the result of the processing of the input image by several convolutional layers with possibly negative weights, the sign of the last convolutional layer's pixels and regression weights cannot be linked to either accelerated aging or decelerated aging, only to the magnitude of the shift that would affect the prediction if each region of the input image was modified. Regression Activation Mapping (RAM) was mentioned as a possible extension of CAM in the original CAM paper[503] and has been used to interpret models CNNs built on retinal images[504] and cortical surfaces[505], but we are to our knowledge the first to describe the generalization of Grad-CAM to a regression task. One notable difference between our implementation and Wang and Yang.'s implementation[504] is that we are taking the absolute value of the final attention map, as mentioned above. We found that not taking the absolute value led to misleading attention maps for participants with high chronological age predictions. The attention map highlights important areas with negative values, which are therefore depicted in blue, a color otherwise associated with unimportant regions in traditional CAMs. Inversely, regions on the input image for which the attention map has a slight positive value are spuriously considered to be the most important and are highlighted in red. We therefore advise that RAM or Grad-RAM be implemented using an absolute value. We coded Grad-RAM using the get_activations and get_gradients_of_activations functions of the keract python library.

It is important to understand that unlike the feature importances described under "Scalar data-based predictors", which describe the model itself, attention maps are sample specific. In other words, they can be used to explain which features drove the predictions for a specific inputted sample but cannot provide an explanation for the way the model is performing predictions in general.

For each aging sub-subdimension, we generated the attention maps for the best performing CNN architecture. We selected representative samples for which we computed the different attention maps. We computed attention maps for the two sexes (female and male), for three age ranges (ten youngest ages, ten middle ages and ten oldest ages of the chronological age distribution) and for three aging rates (accelerated agers, normal agers, decelerated agers). For each intersection of the three categories listed above, we selected the ten most representative samples (e.g. the ten most accelerated agers among young males). The figures in this paper only present the first, most representative of these ten samples. The complete set of samples can be found on the website.

An example for the two attention map filters can be found in Supplementary Figure 78.

### Models ensembling

#### Ensembles hierarchy

The sample size of the intersection between all the aging main dimensions when building the ensemble model at the highest level was small. To mitigate this problem and increase the sample size of the intersection, we excluded the biochemistry subcohorts from the ensemble. We then built three additional separate models at the highest hierarchical level. The first model was built on a subset of the datasets collected during the instances 0 and 1 as a compromise between the breadth of the predictors included and the sample size of the intersection. The datasets included were the eyes, hearing, lungs, biochemistry and blood cells datasets, as well as the non-images arterial and musculoskeletal datasets. We refer to this model as “*instances01”. We built the second model on the datasets collected during the instances 2 and 3, which consist of the brain, arterial, heart, abdomen and musculoskeletal datasets. We refer to this model as “*instances23”. We built the third model on datasets issued from the instances 1.50, 1.51, 1.52 and 1.54. Because the only dataset covering these instances is physical activity, this ensemble model is a “dummy” ensemble identical to the ensemble model built on the aging main dimension “physical activity”. We nonetheless included this model to maintain the symmetry of the pipeline. We refer to this model as “*instances1.5x”. The aging main dimensions included in the four highest level ensemble models can be found in Supplementary Table 44.

All ensemble models are noted with “*” for the parameters over which they have been ensembled. For example, the ensemble model at the second lowest level has “*” for the sub-subdimension field instead of “TowerRearranging” or “TrailMaking” and “*” for the algorithm field instead of “ElasticNet”, “LightGBM” or “NeuralNetwork”.

#### Ensembling algorithm

We built each ensemble model separately on each of the ten data folds. For example, to build the ensemble model on the testing predictions of the data fold #1, we trained and tuned an elastic net on the validation predictions from the data fold #0 using a 10-folds inner cross-validation, as the validation predictions on fold #0 and the testing predictions on fold #1 are generated by the same model (see Methods - Training, tuning and predictions - Images - Scalar data - Nested cross-validation; Methods - Training, tuning and predictions - Images - Cross-validation). We used the same hyperparameters space and Bayesian hyperparameters optimization method as we did for the inner cross-validation we performed during the tuning of the non-ensemble models.

To summarize, the testing ensemble predictions are computed by concatenating the testing predictions generated by ten different elastic nets, each of which was trained and tuned using a 10-folds inner cross-validation on one validation data fold (10% of the full dataset) and tested on one testing fold. This is different from the inner-cross validation performed when training the non-ensemble models, which was performed on the “training+validation” data folds, so on 9 data folds (90% of the dataset).

#### Generating average predictions for each participant

Each ensemble model could only be built on the samples belonging to the intersection of the models included in the ensemble scope. However, the intersection between some of the datasets is limited or non-existent. For example, datasets exclusively collected in instances 0 and 1 (e.g. eye fundus images) have no intersection with datasets exclusively collected in instances 2 and 3 (e.g. heart MRI videos) in terms of sample size. To mitigate this issue, we generated an average prediction for each individual, reported to the instance 0.

We walk through an example. Let us assume a participant had two brain MRI samples collected from them in instances 2 and 3, respectively at age 70 and 80. Let us assume that the age predictions were respectively 64 and 78, so the residuals are respectively -6 years and -2 years, for an average of -4 years. However, we still need to take into account the bias in the residuals, defined as the difference between the participant’s chronological age and the prediction. As explained in more details under Methods - Biological age definition, we observed a bias in the residuals as a function of chronological age. Participants on the younger end of the chronological age distribution tend to be predicted older than they actually are, whereas participants on the older end of the distribution tend to be predicted younger than they actually are. We need to properly account for this bias when translating a prediction from a more recent instance to an older instance. Let us assume that the average bias in the residuals for participants who are 70 and 80 years old is respectively -2 years and -4 years. After correcting for this bias, the predictions are now respectively 64-(-2)=66 and 78-(-4) = 82. Therefore, the corrected residuals for this participant are respectively -4 years and +2 years, for an average of -1 years. Finally, let us assume that the participant was 60 years old in instance 0. We will assign a single brain MRI prediction of 60-1=59 years to the participant, but we still need to un-correct for the bias in residuals. Let us assume that the average bias for the residuals at age 60 is +5 years. We will assign a final prediction for the participant of 59+5=64 years. This new set of predictions reported on the instance 0 is more likely to have a non-zero sample size overlap with other predictors based on datasets collected on instance 0 (e.g. blood biomarkers) and can therefore be leveraged by the ensemble builder.

A key point we would like to highlight here is that we did not actually correct for the bias in the residuals at this step of the pipeline. Instead, we corrected then un-corrected the predictions that we translated from different instances to the instance 0. The actual correction for the residual biases takes place when defining the biological age phenotypes (see Methods - Biological age definition).

To distinguish between raw predictions on the instance 0, and the average predictions reported to the instance 0, we created a new instance which we named instance “*”. We refer to these predictions as “participants predictions”, as opposed to “samples predictions”.

#### Correlation between aging dimensions

To compute the correlations at the first four levels of hierarchy, we used the ensemble models. For example, to estimate the correlation between brain aging and heart aging, we computed the correlation between the corrected residuals for the ensemble model built on all the brain-based models and the corrected residuals for the ensemble model built on all the heart-based models. For example, for the first level of the hierarchy, that is the different biological dimensions, we computed the correlation between the corrected residuals for the ensemble model built on all the brain-based models and the corrected residuals for the ensemble model built on all the heart-based models to estimate the correlation between brain aging and heart aging.

For the second level of the hierarchy, that is the different biological subdimensions, we computed the correlations for the subdimensions of brain aging, eye aging, arterial aging, heart aging, abdomen aging, musculoskeletal aging, physical activity aging and biochemical aging. For brain aging, we computed the correlation between cognitive aging and anatomical (MRI-based) aging. For eye aging, we computed the correlation between OCT-based, fundus, intraocular pressure, acuity and autorefraction aging. For arterial aging, we computed the correlation between pulse wave analysis aging and carotids aging. For heart aging, we computed the correlation between anatomical (MRI-based) aging and electrical (ECG-based) aging. For abdomen aging we computed the correlation between liver and pancreas aging. For musculoskeletal aging we computed the correlation between spine, knees, full body and scalar features-based aging. For physical activity we computed the correlation between "full week" and walking agings. For biochemical aging, we computed the correlation between blood and urine aging.

To compute the correlations at the third level of the hierarchy, that is between the different biological sub-subdimensions, two sub-dimensions had biologically distinct sub-subdimensions: brain cognitive aging and musculoskeletal "scalar features-based" aging. For brain cognitive aging, we computed the correlations between reaction time, matrix pattern completion, tower rearranging, symbol digit substitution, paired associative learning, prospective memory, numeric memory, fluid intelligence, trail making and pairs matching. For musculoskeletal aging we computed the correlations anthropometry, impedance, heel bone densitometry and hand grip strength.

To compute the correlations at the fourth level of the hierarchy, that is between the different views or different features of the same biological dimension, we computed the correlation between the different views for the brain MRI images (sagittal plane, coronal plane, transverse plane, diffusion MRI weighted means, subcortical volumes, gray matter volumes), arterial pulse wave analysis records (scalars and time series), heart ECG images (scalars and time series), heart MRI images (two-chamber, three-chamber and four-chamber views), spine X-rays (sagittal and coronal), full body X-rays (figure and flesh) and full week physical activity (scalars, acceleration, time series features, Gramian angular field-difference, Gramian angular field-summation, Markov transition field, recurrent plot).

To compute the correlations at the fifth level, that is between the different image preprocessing methods, we compute the correlation between raw and contrasted images (heart, liver and pancreas MRI images), between raw and reference images (brain MRI images) and between the full body figure and skeleton images.

To compute the correlations at the sixth level, that is between the different algorithms, we computed the correlation between the algorithms trained on scalar features (elastic net, light GBM and neural network) and, separately, between the CNN architectures (InceptionResNetV2 and InceptionV3).

#### Non-genetic correlates of accelerated aging

Unlike DNA, biomarkers, phenotypes, diseases, family history, environmental variables and socioeconomics can change over life. As a consequence, we compared each biomarker, phenotype and environmental variable with the accelerated aging of the participant at the time the exposure was measured and we used the “Samples predictions”, as opposed to the “Participants predictions” that we used for the identification of genetic correlates (see Methods - Models ensembling - Generating average predictions for each participant).

##### Imputation of the non-genetic X-variables

Most X-variables were not collected on all four instances. Additionally, no X-variables were collected at the same time as the accelerometer data was collected. To identify the non-genetic correlates of accelerated aging, we had to impute the values of the X-variables for the ages of the participants for which they were not available. We considered two imputation methods, which we refer to as the “cross-sectional” and the “longitudinal” imputations.

For the cross-sectional imputation, we computed a linear regression for each X variable as a function of age, adjusting for sex. We then used the slope of the linear regression to extrapolate the value of the XWAS variable at different ages.

For the longitudinal imputation, we first selected, for each X variable, all the participants that had at least two measures taken for this X variable. We then performed a linear regression for each participant. We then averaged the slope of the linear regressions over all the participants of the same sex. Finally, we used this slope to extrapolate the value of the XWAS variable at different ages for all participants depending on their sex, in the same way we did it for the cross-sectional imputation.

It is important to notice that for both the cross-sectional imputation and the longitudinal imputation, data can only be imputed when the XWAS variable has been measured at least once for the participant. This raw measure is then used to extrapolate which value the X variable was likely taking a couple years earlier and/or later.

The advantage of the cross-sectional imputation is larger sample sizes. The advantage of the longitudinal method is that it corrects for generational effects. For example, old people have shorter legs than young people on average[30]. This is not because human legs shrink as we grow older. Instead, people who are old today already had shorter legs when they were young. If the cross-sectional regression is used to impute the length of the participants on instances where it was not measured, it will spuriously assign smaller values to the older samples. In contrast, the longitudinal regression learns the regression coefficient by comparing each participant to themselves as they age and will therefore not capture the generational effect. When used to predict the participants legs' length, it will impute constant values over time. To evaluate which of the two imputation methods should be preferred, we used them to predict X-variables for which we knew the actual values and computed the R-Squared values associated with the predictions. We found that, even with sample sizes as small as 200 samples, longitudinal imputation outperformed cross-sectional imputation. We therefore used longitudinal imputation.

#### X-Wide Association Studies

First, we tested for associations in an univariate context by computing the partial correlation between each X-variable and each of the 28 accelerated aging dimensions. To compute the partial correlation between an X-variable and an aging, we followed a three steps process. (1) We ran a linear regression on each of the two variables, using age, sex and ethnicity as predictors. (2) We computed the residuals for the two variables. (3) We computed the correlation between the two residuals and the associated p-value if their intersection had a sample size of at least ten samples. We used a threshold for significance of 0.05 and corrected the p-values for multiple testing using the Bonferroni correction. We plotted the results using a volcano plot. We refer to this pipeline as an X-Wide Association study [XWAS].

For each aging dimension, we list the biomarkers and phenotypes and environmental variables categories that are significantly associated with both accelerated aging and decelerating aging after Bonferroni correction for multiple testing. We order these X-categories by increasing p-value of their most significantly associated representative, and we provide up to three examples by category. If more than three variables were significantly associated with accelerated aging, we only list the first three and precede the list by “e.g”. If the same variable is present in several X-categories (e.g. heart rate can be found in heart function, ECG, pulse wave analysis and blood pressure), we only report it for the first category listed. For the sake of brevity, we shorten the name of the variables when possible and we do not report highly similar features such as features for paired organs (e.g. left arm and right arm impedance) or brain MRI weighted means highlighting the same region (e.g. weighted mean ICVF in tract anterior thalamic radiation and weighted mean OD in tract anterior thalamic radiation are reported as “anterior thalamic radiation”).

#### Prediction of accelerated aging

We leveraged the pipeline we built to predict chronological age as a function of scalar biomarkers (see Methods - Machine learning algorithms - Scalar data; and Methods - Training, tuning and predictions - Scalar data) to predict accelerated aging for the 28 aging dimensions as a function of the biomarkers, the pathologies and the environmental variables. We leveraged the pipeline to identify which features were driving the predictions, as well (see Methods - Training, tuning and predictions - Interpretability of the predictions - Scalar data-based predictors).

We built a model for each X-variables category (Supplementary Table 6, Supplementary Table 11, Supplementary Table 16, Supplementary Table 21, Supplementary Table 24, Supplementary Table 29). In doing so, we encountered the same difficulty as described under Methods - Data types and Preprocessing - Scalar data - Hierarchical clustering of the predictors: taking the intersection of all the variables yielded small sample sizes. To mitigate this issue and find a compromise between the number of variables included in the model and the sample size of their intersection, we leveraged the hierarchical clustering algorithm described under Methods - Data types and Preprocessing - Scalar data - Hierarchical clustering of the predictors.

#### X-Correlations between the accelerated aging dimensions

We estimated the X-correlations between each pair of aging dimensions, for each X-variables category in two different ways. (1) By computing the correlation between the partial correlations generated during the XWAS, that is in a univariate context. (2) By computing the correlation between the feature importances of the models built to predict accelerated aging, that is in a multivariate context.

##### X-Correlations based on the XWAS results

For the sake of clarity, let us walk through an example. Let us say we want to compute the lifestyle correlation between accelerated brain aging and heart aging. The XWAS generates a vector whose components are the partial correlations between the accelerated aging phenotype and each lifestyle variable, for both accelerated aging and heart aging. We compute three different Pearson correlations between these two partial correlation vectors. (1) The “All” correlation, using all the components of the two vectors. This correlation tends to be inflated by the large number of X-variables whose correlation with both accelerated aging dimensions is close to zero. (2) The “Intersection” correlation, using only the lifestyle variables that were significantly associated with both of the accelerated aging dimensions. Because the cardinality of the intersection can be small, a small number of X-variables can yield very high or very low correlations. (3) The "Union" correlation, using only the lifestyle variables that were significantly associated with at least one of the two accelerated aging dimensions. The "Union" correlation represents a compromise between the "All" and the "Intersection" correlations. The figures in this paper were generated using the "Union" correlation, but all three correlations can be explored on the website.

##### X-Correlations based on the feature importances

We then computed the correlations between the feature importances for different accelerated aging dimensions to estimate the X-correlation between the different dimensions. We used the same method as described above under "X-Correlations based on the XWAS results", replacing the coefficient obtained for each X-variable in a univariate context (using partial correlation with accelerated aging) with the coefficient obtained in a multivariate context (as an accelerated aging predictor in a multivariate model).

### Supplementary Figures

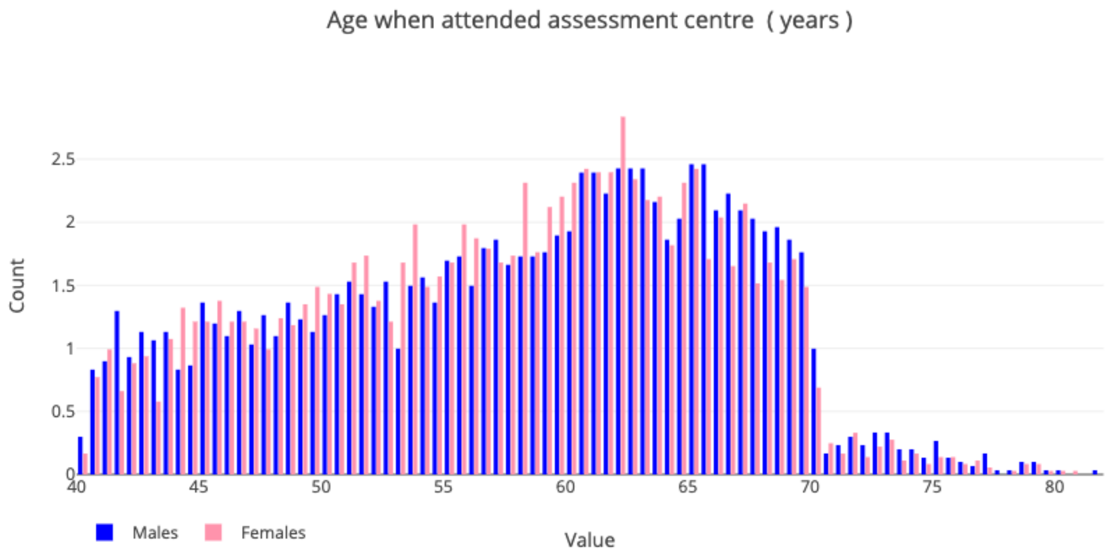

**Supplementary Figure 1: Demographics of the UK Biobank cohort**

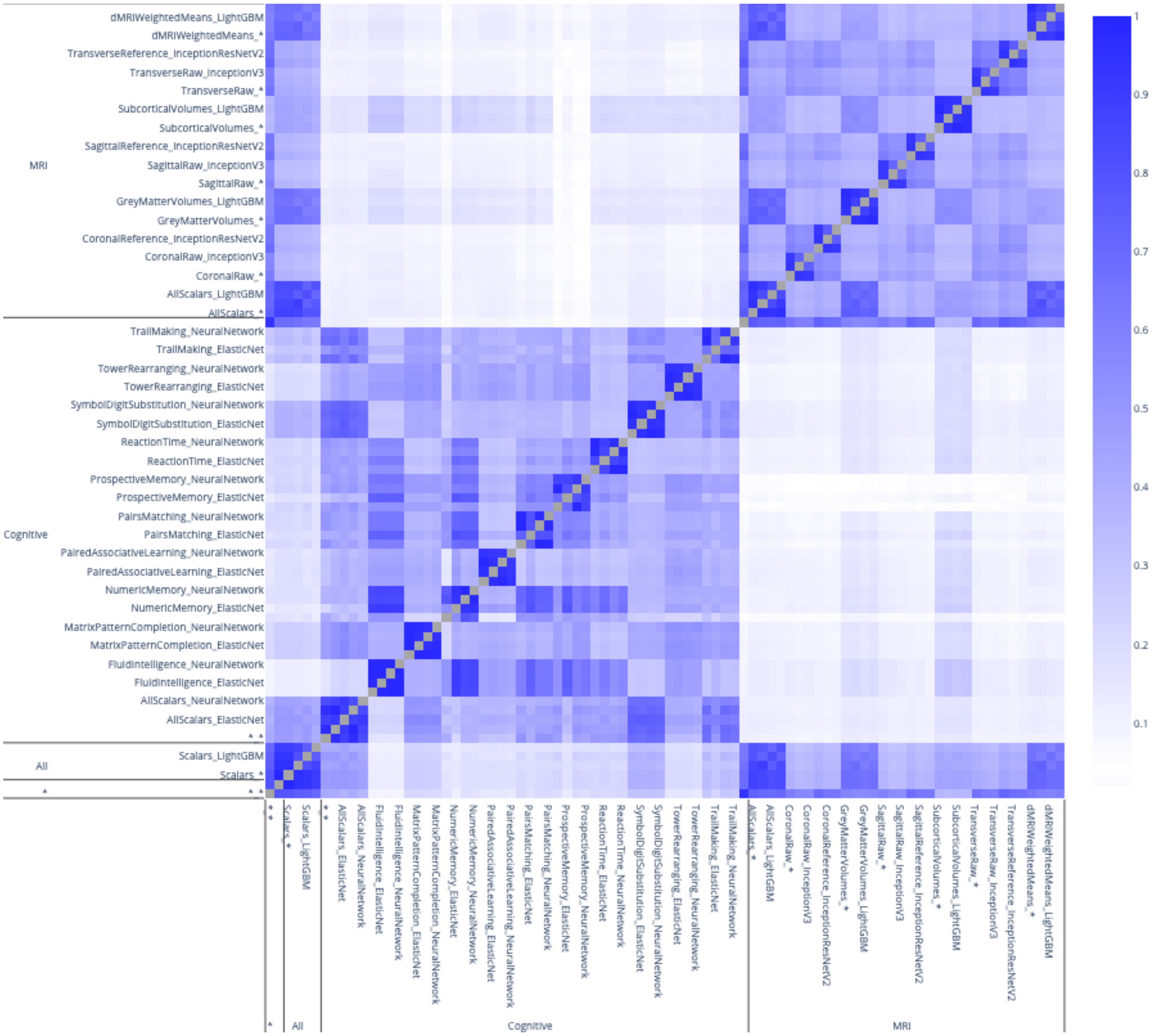

**Supplementary Figure 2: Phenotypic correlation between accelerated brain aging dimensions**

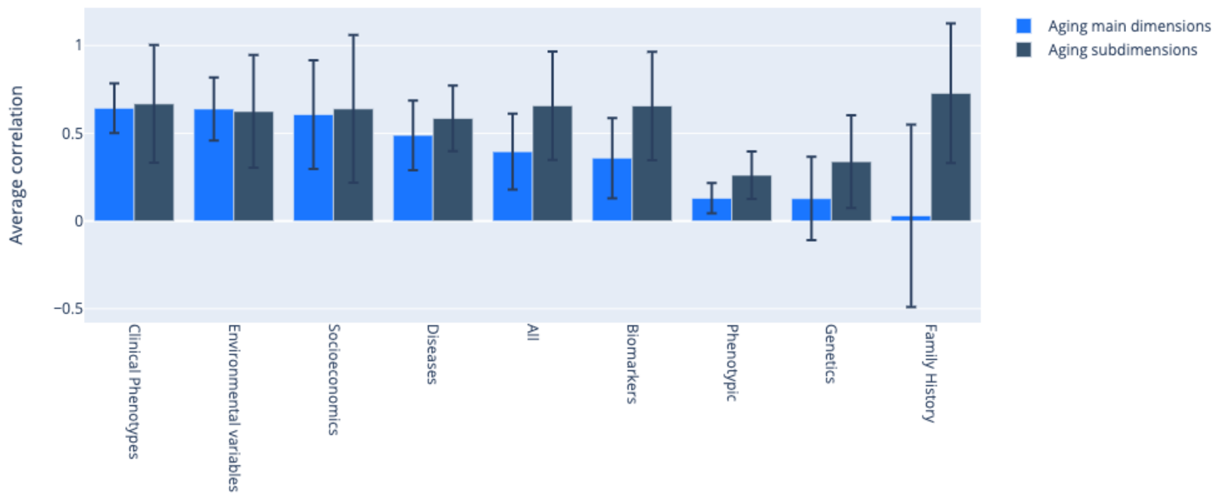

**Supplementary Figure 3: XWAS Summary - Average correlation between aging main dimensions and between aging subdimensions for each X category**

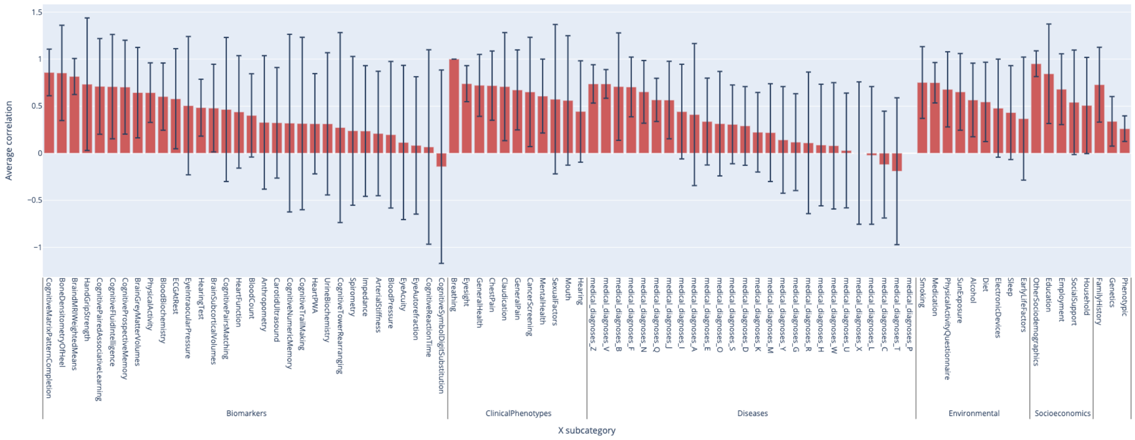

**Supplementary Figure 4: XWAS Summary - Average correlation between aging subdimensions for each X-subcategory**

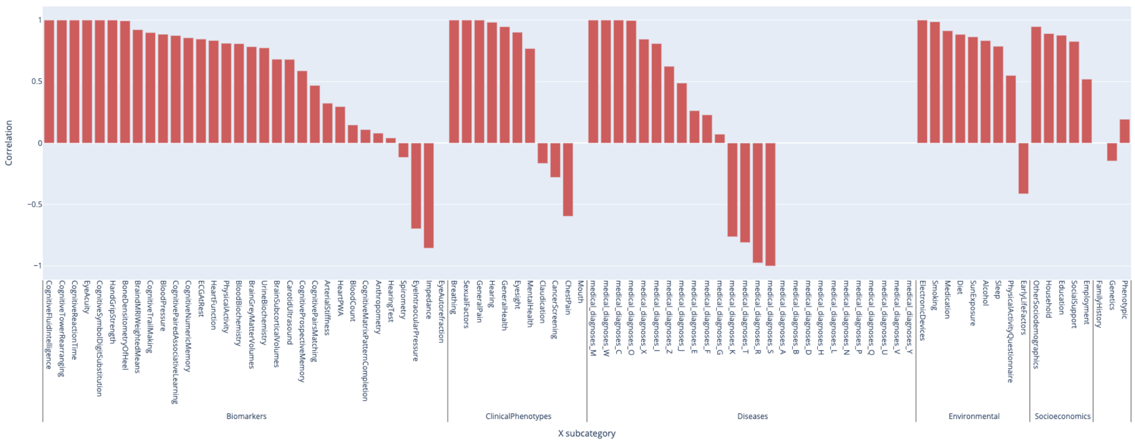

**Supplementary Figure 5: XWAS Summary - Correlation between brain and heart accelerated aging for each X-subcategory**

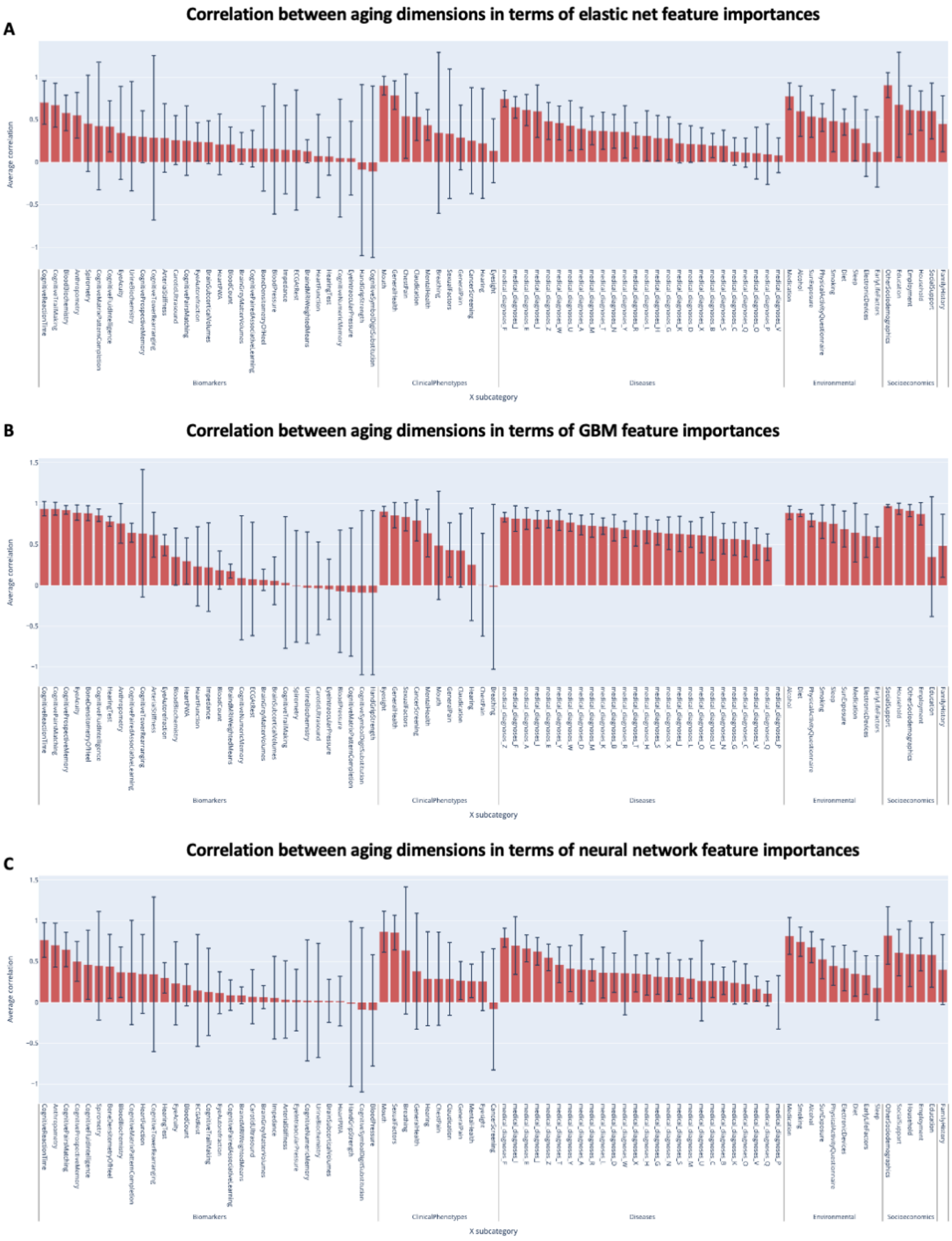

**Supplementary Figure 6: XWAS Summary - Average correlation between aging main dimensions and between aging subdimensions for each X category, based on accelerated aging predictive models' feature importances**

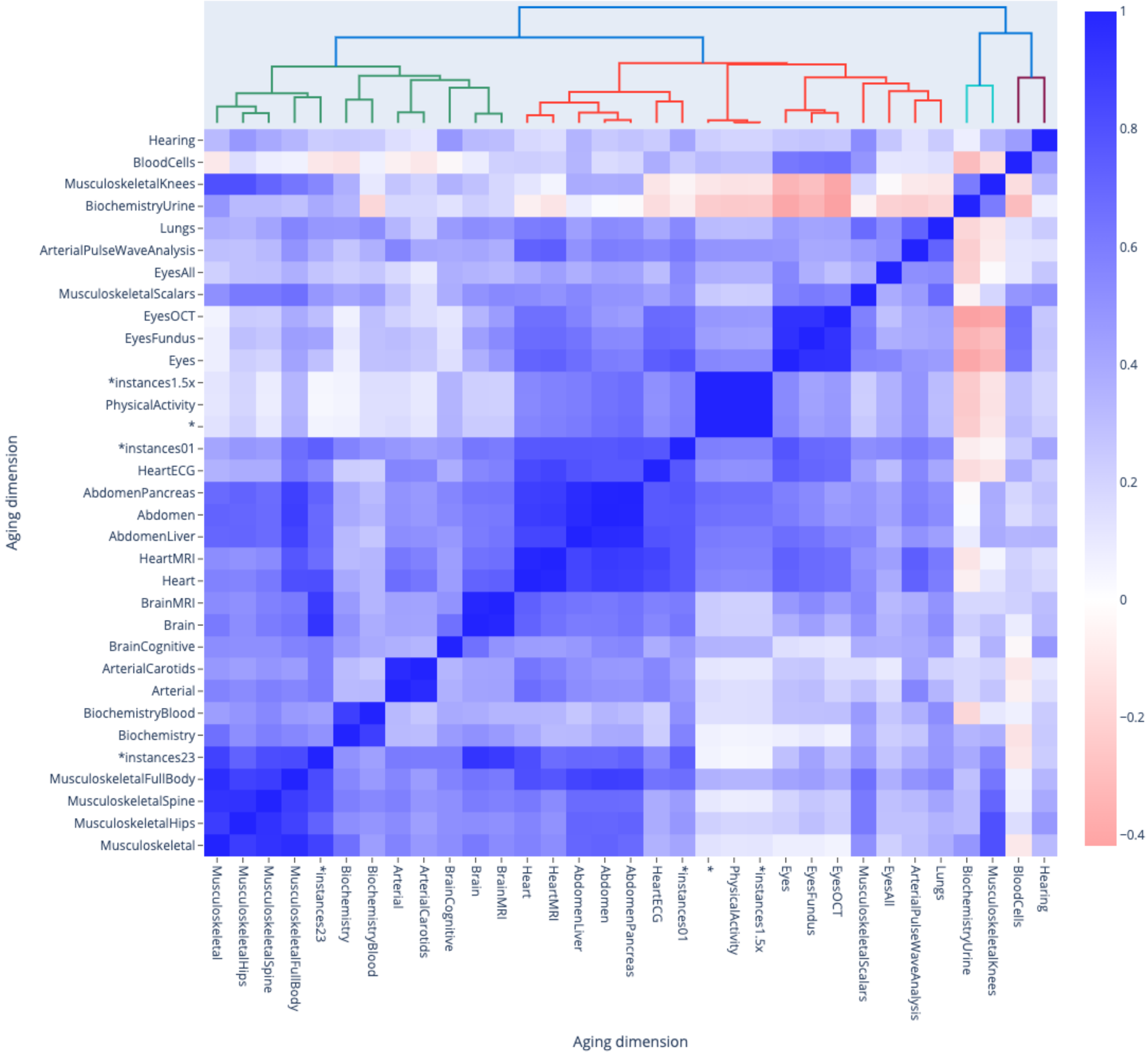

**Supplementary Figure 7: Correlation between the main aging dimensions and 17 selected aging subdimensions in terms of associated biomarkers**

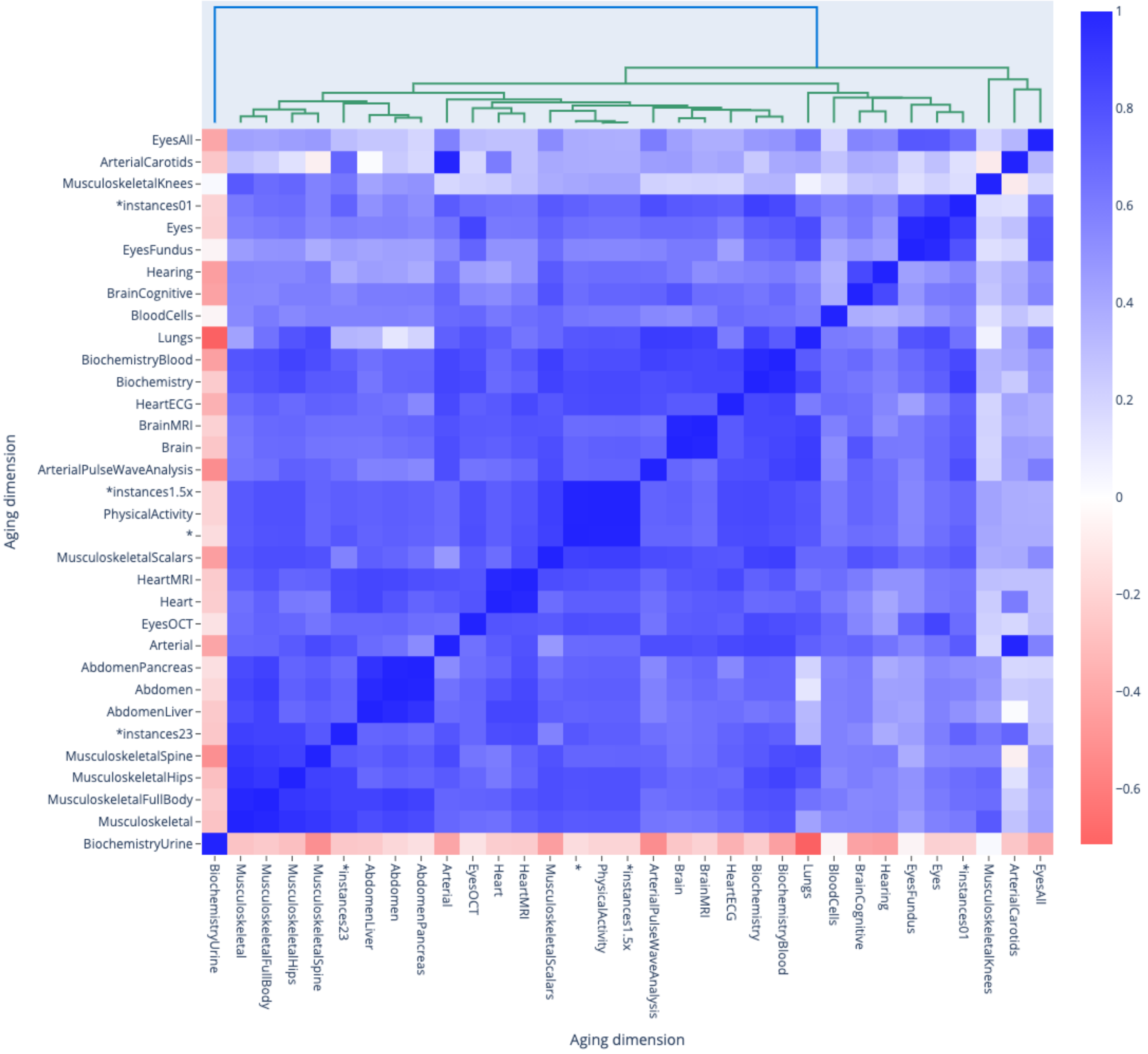

**Supplementary Figure 8: Correlation between the main aging dimensions and 17 selected aging subdimensions in terms of associated clinical phenotypes**

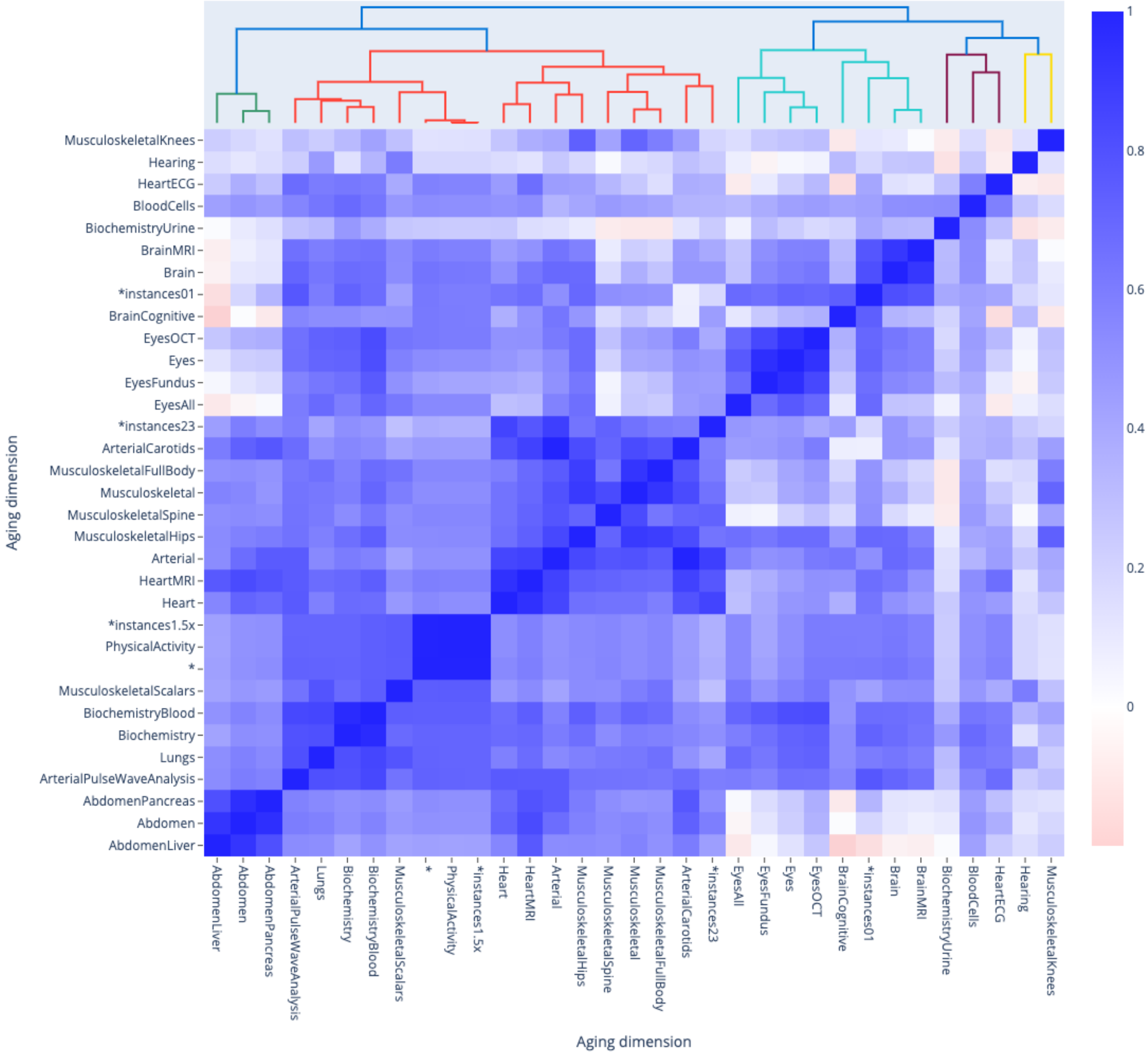

**Supplementary Figure 9: Correlation between the main aging dimensions and 17 selected aging subdimensions in terms of associated diseases**

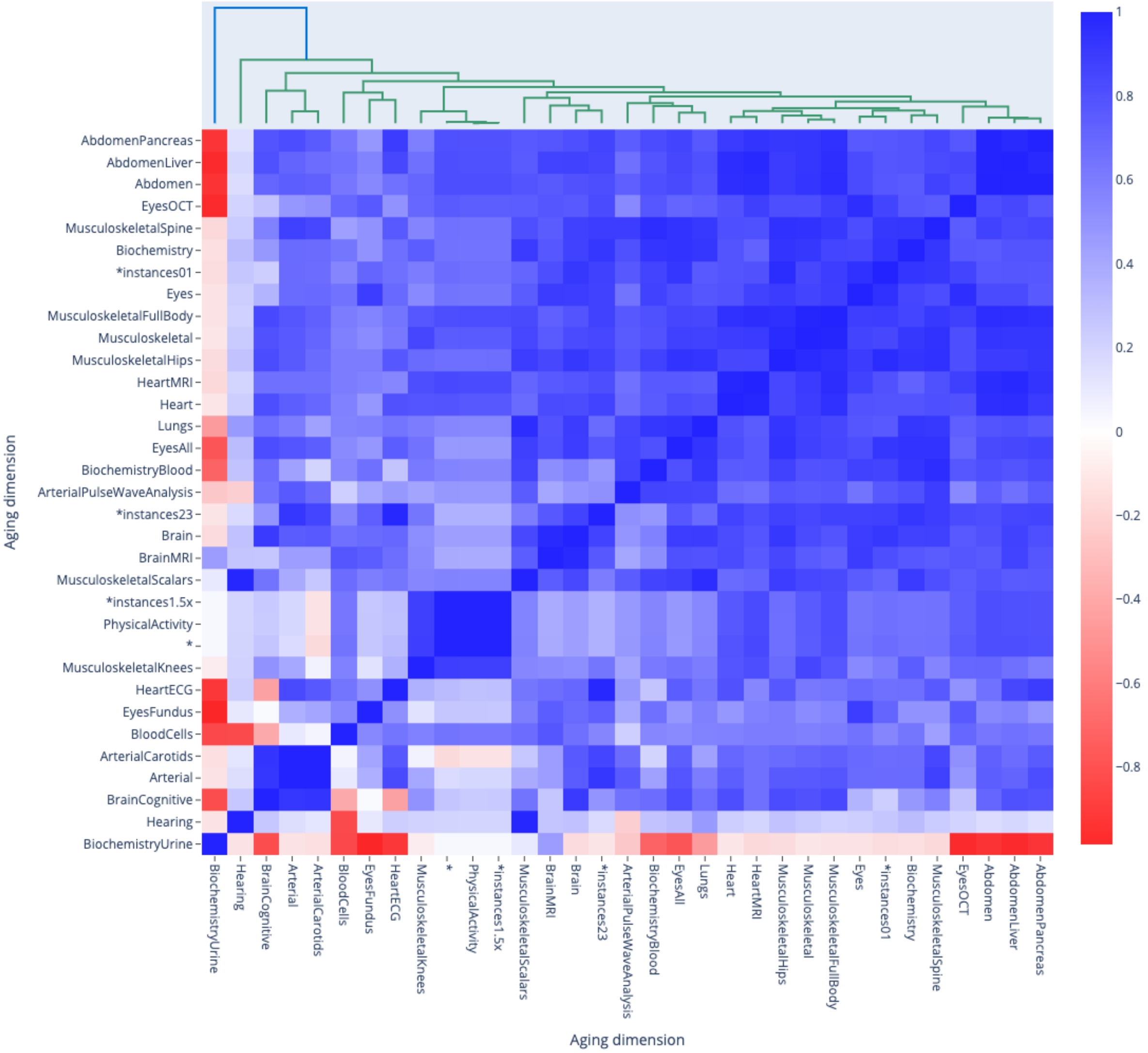

**Supplementary Figure 10: Correlation between the main aging dimensions and 17 selected aging subdimensions in terms of associated socioeconomics**

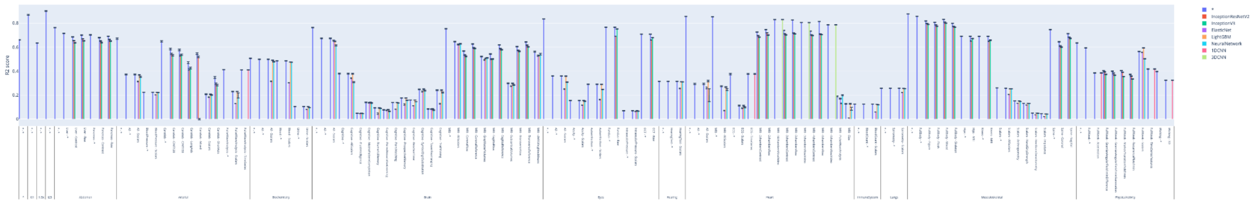

**Supplementary Figure 11: Chronological age prediction accuracy for all models**

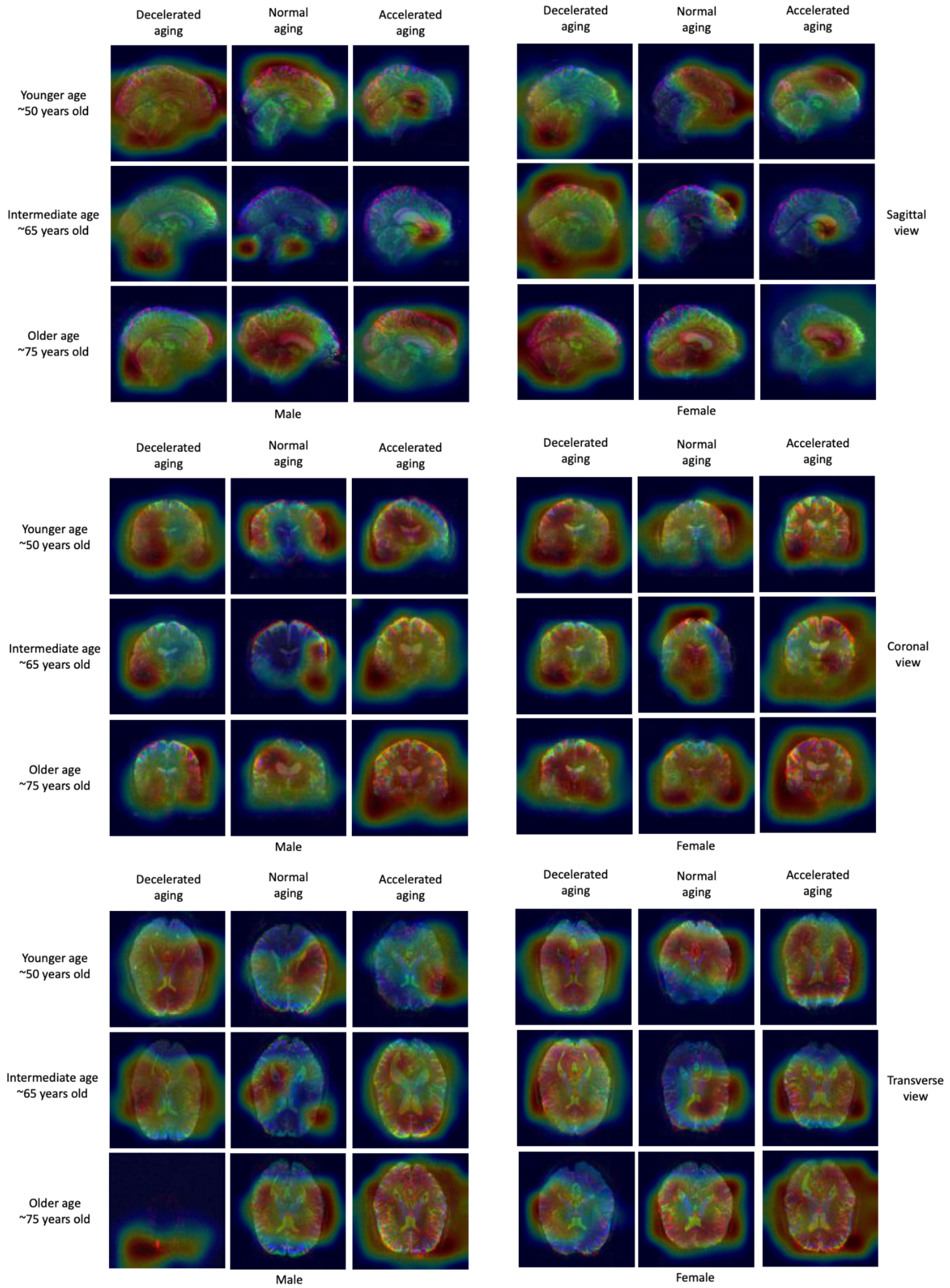

**Supplementary Figure 12: Attention maps for brain MRI images (reference scan)**

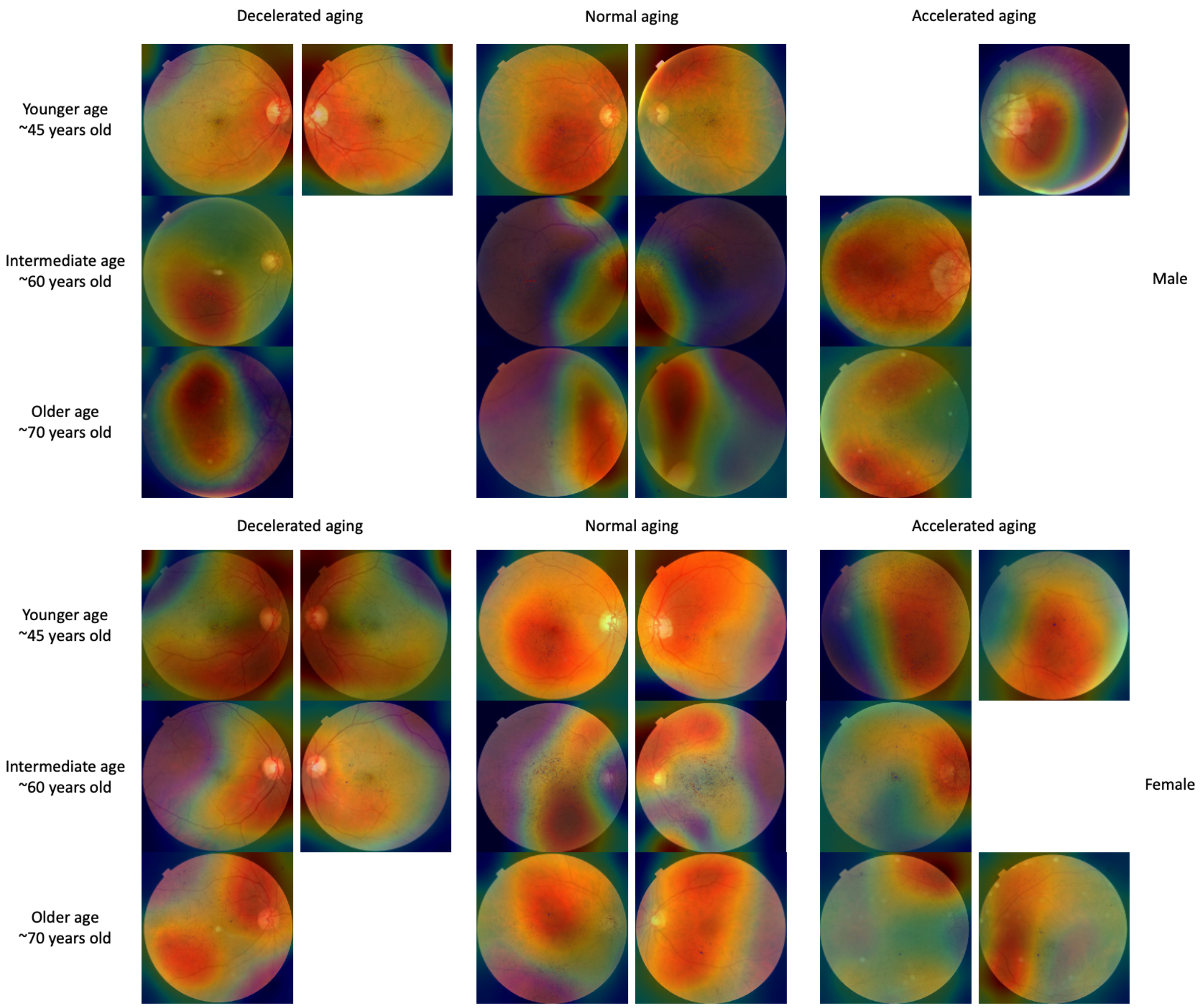

#### Supplementary Figure 13: Attention maps for eye fundus images

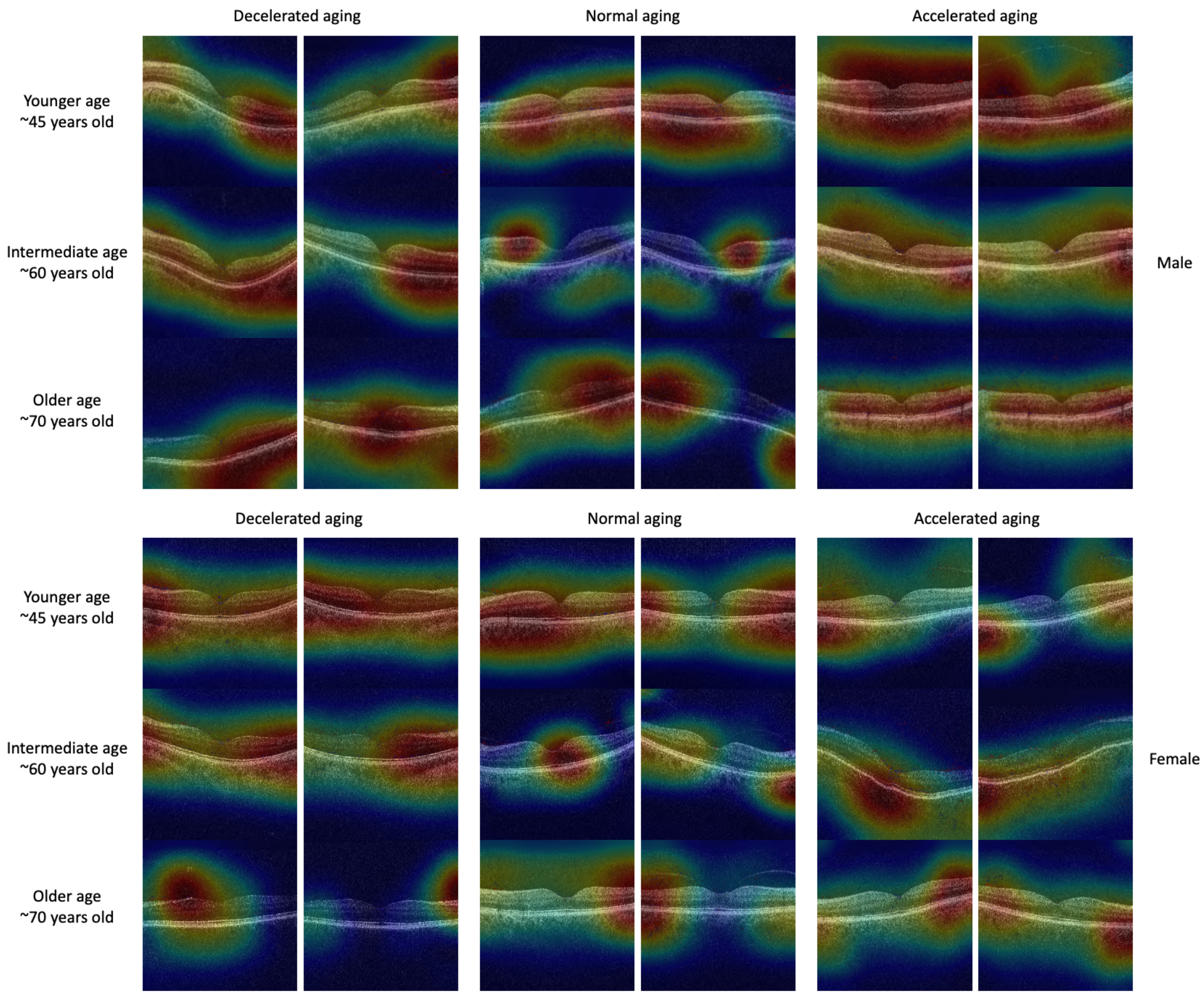

**Supplementary Figure 14: Attention maps for eye OCT images**

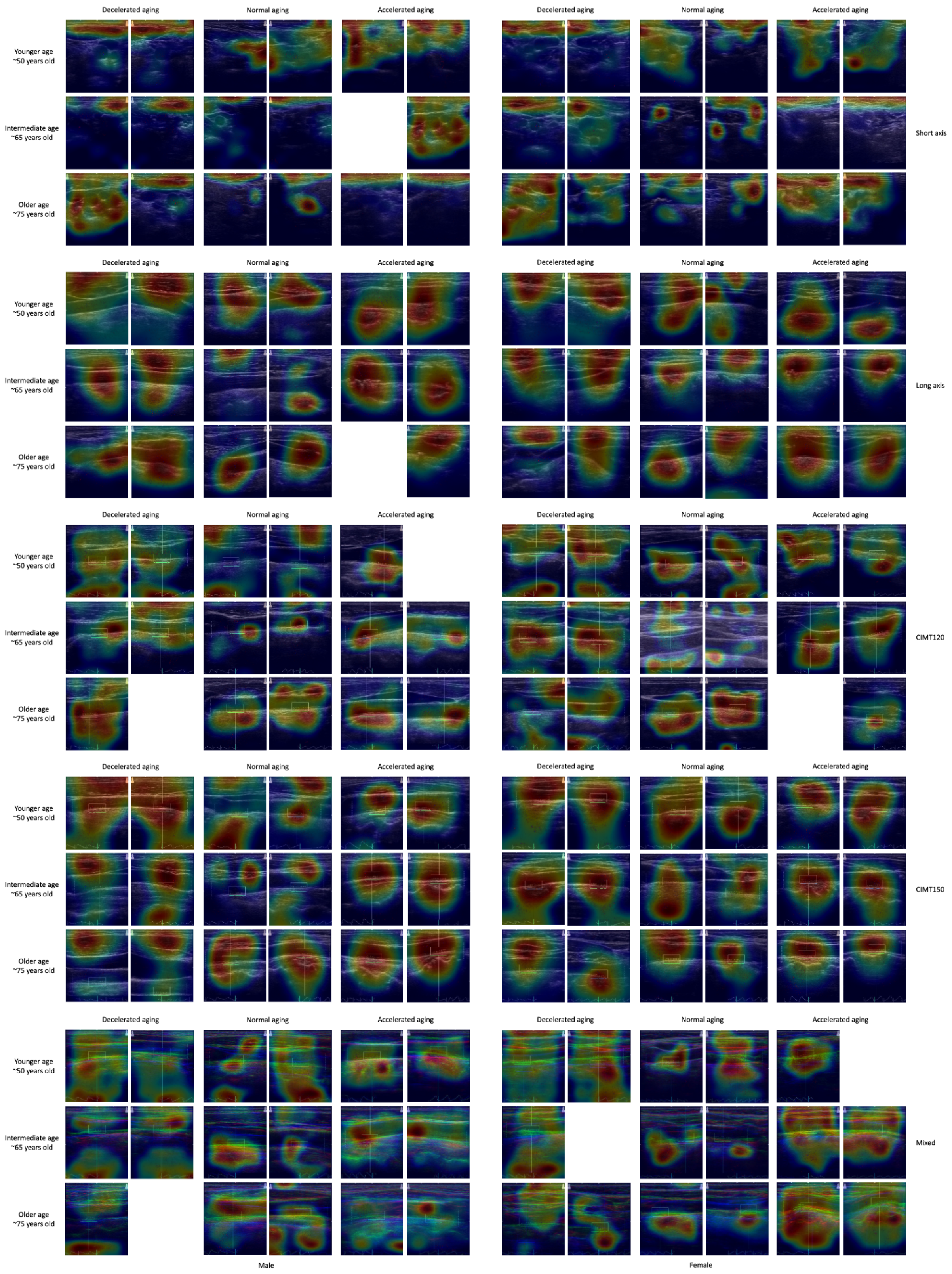

**Supplementary Figure 15: Attention maps for carotid ultrasound images**

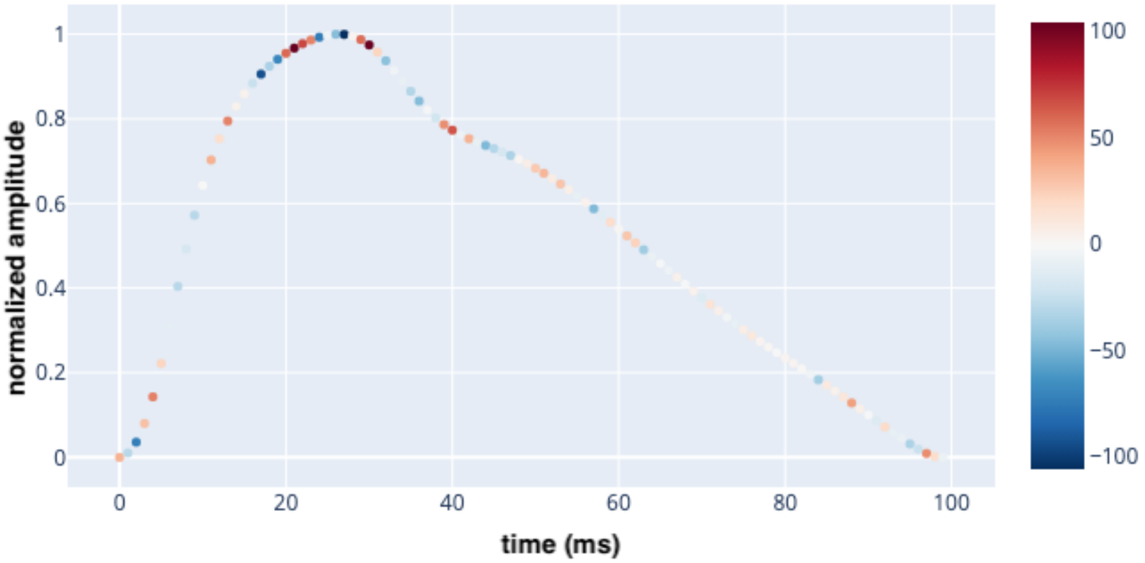

**Supplementary Figure 16: Attention map example for pulse wave analysis time series**

The participant is a correctly predicted 60-65-year-old male. Red data points represent time steps for which a higher value would increase the chronological age prediction, and blue data points represent time steps for which a higher value would decrease the chronological age prediction.

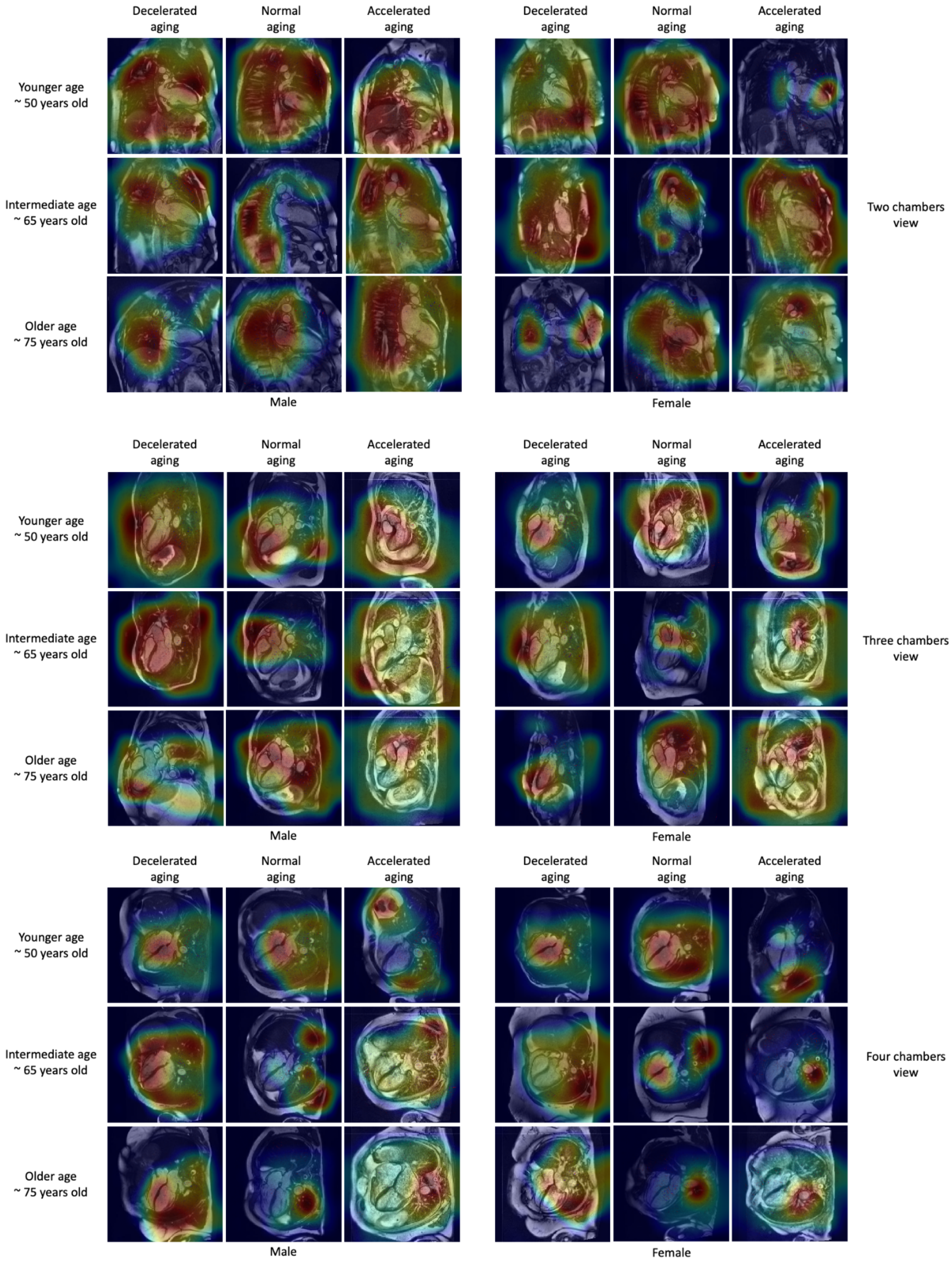

**Supplementary Figure 17: Attention maps for heart MRI images (raw images)**

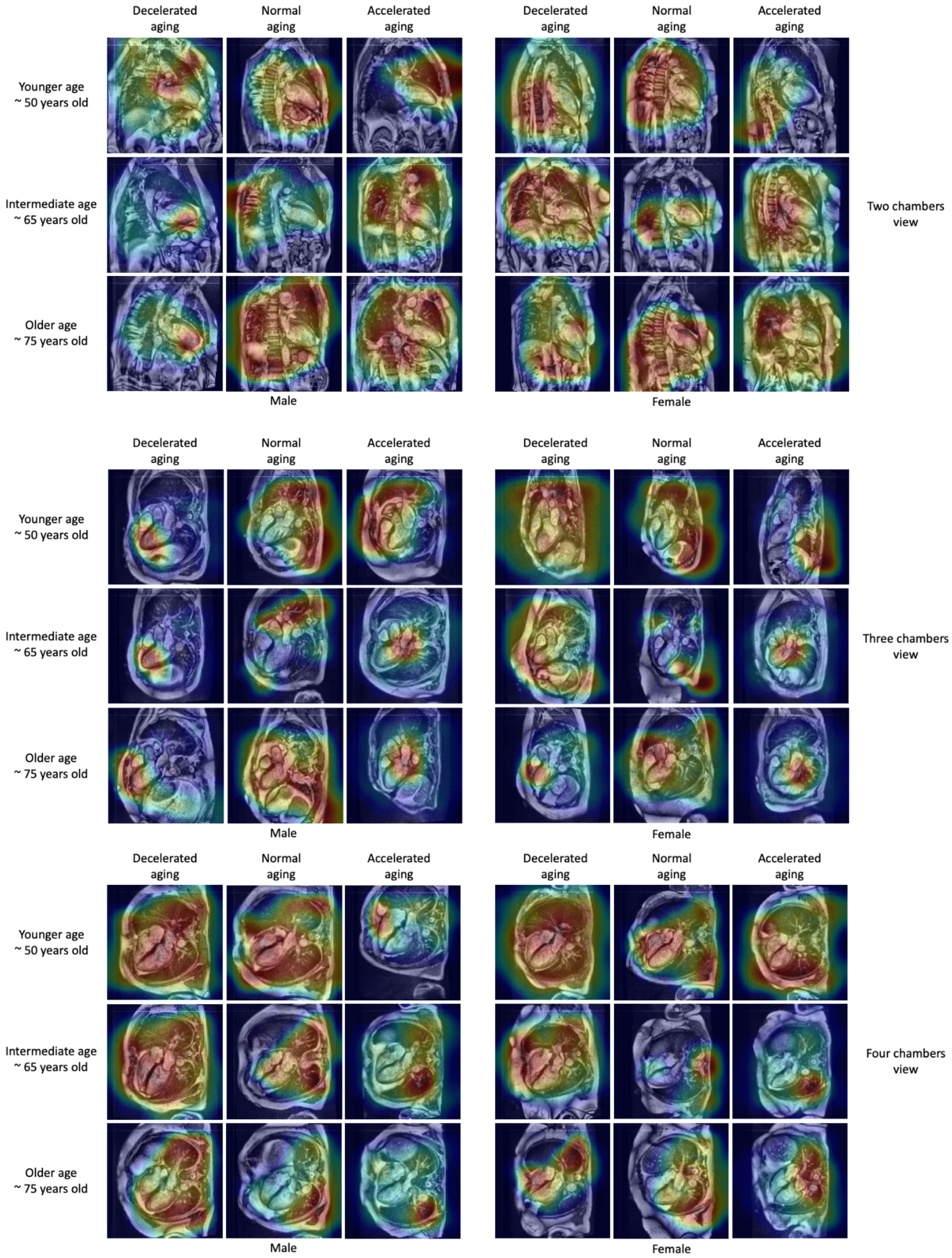

**Supplementary Figure 18: Attention maps for heart MRI images (contrasted images)**

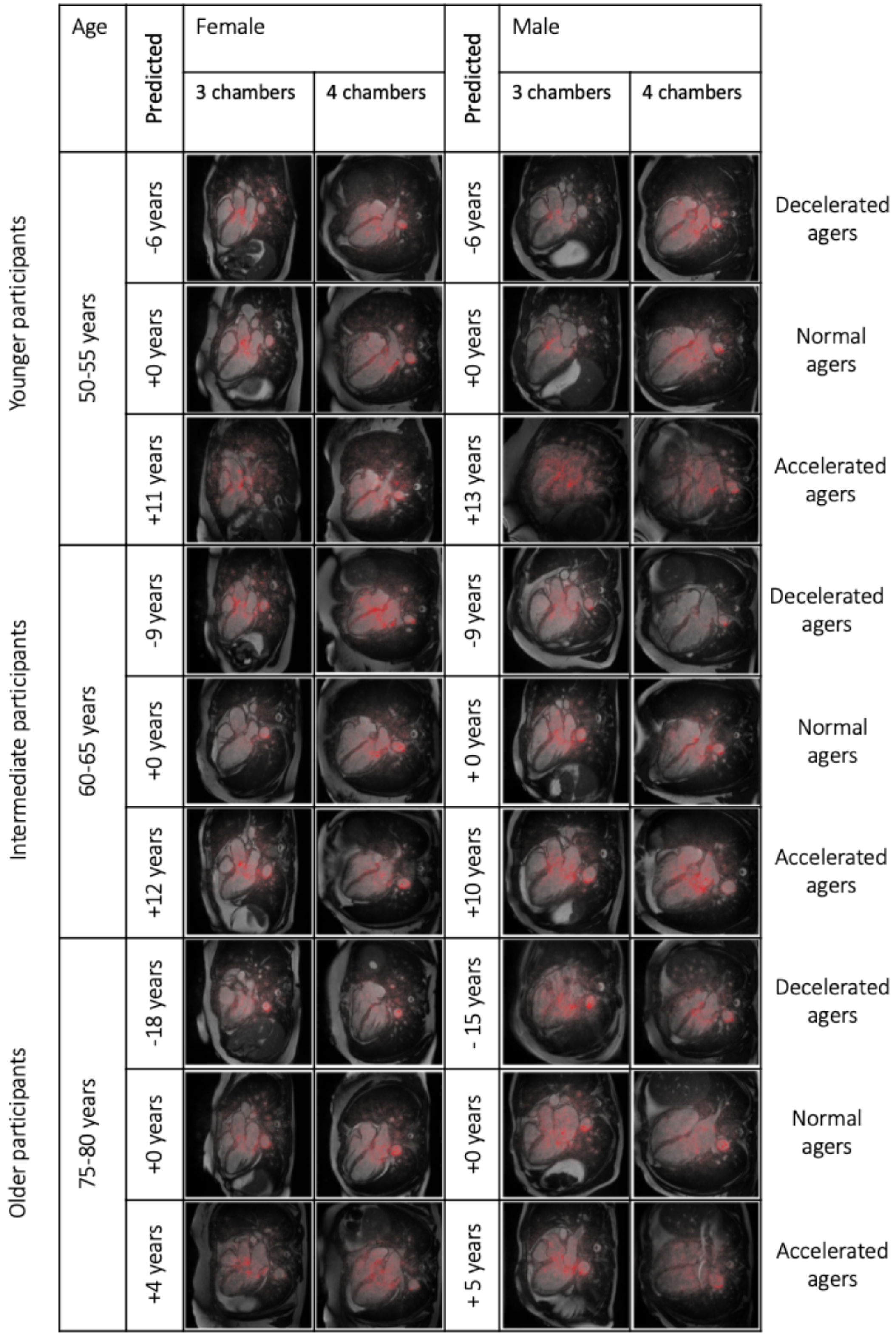

**Supplementary Figure 19: Attention maps for heart MRI videos - summary images**

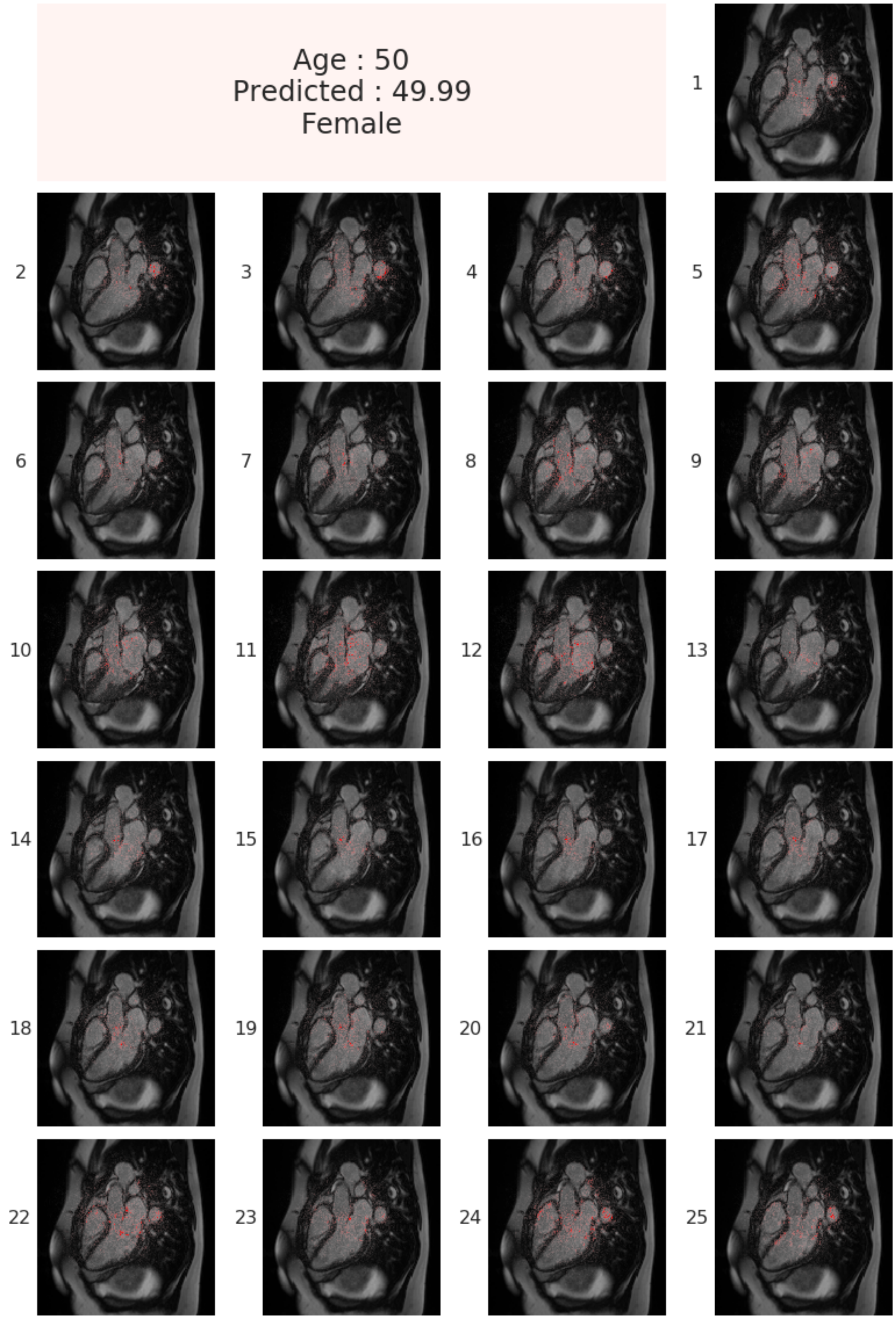

**Supplementary Figure 20: Attention maps for heart MRI videos - time frames XXTODO**

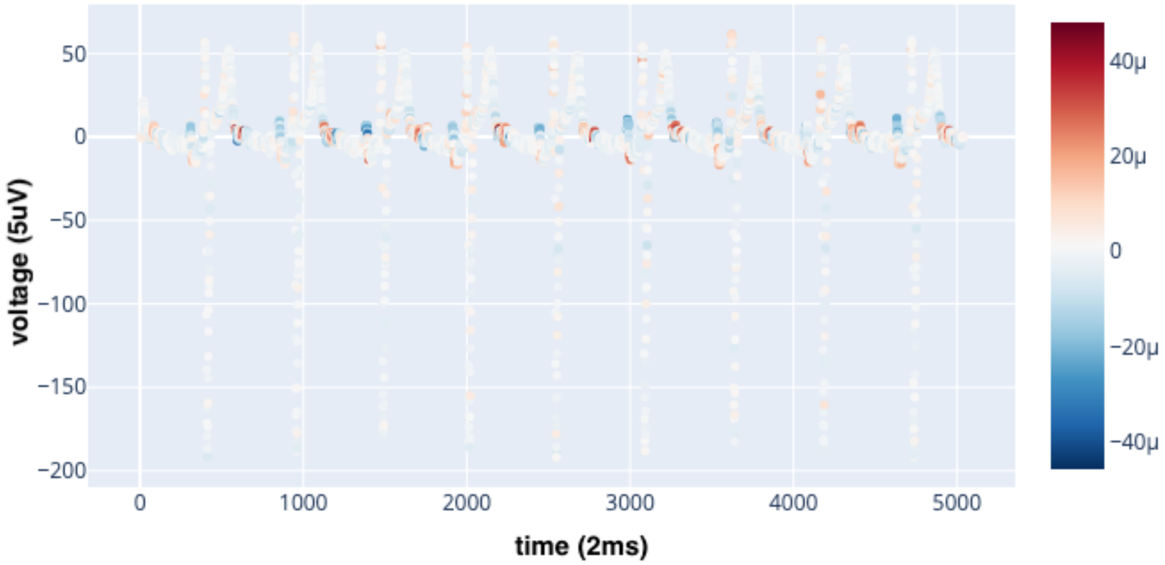

**Supplementary Figure 21: Attention map example for ECG time series**

The participant is a correctly predicted 65-70-year-old male. The signal displayed is the first lead of the ECG. Red data points represent time steps for which a higher value would increase the chronological age prediction, and blue data points represent time steps for which a higher value would decrease the chronological age prediction.

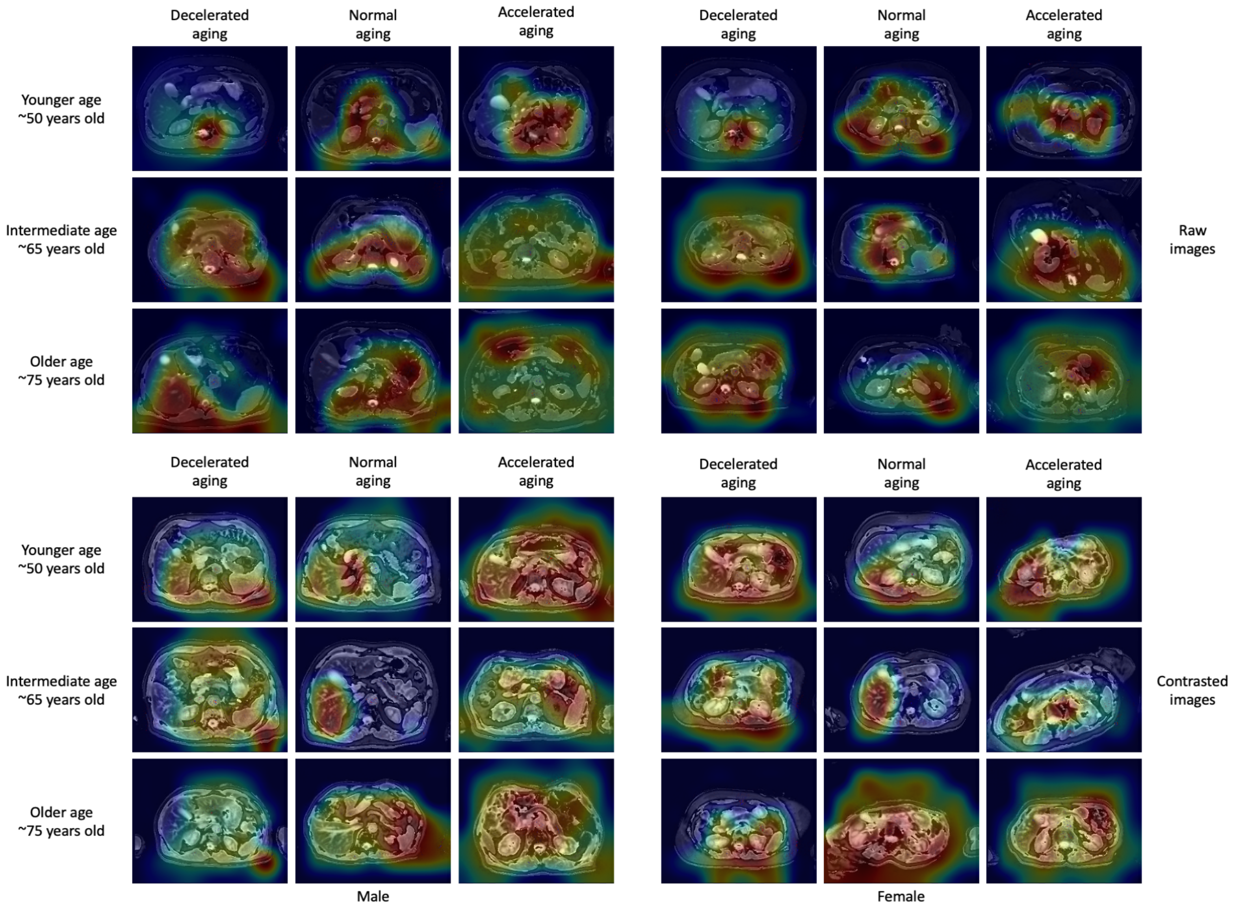

**Supplementary Figure 22: Attention maps for liver MRI images**

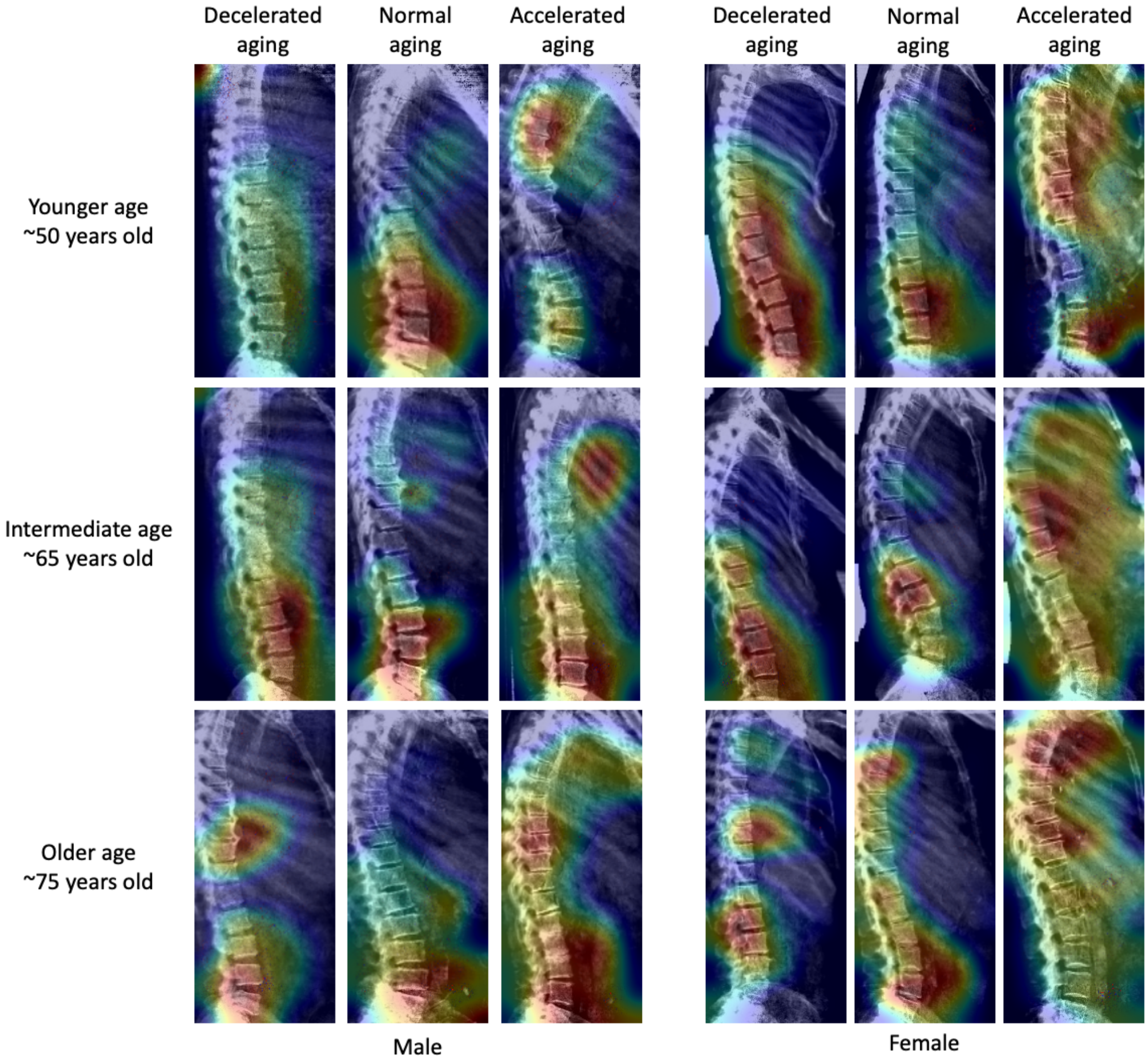

**Supplementary Figure 23: Attention maps for pancreas MRI images**

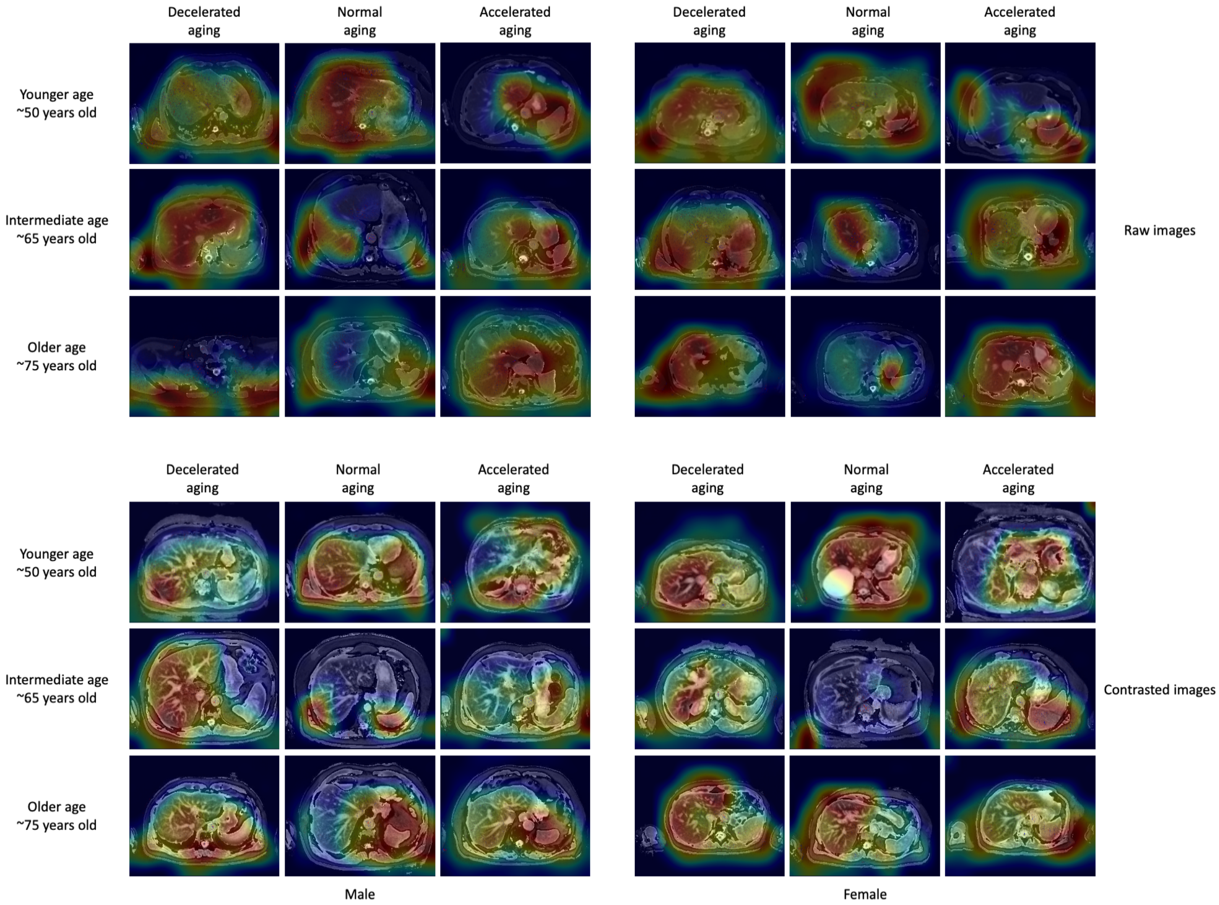

#### Supplementary Figure 24: Attention maps for spine X-ray images (sagittal view)

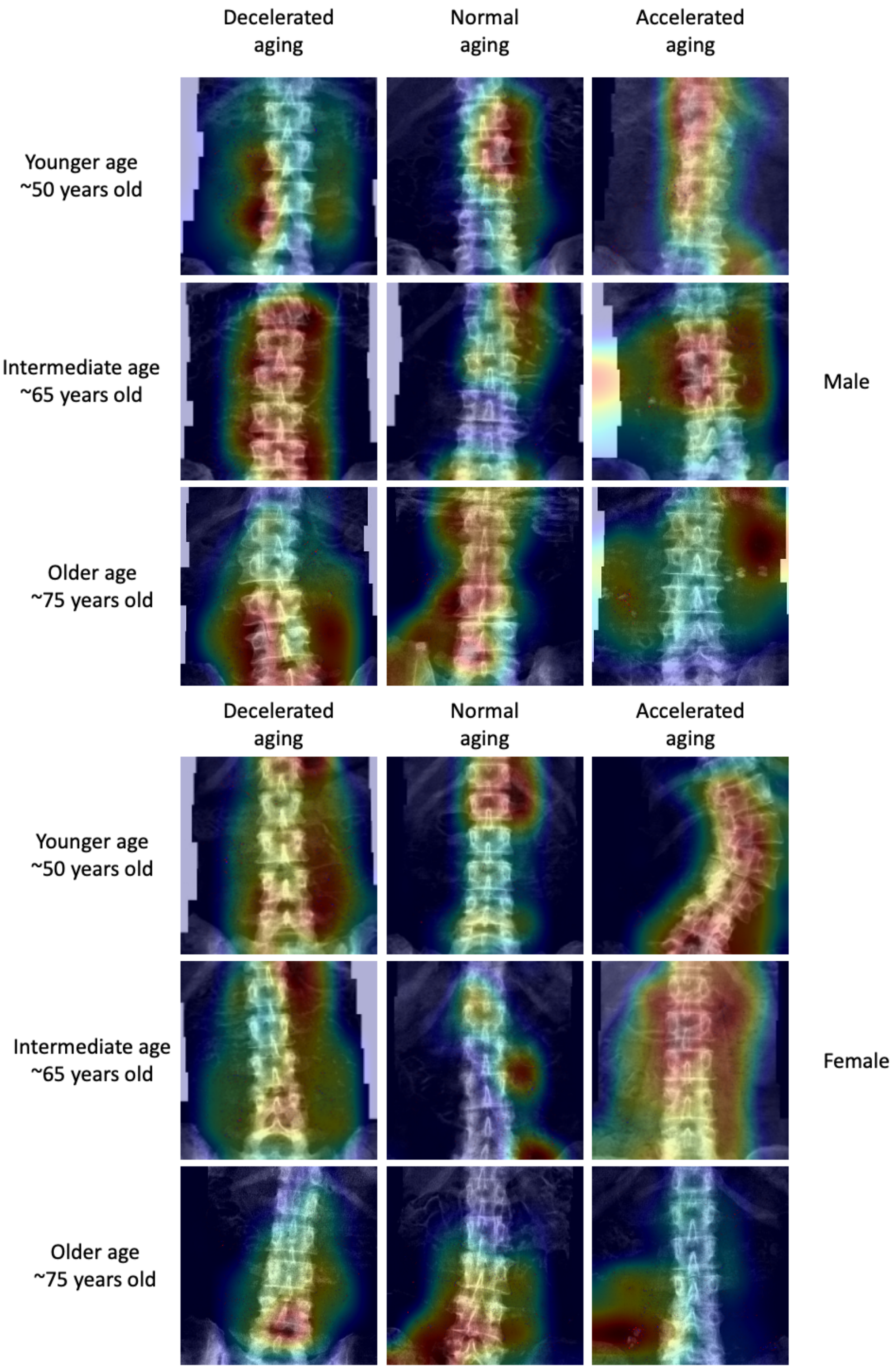

**Supplementary Figure 25: Attention maps for spine X-ray images (coronal view)**

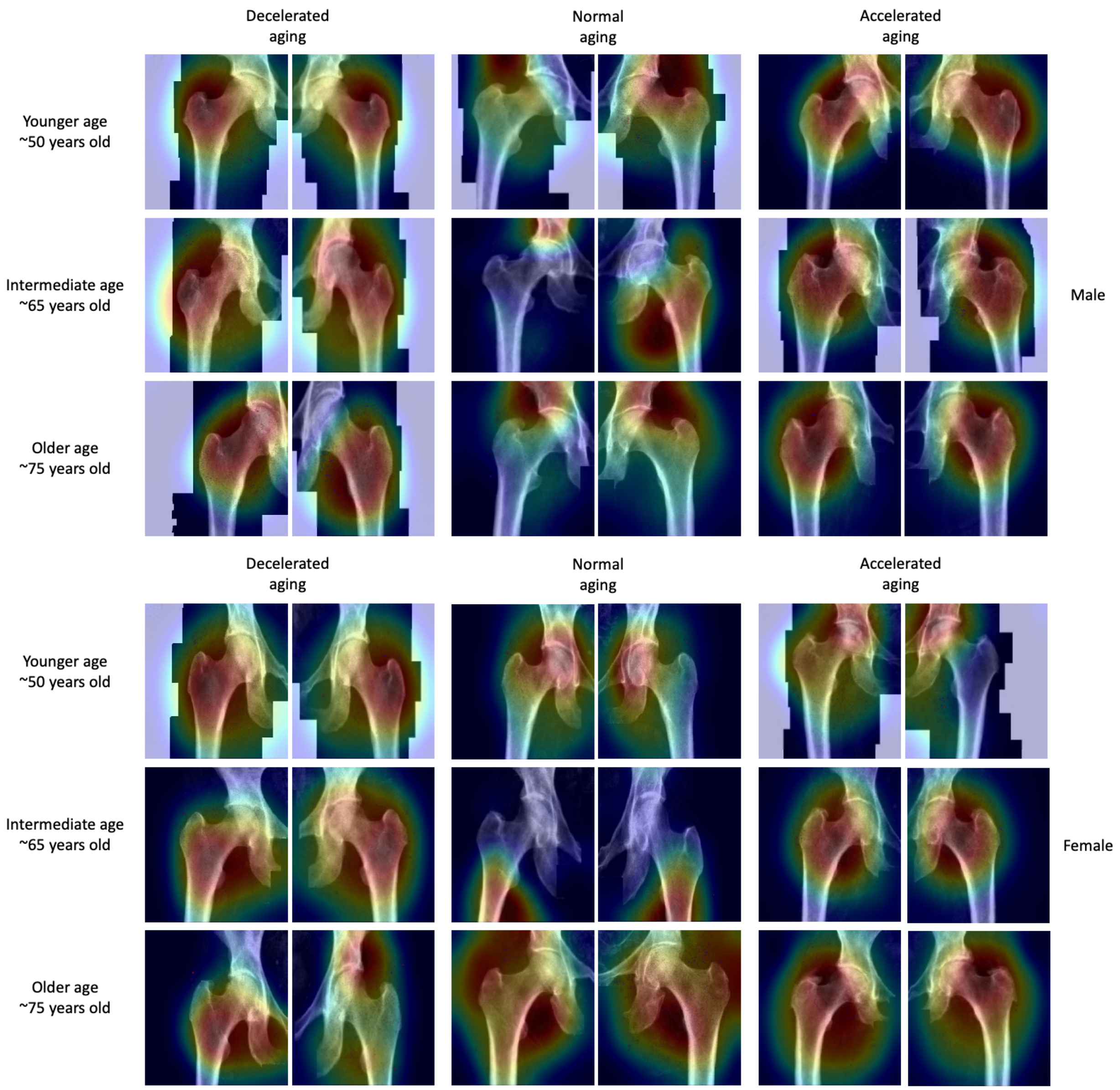

**Supplementary Figure 26: Attention maps for hip X-ray images**

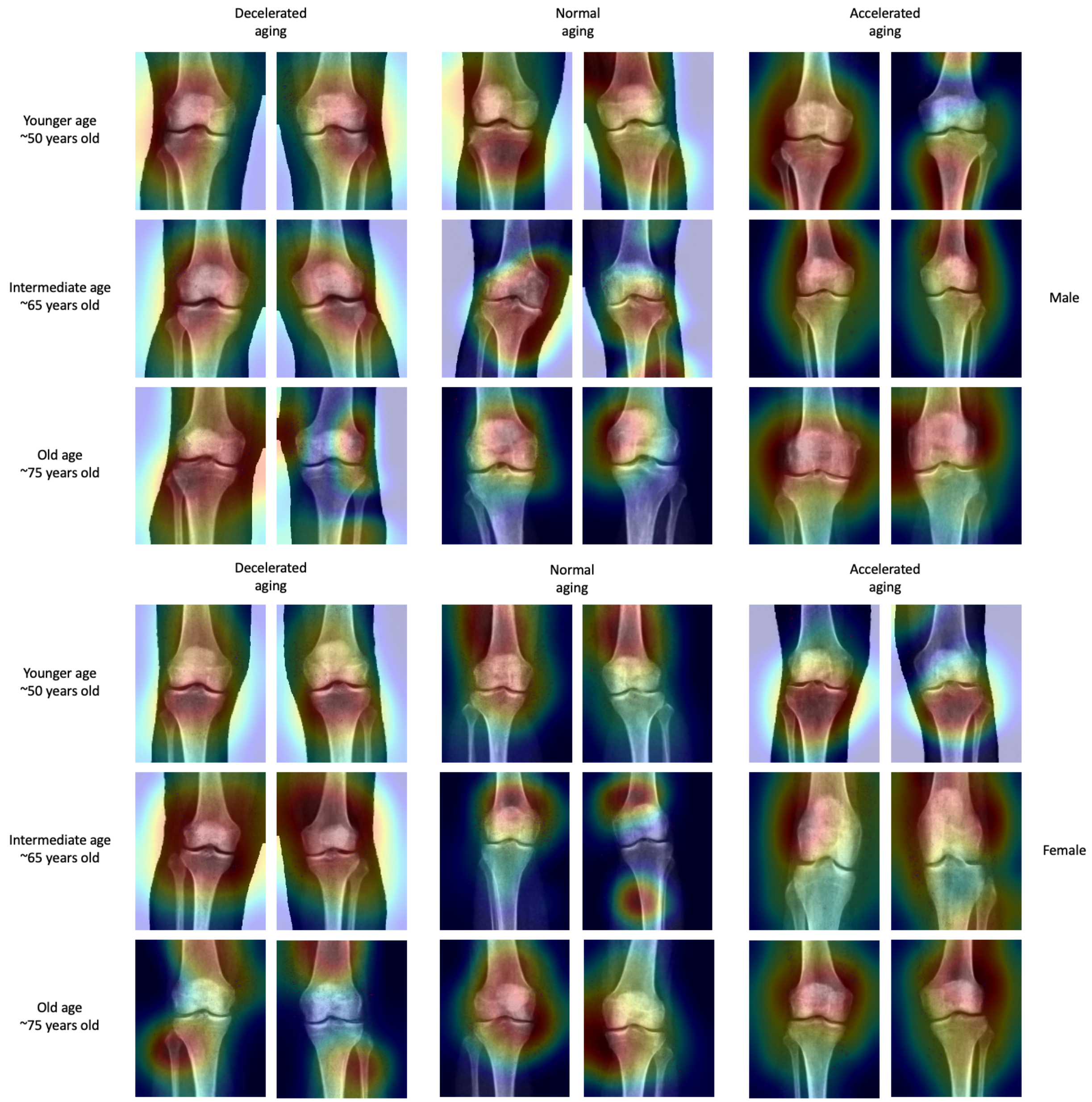

**Supplementary Figure 27: Attention maps for knee X-ray images**

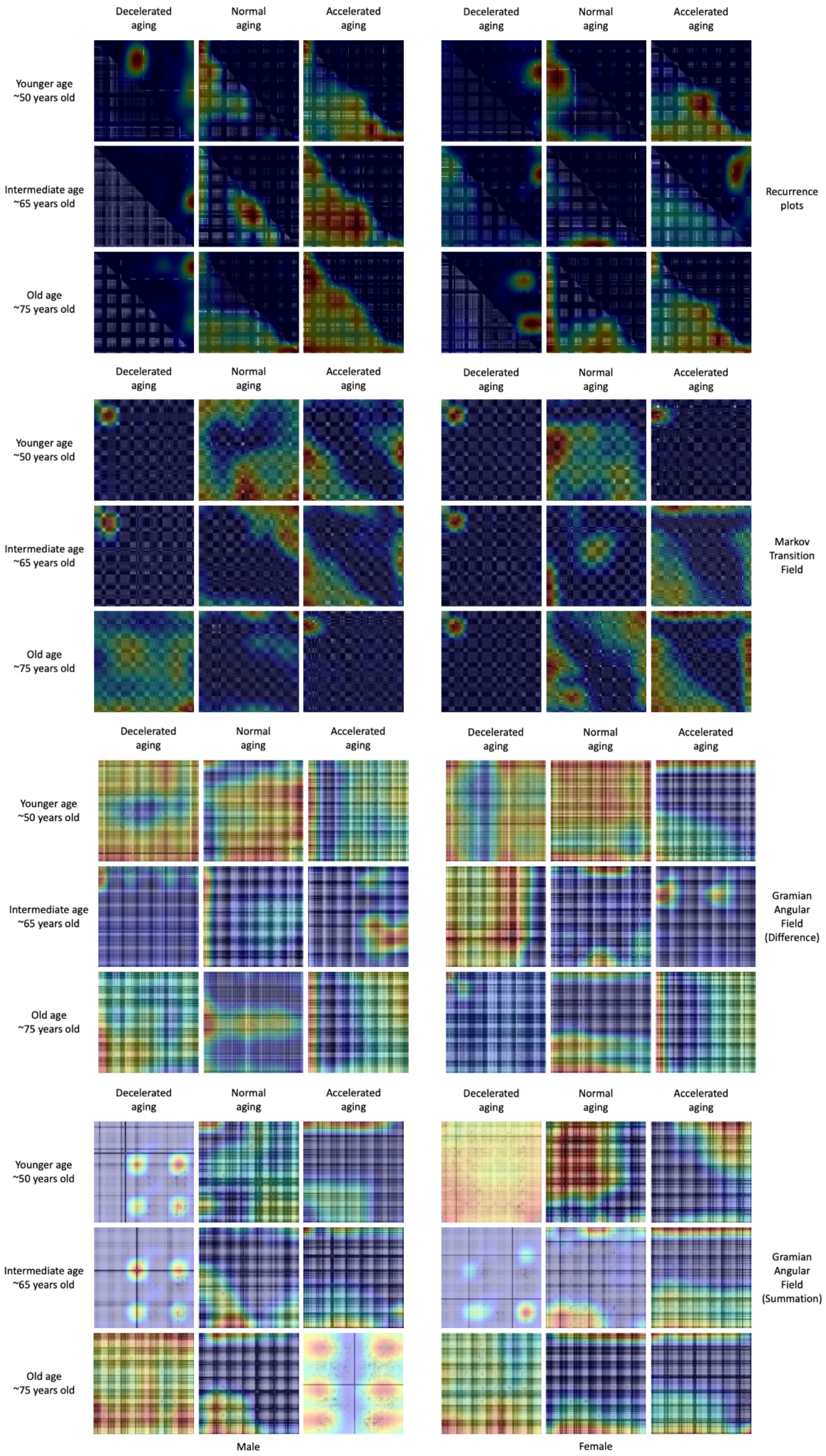

**Supplementary Figure 28: Attention maps for physical activity images**

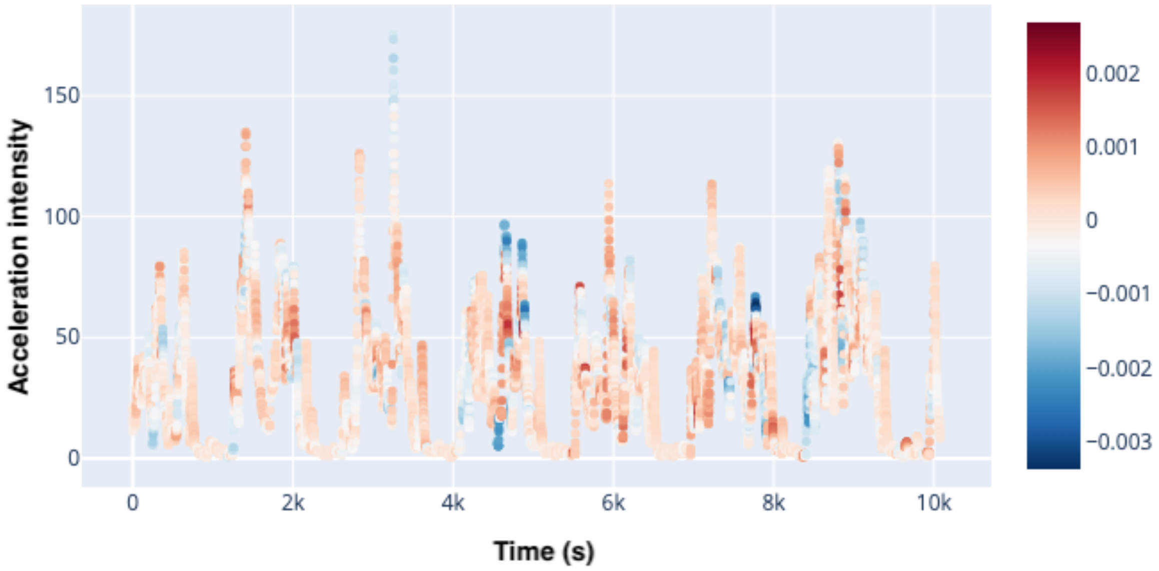

**Supplementary Figure 29: Attention map example for physical activity time series - Full week**

The participant is a correctly predicted 60-65-year-old male. Red data points represent time steps for which a higher value would increase the chronological age prediction, and blue data points represent time steps for which a higher value would decrease the chronological age prediction.

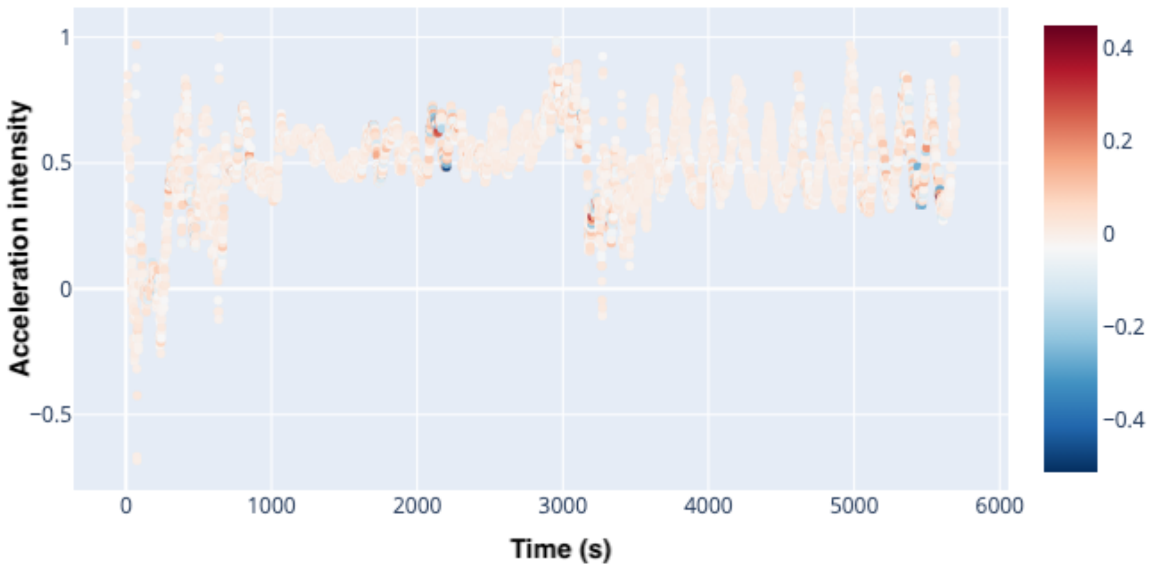

**Supplementary Figure 30: Attention map example for physical activity time series - Walking**

The participant is a correctly predicted 60-65-year-old male. The signal displayed is the accelerometer's z-acceleration. Red data points represent time steps for which a higher value would increase the chronological age prediction, and blue data points represent time steps for which a higher value would decrease the chronological age prediction.

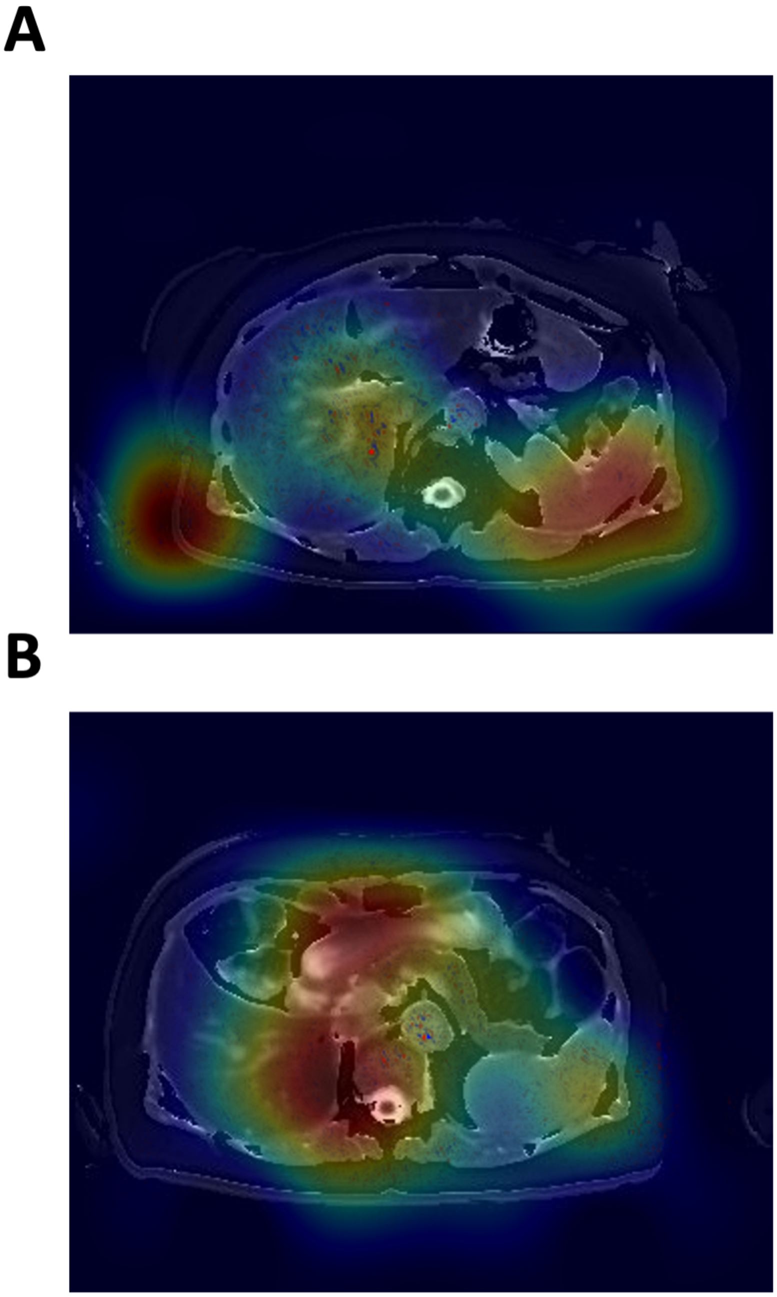

**Supplementary Figure 31: Attention map examples for a liver MRI-based age prediction (A) and a pancreas MRI-based age prediction (B)**

These two abdomen MRI-based age predictors do not particularly rely on the anatomical features of their respective organs to perform their prediction, which explains their correlation.

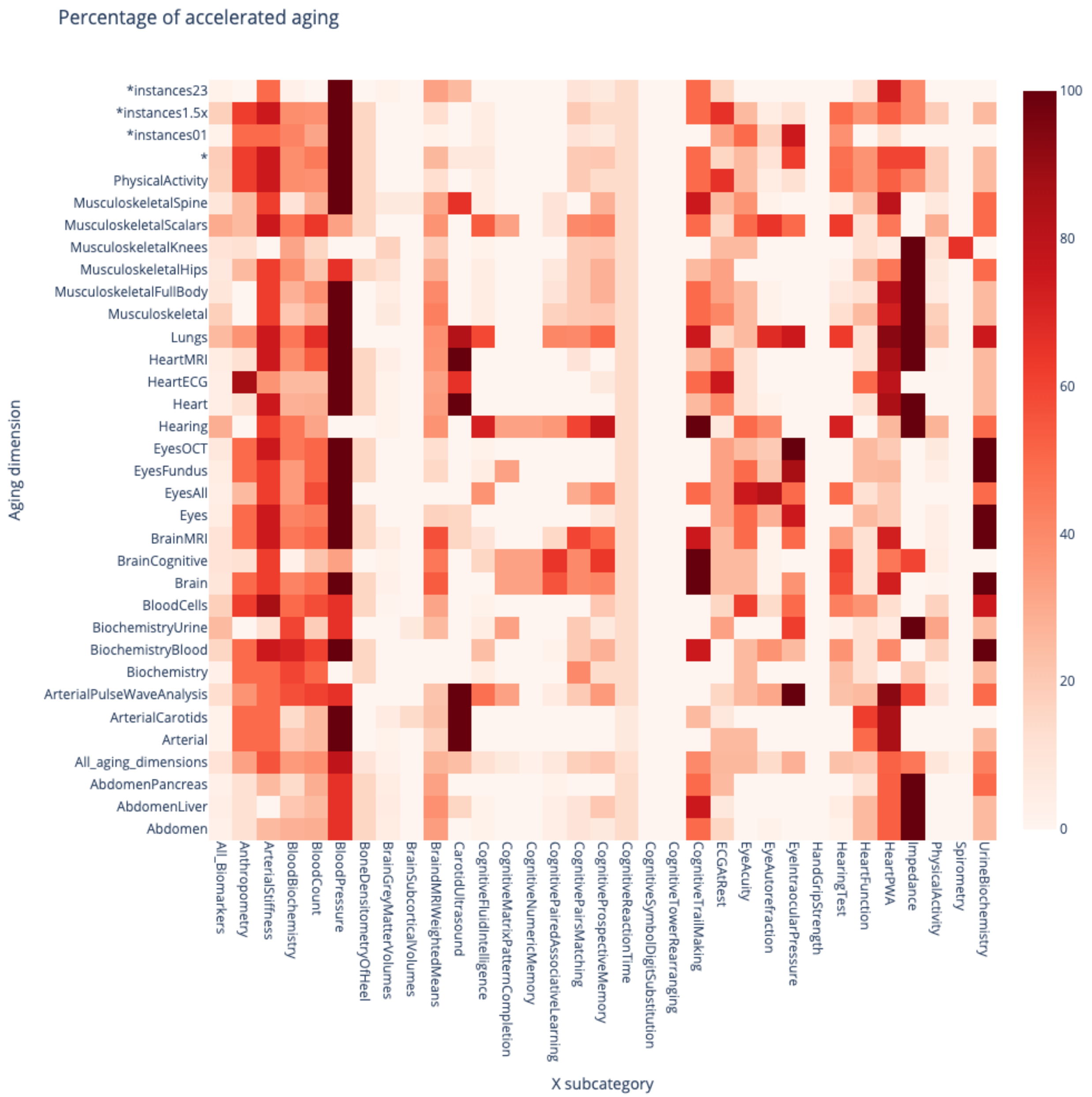

**Supplementary Figure 32**: **Biomarkers Wide Association Study - Proportion of biomarkers significantly associated with accelerated aging in each aging dimension, for each biomarker category**

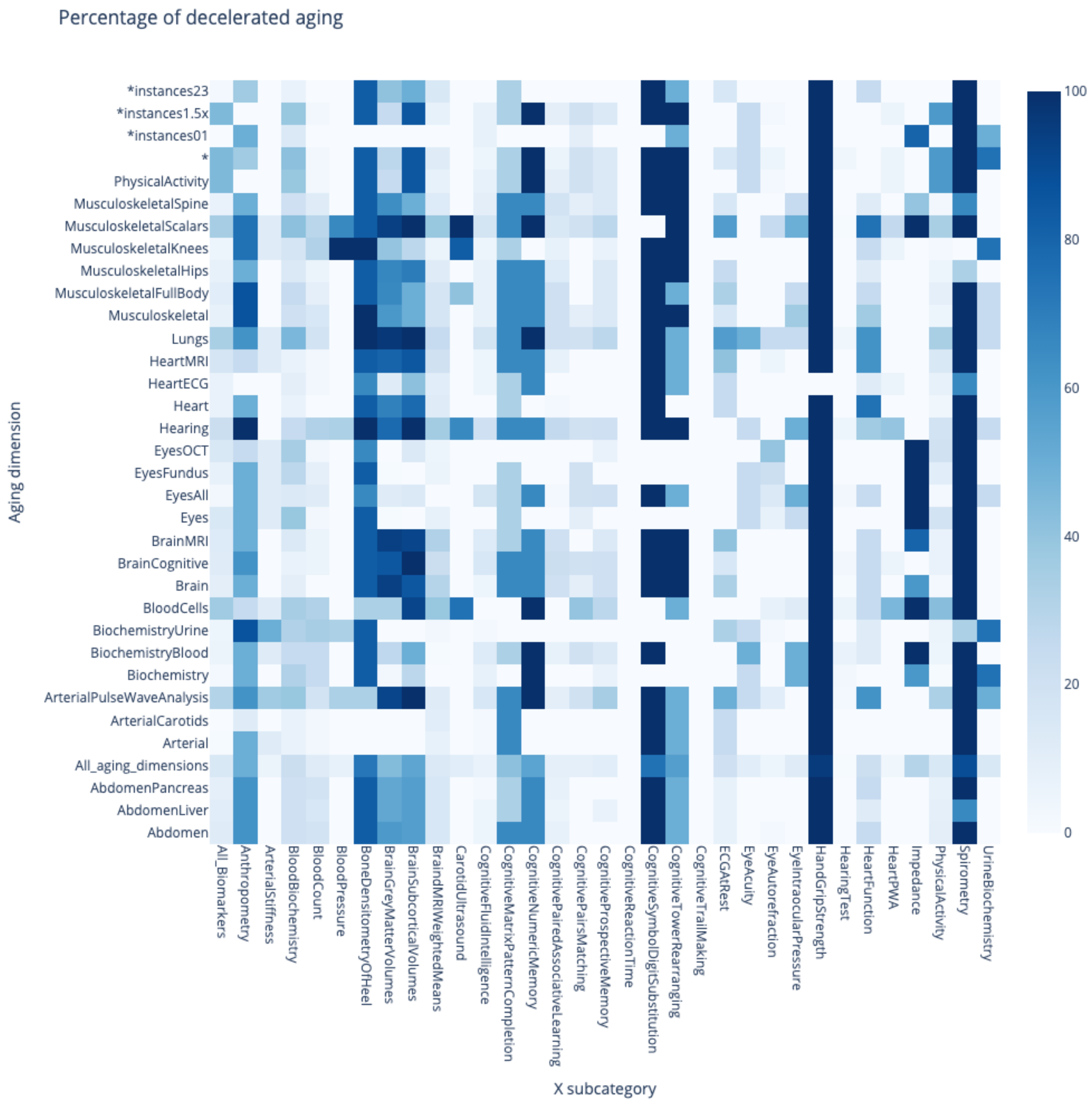

**Supplementary Figure 33**: **Biomarkers Wide Association Study - Proportion of biomarkers significantly associated with decelerated aging in each aging dimension, for each biomarker category**

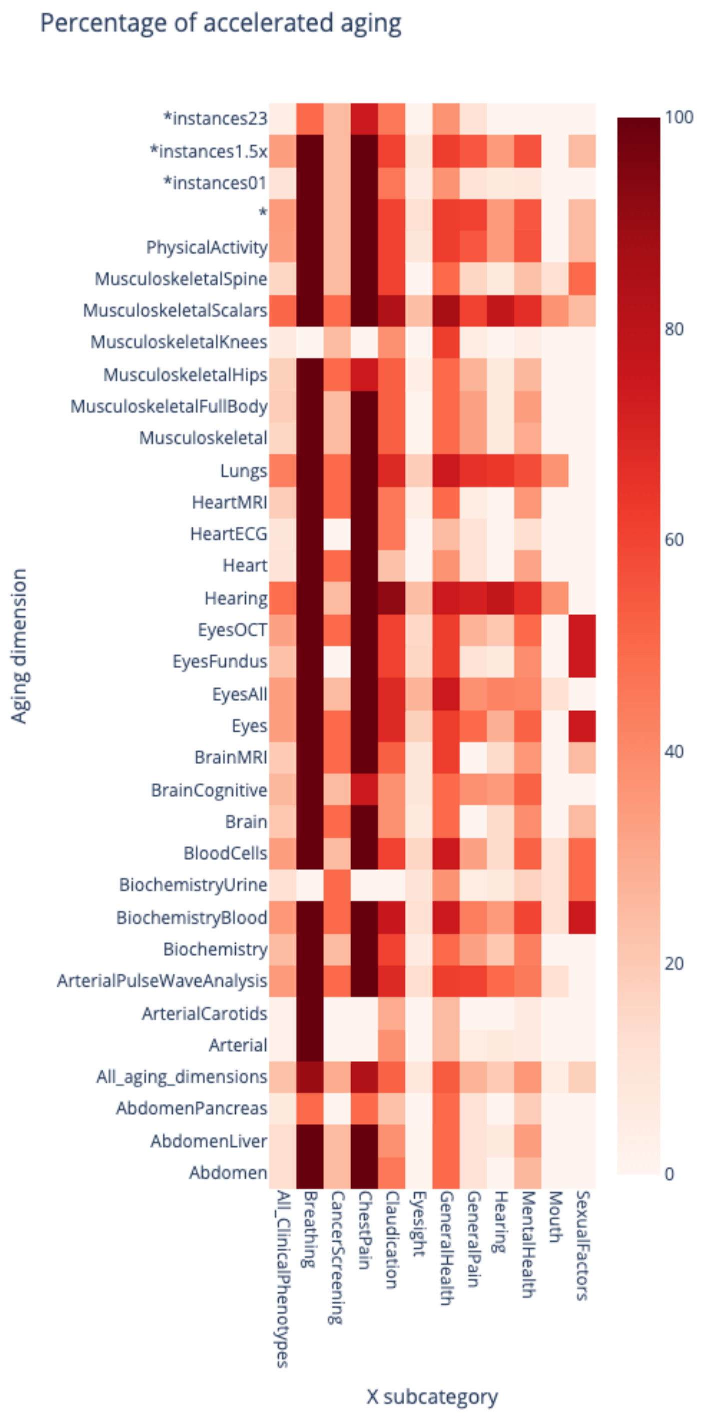

**Supplementary Figure 34**: **Clinical Phenotypes Wide Association Study - Proportion of clinical phenotypes significantly associated with accelerated aging in each aging dimension, for each clinical phenotypes category**

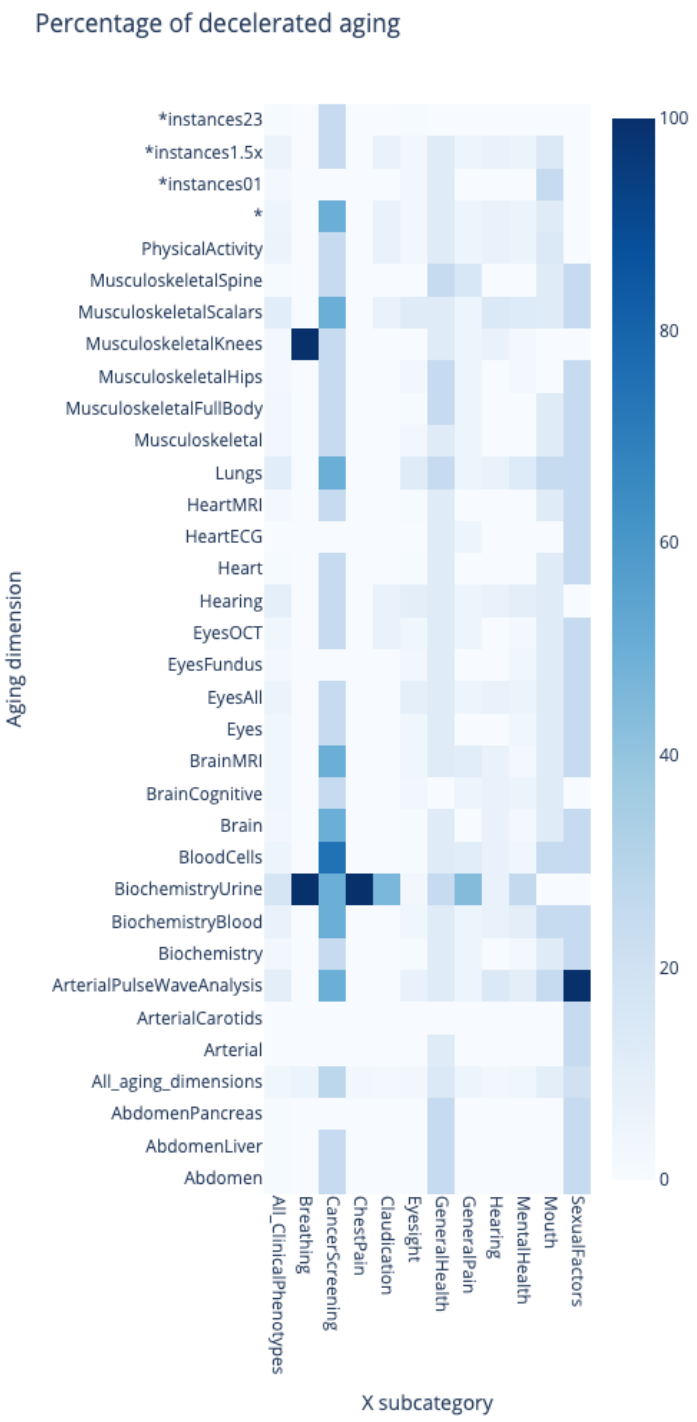

**Supplementary Figure 35**: **Clinical Phenotypes Wide Association Study - Proportion of clinical phenotypes significantly associated with decelerated aging in each aging dimension, for each clinical phenotypes category**

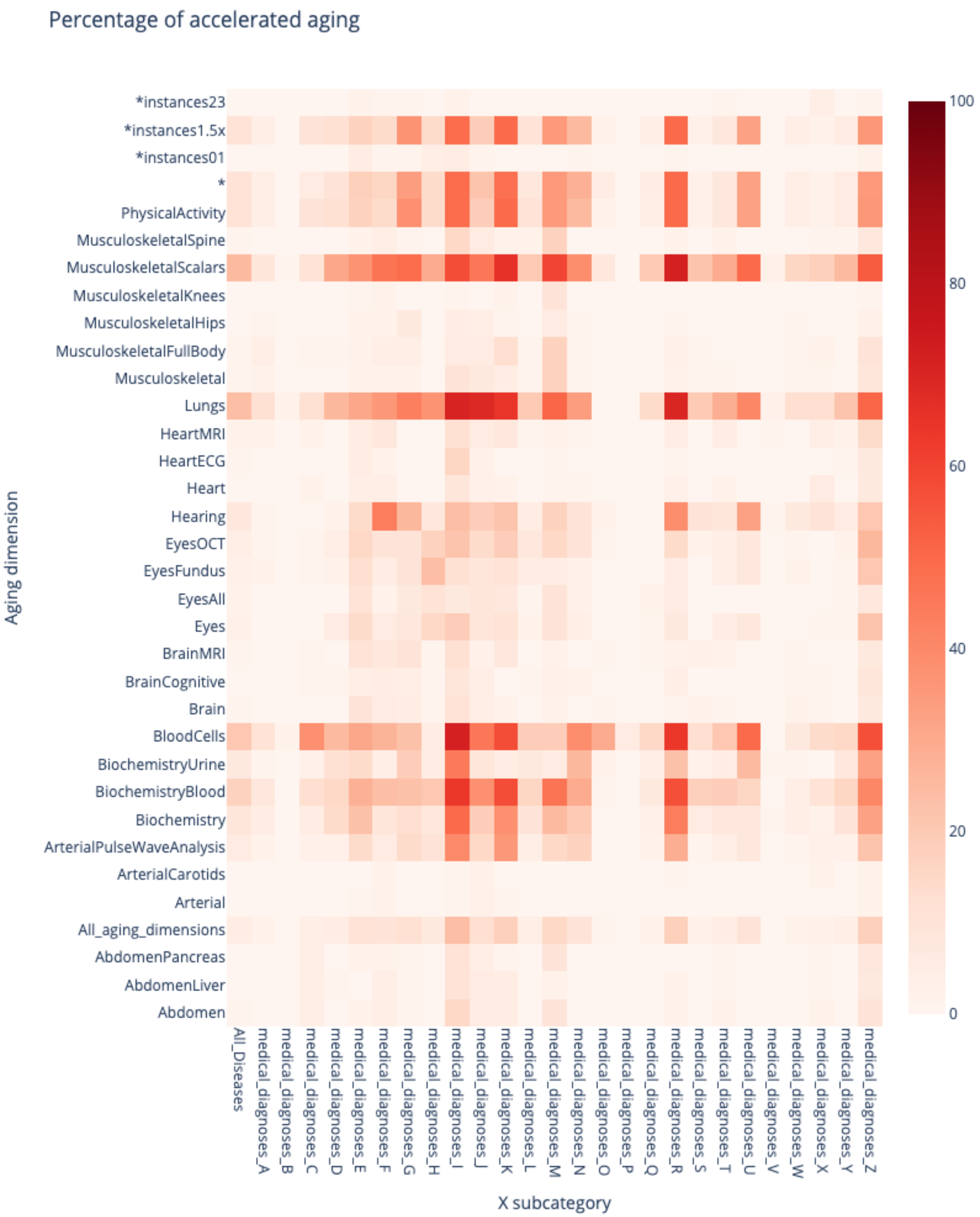

**Supplementary Figure 36: Diseases Wide Association Study - Proportion of diseases significantly associated with accelerated aging in each aging dimension, for each disease category**

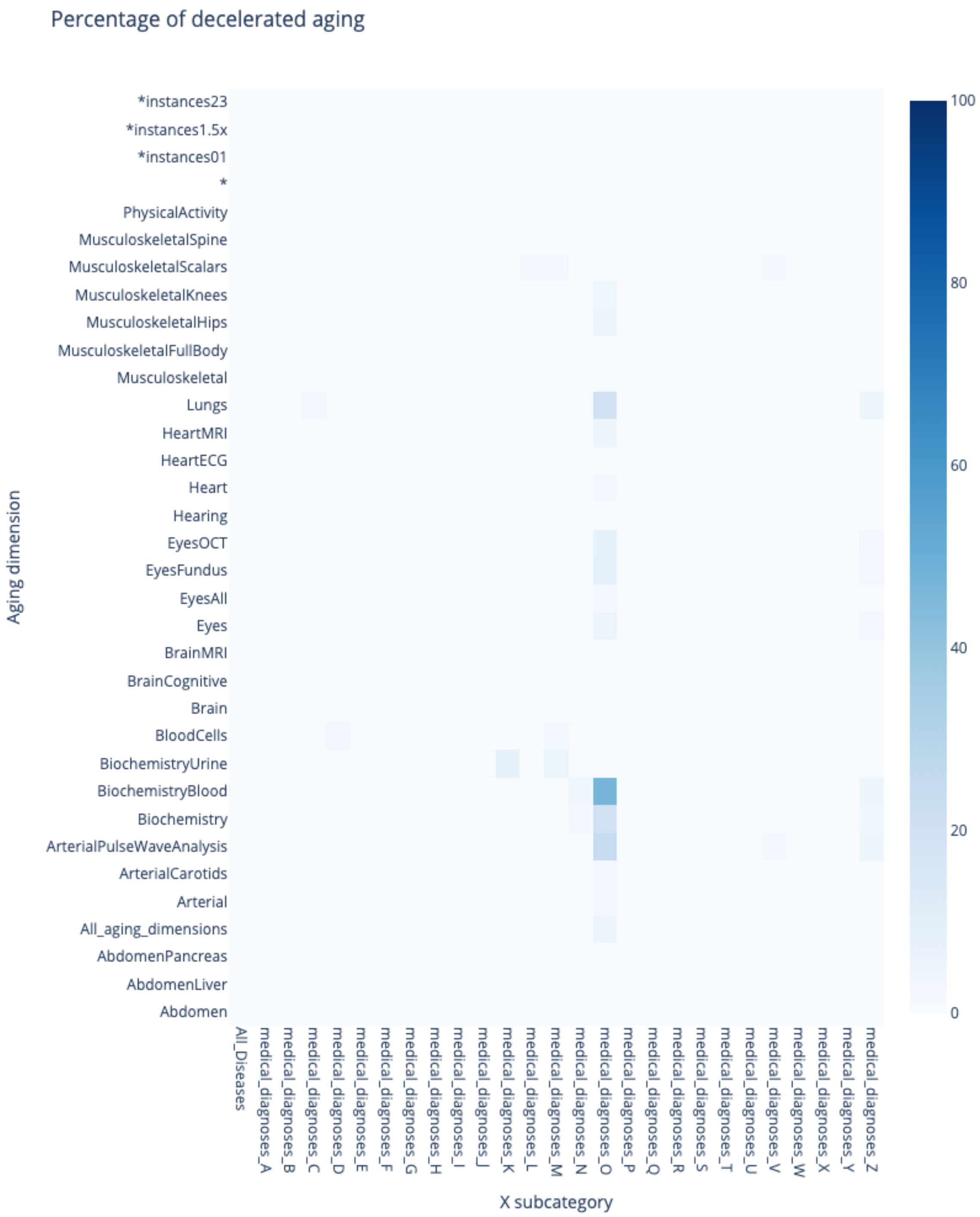

**Supplementary Figure 37**: **Diseases Wide Association Study - Proportion of diseases significantly associated with decelerated aging in each aging dimension, for each disease category**

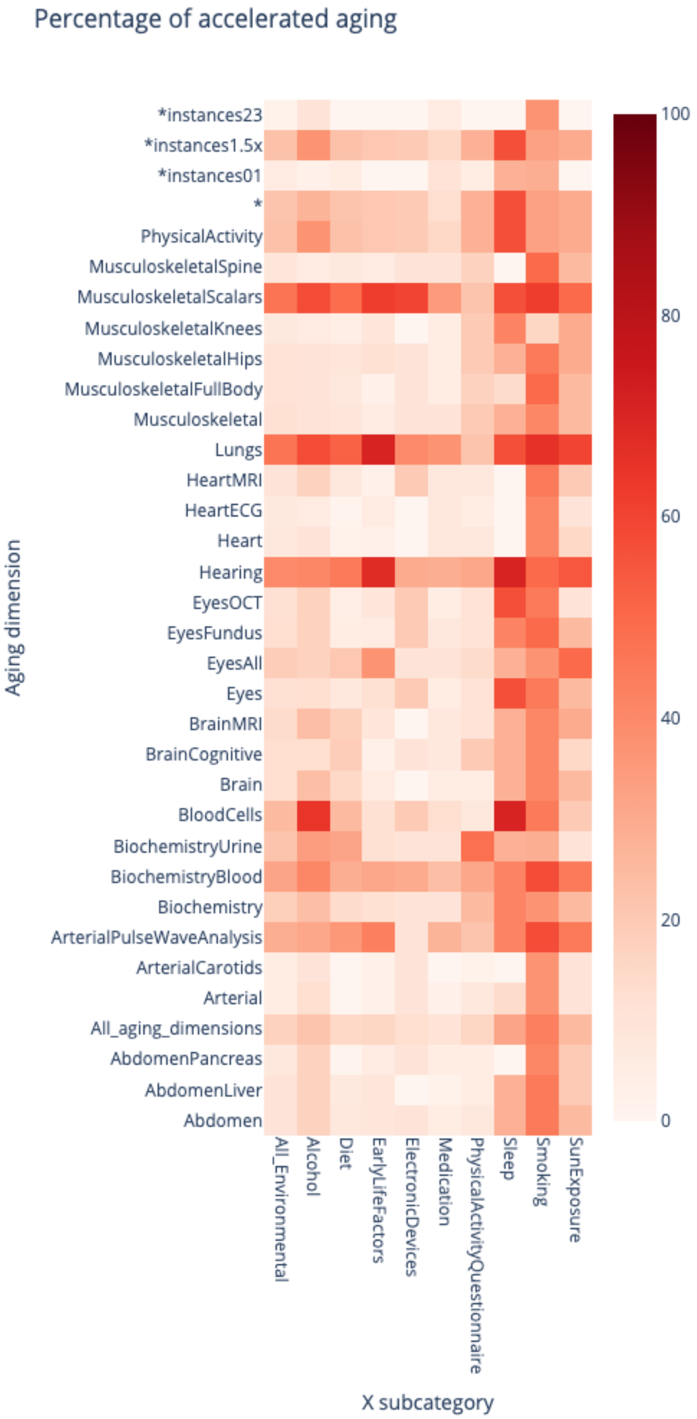

**Supplementary Figure 38**: **Environmental Wide Association Study - Proportion of environmental variables significantly associated with accelerated aging in each aging dimension, for each environmental category**

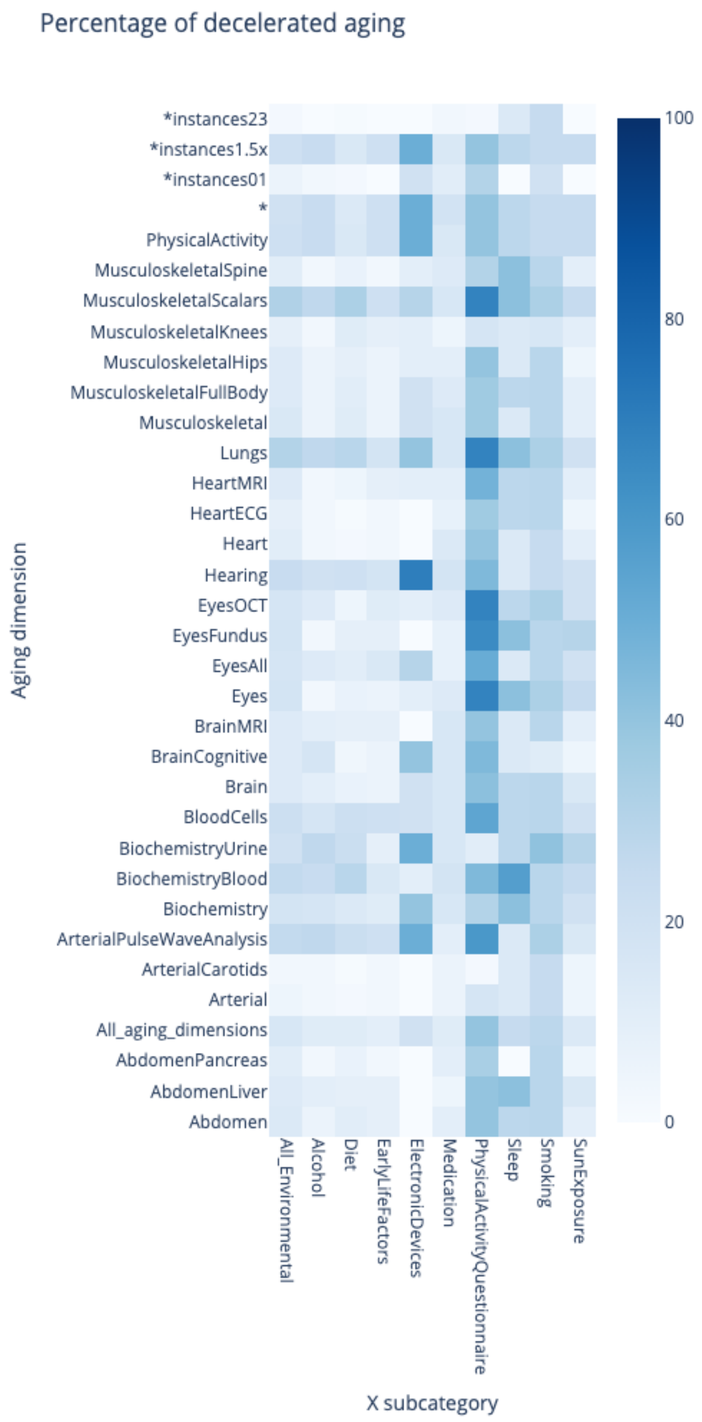

**Supplementary Figure 39**: **Environmental Wide Association Study - Proportion of environmental variables significantly associated with decelerated aging in each aging dimension, for each environmental category**

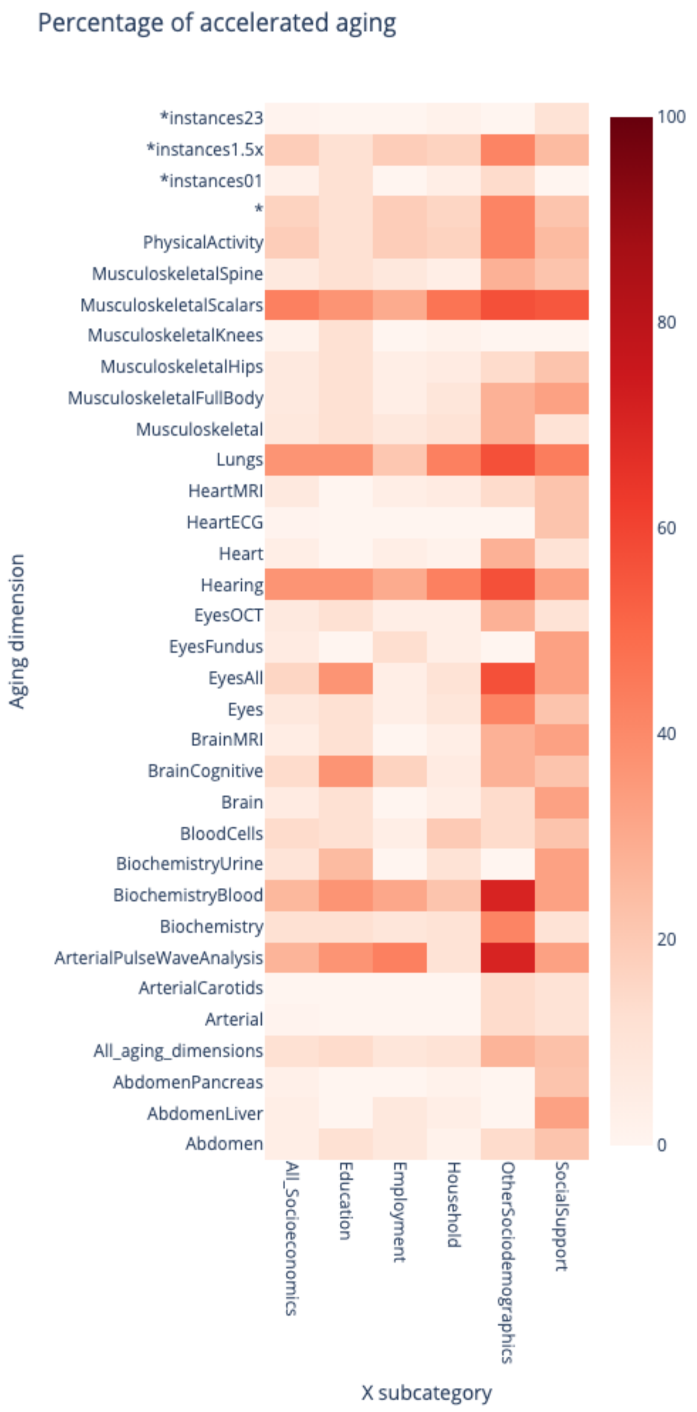

**Supplementary Figure 40**: **Socioeconomics Wide Association Study - Proportion of socioeconomic variables significantly associated with accelerated aging in each aging dimension, for each socioeconomics category**

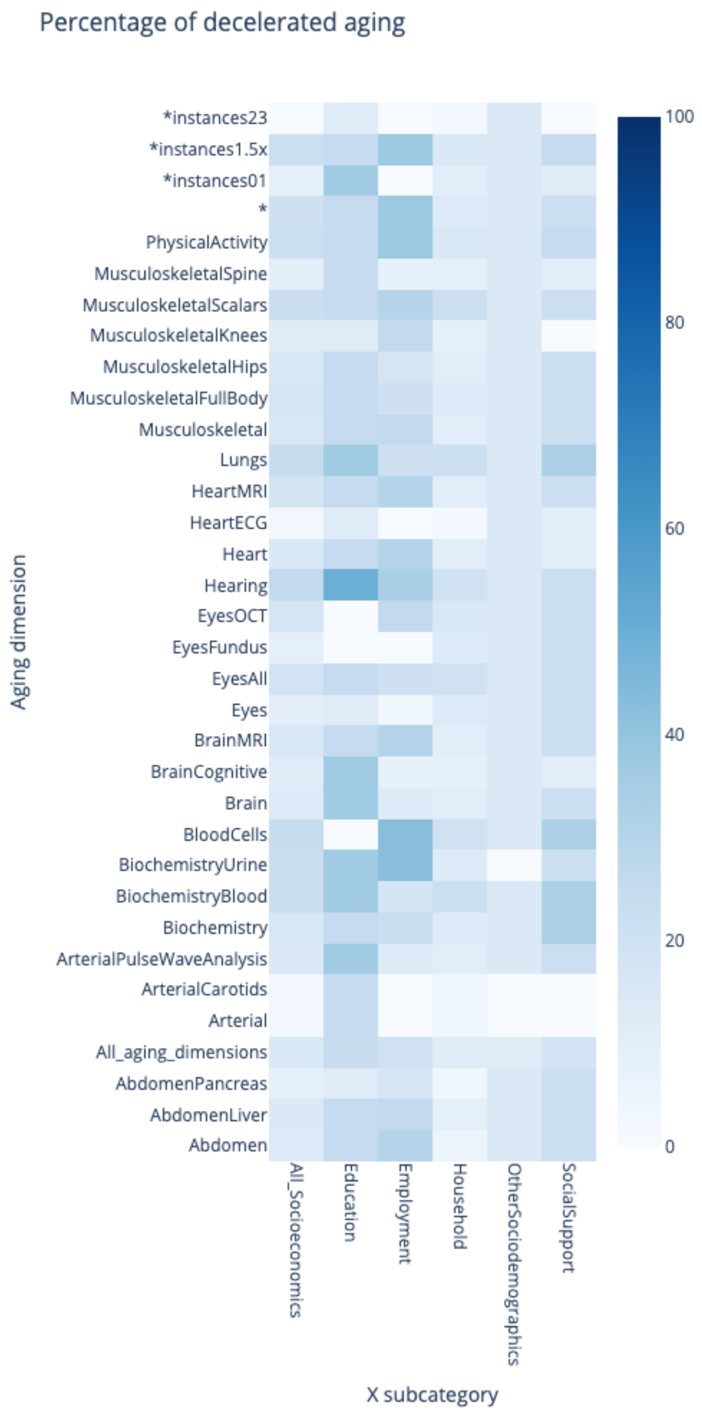

**Supplementary Figure 41**: **Socioeconomics Wide Association Study - Proportion of socioeconomic variables significantly associated with decelerated aging in each aging dimension, for each socioeconomics category**

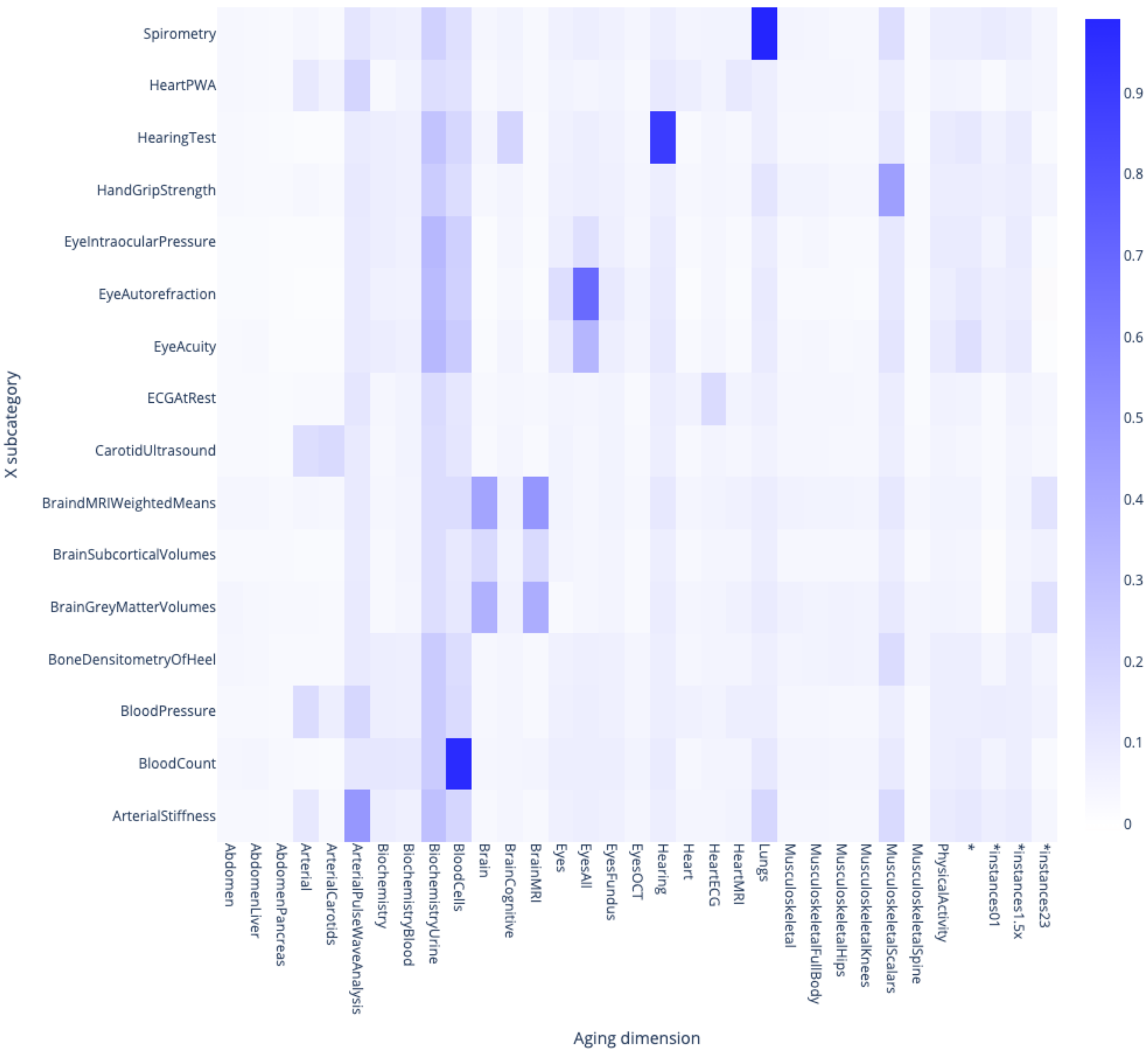

**Supplementary Figure 42: Prediction accuracy for predicting accelerated aging in each aging dimension, from each biomarkers subcategory**

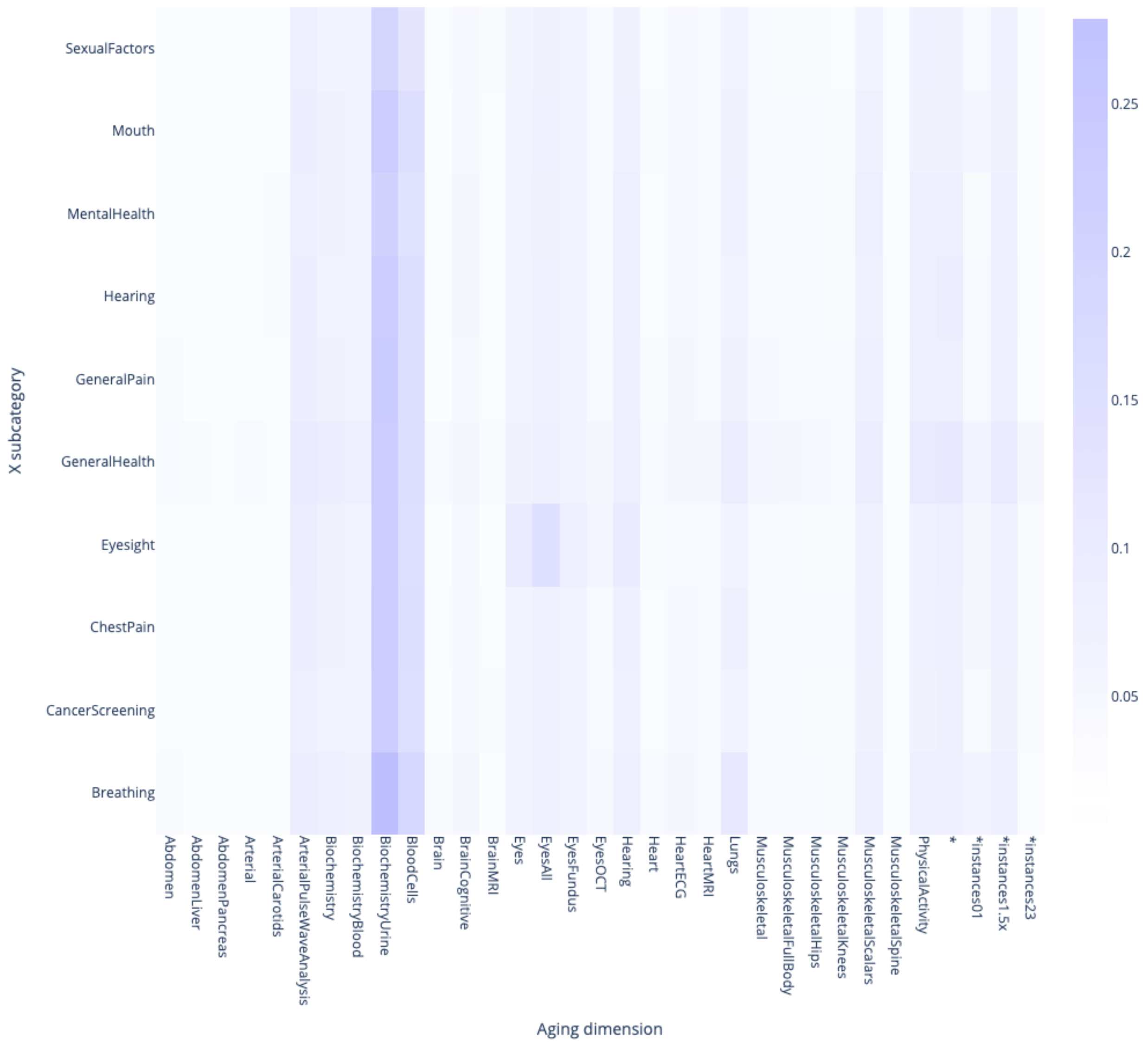

**Supplementary Figure 43: Prediction accuracy for predicting accelerated aging in each aging dimension, from each clinical phenotype subcategory**

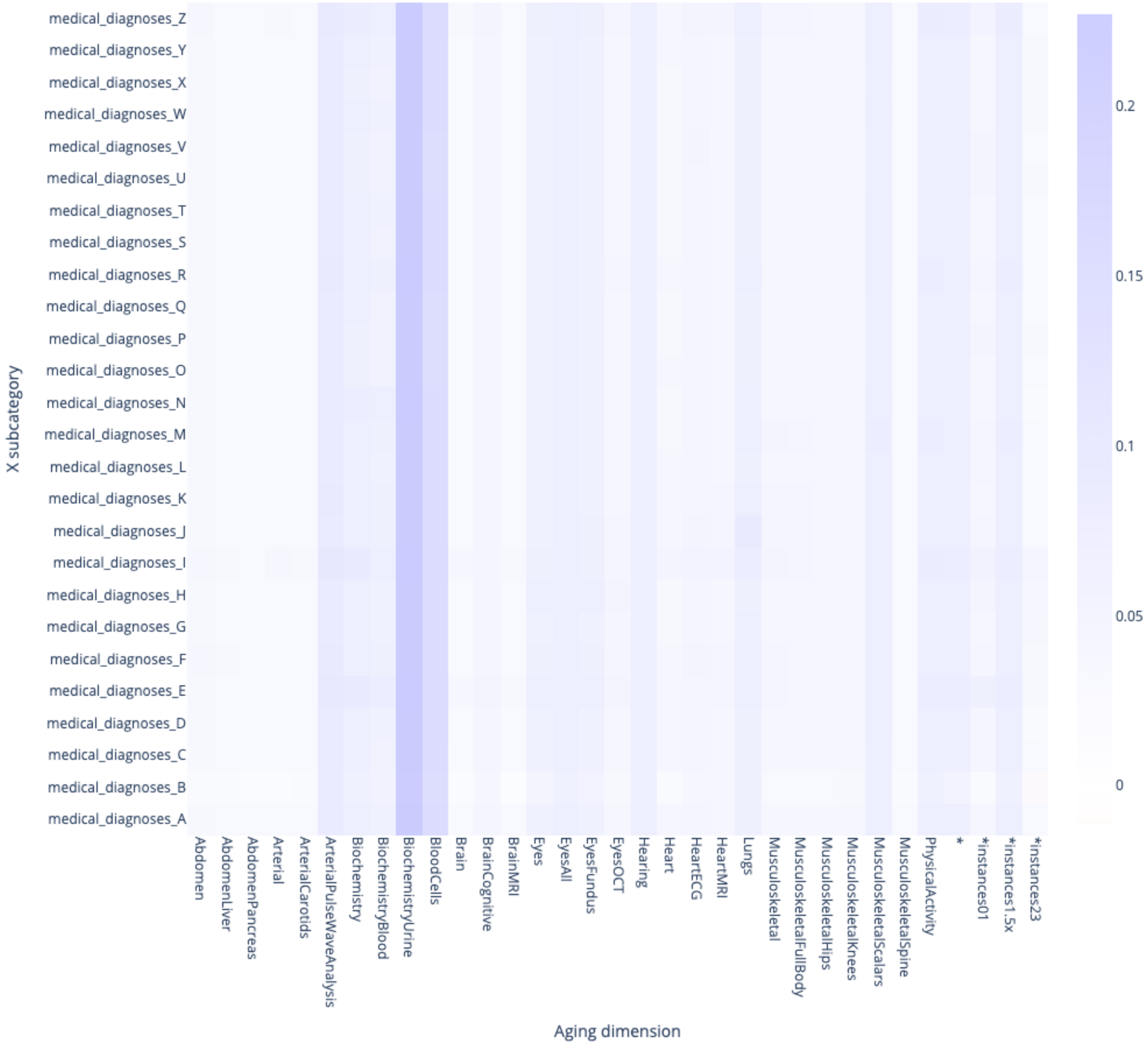

**Supplementary Figure 44: Prediction accuracy for predicting accelerated aging in each aging dimension, from each diseases subcategory**

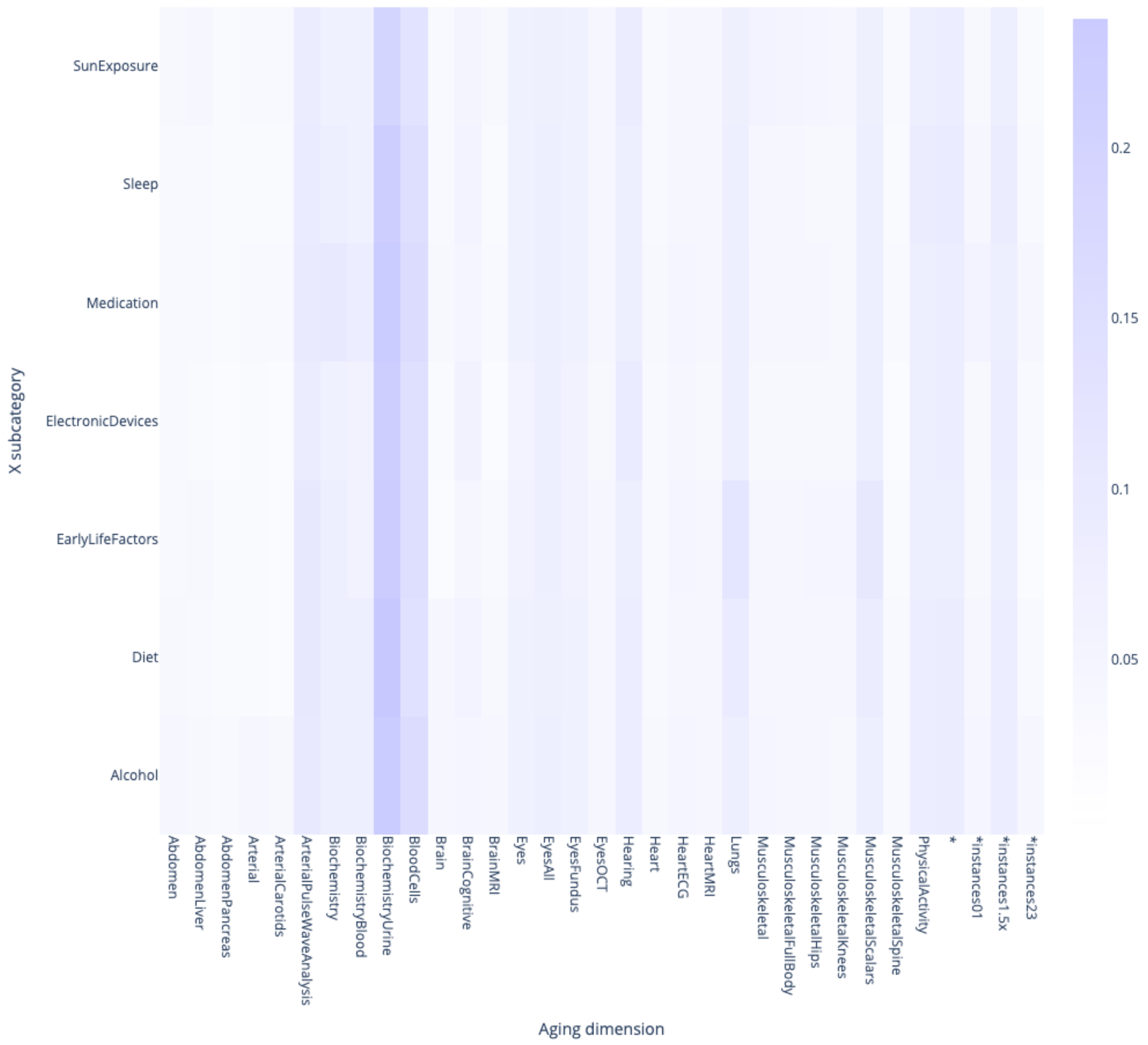

**Supplementary Figure 45: Prediction accuracy for predicting accelerated aging in each aging dimension, from each environmental variable subcategory**

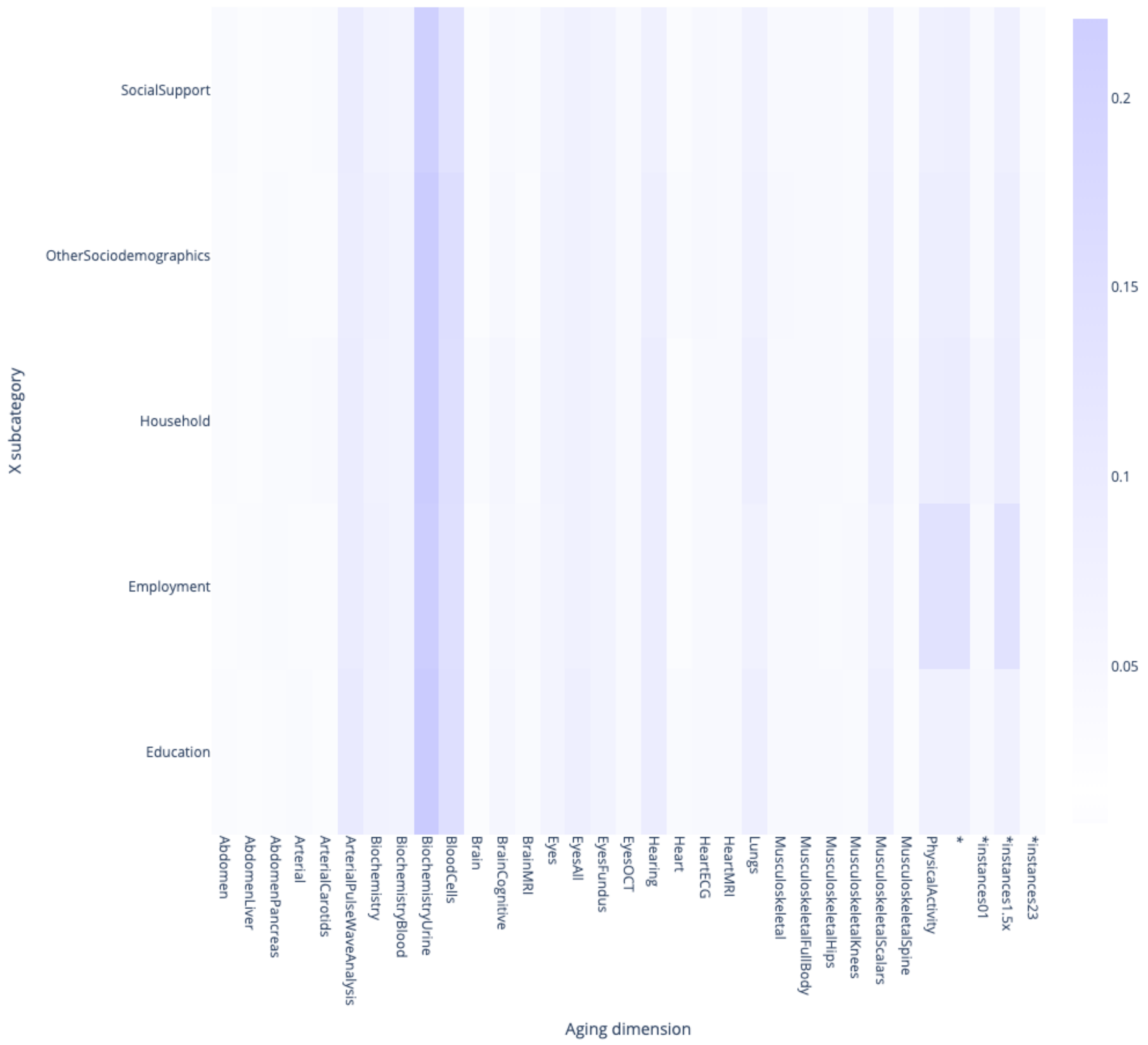

**Supplementary Figure 46: Prediction accuracy for predicting accelerated aging in each aging dimension, from each socioeconomics subcategory**

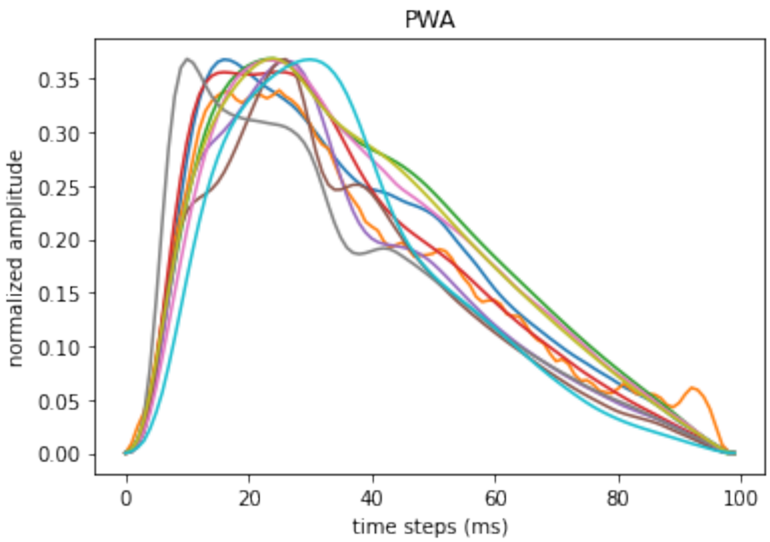

**Supplementary Figure 47: Pulse wave analysis record for ten different participants**

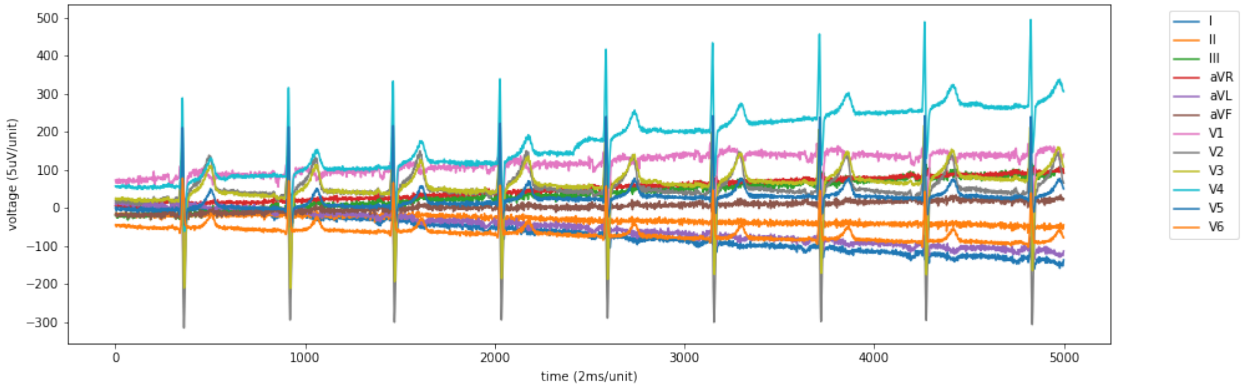

**Supplementary Figure 48: Raw 12-leads ECG sample**

The participant is a 55-60-year-old male participant

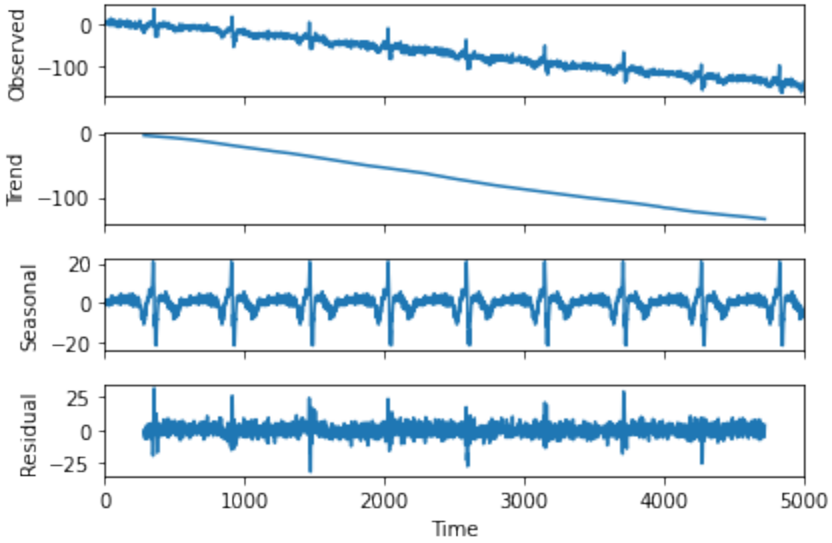

**Supplementary Figure 49: Decomposition of a raw ECG signal into four components**

The first component is the “level” (the mean value), the second is the “trend” (the linear bias over time), the third is the “seasonality” (the periodical component of the signal) and the fourth is the “noise” (the allegedly random remaining variation).

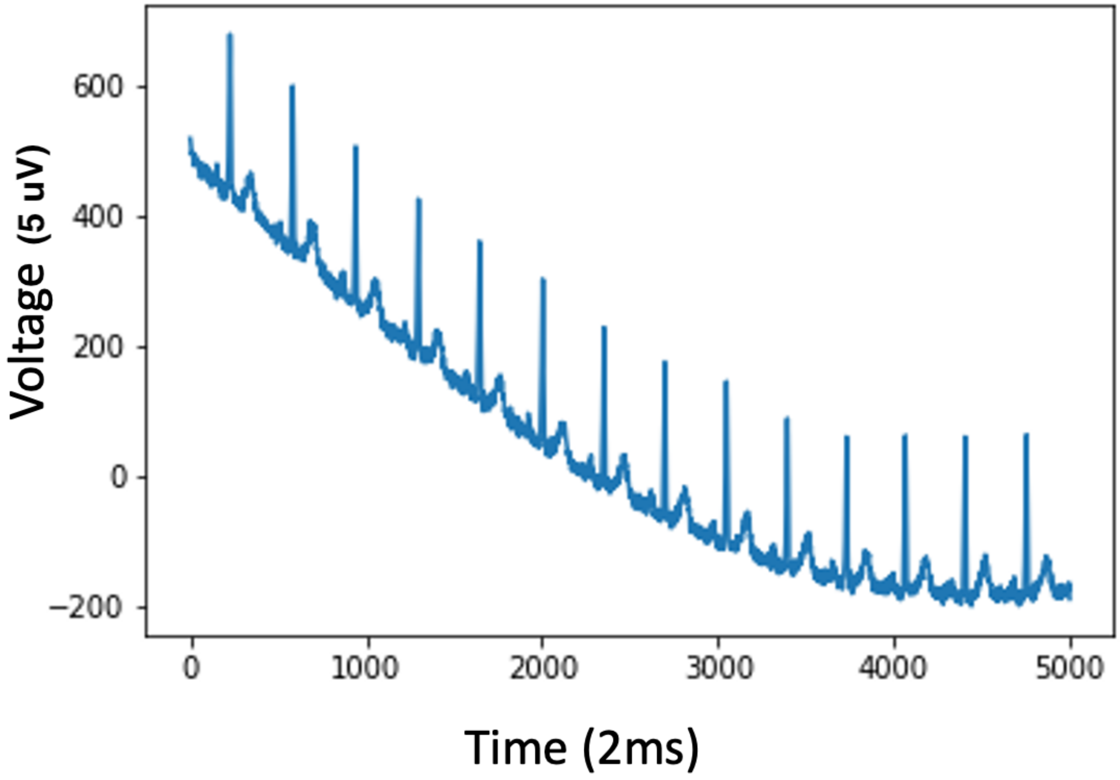

**Supplementary Figure 50: Example of a corrupted ECG recording**

Based on the database the heart rate should be 201, which is not matching the time series.

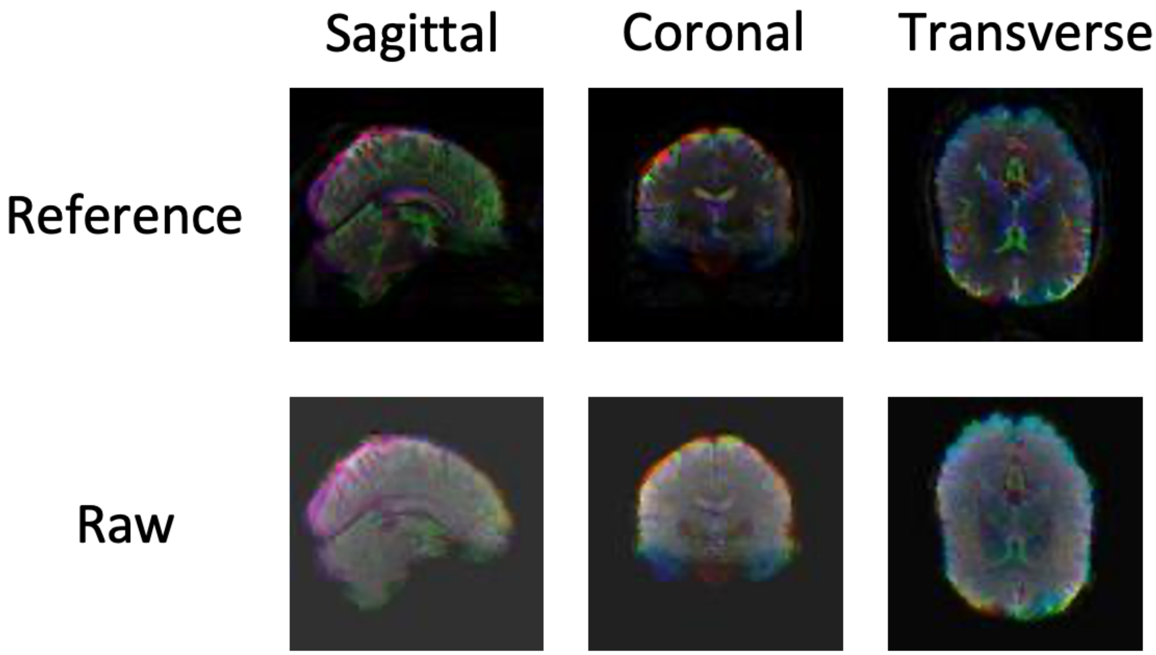

**Supplementary Figure 51**: **Six images (three views and two preprocessings) generated from a brain MRI 3D video**

The participant is a 60-65-year-old male.

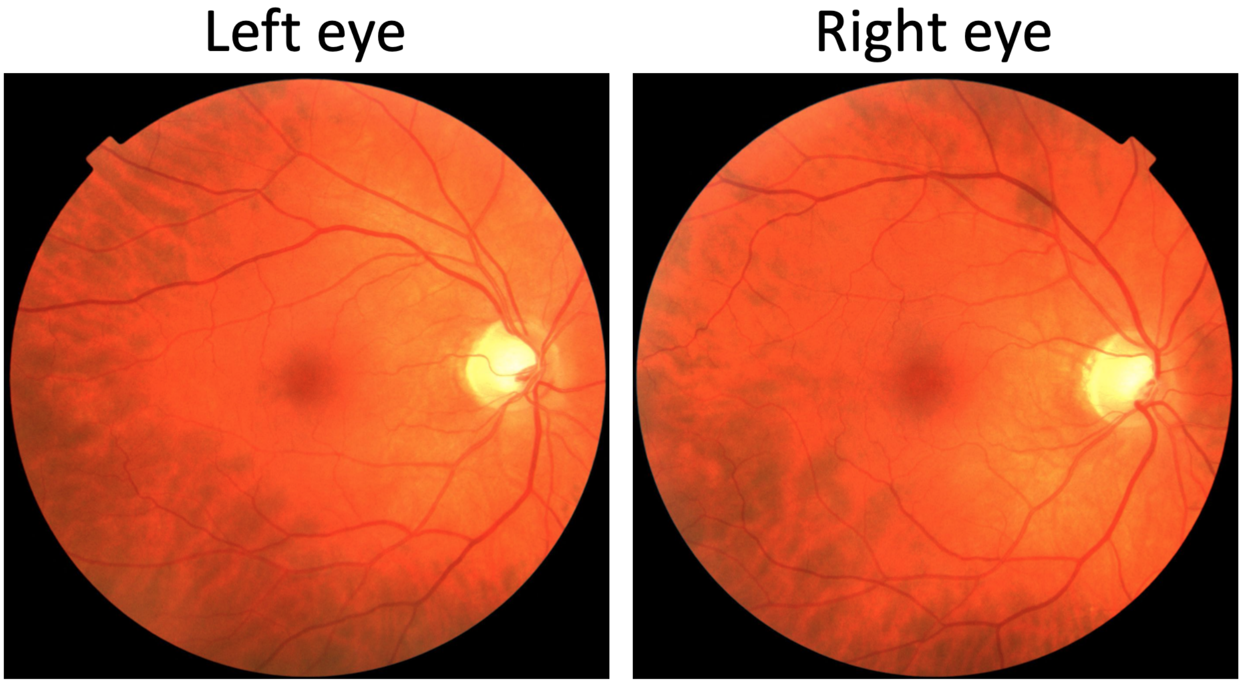

**Supplementary Figure 52**: **Sample preprocessed left and right eye fundus images**

The participant is a 45-50-year-old male.

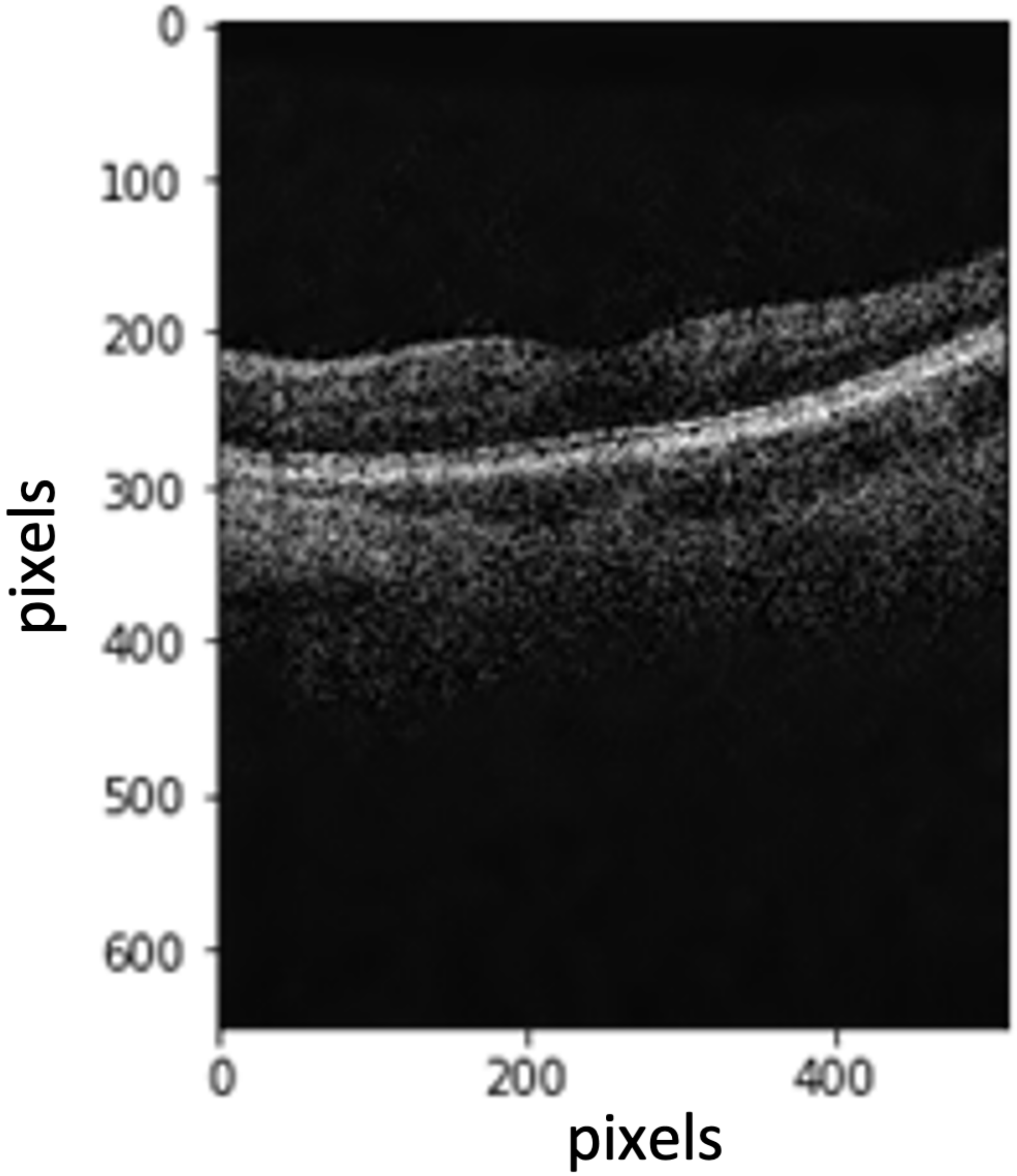

**Supplementary Figure 53: Preprocessing of eye OCT images: raw image**

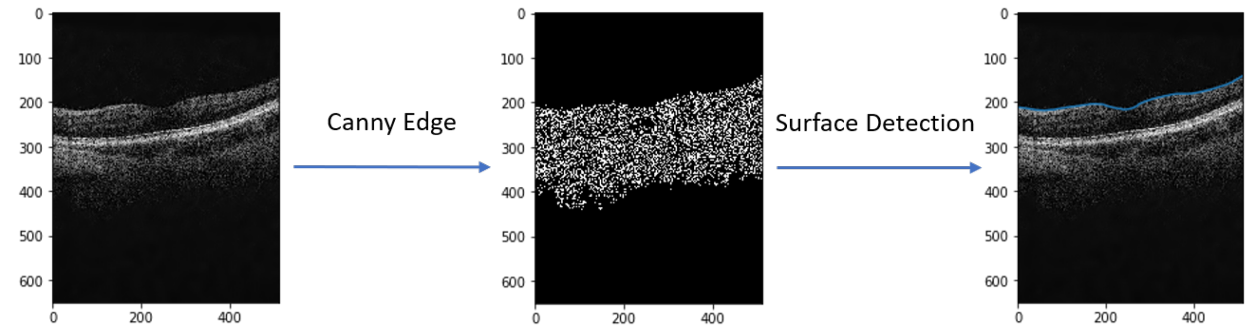

**Supplementary Figure 54: Preprocessing of eye OCT images: detection of the upper surface**

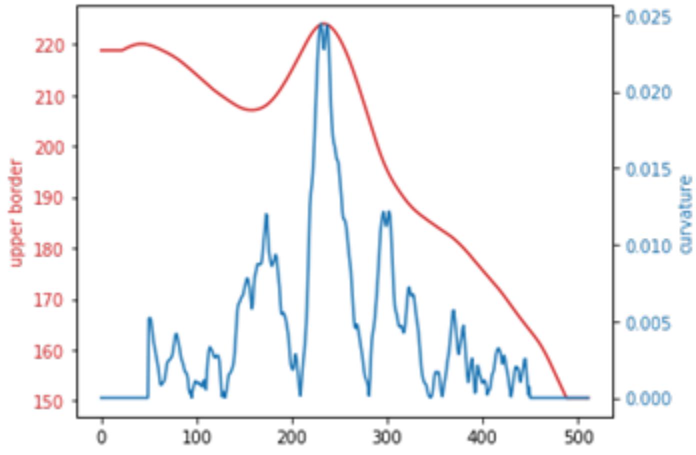

**Supplementary Figure 55: Preprocessing of eye OCT images: computation of the curvature of the upper surface**

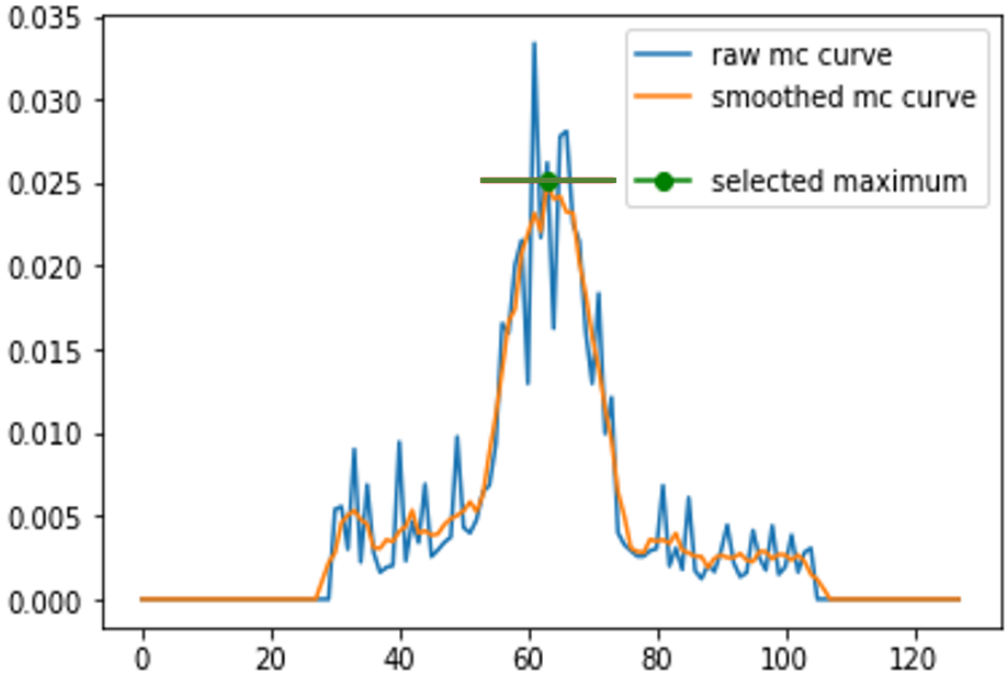

**Supplementary Figure 56: Preprocessing of eye OCT images: detection of image with the maximal curvature after applying the Savitzky–Golay filter**

mc stands for maximal curvature.

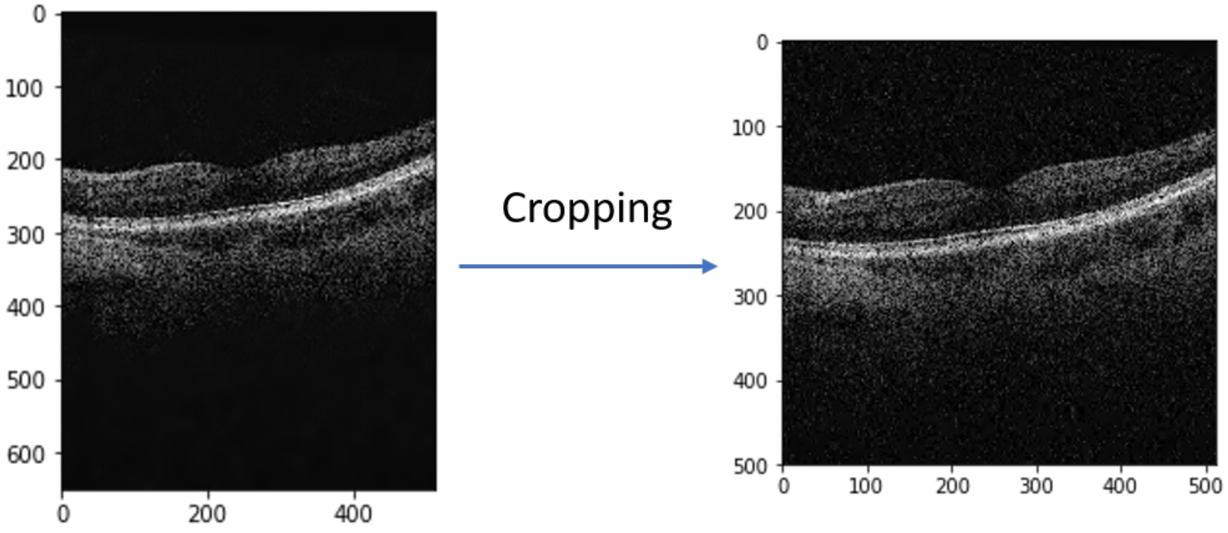

**Supplementary Figure 57: Preprocessing of eye OCT images: raw image vs. cropped image**

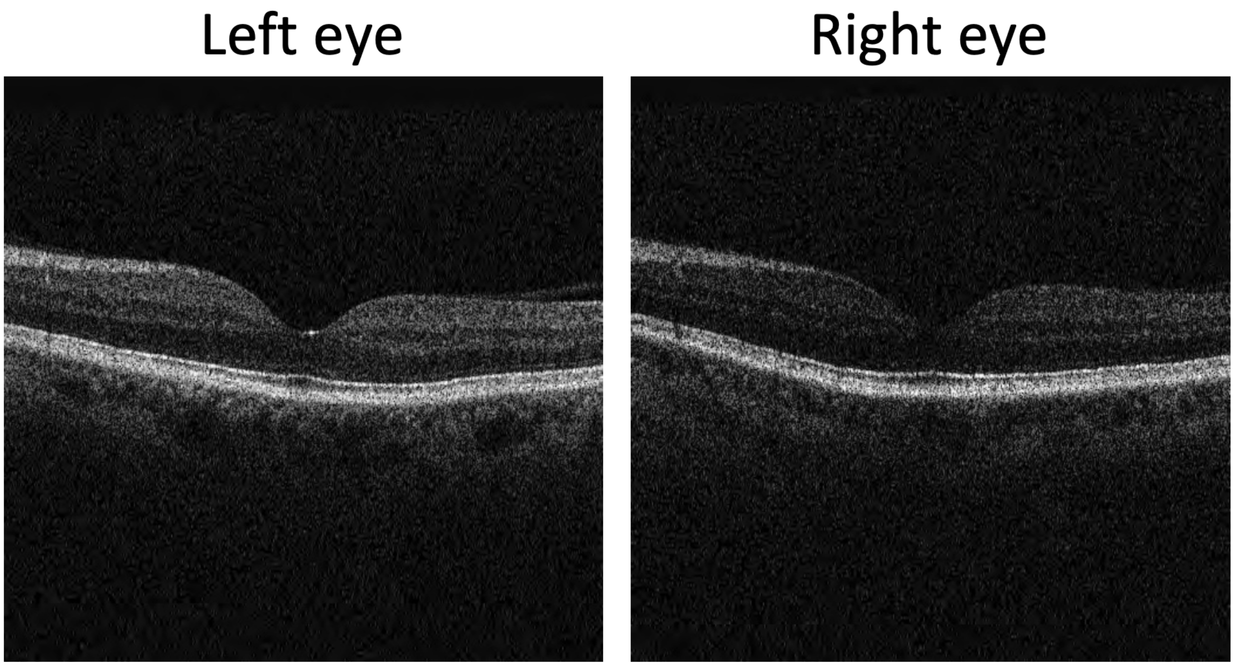

**Supplementary Figure 58: Sample preprocessed OCT left and right eyes images**

The participant is a 60-65-year-old male.

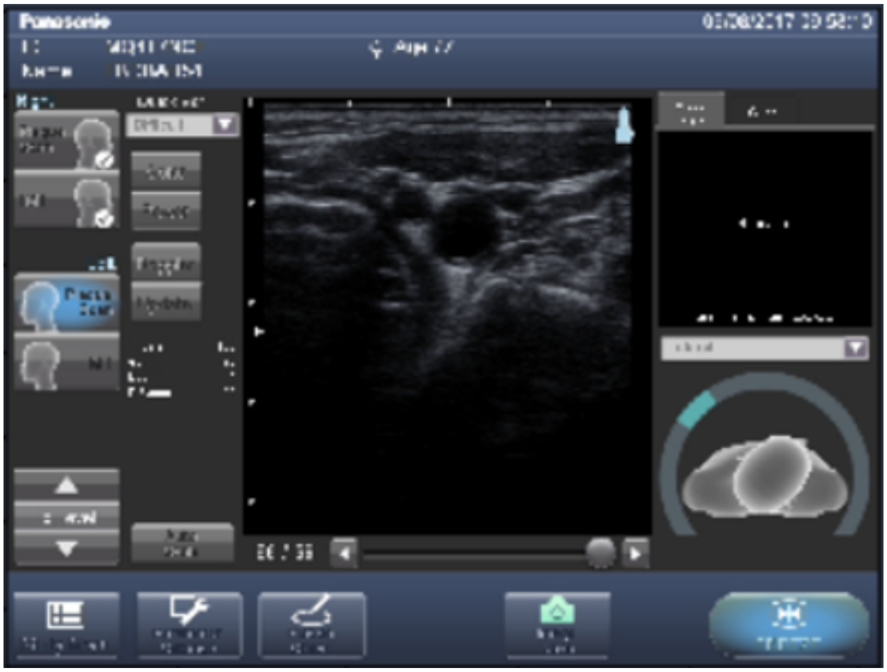

**Supplementary Figure 59: Preprocessing of carotid ultrasound images: screenshot of the software used to record the images as shared by UK Biobank**

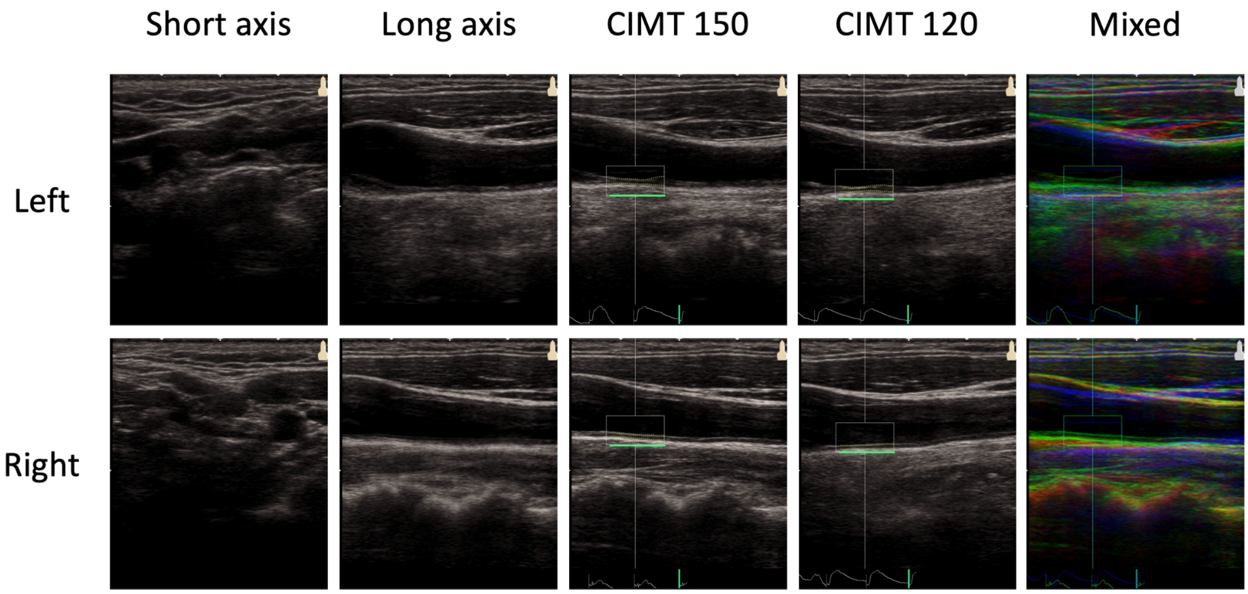

**Supplementary Figure 60**: **Sample preprocessed carotid ultrasound images**

The participant is a 55-60-year-old male.

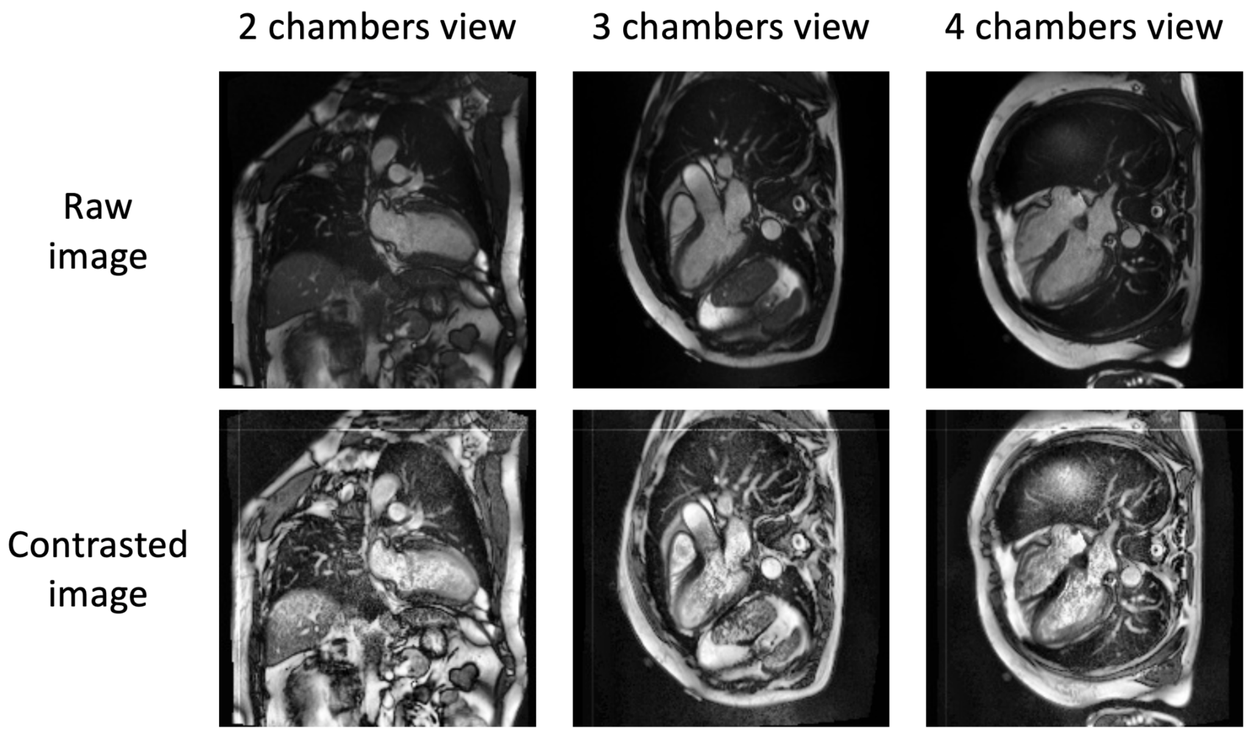

**Supplementary Figure 61**: **Sample preprocessed heart MRI images**

The participant is a 60-65-year-old male.

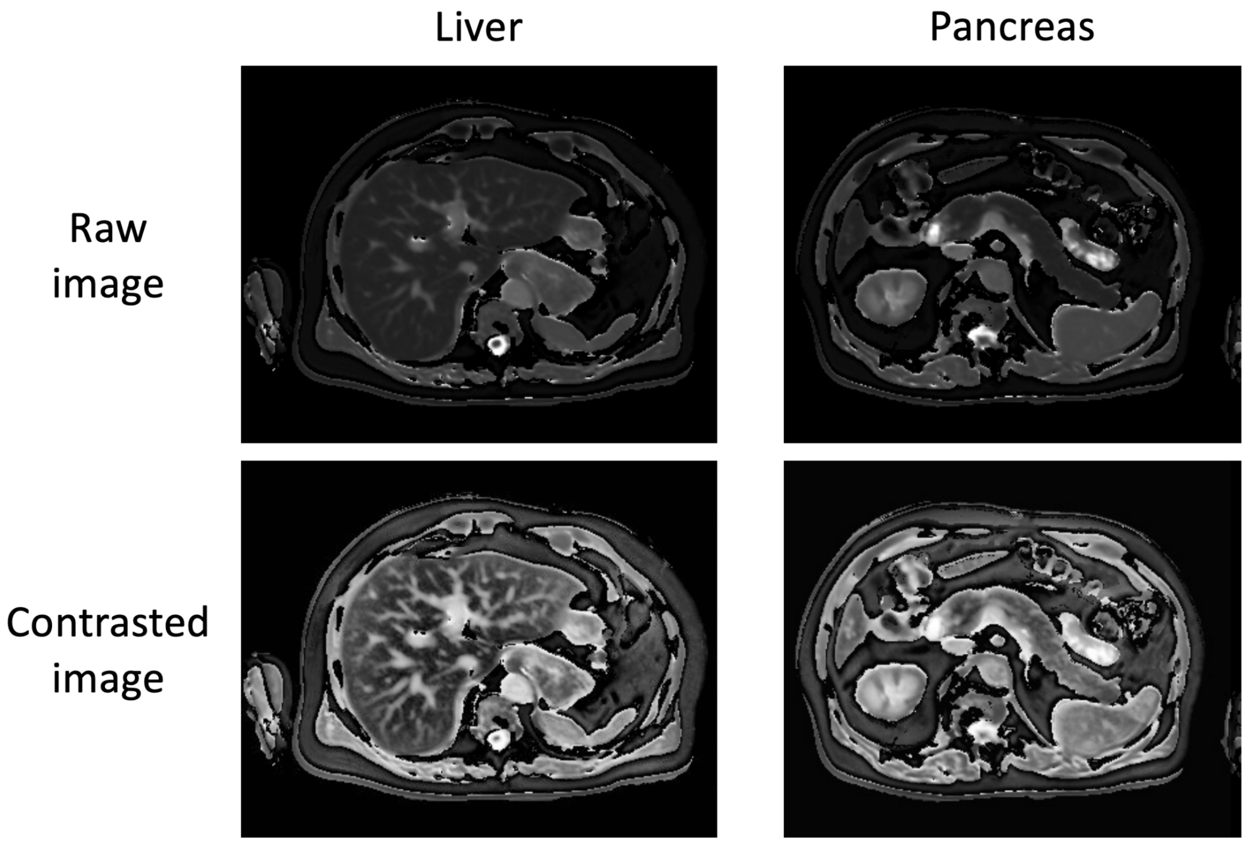

**Supplementary Figure 62**: **Sample preprocessed abdominal (liver and pancreas) MRI images**

The participant is a 60-65-year-old male.

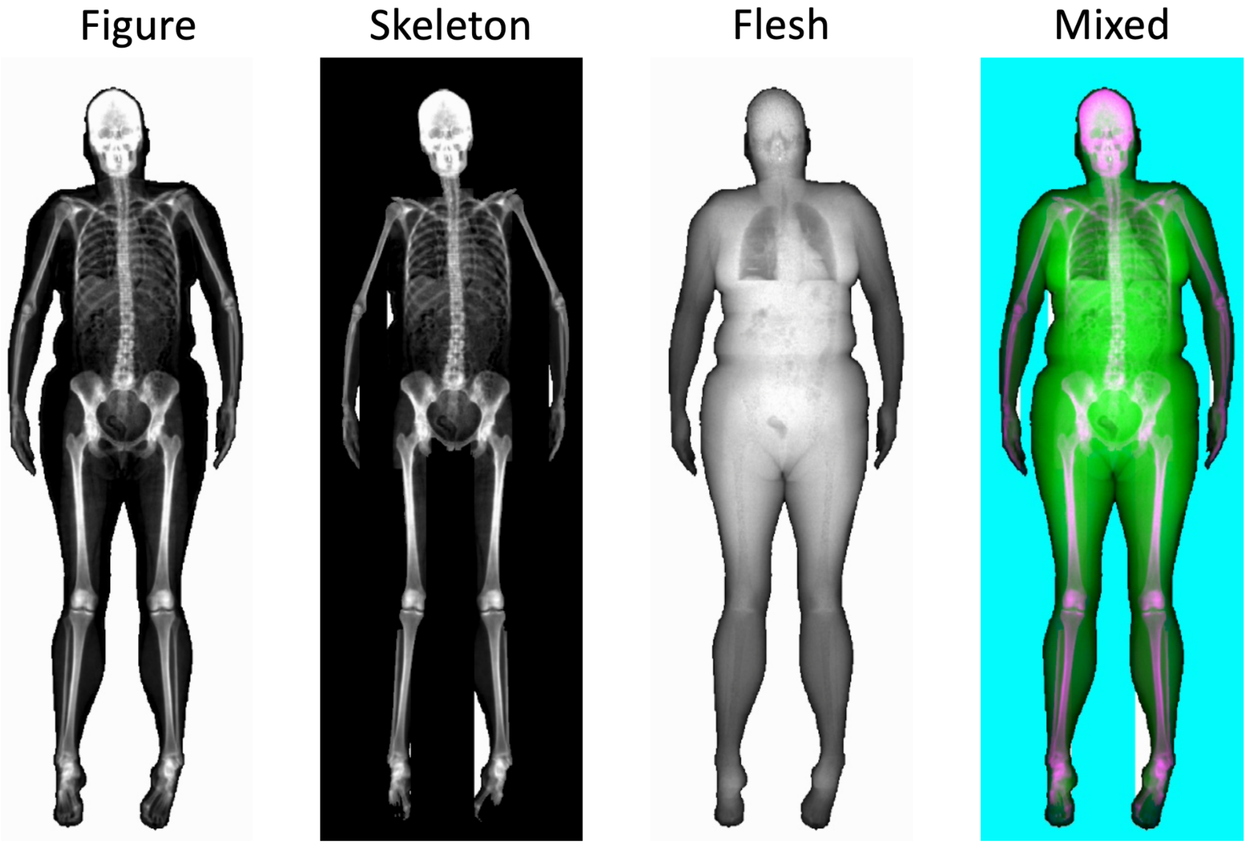

**Supplementary Figure 63**: **Sample preprocessed musculoskeletal full body X-ray images**

The participant is a 50-55-year-old female.

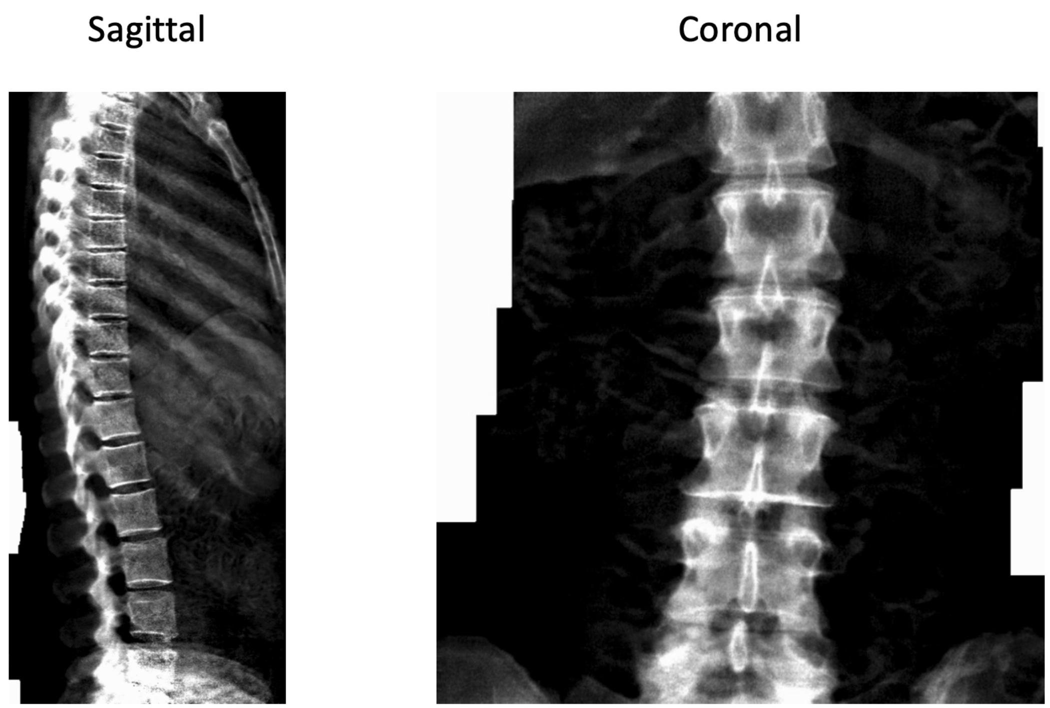

**Supplementary Figure 64: Sample preprocessed spine X-ray images**

The participant is a 50-55-year-old female.

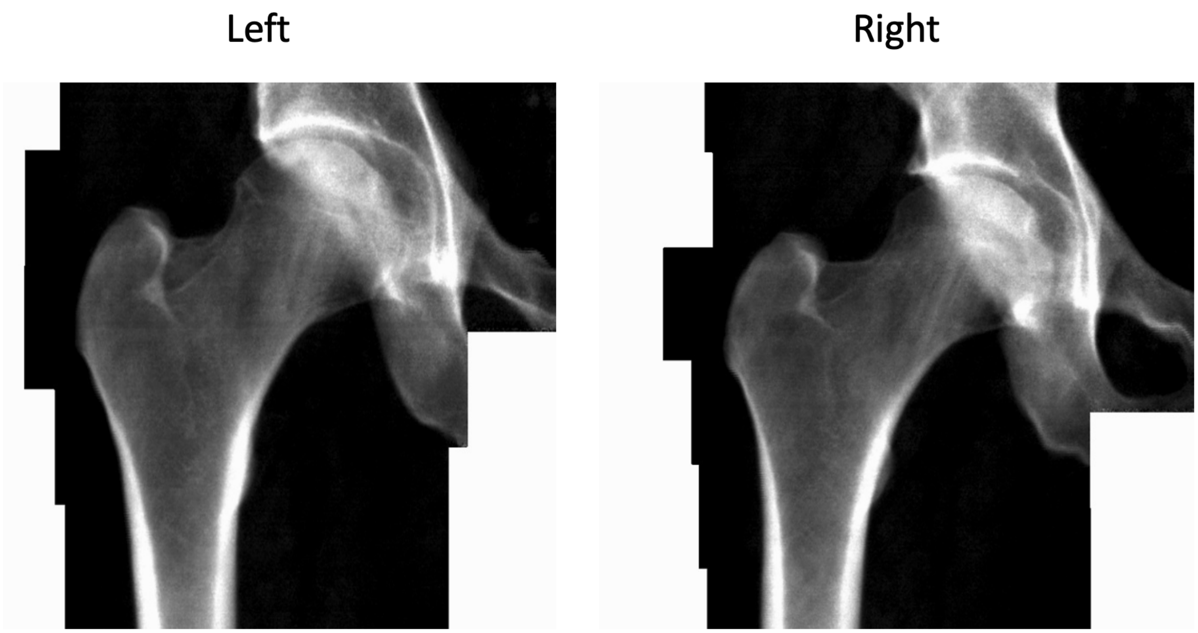

**Supplementary Figure 65: Sample preprocessed hip X-ray images**

The participant is a 60-65-year-old male.

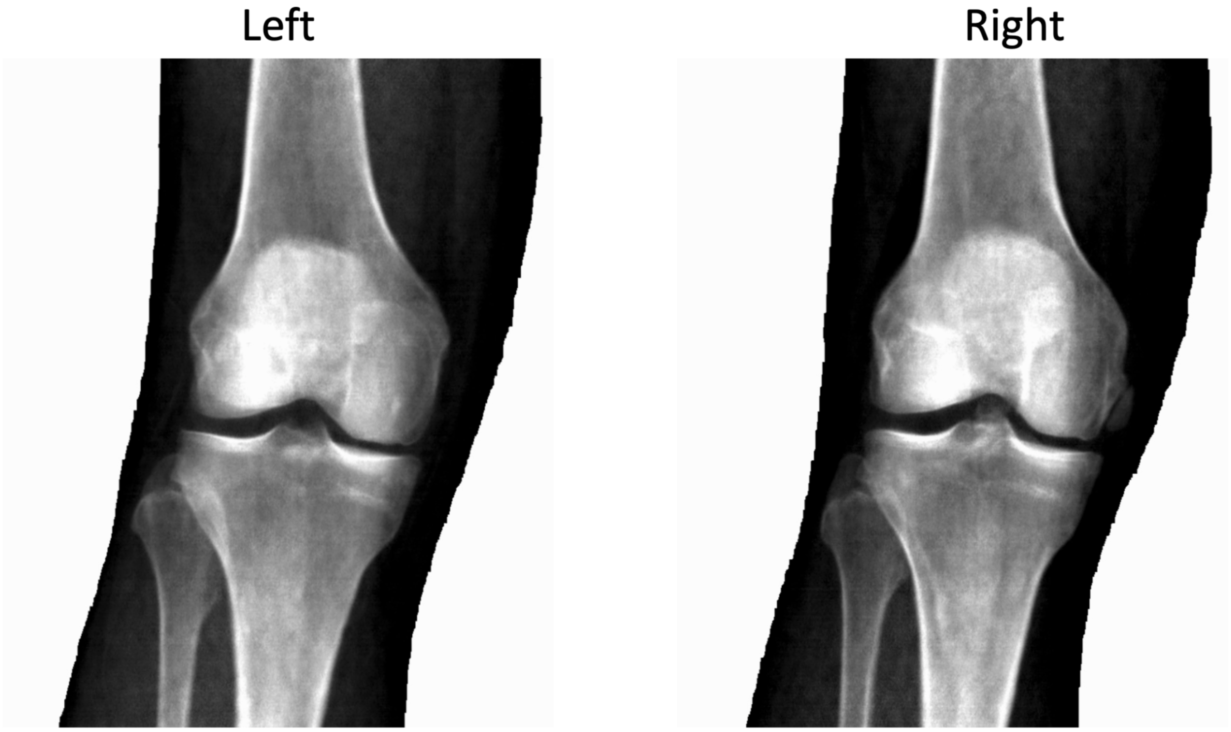

**Supplementary Figure 66: Sample preprocessed knee X-ray images**

The participant is a 60-65-year-old male.

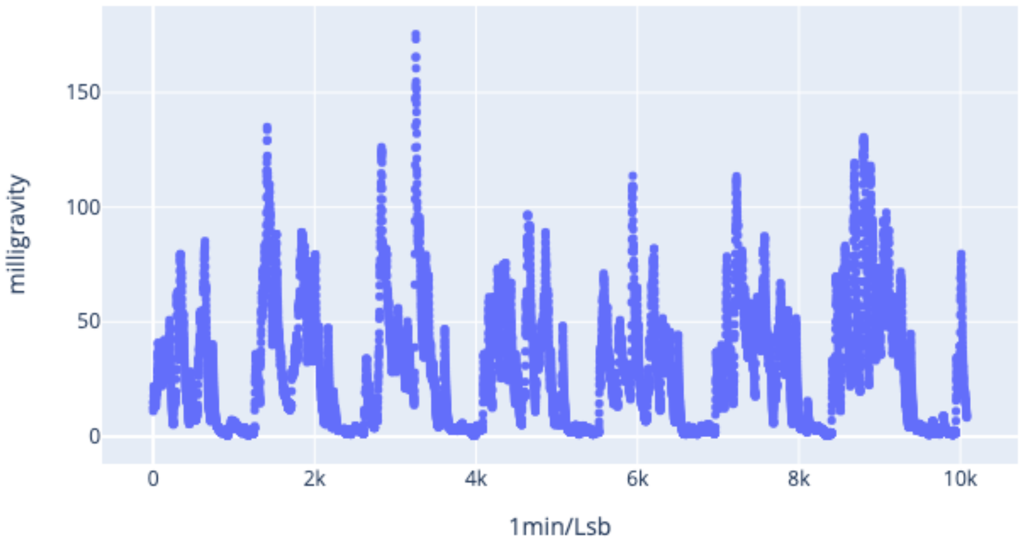

**Supplementary Figure 67**: **Sample preprocessed one-dimensional acceleration time series generated from a participant's wrist accelerometer recording**

The participant is a 65-70-year-old male.

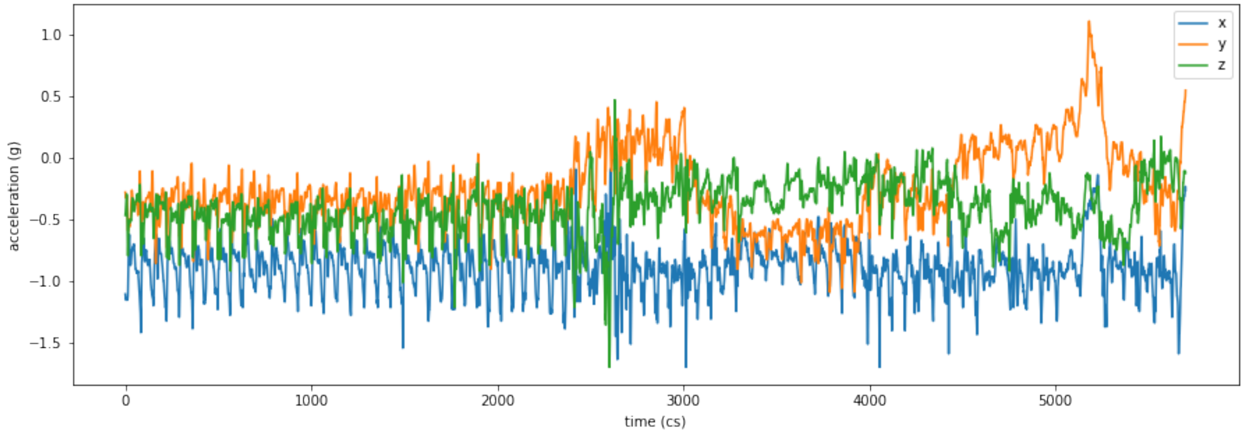

**Supplementary Figure 68**: **Sample preprocessed three-dimensional acceleration time series generated from a participant's wrist accelerometer recording during their walk**

The participant is a 40-45-year-old female participant.

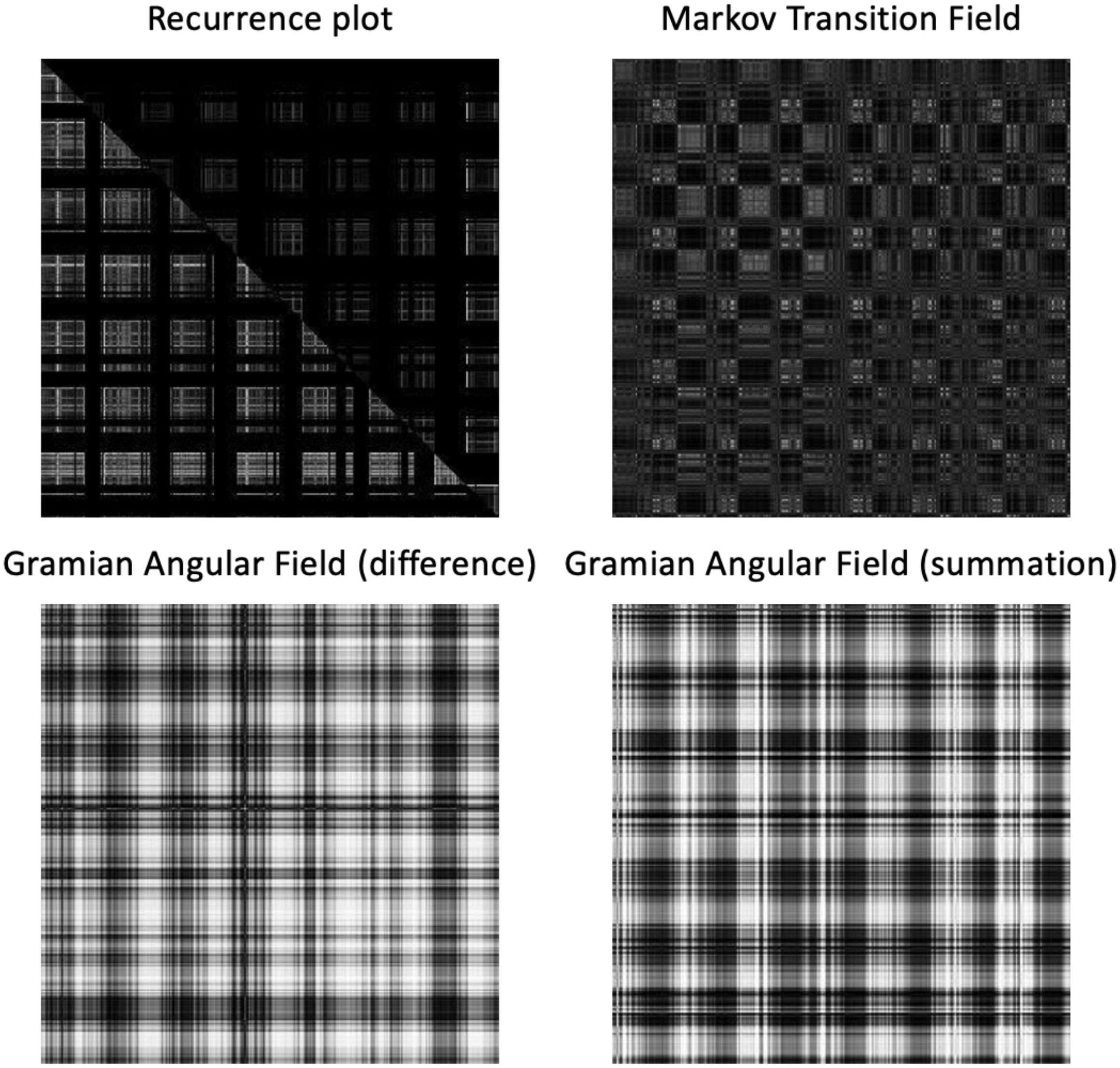

**Supplementary Figure 69: Recurrence plot, Markov transition field and Gramian angular field (difference and summation) generated from a participant's wrist accelerometer recording**

The participant is a 65-70-year-old male. For the recurrence plot, the upper right triangle corresponds to the sleeping timesteps, and the lower left triangle corresponds to the remaining time steps.

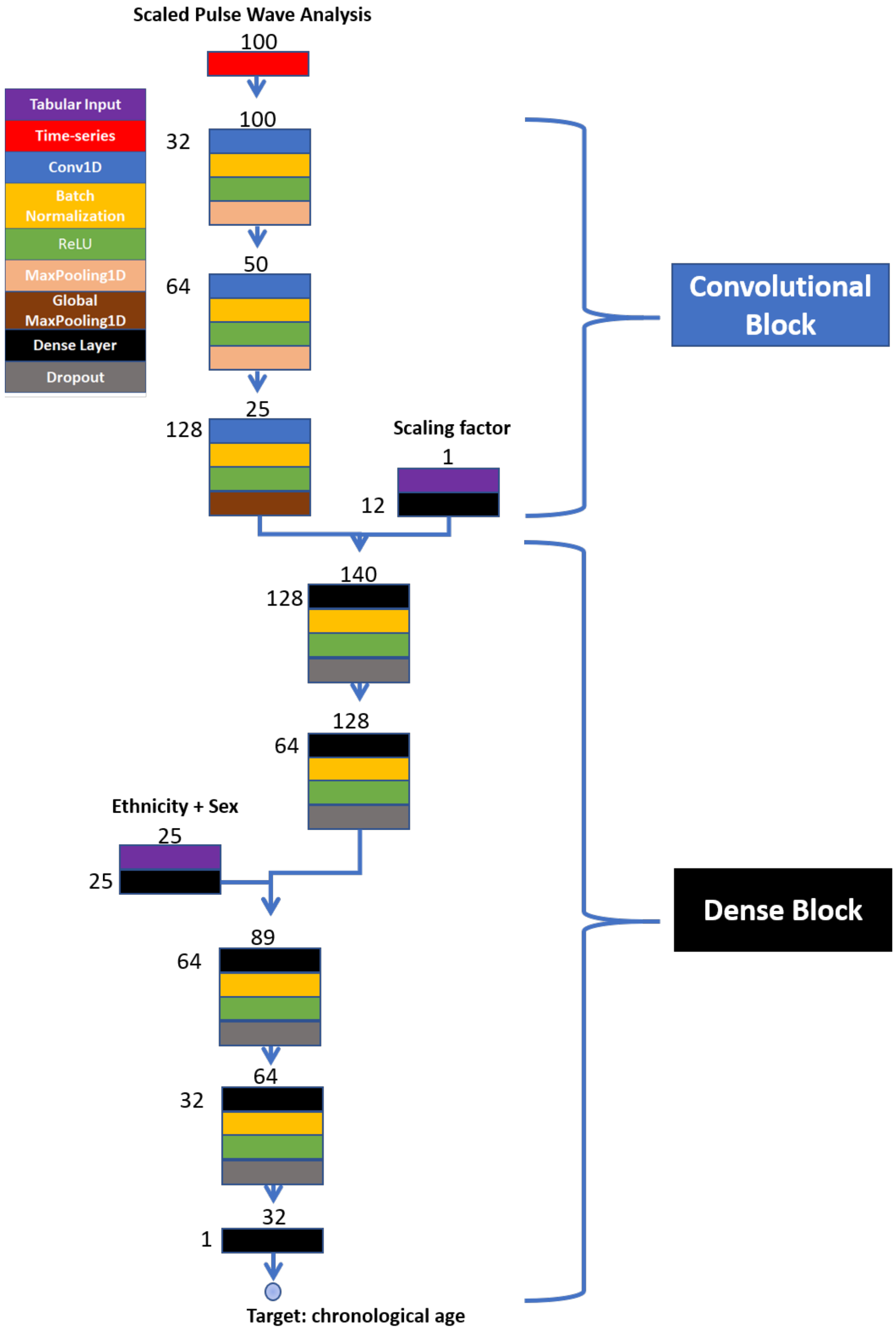

**Supplementary Figure 70: Architecture of the convolutional neural network trained on pulse wave analysis records - Summary figure**

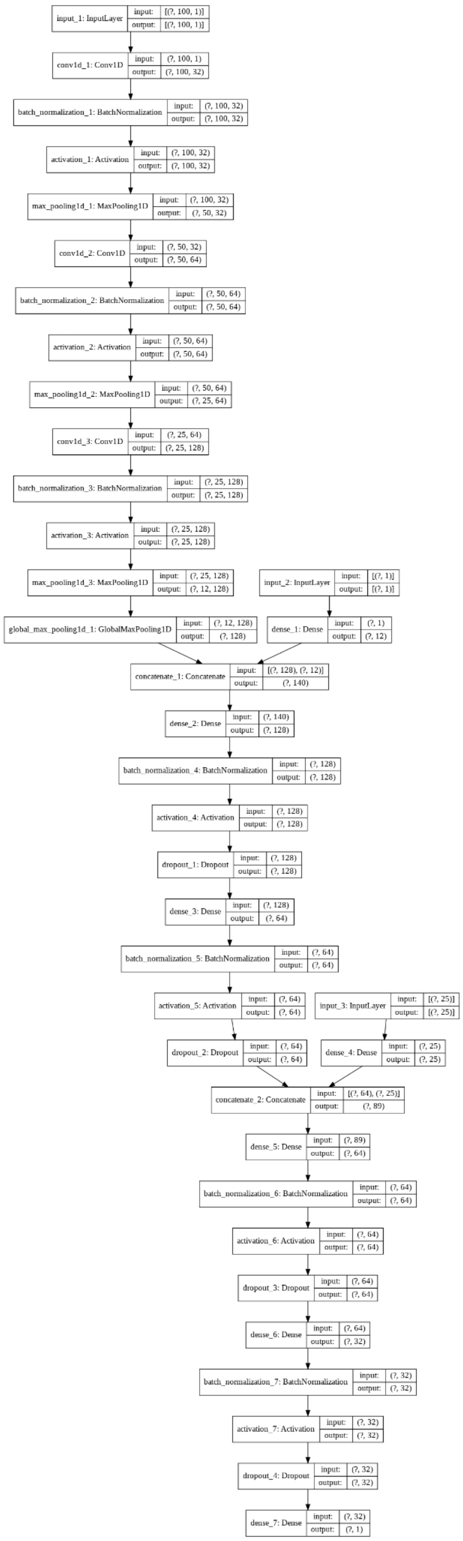

**Supplementary Figure 71: Architecture of the convolutional neural network trained on pulse wave analysis records - Detailed figure**

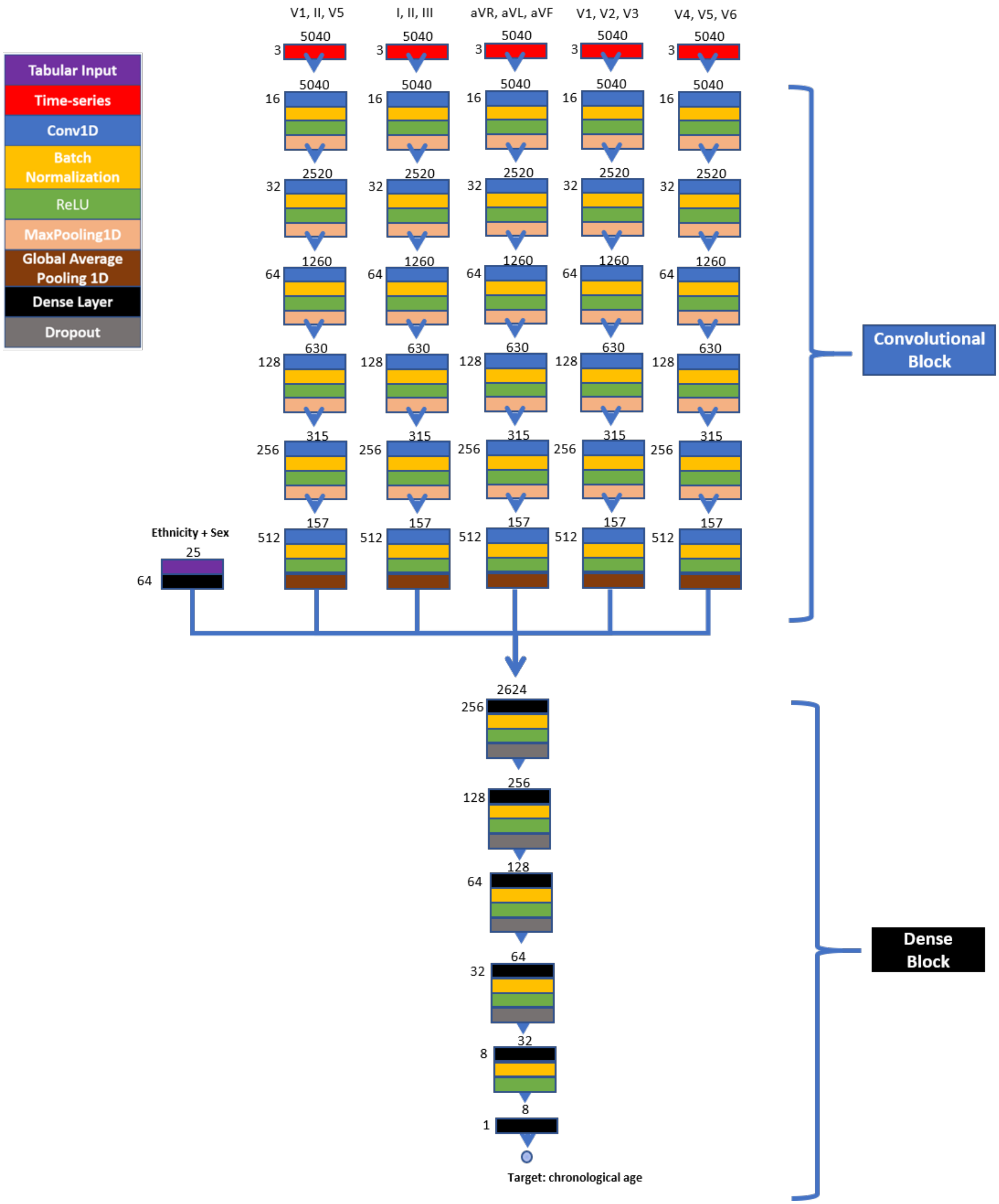

**Supplementary Figure 72: Architecture of the convolutional neural network trained on ECG records - Summary figure**

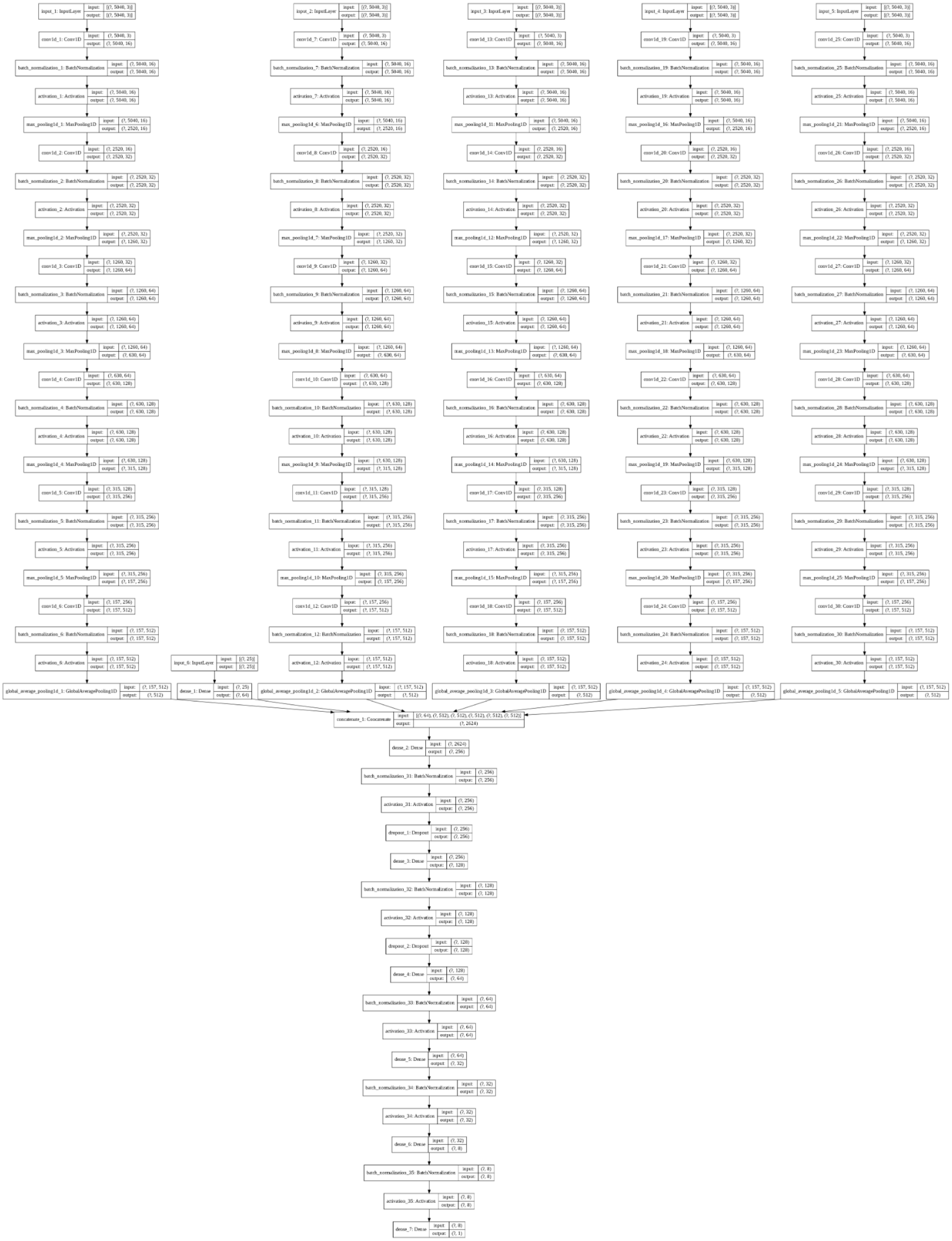

**Supplementary Figure 73: Architecture of the convolutional neural network trained on ECG records - Detailed figure**

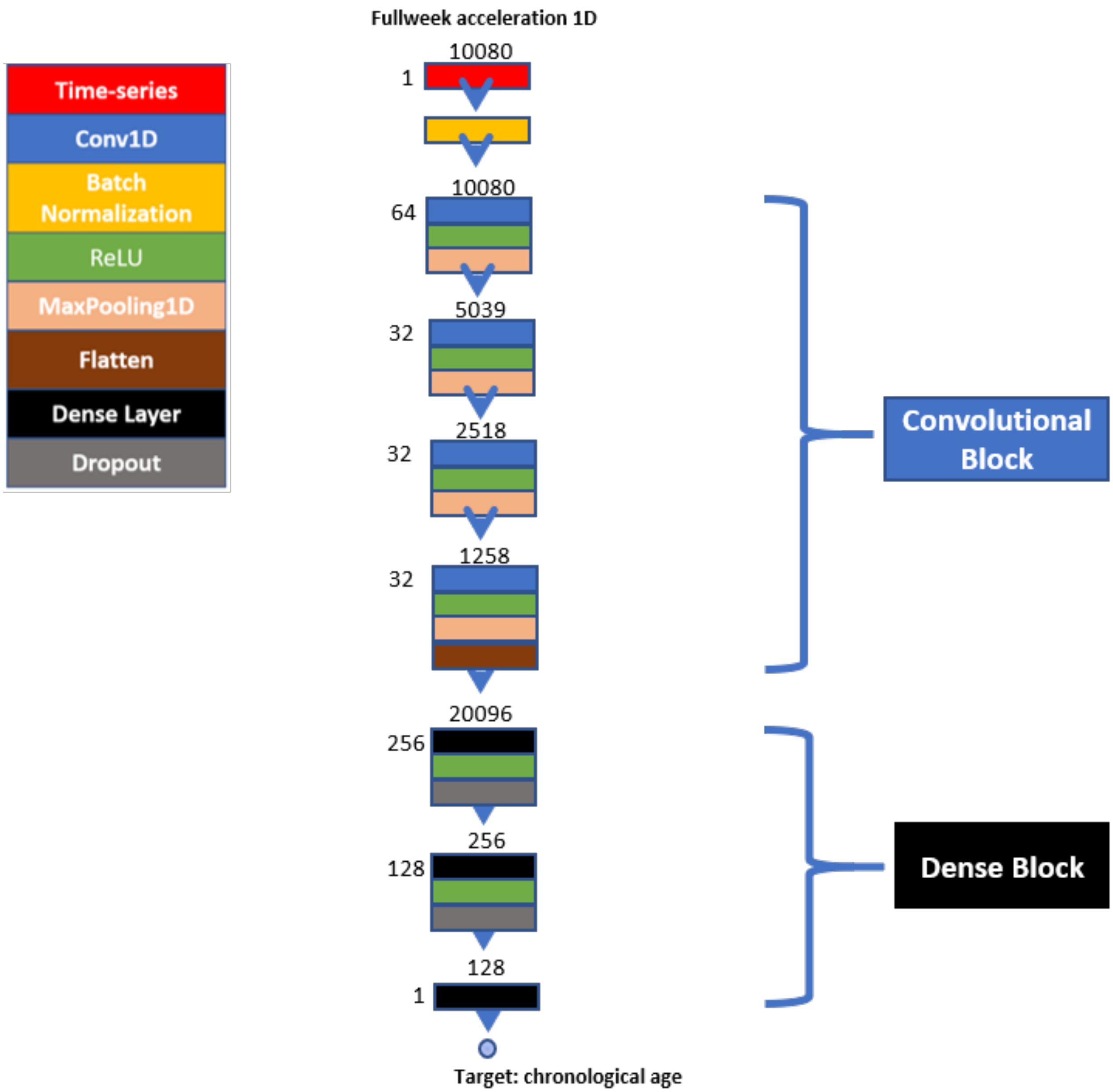

**Supplementary Figure 74: Architecture of the convolutional neural network trained on the raw wrist accelerometer acceleration time series - Summary figure**

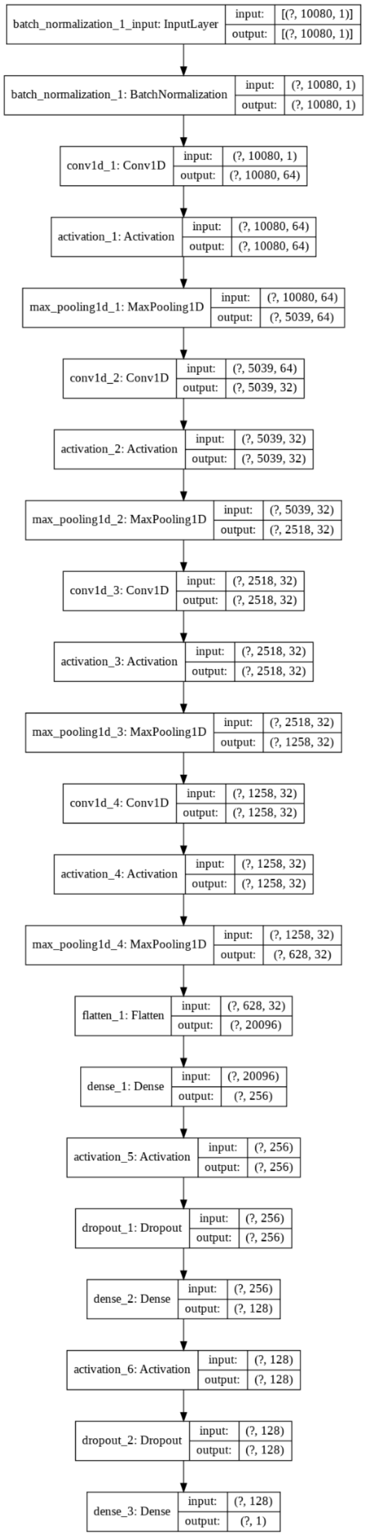

**Supplementary Figure 75: Architecture of the convolutional neural network trained on the raw wrist accelerometer acceleration time series - Detailed figure**

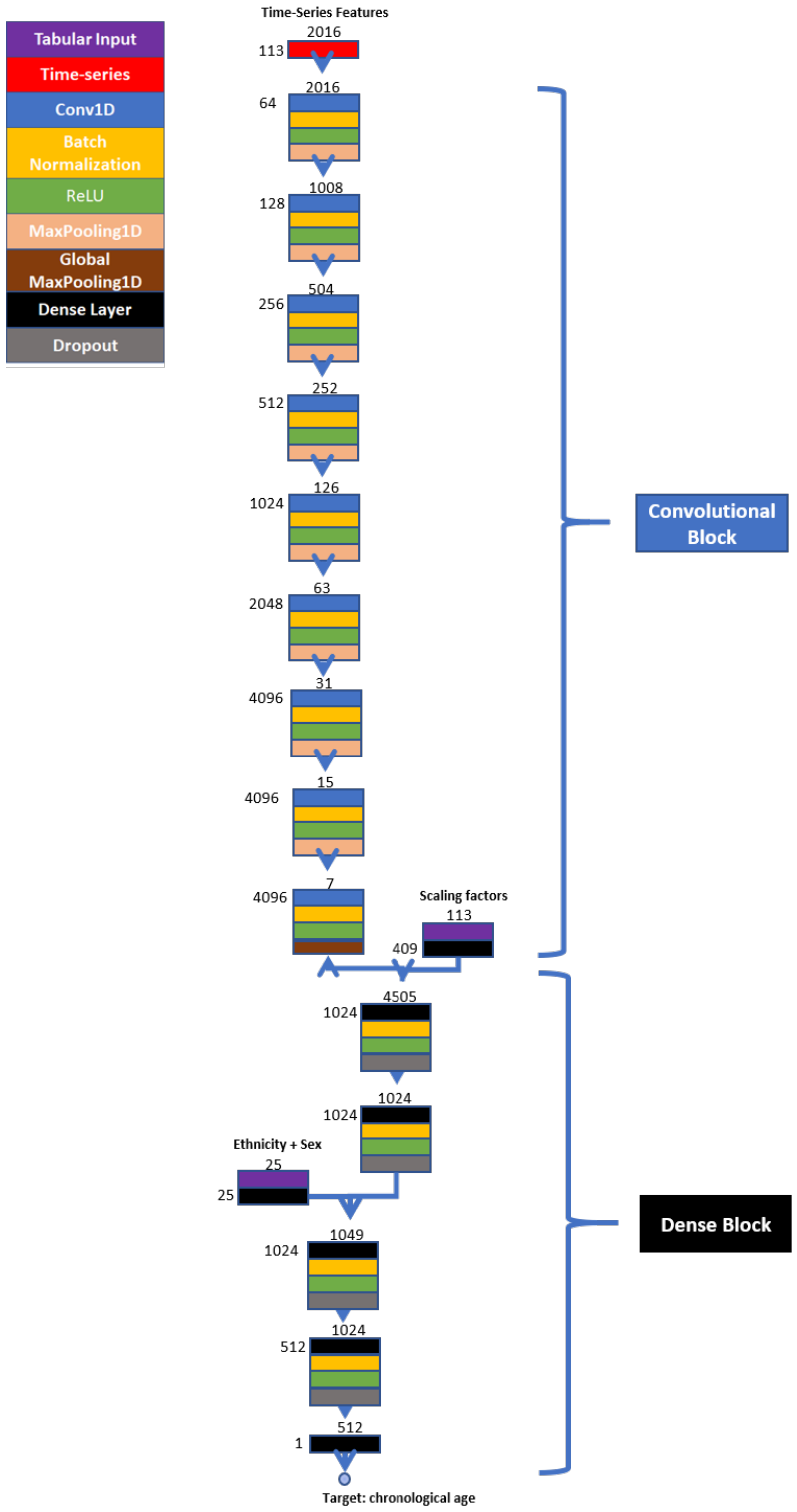

**Supplementary Figure 76: Architecture of the convolutional neural network trained on the wrist accelerometer features time series - Summary figure**

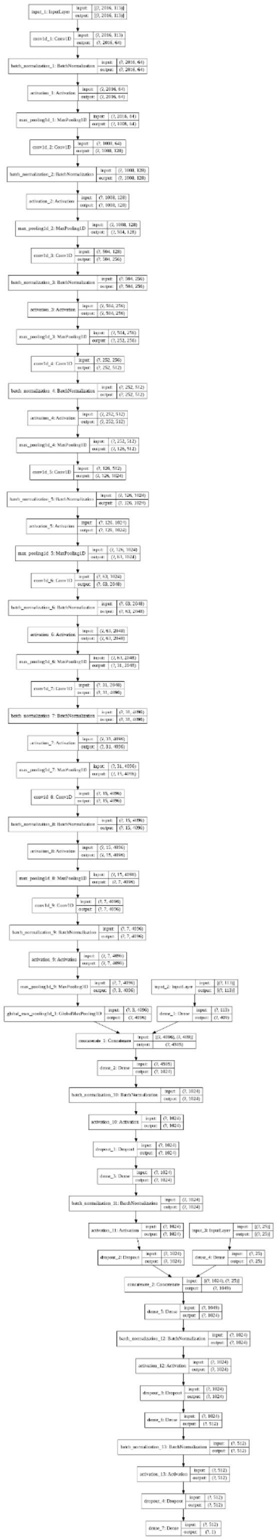

**Supplementary Figure 77: Architecture of the convolutional neural network trained on the wrist accelerometer features time series - Detailed figure**

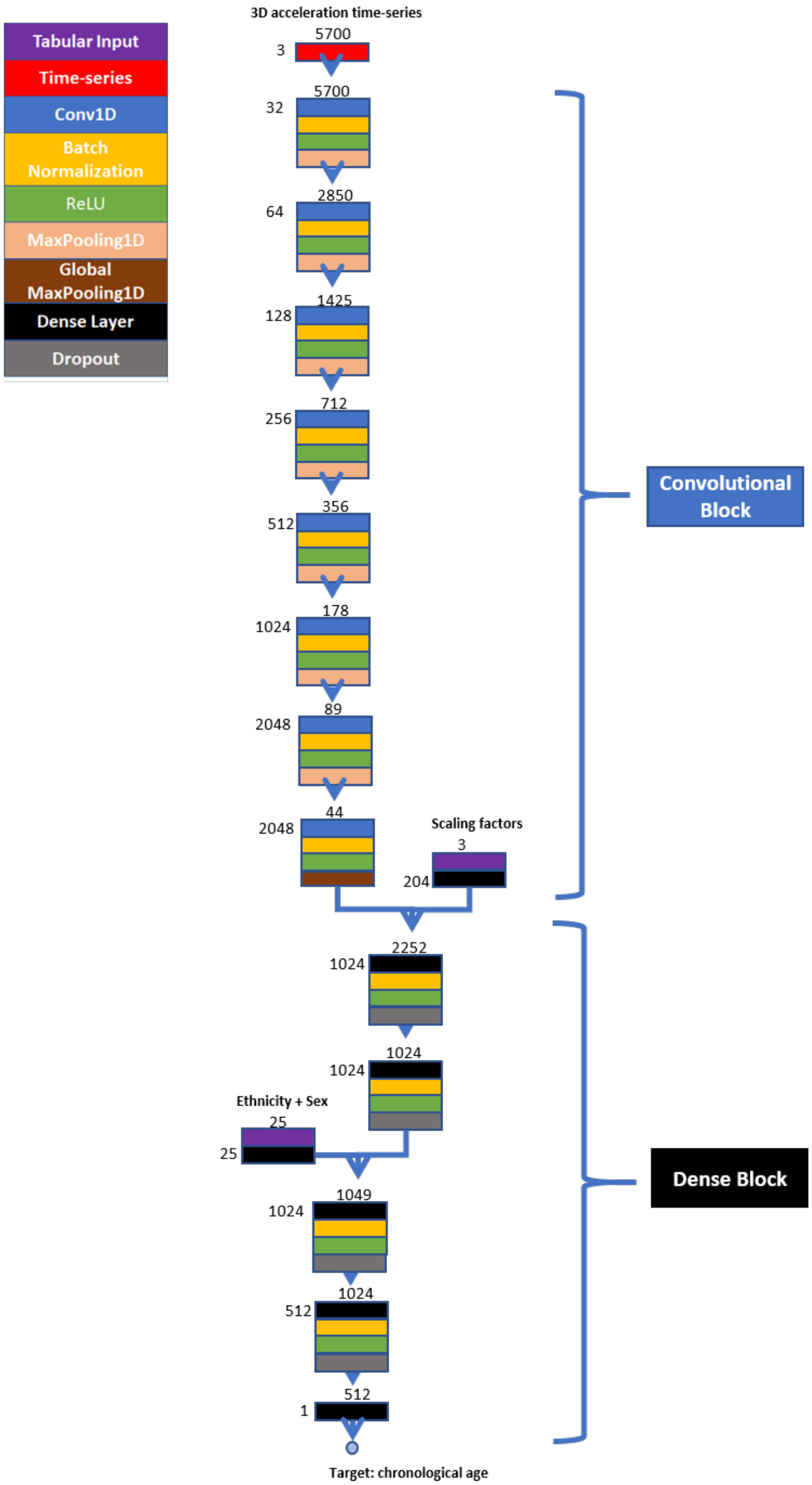

**Supplementary Figure 78: Architecture of the convolutional neural network trained on the wrist accelerometer 3D raw acceleration time series - Summary figure**

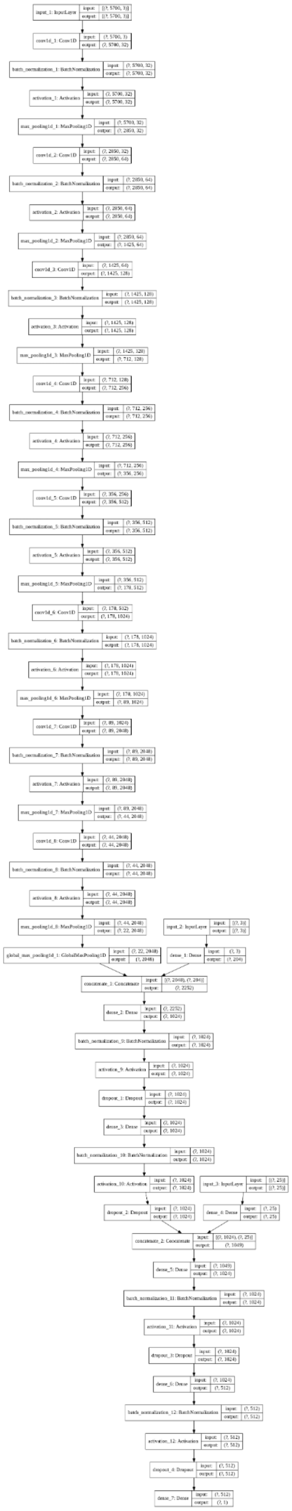

**Supplementary Figure 79: Architecture of the convolutional neural network trained on the wrist accelerometer 3D raw acceleration time series - Detailed figure**

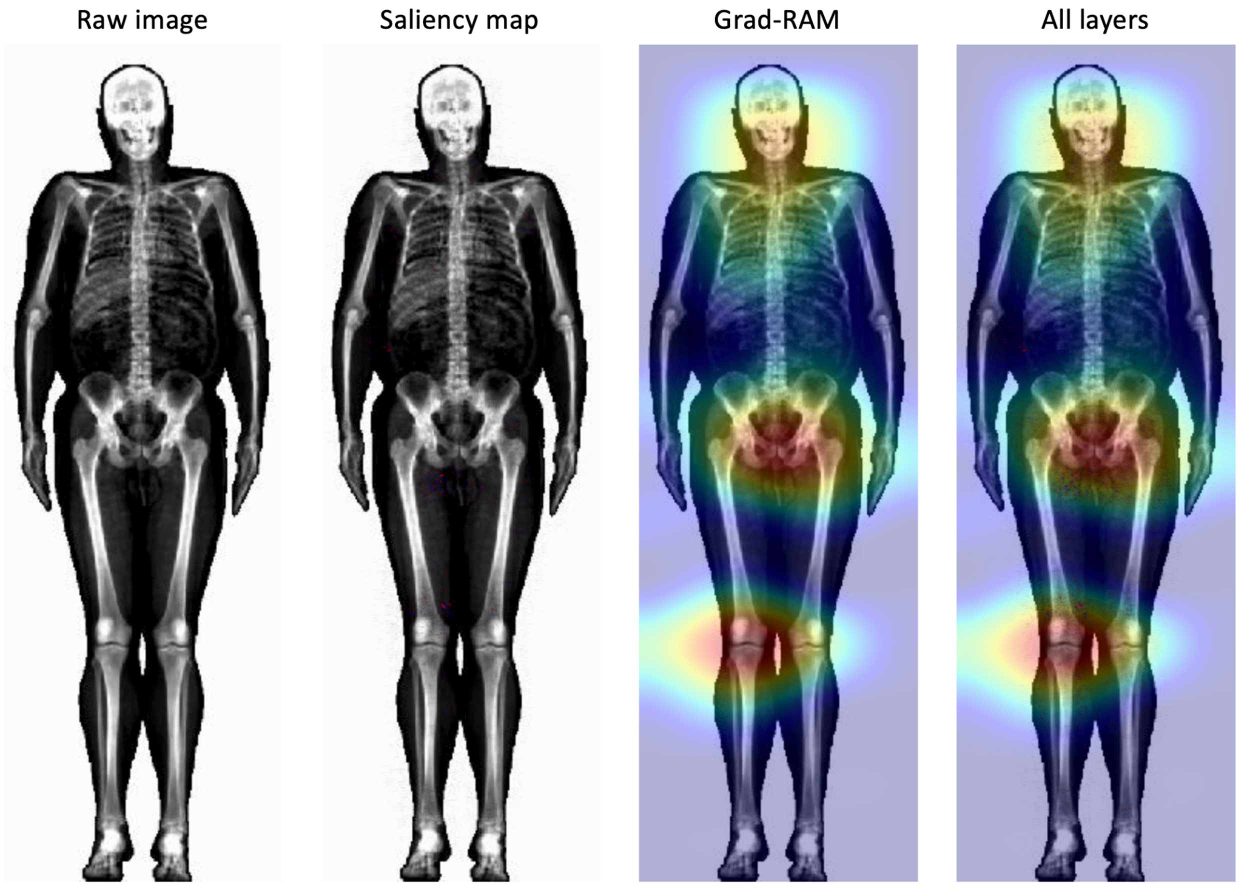

**Supplementary Figure 80: Attention map algorithms: saliency map and Grad-RAM.**

The participant is a 60-65-year-old male whose chronological age was accurately predicted by the machine using a “Figure” musculoskeletal X-ray image.

### Supplementary Tables

**Supplementary Table 1: XWAS Summary - Average correlation between aging main dimensions and between aging subdimensions for each X category**

| X Category | Main dimensions | Aging subdimensions |
|---|---|---|
| **Phenotypic** | .130±.087 | .261±.136 |
| **Genetics** | .128±.237 | .338±.264 |
| **Biomarkers** | .359±.229 | .656±.309 |
| **Clinical phenotypes** | .643±.142 | .668±.336 |
| **Diseases** | .488±.198 | .585±.187 |
| **Family history** | .030±.519 | .728±.397 |
| **Environmental variables** | .639±.180 | .624±.321 |
| **Socioeconomics** | .607±.309 | .639±.421 |

**Supplementary Table 2: Comparison between the models trained on scalar features**

| Main dimension | Subdimension | Sub-subdimension | Number of Predictors (non-demographics) | R-Squared_ElasticNet | R-Squared_LightGBM | R-Squared_NeuralNetwork |
|---|---|---|---|---|---|---|
| Brain | MRI | GreyMatterVolumes | 139 | 0.494±0.012 | 0.505±0.009 | 0.511±0.011 |
| Brain | MRI | SubcorticalVolumes | 14 | 0.274±0.006 | 0.297±0.008 | 0.284±0.009 |
| Brain | MRI | dMRIWeightedMeans | 243 | 0.528±0.014 | 0.531±0.014 | 0.542±0.020 |
| Brain | MRI | AllScalars | 396 | 0.622±0.011 | 0.620±0.010 | 0.627±0.017 |
| Brain | Cognitive | ReactionTime | 14 | 0.112±0.004 | 0.158±0.003 | 0.145±0.003 |
| Brain | Cognitive | MatrixPatternCompletion | 4 | 0.136±0.011 | 0.138±0.010 | 0.133±0.011 |
| Brain | Cognitive | TowerRearranging | 2 | 0.083±0.009 | 0.084±0.009 | 0.074±0.018 |
| Brain | Cognitive | SymbolDigitSubstitution | 2 | 0.230±0.012 | 0.248±0.011 | 0.237±0.013 |
| Brain | Cognitive | PairedAssociativeLearning | 46 | 0.072±0.010 | 0.078±0.011 | 0.064±0.014 |
| Brain | Cognitive | ProspectiveMemory | 14 | 0.122±0.017 | 0.176±0.003 | 0.157±0.010 |
| Brain | Cognitive | NumericMemory | 3 | 0.045±0.021 | 0.092±0.006 | 0.089±0.007 |

| | | | | | | |
|---|---|---|---|---|---|---|
| Brain | Cognitive | FluidIntelligence | 37 | 0.045±0.003 | 0.048±0.003 | 0.045±0.002 |
| Brain | Cognitive | TrailMaking | 4 | 0.128±0.011 | 0.241±0.008 | 0.221±0.010 |
| Brain | Cognitive | PairsMatching | 10 | 0.082±0.004 | 0.138±0.003 | 0.134±0.004 |
| Brain | Cognitive | AllScalars | 135 | 0.340±0.014 | 0.378±0.012 | 0.308±0.060 |
| Brain | All | Scalars | 531 | 0.655±0.013 | 0.648±0.013 | 0.615±0.034 |
| Eyes | Autorefraction | Scalars | 35 | 0.163±0.006 | 0.289±0.005 | 0.247±0.008 |
| Eyes | Acuity | Scalars | 8 | 0.116±0.007 | 0.155±0.005 | 0.148±0.005 |
| Eyes | IntraocularPressure | Scalars | 8 | 0.062±0.004 | 0.069±0.004 | 0.065±0.004 |
| Eyes | All | Scalars | 51 | 0.250±0.004 | 0.358±0.006 | 0.308±0.024 |
| Hearing | HearingTest | Scalars | 126 | 0.255±0.007 | 0.311±0.009 | 0.310±0.008 |
| Lungs | Spirometry | Scalars | 3 | 0.221±0.004 | 0.256±0.005 | 0.254±0.004 |
| Arterial | BloodPressure | Scalars | 3 | 0.202±0.003 | 0.222±0.003 | 0.221±0.003 |
| Arterial | Carotids | Scalars | 12 | 0.183±0.015 | 0.206±0.015 | 0.201±0.018 |
| Arterial | PulseWaveAnalysis | Scalars | 8 | 0.129±0.003 | 0.229±0.003 | 0.197±0.072 |
| Arterial | All | Scalars | 23 | 0.313±0.013 | 0.368±0.014 | 0.353±0.021 |
| Heart | All | Scalars | 35 | 0.259±0.027 | 0.317±0.023 | 0.199±0.155 |
| Heart | MRI | Size | 8 | 0.009±0.028 | 0.127±0.010 | 0.086±0.047 |
| Heart | MRI | PulseWaveAnalysis | 15 | 0.171±0.020 | 0.130±0.027 | 0.199±0.018 |
| Heart | MRI | AllScalars | 23 | 0.070±0.061 | 0.267±0.018 | 0.246±0.040 |
| Heart | ECG | Scalars | 12 | 0.090±0.013 | 0.111±0.019 | 0.097±0.012 |
| Musculoskeletal | Scalars | Impedance | 5 | 0.020±0.001 | 0.042±0.002 | 0.041±0.005 |
| Musculoskeletal | Scalars | Anthropometry | 8 | 0.132±0.003 | 0.151±0.003 | 0.147±0.004 |
| Musculoskeletal | Scalars | HeelBoneDensitometry | 6 | 0.035±0.001 | 0.049±0.002 | 0.048±0.002 |
| Musculoskeletal | Scalars | HandGripStrength | 2 | 0.117±0.003 | 0.131±0.003 | 0.130±0.003 |
| Musculoskeletal | Scalars | AllScalars | 21 | 0.205±0.004 | 0.252±0.004 | 0.252±0.005 |
| PhysicalActivity | FullWeek | Scalars | 2271 | 0.556±0.024 | 0.594±0.006 | 0.503±0.048 |
| Biochemistry | Blood | Scalars | 28 | 0.302±0.005 | 0.476±0.005 | 0.474±0.004 |
| Biochemistry | Urine | Scalars | 4 | 0.081±0.003 | 0.104±0.003 | 0.096±0.003 |
| Biochemistry | All | Scalars | 32 | 0.315±0.010 | 0.491±0.007 | 0.478±0.008 |

| | | | | | | |
|---|---|---|---|---|---|---|
| BloodCells | BloodCount | Scalars | 31 | 0.058±0.004 | 0.122±0.002 | 0.119±0.003 |

**Supplementary Table 3: Pearson correlations between the feature importances for different scalar features-based algorithms**

| Main dimension | Subdimension | Sub-subdimension | CorrelationVersusElasticNet | CorrelationVersusLightGBM | CorrelationVersusNeuralNetwork | ElasticNetVersusLightGBM | ElasticNetVersusNeuralNetwork | LightGBMVersusNeuralNetwork |
|---|---|---|---|---|---|---|---|---|
| Brain | MRI | GreyMatterVolumes | 0.236 | 0.348 | 0.079 | 0.836 | 0.861 | 0.662 |
| Brain | MRI | SubcorticalVolumes | 0.656 | 0.699 | 0.024 | 0.72 | 0.095 | -0.048 |
| Brain | MRI | dMRIWeightedMeans | 0.11 | 0.119 | 0.173 | 0.428 | 0.74 | 0.308 |
| Brain | MRI | AllScalars | 0.107 | 0.037 | 0.085 | 0.427 | 0.753 | 0.296 |
| Brain | Cognitive | ReactionTime | 0.917 | 0.885 | 0.871 | 0.895 | 0.954 | 0.939 |
| Brain | Cognitive | MatrixPatternCompletion | 0.873 | 0.657 | 0.451 | 0.687 | 0.434 | 0.253 |
| Brain | Cognitive | TowerRearranging | 0.938 | 0.958 | 0.429 | 0.899 | 0.377 | 0.358 |
| Brain | Cognitive | SymbolDigitSubstitution | 0.901 | 0.982 | 0.629 | 0.841 | 0.754 | 0.544 |
| Brain | Cognitive | PairedAssociativeLearning | 0.695 | 0.568 | 0.4 | 0.473 | 0.33 | 0.495 |
| Brain | Cognitive | ProspectiveMemory | 0.884 | 0.738 | 0.265 | 0.862 | 0.221 | 0.318 |
| Brain | Cognitive | NumericMemory | 0.658 | 0.399 | 0.561 | 0.849 | 0.614 | 0.744 |
| Brain | Cognitive | FluidIntelligence | 0.635 | 0.139 | 0.515 | 0.42 | 0.522 | 0.187 |
| Brain | Cognitive | TrailMaking | 0.941 | 0.954 | 0.344 | 0.888 | 0.326 | 0.24 |
| Brain | Cognitive | PairsMatching | 0.771 | 0.783 | 0.727 | 0.736 | 0.477 | 0.411 |
| Brain | Cognitive | AllScalars | 0.737 | 0.722 | 0.665 | 0.707 | 0.904 | 0.551 |
| Brain | All | Scalars | 0.24 | 0.346 | 0.151 | 0.507 | 0.757 | 0.568 |
| Eyes | Autorefraction | Scalars | 0.71 | 0.23 | 0.198 | 0.453 | 0.146 | 0.188 |
| Eyes | Acuity | Scalars | 0.803 | 0.61 | 0.541 | 0.878 | 0.394 | 0.147 |
| Eyes | IntraocularPressure | Scalars | 0.4 | 0.296 | 0.633 | 0.521 | 0.888 | 0.389 |
| Eyes | All | Scalars | 0.647 | 0.226 | 0.496 | 0.434 | 0.408 | 0.124 |
| Hearing | HearingTest | Scalars | -0.217 | 0.181 | 0.079 | -0.04 | 0.245 | 0.559 |

| | | | | | | | | |
|---|---|---|---|---|---|---|---|---|
| Lungs | Spirometry | Scalars | 0.531 | 0.937 | 0.751 | 0.425 | 0.878 | 0.615 |
| Arterial | BloodPressure | Scalars | 0.671 | 0.38 | 0.721 | 0.821 | 0.724 | 0.505 |
| Arterial | Carotids | Scalars | 0.552 | 0.912 | -0.058 | 0.779 | 0.241 | 0.07 |
| Arterial | PWA | Scalars | 0.759 | 0.366 | 0.374 | 0.292 | 0.014 | 0.646 |
| Arterial | All | Scalars | 0.171 | 0.35 | -0.093 | 0.566 | 0.411 | 0.589 |
| Heart | All | Scalars | 0.396 | 0.478 | 0.445 | 0.662 | 0.813 | 0.581 |
| Heart | MRI | Size | NA | 0.453 | 0.433 | NA | NA | 0.434 |
| Heart | MRI | PWA | 0.607 | 0.648 | 0.415 | 0.655 | 0.868 | 0.56 |
| Heart | MRI | AllScalars | 0.871 | 0.536 | 0.073 | 0.361 | 0.02 | -0.156 |
| Heart | ECG | Scalars | 0.764 | 0.656 | 0.154 | 0.728 | 0.26 | 0.128 |
| Musculoskeletal | Scalars | Impedance | 0.535 | 0.043 | 0.72 | 0.385 | 0.344 | 0.032 |
| Musculoskeletal | Scalars | Anthropometry | 0.402 | 0.361 | 0.389 | 0.712 | 0.648 | 0.288 |
| Musculoskeletal | Scalars | HeelBoneDensitometry | 0.611 | 0.699 | 0.418 | 0.499 | 0.446 | 0.033 |
| Musculoskeletal | Scalars | HandGripStrength | 0.581 | 0.837 | 0.49 | 0.706 | 0.584 | 0.277 |
| Musculoskeletal | Scalars | AllScalars | 0.337 | 0.352 | 0.241 | 0.588 | 0.495 | 0.213 |
| Biochemistry | Blood | Scalars | 0.595 | 0.406 | 0.143 | 0.601 | 0.185 | -0.102 |
| Biochemistry | Urine | Scalars | 0.721 | 0.508 | 0.491 | 0.821 | 0.257 | 0.073 |
| Biochemistry | All | Scalars | 0.604 | 0.444 | 0.248 | 0.587 | 0.66 | 0.291 |
| Immune System | BloodCount | Scalars | 0.322 | 0.228 | 0.407 | 0.43 | 0.075 | -0.099 |
| Physical Activity | FullWeek | Scalars | 0.067 | 0.184 | 0.05 | 0.674 | 0.737 | 0.621 |
| **Average** | | | 0.568 | 0.504 | 0.369 | 0.618 | 0.497 | 0.337 |
| **Standard deviation** | | | 0.276 | 0.272 | 0.243 | 0.206 | 0.275 | 0.257 |

#### Supplementary Table 4: Spearman correlations between the feature importances for different scalar features-based algorithms

| Main dimension | Subdimension | Sub-subdimension | CorrelationVersusElasticNet | CorrelationVersusLightGBM | CorrelationVersusNeuralNetwork | ElasticNetVersusLightGBM | ElasticNetVersusNeuralNetwork | LightGBMVersusNeuralNetwork |
|---|---|---|---|---|---|---|---|---|
| Brain | MRI | GreyMatterVol | 0.27 | 0.388 | 0.151 | 0.714 | 0.836 | 0.548 |

| | | | | | | | | |
|---|---|---|---|---|---|---|---|---|
| | | umes | | | | | | |
| Brain | MRI | SubcorticalVolumes | 0.722 | 0.681 | 0.403 | 0.725 | 0.36 | 0.415 |
| Brain | MRI | dMRIWeightedMeans | 0.048 | 0.098 | 0.184 | 0.427 | 0.532 | 0.24 |
| Brain | MRI | AllScalars | 0.126 | 0.028 | 0.102 | 0.347 | 0.72 | 0.271 |
| Brain | Cognitive | ReactionTime | 0.7 | 0.535 | 0.732 | 0.58 | 0.731 | 0.818 |
| Brain | Cognitive | MatrixPatternCompletion | 0.647 | 0.531 | 0.659 | 0.464 | 0.312 | 0.54 |
| Brain | Cognitive | TowerRearranging | 0.611 | 0.603 | 0.681 | 0.383 | 0.352 | 0.89 |
| Brain | Cognitive | SymbolDigitSubstitution | 0.635 | 0.628 | 0.237 | 0.516 | 0.305 | 0.484 |
| Brain | Cognitive | PairedAssociativeLearning | 0.555 | 0.571 | 0.611 | 0.397 | 0.37 | 0.71 |
| Brain | Cognitive | ProspectiveMemory | 0.653 | 0.56 | 0.617 | 0.488 | 0.38 | 0.643 |
| Brain | Cognitive | NumericMemory | 0.833 | 0.652 | 0.684 | 0.581 | 0.616 | 0.826 |
| Brain | Cognitive | FluidIntelligence | 0.452 | 0.341 | 0.318 | 0.395 | 0.187 | 0.699 |
| Brain | Cognitive | TrailMaking | 0.641 | 0.666 | 0.346 | 0.566 | 0.293 | 0.432 |
| Brain | Cognitive | PairsMatching | 0.725 | 0.849 | 0.831 | 0.776 | 0.515 | 0.783 |
| Brain | Cognitive | AllScalars | 0.585 | 0.671 | 0.618 | 0.566 | 0.728 | 0.596 |
| Brain | All | Scalars | 0.307 | 0.411 | 0.271 | 0.563 | 0.688 | 0.53 |
| Eyes | Autorefraction | Scalars | 0.745 | 0.154 | 0.323 | 0.397 | 0.304 | 0.295 |
| Eyes | Acuity | Scalars | 0.663 | 0.715 | 0.663 | 0.704 | 0.476 | 0.741 |
| Eyes | IntraocularPressure | Scalars | 0.228 | 0.503 | 0.751 | 0.524 | 0.442 | 0.67 |
| Eyes | All | Scalars | 0.641 | 0.226 | 0.488 | 0.317 | 0.424 | 0.302 |
| Hearing | HearingTest | Scalars | -0.056 | 0.531 | 0.253 | 0.3 | 0.5 | 0.629 |
| Lungs | Spirometry | Scalars | 0.261 | 0.683 | 0.58 | 0.51 | 0.221 | 0.491 |
| Arterial | BloodPressure | Scalars | 0.233 | 0.462 | 0.227 | 0.592 | 0.295 | 0.516 |
| Arterial | Carotids | Scalars | 0.477 | 0.823 | 0.183 | 0.473 | 0.207 | 0.189 |
| Arterial | PWA | Scalars | 0.487 | 0.572 | 0.664 | 0.628 | 0.512 | 0.819 |
| Arterial | All | Scalars | 0.28 | 0.534 | 0.123 | 0.52 | 0.463 | 0.524 |
| Heart | All | Scalars | 0.247 | 0.58 | 0.673 | 0.555 | 0.403 | 0.554 |

| | | | | | | | | |
|---|---|---|---|---|---|---|---|---|
| Heart | MRI | Size | NA | 0.483 | 0.307 | NA | NA | 0.743 |
| Heart | MRI | PWA | 0.594 | 0.637 | 0.527 | 0.753 | 0.408 | 0.446 |
| Heart | MRI | AllScalars | 0.69 | 0.57 | 0.368 | 0.388 | 0.276 | 0.091 |
| Heart | ECG | Scalars | 0.606 | 0.67 | 0.554 | 0.558 | 0.315 | 0.415 |
| Musculoskeletal | Scalars | Impedance | 0.64 | 0.49 | 0.796 | 0.582 | 0.455 | 0.608 |
| Musculoskeletal | Scalars | Anthropometry | 0.329 | 0.563 | 0.588 | 0.685 | 0.523 | 0.739 |
| Musculoskeletal | Scalars | HeelBoneDensitometry | 0.526 | 0.732 | 0.457 | 0.638 | 0.232 | 0.357 |
| Musculoskeletal | Scalars | HandGripStrength | 0.461 | 0.693 | 0.545 | 0.557 | 0.477 | 0.703 |
| Musculoskeletal | Scalars | AllScalars | 0.331 | 0.513 | 0.384 | 0.566 | 0.567 | 0.618 |
| Biochemistry | Blood | Scalars | 0.491 | 0.412 | 0.344 | 0.792 | 0.294 | 0.316 |
| Biochemistry | Urine | Scalars | 0.411 | 0.446 | 0.595 | 0.697 | 0.379 | 0.441 |
| Biochemistry | All | Scalars | 0.532 | 0.519 | 0.722 | 0.818 | 0.712 | 0.711 |
| Immune System | BloodCount | Scalars | 0.354 | 0.271 | 0.585 | 0.394 | 0.241 | 0.228 |
| Physical Activity | FullWeek | Scalars | 0.161 | 0.303 | 0.211 | 0.362 | 0.264 | 0.265 |
| **Average** | | | 0.471 | 0.519 | 0.472 | 0.545 | 0.433 | 0.533 |
| **Standard deviation** | | | 0.213 | 0.182 | 0.208 | 0.138 | 0.167 | 0.202 |

**Supplementary Table 5: GWASs summary - Heritability, number of SNPs and genes associated with accelerated aging in each dimension**

| Aging dimension | Sample size | SNPs | Genes | Heritability (%) | R-Squared for CA prediction (%) |
|---|---|---|---|---|---|
| * | 214 | 0 | 0 | N/A | 66.1±0.2 |
| *instances01 | 238 | 0 | 0 | N/A | 87.0±0.4 |
| *instances1.5x | 73322 | 0 | 0 | 12.5±0.9 | 63.5±0.2 |
| *instances23 | 2287 | 0 | 0 | 20.1±22.7 | 90.1±0.4 |
| Abdomen | 32475 | 1 | 1 | 26.3±1.9 | 76.3±0.2 |
| AbdomenLiver | 40760 | 7 | 6 | 22.3±1.5 | 71.5±0.2 |

| | | | | | |
|---|---|---|---|---|---|
| AbdomenPancreas | 32548 | 0 | 0 | 22.1±1.9 | 70.3±0.3 |
| Arterial | 7549 | 0 | 0 | 32.6±7.3 | 67.1±0.6 |
| ArterialCarotids | 7885 | 0 | 0 | 28.6±7.0 | 64.8±0.6 |
| ArterialPulseWaveAnalysis | 184825 | 192 | 109 | 17.2±0.4 | 41.3±0.2 |
| Biochemistry | 68749 | 165 | 83 | 25.5±1.0 | 50.7±0.3 |
| BiochemistryBlood | 226714 | 1081 | 450 | 26.2±0.3 | 48.6±0.1 |
| BiochemistryUrine | 141595 | 24 | 12 | 10.5±0.5 | 10.4±0.1 |
| BloodCells | 440108 | 2636 | 1056 | 18.1±0.2 | 12.5±0.1 |
| Brain | 22951 | 219 | 25 | 35.9±2.6 | 76.4±0.3 |
| BrainCognitive | 30108 | 27 | 20 | 29.3±2.0 | 37.9±0.4 |
| BrainMRI | 35351 | 235 | 35 | 39.1±1.7 | 75.3±0.2 |
| Eyes | 51456 | 55 | 27 | 28.2±1.2 | 83.6±0.1 |
| EyesFundus | 73148 | 109 | 44 | 26.0±0.9 | 76.6±0.2 |
| EyesOCT | 75690 | 149 | 80 | 23.6±0.9 | 70.8±0.2 |
| Hearing | 181670 | 662 | 243 | 14.1±0.4 | 31.4±0.2 |
| Heart | 15829 | 3 | 3 | 35.2±3.6 | 85.6±0.2 |
| HeartECG | 20585 | 0 | 0 | 21.8±2.9 | 37.6±0.4 |
| HeartMRI | 32267 | 24 | 8 | 37.9±1.9 | 85.3±0.1 |
| Lungs | 354550 | 4286 | 1460 | 30.3±0.2 | 25.6±0.1 |
| Musculoskeletal | 35277 | 11 | 9 | 34.9±1.8 | 87.6±0.1 |
| MusculoskeletalFullBody | 38820 | 8 | 8 | 30.7±1.6 | 85.7±0.1 |
| MusculoskeletalHips | 38806 | 20 | 7 | 27.7±1.6 | 69.0±0.3 |
| MusculoskeletalKnees | 36727 | 12 | 6 | 25.3±1.7 | 69.0±0.3 |
| MusculoskeletalScalars | 407998 | 2631 | 954 | 22.1±0.2 | 25.9±0.1 |
| MusculoskeletalSpine | 37826 | 22 | 13 | 32.9±1.7 | 74.6±0.2 |
| PhysicalActivity | 73322 | 9 | 7 | 12.3±0.9 | 63.5±0.2 |

#### Supplementary Table 6: List of biomarkers by subcategories for the Biomarkers Wide Association Study [BWAS]

See supplementary data

#### Supplementary Table 7: Biomarkers most associated with accelerated aging across all aging dimensions

| X-subcategory | Variables |
|---|---|
| **BloodPressure (66.7%)** | Systolic blood pressure (.093±.083); Diastolic blood pressure (.067±.052); Pulse rate (.057±.022) |
| **AnthropometryImpedance (41.2%)** | Impedance of arm (right) (.051±.031); Impedance of leg (left) (.048±.037); Impedance of whole body (.052±.038) |
| **ArterialStiffness (40.5%)** | Position of the pulse wave peak (.068±.066); Position of the shoulder on the pulse waveform (.073±.065); Absence of notch position in the pulse waveform (.067±.071) |
| **HeartPWA (38.4%)** | Heart rate during PWA (.058±.021); Number of beats in waveform average for PWA (.058±.013); Central systolic blood pressure during PWA (.095±.058) |
| **UrineBiochemistry (31.1%)** | Microalbumin in urine (.041±.016); Sodium in urine (.035±.021); Creatinine (enzymatic) in urine (.031±.023) |
| **BloodCount (29.5%)** | Neutrophill count (.050±.023); White blood cell (leukocyte) count (.053±.023); Monocyte count (.044±.028) |
| **BloodBiochemistry (29.2%)** | Glycated hemoglobin (HbA1c) (.076±.050); Alkaline phosphatase (.063±.035); Cystatin C (.081±.089) |
| **AnthropometryBodySize (29.2%)** | Waist circumference (.087±.035); Body mass index (BMI) (.080±.028); Hip circumference (.064±.033) |
| **BraindMRIWeightedMeans (23.7%)** | Weighted-mean MD in tract superior thalamic radiation (left) (.083±.070); Weighted-mean L3 in tract anterior thalamic radiation (left) (.087±.077); Weighted-mean MD in tract superior thalamic radiation (right) (.078±.064) |
| **CognitiveTrailMaking (22.0%)** | Duration to complete alphanumeric path (trail #2) (.090±.099); Duration to complete numeric path (trail #1) (.105±.125); Total errors traversing alphanumeric path (trail #2) (.071±.060) |
| **ECGAtRest (18.6%)** | Ventricular rate (.062±.023); QTC interval (.068±.034); QRS num (.061±.019) |
| **EyeIntraocularPressure (18.6%)** | Intra-ocular pressure, Goldmann-correlated (right) (.055±.035); Corneal resistance factor (left) (.030±.014); Intra-ocular pressure, Goldmann-correlated (left) (.064±.041) |
| **CarotidUltrasound (18.4%)** | Maximum carotid IMT (intima-medial thickness) at 120 degrees (.096±.109); Maximum carotid IMT (intima-medial thickness) at 240 degrees (.112±.106); Maximum carotid IMT (intima-medial thickness) at 210 degrees (.110±.111) |
| **HearingTest (18.4%)** | Mean signal-to-noise ratio (SNR), (right).14 (.064±.093); Mean signal-to-noise ratio (SNR), (right).13 (.063±.092); Mean signal-to-noise ratio (SNR), (right).15 (.064±.094) |
| **EyeAcuity (15.9%)** | logMAR, final (left) (.075±.059); logMAR, final (right) (.080±.068); Glasses worn/required (right).wearing (.069±.067) |
| **CognitiveProspectiveMemory (14.5%)** | PM: initial answer.Blue square (.058±.049); Time to answer (.065±.052); Number of attempts (.054±.041) |
| **HeartSize (12.5%)** | Average heart rate (.062±.024); Body surface area (.064±.017); LV end systolic volume (.029±.003) |
| **CognitivePairsMatching (12.4%)** | Time to complete round.First round (.051±.053); Time to complete round.Second round (.083±.104); Number of incorrect matches in round.First round (.048±.040) |

| | |
|---|---|
| **EyeAutorefraction (10.9%)** | Cylindrical power (left) (.068±.046); 3mm regularity index (left) (.060±.070); Cylindrical power (right) (.069±.057) |
| **CognitiveReactionTime (10.6%)** | Mean time to correctly identify matches (.065±.088); Mean time to press snap-button (.071±.090) |
| **BoneDensitometryOfHeel (9.1%)** | Ankle spacing width (.058±.026) |
| **PhysicalActivity (8.5%)** | avg_interruptions_in_longest_sleep_V2 (.056±.040); avg_interruptions_in_longest_sleep_V1 (.056±.040); xyCov_min (.066±.046) |
| **CognitiveFluidIntelligence (8.0%)** | FI4 : positional arithmetic.Do not know (.043±.025); FI3 : word interpolation.Grow (.043±.021); FI1 : numeric addition test.Do not know (.025±.011) |
| **CognitiveMatrixPatternCompletion (5.1%)** | Mean time to solve puzzles (.123±.111); Number of puzzles viewed (.033±.014) |
| **CognitivePairedAssociativeLearning (4.8%)** | Word associated with "pretty".dress (.049±.036); Word associated with "huge".house (.037±.018); Word associated with "happy".clown (.071±.057) |
| **CognitiveNumericMemory (3.0%)** | Time to complete test (.053±.029) |
| **BrainGreyMatterVolumes (2.6%)** | Volume of gray matter in Caudate (right) (.067±.036); Volume of gray matter in Caudate (left) (.067±.036); Volume of gray matter in Pallidum (right) (.057±.033) |
| **Spirometry (1.0%)** | Forced vital capacity (FVC) (.048±nan) |
| **BrainSubcorticalVolumes (0.0%)** | |
| **HandGripStrength (0.0%)** | |
| **CognitiveSymbolDigitSubstitution (0.0%)** | |
| **CognitiveTowerRearranging (0.0%)** | |

#### Supplementary Table 8: Biomarkers most associated with accelerated aging for each aging dimension

See supplementary data

#### Supplementary Table 9: Biomarkers most associated with decelerated aging across all aging dimensions

| X-subcategory | Variables |
|---|---|
| **HandGripStrength (97.0%)** | Hand grip strength (left) (.088±.093); Hand grip strength (right) (.088±.094) |
| **Spirometry (79.8%)** | Peak expiratory flow (PEF) (.080±.098); Forced expiratory volume in 1-second (FEV1) (.113±.159); Forced vital capacity (FVC) (.124±.167) |
| **CognitiveSymbolDigitSubstitution (68.2%)** | Number of symbol digit matches made correctly (.100±.132); Number of symbol digit matches attempted (.103±.144) |

| | |
|---|---|
| **BoneDensitometryOfHeel (64.1%)** | Heel bone mineral density (BMD) (.070±.047); Heel bone mineral density (BMD) T-score (.068±.045); Heel quantitative ultrasound index (QUI) (.070±.047) |
| **BrainSubcorticalVolumes (42.2%)** | Volume of thalamus (right) (.082±.073); Volume of accumbens (right) (.070±.073); Volume of thalamus (left) (.088±.074) |
| **AnthropometryBodySize (39.8%)** | Seated height (.079±.093); Sitting height (.102±.109); Seating box height (.050±.053) |
| **BrainGreyMatterVolumes (38.0%)** | Volume of gray matter in Parahippocampal Gyrus, anterior division (right) (.059±.033); Volume of gray matter in Amygdala (right) (.075±.059); Volume of gray matter in X Cerebellum (left) (.065±.041) |
| **CognitiveNumericMemory (27.3%)** | Maximum digits remembered correctly (.068±.049); Number of rounds of numeric memory test performed (.060±.037); Time to complete test (.025±.007) |
| **AnthropometryImpedance (23.6%)** | Impedance of leg (left) (.046±.015); Impedance of leg (right) (.043±.014); Impedance of whole body (.037±.008) |
| **CognitiveMatrixPatternCompletion (23.2%)** | Number of puzzles correctly solved (.080±.071); Number of puzzles viewed (.132±.103); Mean time to solve puzzles (.038±.014) |
| **CognitiveTowerRearranging (19.7%)** | Number of puzzles correct (.092±.095); Number of puzzles attempted (.150±.105) |
| **HeartSize (18.6%)** | LV ejection fraction (.047±.015); Body surface area (.083±.051); LV stroke volume (.063±.042) |
| **BloodBiochemistry (17.9%)** | IGF-1 (.077±.065); Creatinine (.056±.028); Vitamin D (.032±.015) |
| **PhysicalActivity (14.8%)** | p625_max (.074±.044); p1_max (.078±.057); p2_max (.077±.052) |
| **ECGAtRest (14.4%)** | PP interval (.065±.020); RR interval (.066±.020); R axis (.069±.039) |
| **BraindMRIWeightedMeans (11.8%)** | Weighted-mean FA in tract posterior thalamic radiation (left) (.061±.046); Weighted-mean FA in tract anterior thalamic radiation (left) (.069±.050); Weighted-mean FA in tract anterior thalamic radiation (right) (.062±.041) |
| **UrineBiochemistry (9.8%)** | Creatinine (enzymatic) in urine (.150±.222); Potassium in urine (.074±.085); Sodium in urine (.277±.309) |
| **BloodCount (8.7%)** | Lymphocyte percentage (.033±.015); Platelet distribution width (.026±.009); Red blood cell (erythrocyte) count (.041±.015) |
| **CognitiveProspectiveMemory (7.4%)** | PM: initial answer.Orange circle (.059±.050); PM: final answer.Orange circle (.047±.034); Time to answer (.038±.012) |
| **CarotidUltrasound (6.8%)** | Mean carotid IMT (intima-medial thickness) at 150 degrees (.023±.004); Maximum carotid IMT (intima-medial thickness) at 150 degrees (.021±.005); Minimum carotid IMT (intima-medial thickness) at 240 degrees (.030±.006) |
| **EyeIntraocularPressure (6.8%)** | Intra-ocular pressure, Goldmann-correlated (right) (.030±.005); Intra-ocular pressure, corneal-compensated (right) (.034±.004); Intra-ocular pressure, corneal-compensated (left) (.032±.013) |
| **CognitiveFluidIntelligence (6.2%)** | Fluid intelligence score (.063±.052); FI3 : word interpolation.Adult (.050±.025); FI4 : positional arithmetic.6 (.055±.029) |
| **CognitivePairedAssociativeLearning (6.1%)** | Number of word pairs correctly associated (.059±.051); Word associated with "pretty".dish (.052±.048); Word associated with "new".plant (.057±.050) |
| **EyeAcuity (6.1%)** | Glasses worn/required (left).none (.072±.062); Glasses worn/required (right).none (.092±.068); Glasses worn/required (left).wearing (.029±.016) |
| **BloodPressure (6.1%)** | Diastolic blood pressure (.029±.012); Pulse rate (.030±.004); Systolic blood pressure (.023±nan) |
| **CognitivePairsMatching (6.1%)** | Number of correct matches in round.Second round (.030±.017); Number of correct matches in round.First round (.031±.016) |
| **HeartPWA (3.8%)** | End systolic pressure index during PWA (.047±.006); Diastolic brachial blood pressure during PWA (.086±.033); Mean arterial pressure during PWA (.069±.036) |
| **ArterialStiffness (3.8%)** | Pulse wave peak to peak time (.057±.065); Pulse rate (.036±.014); Position of the shoulder on |

| | |
|---|---|
| | the pulse waveform (.023±nan) |
| **EyeAutorefraction (3.4%)** | 6mm cylindrical power (left) (.029±.010); 6mm cylindrical power (right) (.027±.011); 3mm cylindrical power (right) (.024±.008) |
| **HearingTest (1.0%)** | Triplet correct (left).1 (.035±.031); Triplet correct (right).1 (.039±.036); Triplet correct (right).2 (.033±.028) |
| **CognitiveReactionTime (0.0%)** | |
| **CognitiveTrailMaking (0.0%)** | |

#### Supplementary Table 10: Biomarkers most associated with decelerated aging for each aging dimension

See supplementary data

#### Supplementary Table 11: List of clinical phenotypes by subcategories for the Clinical Phenotypes Wide Association Study [CWAS]

See supplementary data

#### Supplementary Table 12: Clinical phenotypes most associated with accelerated aging across all aging dimensions

| X-subcategory | Variables |
|---|---|
| **Breathing (81.8%)** | Shortness of breath walking on level ground (.062±.029); Wheeze or whistling in the chest in last year (.047±.033) |
| **ChestPain (59.8%)** | Chest pain or discomfort walking normally (.036±.013); Chest pain due to walking ceases when standing still (.041±.018); Chest pain or discomfort (.035±.016) |
| **GeneralHealth (47.3%)** | Long-standing illness, disability or infirmity (.073±.032); Overall health rating (.099±.036); Falls in the last year (.039±.015) |
| **Claudication (37.8%)** | Leg pain when walking uphill or hurrying (.052±.020); Leg pain on walking (.050±.019); Leg pain when walking normally (.044±.016) |
| **MentalHealth (27.3%)** | Fed-up feelings (.033±.012); Frequency of tiredness / lethargy in last 2 weeks (.046±.019); Frequency of depressed mood in last 2 weeks (.034±.012) |
| **GeneralPain (19.7%)** | Back pain for 3+ months (.032±.010); Knee pain for 3+ months (.032±.012); Hip pain for 3+ months (.031±.012) |
| **CancerScreening (19.7%)** | Ever had bowel cancer screening (.029±.008); Time since last prostate specific antigen (PSA) test (.028±.010); Most recent bowel cancer screening (.018±.005) |
| **Hearing (13.2%)** | Noisy workplace (.043±.021); Tinnitus severity/nuisance (.027±.012); Hearing difficulty/problems with background noise (.032±.018) |
| **Eyesight (6.6%)** | Eye problems/disorders.Diabetes related eye disease (.051±.023); Age when diabetes-related eye disease diagnosed (.037±.020); Other eye problems (.036±.020) |

| | |
|---|---|
| **SexualFactors (5.3%)** | Ever had same-sex intercourse (.017±.004); Age first had sexual intercourse (.019±nan) |
| **Mouth (3.5%)** | Mouth/teeth dental problems.Prefer not to answer (.016±.004); Mouth/teeth dental problems.Painful gums (.031±.000); Mouth/teeth dental problems.None of the above (.024±nan) |

#### Supplementary Table 13: Clinical phenotypes most associated with accelerated aging for each aging dimension

See supplementary data

#### Supplementary Table 14: Clinical phenotypes most associated with decelerated aging across all aging dimensions

| X-subcategory | Variables |
|---|---|
| **CancerScreening (18.2%)** | Most recent bowel cancer screening (.030±.008); Ever had prostate specific antigen (PSA) test (.028±.009); Ever had bowel cancer screening (.014±.003) |
| **SexualFactors (15.9%)** | Age first had sexual intercourse (.034±.014); Ever had same-sex intercourse (.023±nan); Lifetime number of sexual partners (.011±nan) |
| **GeneralHealth (11.0%)** | Weight change compared with 1 year ago.No - weigh about the same (.034±.014); Weight change compared with 1 year ago.Yes - gained weight (.042±.018); Weight change compared with 1 year ago.Yes - lost weight (.016±.001) |
| **Mouth (8.5%)** | Mouth/teeth dental problems.None of the above (.032±.015); Mouth/teeth dental problems.Bleeding gums (.048±nan); Mouth/teeth dental problems.Mouth ulcers (.008±nan) |
| **GeneralPain (3.9%)** | Pain type(s) experienced in last month.None of the above (.035±.009); Pain type(s) experienced in last month.Headache (.013±.001); Hip pain for 3+ months (.016±nan) |
| **Breathing (3.0%)** | Shortness of breath walking on level ground (.025±nan); Wheeze or whistling in the chest in last year (.032±nan) |
| **MentalHealth (2.6%)** | Illness, injury, bereavement, stress in last 2 years.None of the above (.029±.010); Risk taking (.031±.015); Manic/hyper symptoms.None of the above (.043±.016) |
| **Eyesight (2.4%)** | Eye problems/disorders.None of the above (.049±.019); Reason for glasses/contact lenses.For short-sightedness, i.e. only or mainly for distance viewing such as driving, cinema etc (called myopia) (.041±.033); Age started wearing glasses or contact lenses (.026±.012) |
| **Hearing (2.2%)** | Tinnitus.No, never (.022±.010); Tinnitus.Yes, now most or all of the time (.021±.007); Noisy workplace (.031±nan) |
| **Claudication (1.6%)** | Leg pain on walking : effect of standing still.Pain usually disappears in less than 10 minutes (.026±.007); Leg pain on walking : action taken (.026±nan); Leg pain on walking (.027±nan) |
| **ChestPain (1.5%)** | Chest pain or discomfort (.018±nan); Chest pain due to walking ceases when standing still (.011±nan) |

#### Supplementary Table 15: Clinical phenotypes most associated with decelerated aging for each aging dimension

See supplementary data

**Supplementary Table 16: List of diseases by subcategories for the Diseases Wide Association Study [DWAS]**

See supplementary data

**Supplementary Table 17: Diseases most associated with accelerated aging across all aging dimensions**

| X-subcategory | Variables |
|---|---|
| **medical_diagnoses _I (18.9%)** | I10 Essential (primary) hypertension (.074±.031); I25 Chronic ischemic heart disease (.040±.017); I48 Atrial fibrillation and flutter (.040±.013) |
| **medical_diagnoses _Z (14.2%)** | Z86 Personal history of certain other diseases (.045±.016); Z72 Problems related to lifestyle (.035±.012); Z92 Personal history of medical treatment (.043±.017) |
| **medical_diagnoses _R (14.2%)** | R07 Pain in throat and chest (.033±.012); R06 Abnormalities of breathing (.025±.009); R10 Abdominal and pelvic pain (.024±.009) |
| **medical_diagnoses _K (12.5%)** | K29 Gastritis and duodenitis (.027±.008); K52 Other non-infective gastro-enteritis and colitis (.024±.009); K44 Diaphragmatic hernia (.029±.010) |
| **medical_diagnoses _M (10.2%)** | M06 Other rheumatoid arthritis (.029±.010); M54 Dorsalgia (.027±.011); M79 Other soft tissue disorders, not elsewhere classified (.028±.012) |
| **medical_diagnoses _J (9.3%)** | J44 Other chronic obstructive pulmonary disease (.038±.018); J45 Asthma (.032±.022); J22 Unspecified acute lower respiratory infection (.028±.012) |
| **medical_diagnoses _E (8.8%)** | E78 Disorders of lipoprotein metabolism and other lipidaemias (.044±.022); E11 Non-insulin-dependent diabetes mellitus (.061±.027); E87 Other disorders of fluid, electrolyte and acid-base balance (.031±.012) |
| **medical_diagnoses _G (8.1%)** | G56 Mononeuropathies of upper limb (.022±.010); G63 Polyneuropathy in diseases classified elsewhere (.021±.006); G40 Epilepsy (.027±.011) |
| **medical_diagnoses _F (7.2%)** | F17 Mental and behavioural disorders due to use of tobacco (.046±.013); F10 Mental and behavioural disorders due to use of alcohol (.034±.010); F32 Depressive episode (.035±.011) |
| **medical_diagnoses _N (6.7%)** | N17 Acute renal failure (.034±.014); N18 Chronic renal failure (.037±.019); N39 Other disorders of urinary system (.023±.008) |
| **medical_diagnoses _H (4.1%)** | H36 Retinal disorders in diseases classified elsewhere (.033±.012); H35 Other retinal disorders (.018±.006); H40 Glaucoma (.023±.011) |
| **medical_diagnoses _D (3.6%)** | D64 Other anaemias (.032±.012); D50 Iron deficiency anaemia (.026±.009); D69 Purpura and other haemorrhagic conditions (.019±.008) |
| **medical_diagnoses _T (3.4%)** | T82 Complications of cardiac and vascular prosthetic devices, implants and grafts (.023±.007); T81 Complications of procedures, not elsewhere classified (.029±.009); T43 Poisoning by psychotropic drugs, not elsewhere classified (.016±.003) |
| **medical_diagnoses _L (3.1%)** | L97 Ulcer of lower limb, not elsewhere classified (.023±.007); L03 Cellulitis (.026±.010); L40 Psoriasis (.019±.004) |
| **medical_diagnoses _Y (2.6%)** | Y83 Surgical operation and other surgical procedures as the cause of abnormal reaction of the patient, or of later complication, without mention of misadventure at the time of the procedure (.033±.011); Y84 Other medical procedures as the cause of abnormal reaction of the patient, or of later complication, without mention of misadventure at the time of the procedure (.017±.002); Y95 Nosocomial condition (.019±.003) |
| **medical_diagnoses _U (2.5%)** | U82 Resistance to betalactam antibiotics (.007±.001); U80 Agent resistant to penicillin and related antibiotics (.012±.001); U83 Resistance to other antibiotics (.007±.001) |

| | |
|---|---|
| **medical_diagnoses_C (2.4%)** | C79 Secondary malignant neoplasm of other sites (.015±.004); C34 Malignant neoplasm of bronchus and lung (.017±.010); C50 Malignant neoplasm of breast (.025±.012) |
| **medical_diagnoses_X (2.1%)** | X61 Intentional self-poisoning by and exposure to antiepileptic, sedative-hypnotic, anti-Parkinsonism and psychotropic drugs, not elsewhere classified (.017±.003); X62 Intentional self-poisoning by and exposure to narcotics and psychodysleptics [hallucinogens], not elsewhere classified (.016±.005); X44 Accidental poisoning by and exposure to other and unspecified drugs, medicaments and biological substances (.012±.004) |
| **medical_diagnoses_S (1.9%)** | S00 Superficial injury of head (.018±.004); S09 Other and unspecified injuries of head (.015±.002); S01 Open wound of head (.018±.003) |
| **medical_diagnoses_W (1.6%)** | W19 Unspecified fall (.022±.007); W01 Fall on same level from slipping, tripping and stumbling (.019±.006); W18 Other fall on same level (.014±.004) |
| **medical_diagnoses_A (1.4%)** | A41 Other septicaemia (.022±.008); A09 Diarrhoea and gastro-enteritis of presumed infectious origin (.023±.004); A08 Viral and other specified intestinal infections (.011±.002) |
| **medical_diagnoses_Q (1.1%)** | Q61 Cystic kidney disease (.020±.011); Q21 Congenital malformations of cardiac septa (.010±.001); Q85 Phakomatoses, not elsewhere classified (.009±.001) |
| **medical_diagnoses_O (1.0%)** | O99 Other maternal diseases classifiable elsewhere but complicating pregnancy, childbirth and the puerperium (.014±.005); O63 Long labor (.011±.001); O70 Perineal laceration during delivery (.016±.004) |
| **medical_diagnoses_B (0.1%)** | B95.6 Staphylococcus aureus as the cause of diseases classified to other chapters (.021±.003); B96.5 Pseudomonas (aeruginosa)(mallei)(pseudomallei) as the cause of diseases classified to other chapters (.019±.001); B96.2 Escherichia coli [E. coli] as the cause of diseases classified to other chapters (.023±.003) |
| **medical_diagnoses_V (0.1%)** | V03 Pedestrian injured in collision with car, pick-up truck or van (.010±.002); V43 Car occupant injured in collision with car, pick-up truck or van (.008±nan) |
| **medical_diagnoses_P (0.0%)** | |

#### Supplementary Table 18: Diseases most associated with accelerated aging for each aging dimension

See supplementary data

#### Supplementary Table 19: Diseases most associated with decelerated aging across all aging dimensions

| X-subcategory | Variables |
|---|---|
| **medical_diagnoses_O (2.8%)** | O70 Perineal laceration during delivery (.027±.010); O80 Single spontaneous delivery (.021±.005); O63 Long labor (.018±.005) |
| **medical_diagnoses_Z (0.9%)** | Z37 Outcome of delivery (.038±.011); Z34 Supervision of normal pregnancy (.014±.004); Z39 Postpartum care and examination (.009±.001) |
| **medical_diagnoses_K (0.3%)** | K40 Inguinal hernia (.009±.003); K57 Diverticular disease of intestine (.012±nan); K80 Cholelithiasis (.015±nan) |
| **medical_diagnoses_M (0.2%)** | M23 Internal derangement of knee (.009±.002); M81 Osteoporosis without pathological fracture (.007±nan); M17 Gonarthrosis [arthrosis of knee] (.017±nan) |
| **medical_diagnoses_N (0.1%)** | N97 Female infertility (.015±.003); N86 Erosion and ectropion of cervix uteri (.009±nan); N40 Hyperplasia of prostate (.015±nan) |

| | |
|---|---|
| **medical_diagnoses_C (0.1%)** | C61 Malignant neoplasm of prostate (.013±.007); C50 Malignant neoplasm of breast (.012±nan); C43 Malignant melanoma of skin (.006±nan) |
| **medical_diagnoses_I (0.1%)** | I83 Varicose veins of lower extremities (.010±.003); I84 Haemorrhoids (.013±nan) |
| **medical_diagnoses_V (0.1%)** | V18 Pedal cyclist injured in noncollision transport accident (.016±.007); V13 Pedal cyclist injured in collision with car, pick-up truck or van (.012±nan) |
| **medical_diagnoses_D (0.1%)** | D56 Thalassaemia (.009±nan); D22 Melanocytic naevi (.009±nan) |
| **medical_diagnoses_L (0.0%)** | L82 Seborrhoeic keratosis (.007±nan) |
| **medical_diagnoses_E (0.0%)** | E66 Obesity (.017±nan) |
| **medical_diagnoses_F (0.0%)** | F64 Gender identity disorders (.010±nan) |
| **medical_diagnoses_W (0.0%)** | W02 Fall involving ice-skates, skis, roller-skates or skateboards (.008±nan) |
| **medical_diagnoses_B (0.0%)** | |
| **medical_diagnoses_G (0.0%)** | |
| **medical_diagnoses_H (0.0%)** | |
| **medical_diagnoses_X (0.0%)** | |
| **medical_diagnoses_J (0.0%)** | |
| **medical_diagnoses_U (0.0%)** | |
| **medical_diagnoses_Y (0.0%)** | |
| **medical_diagnoses_P (0.0%)** | |
| **medical_diagnoses_Q (0.0%)** | |
| **medical_diagnoses_R (0.0%)** | |
| **medical_diagnoses_S (0.0%)** | |
| **medical_diagnoses_T (0.0%)** | |
| **medical_diagnoses_A (0.0%)** | |

##### Supplementary Table 20: Diseases most associated with decelerated aging for each aging dimension

See supplementary data

##### Supplementary Table 21: List of family history variables by subcategories for the Family History Phenotypes Wide Association Study [FWAS]

See supplementary data

#### Supplementary Table 22: Family history variables most associated with accelerated aging for each aging dimension

| Aging dimension | X-subcategory | Variables |
|---|---|---|
| **Abdomen** | FamilyHistory (1.1%) | |
| **AbdomenLiver** | FamilyHistory (1.1%) | |
| **AbdomenPancreas** | FamilyHistory (0.0%) | |
| **Arterial** | FamilyHistory (0%) | |
| **ArterialCarotids** | FamilyHistory (0%) | |
| **ArterialPulseWaveAnalysis** | FamilyHistory (0.0%) | |
| **Biochemistry** | FamilyHistory (2.3%) | |
| **BiochemistryBlood** | FamilyHistory (3.3%) | Illnesses of mother.Diabetes (.043) |
| **BiochemistryUrine** | FamilyHistory (0.0%) | |
| **Brain** | FamilyHistory (0.0%) | |
| **BrainCognitive** | FamilyHistory (6.5%) | Illnesses of father.Lung cancer (.100); Illnesses of adopted mother.Lung cancer (.096); Illnesses of adopted siblings.Stroke (.073) |
| **BrainMRI** | FamilyHistory (1.1%) | |
| **Eyes** | FamilyHistory (3.4%) | |
| **EyesAll** | FamilyHistory (3.3%) | |
| **EyesFundus** | FamilyHistory (0.0%) | |
| **EyesOCT** | FamilyHistory (0.0%) | |
| **Hearing** | FamilyHistory (2.2%) | Illnesses of siblings.Prefer not to answer1 (.014); Illnesses of siblings.Severe depression (.010) |
| **Heart** | FamilyHistory (0%) | |
| **HeartECG** | FamilyHistory (1.1%) | |
| **HeartMRI** | FamilyHistory (0%) | |
| **ImmuneSystem** | FamilyHistory (1.1%) | Illnesses of mother.Alzheimer's disease/dementia (.015) |
| **Lungs** | FamilyHistory (6.5%) | Illnesses of father.Chronic bronchitis/emphysema (.121); Illnesses of mother.Chronic bronchitis/emphysema (.092); Illnesses of adopted mother.Diabetes (.054) |
| **Musculoskeletal** | FamilyHistory (0.0%) | |
| **MusculoskeletalFullBody** | FamilyHistory (1.1%) | |
| **MusculoskeletalHips** | FamilyHistory (1.1%) | |

| | | |
|---|---|---|
| **MusculoskeletalKnees** | FamilyHistory (0.0%) | |
| **MusculoskeletalScalars** | FamilyHistory (3.3%) | Illnesses of siblings.Prefer not to answer1 (.013); Illnesses of siblings.Severe depression (.011); Illnesses of mother.Severe depression (.009) |
| **MusculoskeletalSpine** | FamilyHistory (0.0%) | |
| **PhysicalActivity** | FamilyHistory (1.4%) | |

#### Supplementary Table 23: Family history variables most associated with decelerated aging for each aging dimension

| Aging dimension | X-subcategory | Variables |
|---|---|---|
| **Abdomen** | FamilyHistory (0.0%) | |
| **AbdomenLiver** | FamilyHistory (0.0%) | |
| **AbdomenPancreas** | FamilyHistory (1.1%) | |
| **Arterial** | FamilyHistory (0%) | |
| **ArterialCarotids** | FamilyHistory (0%) | |
| **ArterialPulseWaveAnalysis** | FamilyHistory (1.1%) | Illnesses of mother.Alzheimer's disease/dementia (.023) |
| **Biochemistry** | FamilyHistory (2.3%) | |
| **BiochemistryBlood** | FamilyHistory (2.2%) | Illnesses of father.Prostate cancer (.012) |
| **BiochemistryUrine** | FamilyHistory (1.1%) | |
| **Brain** | FamilyHistory (1.1%) | |
| **BrainCognitive** | FamilyHistory (1.1%) | |
| **BrainMRI** | FamilyHistory (1.1%) | Illnesses of father.Prostate cancer (.032) |
| **Eyes** | FamilyHistory (1.1%) | |
| **EyesAll** | FamilyHistory (0.0%) | |
| **EyesFundus** | FamilyHistory (3.3%) | |
| **EyesOCT** | FamilyHistory (3.3%) | |
| **Hearing** | FamilyHistory (6.5%) | Illnesses of adopted siblings.Stroke (.126); Illnesses of adopted mother.Diabetes (.108); Illnesses of adopted father.Lung cancer (.061) |
| **Heart** | FamilyHistory (0%) | |
| **HeartECG** | FamilyHistory (0.0%) | |
| **HeartMRI** | FamilyHistory (0%) | |
| **ImmuneSystem** | FamilyHistory (3.3%) | Illnesses of adopted siblings.Stroke (.130); Illnesses of father.Do not know (.076) |

| | | |
|---|---|---|
| **Lungs** | FamilyHistory (2.2%) | Illnesses of mother.Alzheimer's disease/dementia (.022); Illnesses of father.Prostate cancer (.012) |
| **Musculoskeletal** | FamilyHistory (1.1%) | |
| **MusculoskeletalFullBody** | FamilyHistory (0.0%) | |
| **MusculoskeletalHips** | FamilyHistory (1.1%) | Illnesses of father.Prostate cancer (.027) |
| **MusculoskeletalKnees** | FamilyHistory (3.3%) | Illnesses of father.Prostate cancer (.025); Illnesses of mother.Severe depression (.025) |
| **MusculoskeletalScalars** | FamilyHistory (6.5%) | Illnesses of father.Lung cancer (.141); Illnesses of adopted mother.Chronic bronchitis/emphysema (.124); Illnesses of adopted father.Lung cancer (.121) |
| **MusculoskeletalSpine** | FamilyHistory (1.1%) | |
| **PhysicalActivity** | FamilyHistory (0.0%) | |

#### Supplementary Table 24: List of environmental variables by subcategories for the Environmental Wide Association Study [EWAS]

See supplementary data

#### Supplementary Table 25: Environmental variables most associated with accelerated aging across all aging dimensions

| X-subcategory | Variables |
|---|---|
| **Smoking (36.4%)** | Current tobacco smoking.Yes, on most or all days (.056±.023); Pack years adult smoking as proportion of life span exposed to smoking (.084±.031); Difficulty not smoking for 1 day (.058±.023) |
| **Sleep (24.7%)** | Nap during day (.047±.025); Daytime dozing / sleeping (narcolepsy) (.039±.016); Sleeplessness / insomnia (.038±.016) |
| **SunExposure (19.7%)** | Facial ageing.About your age (.032±.014); Time spend outdoors in summer (.046±.014); Facial ageing.Older than you are (.025±.007) |
| **Alcohol (15.9%)** | Average weekly beer plus cider intake (.043±.021); Reason for reducing amount of alcohol drunk.Illness or ill health (.045±.017); Average weekly spirits intake (.035±.011) |
| **EarlyLifeFactors (12.2%)** | Comparative height size at age 10.Taller (.044±.012); Comparative body size at age 10.Plumper (.028±.013); Breastfed as a baby.Do not know (.024±.008) |
| **PhysicalActivityQuestionnaire (11.6%)** | Types of physical activity in last 4 weeks.None of the above (.039±.017); Time spent watching television (TV) (.061±.033); Types of transport used (excluding work).Public transport (.052±.014) |
| **Diet (11.3%)** | Major dietary changes in the last 5 years.Yes, because of illness (.047±.021); Bread type.White (.041±.020); Processed meat intake (.037±.011) |
| **Medication (8.4%)** | Medication for cholesterol, blood pressure or diabetes.Cholesterol lowering medication (.060±.036); Medication for pain relief, constipation, heartburn.Aspirin (.046±.021); Medication for cholesterol, blood pressure or diabetes.Blood pressure medication (.045±.019) |
| **ElectronicDevices (7.9%)** | Plays computer games (.029±.010); Usual side of head for mobile phone use.Left (.018±.006); Weekly usage of mobile phone in last 3 months (.011±.001) |

#### Supplementary Table 26: Environmental variables most associated with accelerated aging for each aging dimension

See supplementary data

#### Supplementary Table 27: Environmental variables most associated with decelerated aging across all aging dimensions

| X-subcategory | Variables |
|---|---|
| **PhysicalActivityQuestionnaire (34.6%)** | Usual walking pace (.070±.037); Drive faster than motorway speed limit (.053±.029); Duration of other exercises (.053±.024) |
| **Smoking (24.9%)** | Time from waking to first cigarette (.058±.023); Age started smoking in current smokers (.057±.023); Current tobacco smoking.No (.054±.021) |
| **Sleep (13.0%)** | Snoring (.030±.010); Getting up in morning (.031±.012); Sleep duration (.020±.005) |
| **ElectronicDevices (11.8%)** | Hands-free device/speakerphone use with mobile phone in last 3 month (.032±.020); Length of mobile phone use (.044±.026); Difference in mobile phone use compared to two years previously (.035±.018) |
| **Medication (10.5%)** | Medication for cholesterol, blood pressure or diabetes.None of the above (.094±.042); Medication for cholesterol, blood pressure, diabetes, or take exogenous hormones.None of the above (.057±.027); Medication for pain relief, constipation, heartburn.None of the above (.041±.017) |
| **Diet (9.3%)** | Major dietary changes in the last 5 years.No (.034±.012); Cereal intake (.047±.020); Never eat eggs, dairy, wheat, sugar.I eat all of the above (.036±.014) |
| **Alcohol (9.1%)** | Alcohol usually taken with meals (.041±.018); Alcohol drinker status (.035±.011); Average weekly red wine intake (.039±.014) |
| **SunExposure (8.9%)** | Facial ageing.Younger than you are (.045±.023); Skin colour (.026±.013); Hair colour (natural, before graying).Dark brown (.018±.006) |
| **EarlyLifeFactors (7.6%)** | Comparative height size at age 10.Shorter (.039±.012); Breastfed as a baby.Yes (.031±.010); Comparative body size at age 10.About average (.024±.008) |

#### Supplementary Table 28: Environmental variables most associated with decelerated aging for each aging dimension

See supplementary data

#### Supplementary Table 29: List of socioeconomic variables by subcategories for the Socioeconomics Wide Association Study [SWAS]

See supplementary data

#### Supplementary Table 30: Socioeconomic variables most associated with accelerated aging across all aging dimensions

| X-subcategory | Variables |
|---|---|
| **OtherSociodemographics (16.5%)** | Private healthcare (.035±.011); Attendance/disability/mobility allowance.Attendance allowance (.026±.008); Attendance/disability/mobility allowance.Do not know (.016±.005) |
| **SocialSupport (13.9%)** | Leisure/social activities.None of the above (.029±.008); Leisure/social activities.Pub or social club (.034±.014); Leisure/social activities.Religious group (.065±.023) |
| **Education (11.0%)** | Qualifications.None of the above (.056±.031); Qualifications.CSEs or equivalent (.039±.012); Qualifications.Prefer not to answer (.021±.008) |
| **Household (8.5%)** | Own or rent accommodation lived in.Rent - from local authority, local council, housing association (.048±.020); Type of accommodation lived in.A flat, maisonette or apartment (.035±.010); Gas or solid-fuel cooking/heating.A gas fire that you use regularly in winter time (.039±.014) |
| **Employment (5.0%)** | Job involves night shift work (.026±.007); Job involves heavy manual or physical work (.025±.014); Current employment status.Prefer not to Answer (.014±.004) |

#### Supplementary Table 31: Socioeconomic variables most associated with accelerated aging for each aging dimension

See supplementary data

#### Supplementary Table 32: Socioeconomic variables most associated with decelerated aging across all aging dimensions

| X-subcategory | Variables |
|---|---|
| **Employment (16.4%)** | Current employment status.In paid employment or self-employed (.061±.057); Length of working week for main job (.065±.068); Frequency of travelling from home to job workplace (.057±.068) |
| **Education (16.3%)** | Qualifications.College or University degree (.059±.021); Qualifications.A levels/AS levels or equivalent (.044±.013); Qualifications.O levels/GCSEs or equivalent (.034±.015) |
| **SocialSupport (13.9%)** | Leisure/social activities.Sports club or gym (.051±.024); Able to confide (.026±.011); Leisure/social activities.Pub or social club (.048±.003) |
| **OtherSociodemographics (11.7%)** | Attendance/disability/mobility allowance.None of the above (.056±.025) |
| **Household (10.0%)** | Average total household income before tax (.079±.037); Number of vehicles in household (.052±.019); Number in household (.039±.014) |

#### Supplementary Table 33: Socioeconomic variables most associated with decelerated aging for each aging dimension

See supplementary data

**Supplementary Table 34: Image sizes after resizing**

| Main dimension | Subdimension | Sub-subdimension | Size before resizing | Size after resizing |
|---|---|---|---|---|
| Brain | MRI | SagittalRaw | 88, 88 | 316, 316 |
| | | SagittalReference | 88, 88 | 316, 316 |
| | | CoronalRaw | 88, 88 | 316, 316 |
| | | CoronalReference | 88, 88 | 316, 316 |
| | | TransverseRaw | 88, 88 | 316, 316 |
| | | TransverseReference | 88, 88 | 316, 316 |
| Eyes | Fundus | Raw | 1388, 1388 | 316, 316 |
| | OCT | Raw | 500, 512 | 312, 320 |
| Arterial | Carotids | Mixed | 505, 436 | 337, 291 |
| | | LongAxis | 505, 436 | 337, 291 |
| | | CIMT120 | 505, 436 | 337, 291 |
| | | CIMT150 | 505, 436 | 337, 291 |
| | | ShortAxis | 505, 436 | 337, 291 |
| Heart | MRI | 2chambersRaw | 200, 200 | 316, 316 |
| | | 2chambersContrast | 200, 200 | 316, 316 |
| | | 3chambersRaw | 200, 200 | 316, 316 |
| | | 3chambersContrast | 200, 200 | 316, 316 |
| | | 4chambersRaw | 200, 200 | 316, 316 |
| | | 4chambersContrast | 200, 200 | 316, 316 |
| | | 3chambersRawVideo | 200, 200 | 150, 150 |
| | | 4chambersRawVideo | 200, 200 | 150, 150 |
| | | 34chambersRawVideo | 200, 200 | 150, 150 |

| | | | | |
|---|---|---|---|---|
| Abdomen | Liver | Raw | 288, 364 | 288, 364 |
| | | Contrast | 288, 364 | 288, 364 |
| | Pancreas | Raw | 288, 350 | 288, 350 |
| | | Contrast | 288, 350 | 288, 350 |
| Musculoskeletal | Spine | Sagittal | 1513, 684 | 466, 211 |
| | | Coronal | 724, 720 | 315, 313 |
| | Hips | MRI | 626, 680 | 329, 303 |
| | Knees | MRI | 851, 700 | 347, 286 |
| | FullBody | Mixed | 811, 272 | 541, 181 |
| | | Figure | 811, 272 | 541, 181 |
| | | Skeleton | 811, 272 | 541, 181 |
| | | Flesh | 811, 272 | 541, 181 |
| PhysicalActivity | FullWeek | GramianAngularField1minDifference | 316, 316 | 316, 316 |
| | | GramianAngularField1minSummation | 316, 316 | 316, 316 |
| | | MarkovTransitionField1min | 316, 316 | 316, 316 |
| | | RecurrencePlots1min | 316, 316 | 316, 316 |

**Supplementary Table 35: Scalar features generated from the wrist accelerometer data**

See supplementary data

**Supplementary Table 36: Physical activity scalar features outputted by UKB's software and removed from our list of predictors because non-biological**

See supplementary data

**Supplementary Table 37: Data augmentation hyperparameters for images**

| Dataset | Rotation range | Horizontal shift percentage range | Vertical shift percentage range | Zoom range |
|---|---|---|---|---|
| Brain MRI | 10 | 0.05 | 0.1 | 0 |
| Eye Fundus | 20 | 0.02 | 0.02 | 0 |
| Eye OCT | 30 | 0.1 | 0.2 | 0 |
| Arterial Carotids | 0 | 0.2 | 0 | 0 |
| Heart MRI | 10 | 0.1 | 0.1 | 0 |
| Abdomen Liver | 10 | 0.1 | 0.1 | 0 |
| Abdomen Pancreas | 10 | 0.1 | 0.1 | 0 |
| Musculoskeletal Spine | 10 | 0.1 | 0.1 | 0 |
| Musculoskeletal Hips | 10 | 0.1 | 0.1 | 0.1 |
| Musculoskeletal Knees | 10 | 0.1 | 0.1 | 0.1 |
| Musculoskeletal FullBody | 10 | 0.05 | 0.02 | 0 |
| PhysicalActivity FullWeek | 0 | 0 | 0 | 0 |
| Heart MRI (Videos) | 0 | 0.1 | 0.1 | 0 |

**Supplementary Table 38: Hyperparameter space for scalar features-based models Bayesian optimization**

| **Algorithm** | **Hyperparameter** | **Scale** | **Low** | **High** |
|---|---|---|---|---|
| Elastic net | alpha | loguniform | -10 | 0 |
| | l1_ratio | uniform | 0 | 1 |
| Gradient Boosted Machine | num_leaves | quniform | 5 | 45 |
| | min_child_samples | quniform | 100 | 500 |
| | min_child_weight | loguniform | -5 | 4 |
| | subsample | uniform | 0.2 | 0.8 |
| | colsample | uniform | 0.4 | 0.6 |
| | reg_alpha | loguniform | -2 | 2 |

| | | | | |
|---|---|---|---|---|
| | reg_lambda | loguniform | -2 | 2 |
| | n_estimators | quniform | 150 | 450 |
| Neural network | learning_rate_init | loguniform | -5 | -1 |
| | apha | loguniform | -6 | 3 |

**Supplementary Table 39: Architecture of the three-dimensional convolutional neural network trained on heart MRI videos**

| Layer type | Kernel size | | Stride size | | Elements |
|---|---|---|---|---|---|
| | *Time* | *Space* | *Time* | *Space* | |
| Conv 3D | 3 | (3, 3) | 1 | (1, 1) | 16 |
| Max Pooling | 1 | (2, 2) | 1 | (2, 2) | NA |
| Conv 3D | 3 | (3, 3) | 1 | (1, 1) | 64 |
| Max Pooling | 1 | (5,5) | 1 | (2, 2) | NA |
| Conv 3D | 5 | (5,5) | 1 | (1, 1) | 512 |
| Max Pooling | 5 | (5,5) | 1 | (2, 2) | NA |
| Conv 3D | 5 | (7,7) | 1 | (1, 1) | 1024 |
| Max Pooling | 1 | (5,5) | 1 | (2,2) | NA |
| Batch Norm | NA | | | | |
| Fully Connected | NA | 1024 | NA | NA | 1024 |
| Dropout | NA | | | | |
| Fully Connected | NA | 1 | NA | NA | 1 |

**Supplementary Table 40: Nested Cross-Validation pipeline**

**10x9-fold Nested Cross-Validation [CV]**
**10-fold Outer Cross-Validation; 9-fold Inner Cross-Validation**
**(N_CV_inner = N_CV_outer -1)**

| | Data Fold | N=100 |
|---|---|---|
| **INITIAL DATA SPLIT** | F0 | N=10 |
| | F1 | N=10 |
| | F2 | N=10 |
| | F3 | N=10 |
| | F4 | N=10 |
| | F5 | N=10 |
| | F6 | N=10 |
| | F7 | N=10 |
| | F8 | N=10 |
| | F9 | N=10 |

**OUTER CROSS-VALIDATION FOLD 0**

| Data Fold | Outer CV folds 0 | | Inner CV folds 0 | | Inner CV folds 1 | | Inner CV folds 2 | | Inner CV folds 3 | | Inner CV folds 4 | | Inner CV folds 5 | | Inner CV folds 6 | | Inner CV folds 7 | | Inner CV folds 8 |
|---|---|---|---|---|---|---|---|---|---|---|---|---|---|---|---|---|---|---|---|
| F0 | Train | | Train_train | | Train_train | | Train_train | | Train_train | | Train_train | | Train_train | | Train_train | | Train_train | | Train_validation |
| F1 | Train | | Train_train | | Train_train | | Train_train | | Train_train | | Train_train | | Train_train | | Train_train | | Train_validation | | Train_train |
| F2 | Train | | Train_train | | Train_train | | Train_train | | Train_train | | Train_train | | Train_train | | Train_validation | | Train_train | | Train_train |
| F3 | Train | | Train_train | | Train_train | | Train_train | | Train_train | | Train_train | | Train_validation | | Train_train | | Train_train | | Train_train |
| F4 | Train | zoom on train => | Train_train | + | Train_train | + | Train_train | + | Train_train | + | Train_validation | + | Train_train | + | Train_train | + | Train_train | + | Train_train |
| F5 | Train | | Train_train | | Train_train | | Train_train | | Train_validation | | Train_train | | Train_train | | Train_train | | Train_train | | Train_train |
| F6 | Train | | Train_train | | Train_train | | Train_validation | | Train_train | | Train_train | | Train_train | | Train_train | | Train_train | | Train_train |
| F7 | Train | | Train_train | | Train_validation | | Train_train | | Train_train | | Train_train | | Train_train | | Train_train | | Train_train | | Train_train |
| F8 | Train | | Train_validation | | Train_train | | Train_train | | Train_train | | Train_train | | Train_train | | Train_train | | Train_train | | Train_train |
| F9 | Test | | x | | x | | x | | x | | x | | x | | x | | x | | x |

**OUTER CROSS-VALIDATION FOLD 1**

| Data Fold | Outer CV folds 1 | | Inner CV folds 0 | | Inner CV folds 1 | | Inner CV folds 2 | | Inner CV folds 3 | | Inner CV folds 4 | | Inner CV folds 5 | | Inner CV folds 6 | | Inner CV folds 7 | | Inner CV folds 8 |
|---|---|---|---|---|---|---|---|---|---|---|---|---|---|---|---|---|---|---|---|
| F0 | Train | | Train_train | | Train_train | | Train_train | | Train_train | | Train_train | | Train_train | | Train_train | | Train_validation | | Train_train |
| F1 | Train | | Train_train | | Train_train | | Train_train | | Train_train | | Train_train | | Train_train | | Train_validation | | Train_train | | Train_train |
| F2 | Train | | Train_train | | Train_train | | Train_train | | Train_train | | Train_train | | Train_validation | | Train_train | | Train_train | | Train_train |
| F3 | Train | | Train_train | | Train_train | | Train_train | | Train_train | | Train_validation | | Train_train | | Train_train | | Train_train | | Train_train |
| F4 | Train | zoom on train => | Train_train | + | Train_train | + | Train_train | + | Train_validation | + | Train_train | + | Train_train | + | Train_train | + | Train_train | + | Train_train |
| F5 | Train | | Train_train | | Train_train | | Train_validation | | Train_train | | Train_train | | Train_train | | Train_train | | Train_train | | Train_train |
| F6 | Train | | Train_train | | Train_validation | | Train_train | | Train_train | | Train_train | | Train_train | | Train_train | | Train_train | | Train_train |
| F7 | Train | | Train_validation | | Train_train | | Train_train | | Train_train | | Train_train | | Train_train | | Train_train | | Train_train | | Train_train |
| F8 | Test | | x | | x | | x | | x | | x | | x | | x | | x | | x |
| F9 | Train | | Train_train | | Train_train | | Train_train | | Train_train | | Train_train | | Train_train | | Train_train | | Train_train | | Train_validation |

.

.

**OUTER CROSS-VALIDATION FOLD 9**

| Data Fold | Outer CV folds 9 | | Inner CV folds 0 | | Inner CV folds 1 | | Inner CV folds 2 | | Inner CV folds 3 | | Inner CV folds 4 | | Inner CV folds 5 | | Inner CV folds 6 | | Inner CV folds 7 | | Inner CV folds 8 |
|---|---|---|---|---|---|---|---|---|---|---|---|---|---|---|---|---|---|---|---|
| F0 | Test | | x | | x | | x | | x | | x | | x | | x | | x | | x |
| F1 | Train | | Train_train | | Train_train | | Train_train | | Train_train | | Train_train | | Train_train | | Train_train | | Train_train | | Train_validation |
| F2 | Train | | Train_train | | Train_train | | Train_train | | Train_train | | Train_train | | Train_train | | Train_train | | Train_validation | | Train_train |
| F3 | Train | | Train_train | | Train_train | | Train_train | | Train_train | | Train_train | | Train_train | | Train_validation | | Train_train | | Train_train |
| F4 | Train | zoom on train => | Train_train | + | Train_train | + | Train_train | + | Train_train | + | Train_train | + | Train_validation | + | Train_train | + | Train_train | + | Train_train |
| F5 | Train | | Train_train | | Train_train | | Train_train | | Train_train | | Train_validation | | Train_train | | Train_train | | Train_train | | Train_train |
| F6 | Train | | Train_train | | Train_train | | Train_train | | Train_validation | | Train_train | | Train_train | | Train_train | | Train_train | | Train_train |
| F7 | Train | | Train_train | | Train_train | | Train_validation | | Train_train | | Train_train | | Train_train | | Train_train | | Train_train | | Train_train |
| F8 | Train | | Train_train | | Train_validation | | Train_train | | Train_train | | Train_train | | Train_train | | Train_train | | Train_train | | Train_train |
| F9 | Train | | Train_validation | | Train_train | | Train_train | | Train_train | | Train_train | | Train_train | | Train_train | | Train_train | | Train_train |

+

**SPECIAL "ALL FOLDS" PERMUTATION for feature importance**

| Data Fold | Outer CV folds All | | Inner CV folds 0 | | Inner CV folds 1 | | Inner CV folds 2 | | Inner CV folds 3 | | Inner CV folds 4 | | Inner CV folds 5 | | Inner CV folds 6 | | Inner CV folds 7 | | Inner CV folds 8 | | Inner CV folds 9 |
|---|---|---|---|---|---|---|---|---|---|---|---|---|---|---|---|---|---|---|---|---|---|
| F0 | Train | | Train_train | | Train_train | | Train_train | | Train_train | | Train_train | | Train_train | | Train_train | | Train_train | | Train_train | | Train_validation |
| F1 | Train | | Train_train | | Train_train | | Train_train | | Train_train | | Train_train | | Train_train | | Train_train | | Train_train | | Train_validation | | Train_train |
| F2 | Train | | Train_train | | Train_train | | Train_train | | Train_train | | Train_train | | Train_train | | Train_train | | Train_validation | | Train_train | | Train_train |
| F3 | Train | | Train_train | | Train_train | | Train_train | | Train_train | | Train_train | | Train_train | | Train_validation | | Train_train | | Train_train | | Train_train |
| F4 | Train | zoom on train => | Train_train | + | Train_train | + | Train_train | + | Train_train | + | Train_train | + | Train_validation | + | Train_train | + | Train_train | + | Train_train | + | Train_train |
| F5 | Train | | Train_train | | Train_train | | Train_train | | Train_train | | Train_validation | | Train_train | | Train_train | | Train_train | | Train_train | | Train_train |
| F6 | Train | | Train_train | | Train_train | | Train_train | | Train_validation | | Train_train | | Train_train | | Train_train | | Train_train | | Train_train | | Train_train |
| F7 | Train | | Train_train | | Train_train | | Train_validation | | Train_train | | Train_train | | Train_train | | Train_train | | Train_train | | Train_train | | Train_train |
| F8 | Train | | Train_train | | Train_validation | | Train_train | | Train_train | | Train_train | | Train_train | | Train_train | | Train_train | | Train_train | | Train_train |
| F9 | Train | | Train_validation | | Train_train | | Train_train | | Train_train | | Train_train | | Train_train | | Train_train | | Train_train | | Train_train | | Train_train |

| METHODS: | |
|---|---|
| **General comments** | The Nested Cross-Validation is a normal double split train/val/test method, for which not just one, but the two splits are replaced with a CV instead. |
| **Outer Cross-Validation** | The Outer Cross-Validation is used to replace the split between train + val on one side, vs. test on the other side. Thanks to the Outer Cross-Validation, an unbiased testing prediction can be obtained for every sample.<br>Three common sources of confusion clarified:<br>**Yes**, using an Outer Cross-Validation does mean that the predictions do not all come from the same model. (For exemple for testing, the prediction for each fold comes from a different model.)<br>**No**, the Outer Cross-Validation has ***nothing*** to do with hyperparameter tuning or model selection.<br>Tuning a model only requires one Outer Cross-Validation. In contrast, it requires N_CV_folds distinct Inner Cross-Validations, one for each fold of the Outer Cross Validation. (see below) |
| **Inner Cross-Validation** | The Inner Cross-Validation is used to replace the split between the typical training set and validation set.<br>Thanks to the Inner Cross-Validation, hyperparameter selection can be decided based on a performance calculated on the full training set and validation set sample size, instead of the sole validation set sample size.<br>Two common sources of confusion clarified:<br>#1 - The whole purpose of performing an Inner Cross-Validation is ***hyperparameter selection*.**<br>The Inner Cross-Validation tunes the model that will then be used to generate predictions on the left out testing fold.<br>Therefore, the Inner Cross-Validation does ***not*** generate a testing prediction for more samples. That is the role of the Outer Cross-Validation.<br>One last time, the Inner Cross-Validation is ***only*** about improving the hyperparameter tuning of the model.<br>#2 - Tuning a model requires N_CV_folds distinct Inner Cross-Validations, one for each fold of the Outer Cross Validation. In contrast, it only requires one Outer Cross-Validation. (see above) |
| **Special Fold 0** | Special "All folds" permutation is only here for feature importance purposes.<br>We could extract the features importance from the 10 models used to generate the predictions on the testing set, but we would have to take the average of features importances between several model.<br>Addtionally each of these models only saw 90% of the data (10% was left aside for testing purposes).<br>If we only care about feature importance, we actually do not need to leave samples aside for testing. We can use 100% of the data to train a model using a 10-fold (not 9-fold this time, since the test fold can be used too!) ***Inner*** Cross-Validation to train a single model.<br>This model will never be tested, but it does not matter: its role is to provide the best estimate for the feature importance, which it does well.<br>Two common sources of confusion clarified:<br>#1 - The Nested Cross-Validation pipeline and the Special "All Folds" permutation pipeline are ***not*** intertwined and serve different purposes.<br>The purpose of the Nested Cross-Validation pipeline is to generate accurate and a unbiased testing prediction for every sample, whereas the purpose of the Special "All Folds" pipeline is to generate feature importance.<br>#2 - **Yes**, it does mean that the feature importance reported do not correspond to any of the 10 models (trained in the 10 Outer Cross-Validation folds) that generated the final testing predictions.<br>The feature importance comes from a model that was never used to generate predictions. |

| RESULTS: | **Example:** cells corresponding to the values used for each prediction set | **Explanation:** |
|---|---|---|
| **Training** | "E17+G17+I17+K17+M17+O17+Q17+S17+<br>E29+G29+I29+K29+M29+O29+Q29+U29+..." | To obtain the training prediction for the Data Fold F1, take the mean of every training prediction available on this data fold (10*8=80 of them). |
| **Validation** | "E25" | To obtain the validation prediction for the Data Fold F1, take the prediction outputted by the model trained on the remaining 8 data folds with the best hyperparameters values obtained for this Outer Cross-Validation fold.<br>These validation predictions can then be used to tune the ensemble models. /!\ Warning /!\: This can only be properly done if data folds split used for the Outer Cross-Validation and the Inner Cross-Validation is the same. |
| **Testing** | "C26" | No extra step is needed for the testing prediction for the Data Fold 1. Simply take the only testing prediction available for this fold. |
| **Feature importance:** | N/A | Extract the features importance from the special "outer fold" "All".<br>If want to compute the standard deviation for the feature importance as well, we can leverage the feature importance computed on each of the Outer Cross-Validation folds and take the square root of their variance. |

**SUPPLEMENTARY:**

**Comparison between Nested Cross-Validation and other validation methods:**
**#1**-Single split train/test.
Split train/test works when there are no hyperparameters to tune. If one uses a simple train/test split and tune the hyperparameters on the testing set, some overfitting is to be expected.
**#2**-Double split train/validation/test.
Because of the above, a train/validation/test double split is commonly used when hyperparameters tuning is required.
The training is performed on the training set, the tuning of the hyperparameters on the validation set, and the evaluation of the unbiased model performanc on the untouched test dataset.
**#3**-Cross-Validation(train/val) and split (train+val)/test.
With the pipeline #2 (double split train/validation/test), the tuning of the hyperparameters is not leveraging the dataset as much as with a Cross-Validation, as only one data fold is used to tune the hyperparameters.
Therefore machine learning practitioners usually replace the "inner split" (the training/validation split), by a Cross-Validation instead, but they keep the "outer split" (between the training+validation dataset used for their Cross-Validation and the testing set).
**#4**-Split train/val and Cross-Validation((train+val)/test).
Pipeline #3 only generate an unbiased testing prediction for a fraction of the dataset (the testing set).
To generate an unbiased prediction on every sample of the dataset, pipeline #4 can be used.
However, because there is a simple split between the training set and the validation set (one for each Cross-Validation fold), that means that the hypperparameter tuning only leverages a fraction of the dataset.
Pipeline #4 is preferred when generating an unbiased testing prediction for every sample is needed but the models are too time consuming to train to be able to afford a Nested Cross-Validation (#5).
Pipeline #4 is therefore the pipeline we used to predict chronological age using medical images or videos, for example.
**#5**-Nested Cross-Validation.
To fully leverage the whole dataset, a Nested Cross-Validation replaces both the outer split (train+val/test) and the inner splits (train/val) by a Cross-Validation, hence the nesting.
It requires the training of a large number of models, and it is therefore usually reserved for models that can be trained relatively quickly.
In our case, we used it to train our scalar features-based models (elastic nets, light gradient boosted machines, and shallow fully connected neural networks).

**Supplementary Table 41: Hyperparameters tuning upstream of the cross-validation for images-based models**

See supplementary data

**Supplementary Table 42: Outer Cross-Validation with inner split pipeline**

The values displayed are validation RMSE values. Lower values are associated with better hyperparameter tuning. When two values are displayed (value1/value2), the second value corresponds to the training RMSE. The architecture used was InceptionV3, with an initial learning rate of 0.001. The model was trained on the data folds 2-9, and validated on the data fold #0. The data fold #1 was set aside as the testing set and was not used.

| Data Fold | N=100 | Outer CV folds 0 | Outer CV folds 1 | Outer CV folds 2 | Outer CV folds 3 | Outer CV folds 4 | Outer CV folds 5 | Outer CV folds 6 | Outer CV folds 7 | Outer CV folds 8 | Outer CV folds 9 |
|---|---|---|---|---|---|---|---|---|---|---|---|
| F0 | N=10 | Train | Train | Train | Train | Train | Train | Train | Train | Validation | Test |
| F1 | N=10 | Train | Train | Train | Train | Train | Train | Train | Validation | Test | Train |
| F2 | N=10 | Train | Train | Train | Train | Train | Train | Validation | Test | Train | Train |
| F3 | N=10 | Train | Train | Train | Train | Train | Validation | Test | Train | Train | Train |
| F4 | N=10 | Train | Train | Train | Train | Validation | Test | Train | Train | Train | Train |
| F5 | N=10 | Train | Train | Train | Validation | Test | Train | Train | Train | Train | Train |
| F6 | N=10 | Train | Train | Validation | Test | Train | Train | Train | Train | Train | Train |
| F7 | N=10 | Train | Validation | Test | Train | Train | Train | Train | Train | Train | Train |
| F8 | N=10 | Validation | Test | Train | Train | Train | Train | Train | Train | Train | Train |
| F9 | N=10 | Test | Train | Train | Train | Train | Train | Train | Train | Train | Validation |

**Supplementary Table 43: Batch size for each transferred convolutional neural network architecture**

| Architecture | Batch size |
|---|---|
| DenseNet121 | 16 |
| DenseNet169 | 16 |
| DenseNet201 | 16 |
| InceptionResNetV2 | 8 |
| InceptionV3 | 32 |
| MobileNet | 128 |
| MobileNetV2 | 64 |
| NASNetLarge | 4 |
| NASNetMobile | 64 |
| ResNet101 | 16 |
| ResNet101V2 | 16 |
| ResNet152 | 16 |
| ResNet152V2 | 16 |
| ResNet50 | 32 |
| ResNet50V2 | 32 |
| ResNeXt50 | 4 |
| VGG16 | 32 |
| VGG19 | 32 |
| Xception | 32 |

**Supplementary Table 44: Aging dimensions included in each of the four general aging models**

| | Aging dimensions included in the ensemble | | | | | | | | | | |
|---|---|---|---|---|---|---|---|---|---|---|---|
| General aging model | Brain | Eyes | Hearing | Lungs | Arterial | Heart | Abdomen | Musculoskeletal | Physical Activity | Biochemistry | BloodCells |
| * | TRUE | TRUE | TRUE | TRUE | TRUE | TRUE | TRUE | TRUE | TRUE | FALSE | TRUE |

| | | | | | | | | | | | |
|---|---|---|---|---|---|---|---|---|---|---|---|
| *instances01 | FALSE | TRUE | TRUE | TRUE | non-images only | FALSE | FALSE | non-images only | FALSE | TRUE | TRUE |
| *instances1.5x | FALSE | FALSE | FALSE | FALSE | FALSE | FALSE | FALSE | FALSE | TRUE | FALSE | FALSE |
| *instances23 | TRUE | FALSE | FALSE | FALSE | TRUE | TRUE | TRUE | TRUE | FALSE | FALSE | FALSE |

**Supplementary Table 45: Hyperparameters values selected during the tuning of time series-based models**

| | Hyperparameter | | | | |
|---|---|---|---|---|---|
| Dataset | learning_rate | dropout | kernel_regularizer | bias_regularizer | activity_regularizer |
| Pulse Wave Analysis | 0.001 | 0.2 | 1.00E-06 | 1.00E-06 | None |
| ECG | 0.001 | 0.2 | 1.00E-06 | 1.00E-04 | None |
| Physical Activity Fullweek 1D acceleration | 0.001 | 0.5 | 1.00E-04 | None | None |
| Physical Activity Fullweek features | 0.001 | 0.2 | None | None | None |
| Physical Activity Walking 3D acceleration | 0.001 | 0.2 | 1.00E-05 | 1.00E-04 | None |
